# Weight loss and permanent discontinuation across obesity treatment-initiation strategies: Bayesian longitudinal network meta-analysis

**DOI:** 10.64898/2026.11.13.26367524

**Authors:** Xinbo Yin, Minhui Dai, Zhuying Xia, Dongliang Liu, Guoqing Huang, Xiangmin Li, Xiaokai Wang

**Affiliations:** Department of Emergency Medicine, Xiangya Hospital, Central South University. Changsha. Hunan. China; Teaching and Research Section of Clinical Nursing, Xiangya Hospital, Central South University. Changsha. Hunan. China; National Clinical Research Center for Geriatric Disease (Xiangya Hospital), Central South University. Changsha. Hunan. China; Department of Clinical Nutrition, Xiangya Hospital, Central South University, Changsha. Hunan. China; Department of Endocrinology, Xiangya Hospital, Central South University, Changsha. Hunan. China

## Abstract

**Background:** Obesity pharmacotherapy strategies differ in maintenance dose, titration and lifestyle support, but existing syntheses largely compare drugs rather than treatment-initiation decisions. We assessed 1-year weight reduction, permanent discontinuation and incremental benefit–discontinuation trade-offs under aligned estimands, while mapping attrition of eligible trial evidence.

**Methods:** We searched eight databases and two trial registries from inception to October, 2026, for randomised trials (≥12 weeks) of obesity pharmacotherapy initiation in adults with overweight or obesity. Maintenance, withdrawal and switching trials were excluded. Two reviewers independently selected studies, extracted data and assessed bias and certainty. We mapped analytical attrition and used Bayesian longitudinal network meta-analysis to estimate treatment-policy-compatible weight change at 52 weeks, alongside reference networks and analyses of discontinuation, dose escalation, titration, lifestyle support and diabetes-related attenuation. The protocol was prospectively registered with PROSPERO (CRD420261532130).

**Findings:** We included 180 reports from 178 trials (196 randomisation units; 81 014 participants). Only 78 units met criteria for 1-year assessment; the primary longitudinal model in adults without diabetes incorporated 50 units (including trials reporting 12–24-week interim data), and 45 of 160 target strategies had non-extrapolated 52-week support. The largest placebo-adjusted weight reductions were with retatrutide 12 mg (−19·80 percentage points, 95% CrI −21·81 to −17·78), tirzepatide 15 mg (−16·87, −18·07 to −15·51) and cagrilintide–semaglutide 2·4/2·4 mg (−16·37, −18·10 to −14·70). Only tirzepatide 15 mg and semaglutide 2·4 mg had high-certainty evidence. Thirteen of 32 evaluated strategies significantly increased adverse-event-related permanent discontinuation versus placebo, whereas none significantly increased all-cause discontinuation. Five of 22 adjacent dose steps had ≥0·80 probability of adding >2 percentage points of weight reduction, although discontinuation effects were uncertain or unavailable for some steps. Only eight 1-year titration contrasts and three trials of randomised lifestyle-support increments were identified. Weight reduction in adults with type 2 diabetes was 62% of that without diabetes (ratio 0·62, 95% CI 0·58–0·66).

**Interpretation:** Only a small fraction of eligible trials directly informs 1-year treatment-initiation decisions. Higher-dose and multi-receptor strategies offer substantial weight reduction, but certainty and discontinuation trade-offs vary. Standardised treatment-policy reporting and randomised evidence on titration and lifestyle support are needed.

**Funding:** None.

## Introduction

Obesity affects more than one billion people worldwide, with prevalence projected to rise through 2050.^1,2^ Semaglutide 2·4 mg and tirzepatide achieve substantial weight reduction, and newer multi-receptor, oral and long-acting agents are expanding treatment options.^3–6^ Yet initiating treatment involves decisions beyond drug choice: the target maintenance dose, escalation schedule and intensity of adjunct lifestyle support. These decisions must balance additional weight reduction against tolerability and permanent discontinuation, because adverse events increase with dose, cessation can lead to weight regain and discontinuation is common in routine practice.^7–10^

Previous network meta-analyses established comparative drug efficacy,^11–13^ but compared drugs or standard maintenance doses rather than incremental initiation decisions, and longitudinal and dose–response methods^14,15^have rarely been applied to aligned initiation strategies. Four features of the evidence complicate such comparisons. First, treatment-policy and hypothetical or trial-product estimands can yield materially different estimates, as in trials of cagrilintide–semaglutide and orforglipron.^5,16,17^ Second, trials randomising after weight-loss induction or during withdrawal address maintenance rather than initiation.^8,9,18^ Third, lifestyle support is usually identical across arms, precluding estimation of its incremental benefit.^19^ Fourth, weight reduction differs by type 2 diabetes status, requiring separate analyses.^20,21^

We therefore conducted an estimand-aligned systematic review and Bayesian longitudinal network meta-analysis to quantify how much eligible evidence informs 1-year initiation decisions. We estimated strategy-specific weight reduction and permanent discontinuation, assessed the incremental effects of dose escalation, titration and lifestyle support, bounded joint response without discontinuation and examined attenuation of treatment effects in type 2 diabetes.

## Methods

### Study design and search strategy

We conducted a systematic review and network meta-analysis of randomised controlled trials following PRISMA 2020^22^, PRISMA-NMA^23^ and PRISMA-S^24^ guidance (completed checklists in appendices 2–5; full search strategies in appendix 1). The protocol was prospectively registered with PROSPERO (CRD420261532130) on 7 October 2026, before data extraction. Pre-specified analyses were established in version 1.0; dose-step benefit–discontinuation analyses and the joint response-without-discontinuation analysis were incorporated in version 1.1 after initial data extraction. Evidence-funnel layering, the three-level meta-regression of diabetes-related attenuation, posterior probability thresholds (5 and 10 percentage points), the 5-percentage-point clinical importance threshold for GRADE, and selection of the maridebart cafraglutide 420 mg reference arm were defined after version 1.1. In deviation from protocol section 50 (which prespecified Bayesian binomial models reporting risk ratios), binary outcomes were synthesised using contrast-based frequentist random-effects models reporting odds ratios with continuity corrections for sparse events (appendix).

### Eligibility criteria and treatment strategies

We included randomised trials of at least 12 weeks in adults (≥ 18 years) with overweight or obesity and an explicit weight-management objective; comorbidity-selected trials were eligible if weight management was prespecified. Eligible interventions, including investigational agents, were GLP-1-related agents, amylin-related agents, orlistat, phentermine–topiramate and naltrexone–bupropion. We excluded trials of randomised withdrawal, switching, de-escalation or post-bariatric management, trials randomising after active weight-loss induction, lifestyle-only trials and ineligible populations. Two reviewers independently screened records, resolving disagreements by consensus or a third reviewer. Reports were linked to parent trials, and independently randomised cohorts were counted as separate randomisation units. Inclusion and exclusion criteria, together with the reasons recorded at each screening stage, are tabulated in appendix 7.

Each strategy was defined by molecule or combination, formulation, route, dosing frequency, target maintenance dose and titration policy, and formed a separate network node. Maximum-tolerated-dose regimens were analysed as randomised, and non-target interventions within eligible trials were retained as connector nodes. Lifestyle support was classified by behavioural, nutritional and exercise components, with delivery intensity recorded separately. Coding rules and the post-hoc selection of the maridebart cafraglutide 420 mg reference arm are in the appendix.^6^

### Outcomes and estimands

The primary outcome was percentage body-weight change from randomisation at 52 weeks (window 48–56 weeks), expressed as a between-group mean difference in percentage points. Secondary time points were 12 weeks (10–14) and 24 weeks (20–28); observations outside these windows retained their actual follow-up times.

The principal implementation outcome was all-cause permanent discontinuation of the assigned drug strategy, distinguished from temporary interruption, dose reduction and study withdrawal. Secondary outcomes were weight-loss thresholds (≥5%, ≥10%, ≥15% and ≥20%), absolute weight change, waist circumference, physical function, body composition, adverse-event-related permanent discontinuation, serious adverse events and gastrointestinal events.

Two reviewers independently extracted or verified data from each report using a standardised electronic form; the form, with the definition, permissible values and source hierarchy of every field, is provided with a worked example in appendix 6. Extracted items included trial and report identifiers, design, phase and number of sites, population and baseline characteristics, the full strategy definition (molecule, formulation, route, dosing frequency, target maintenance dose and titration policy), background lifestyle support, outcome definitions with time points and denominators, estimand-relevant trial conduct (handling of treatment discontinuation, rescue treatment, switching and missing data), effect estimates with precision, funding source and risk-of-bias judgements. Estimates were prioritised by prespecification, degree of adjustment and source reliability, and figure-derived values were used when necessary; disagreements were resolved against the source report or by a third reviewer. We wrote to the corresponding authors of trials with missing or unusable data; no additional data were provided in reply. Estimands were classified from each trial’s handling of treatment discontinuation, rescue treatment, switching and missing data rather than terminology alone.^17^ Primary analyses used treatment-policy-compatible estimates; hypothetical, trial-product, on-treatment and historical last-observation-carried-forward (LOCF) estimates were analysed separately. Analyses were stratified by type 2 diabetes status, with mixed populations as a separate stratum.

### Risk of bias and certainty of evidence

Two reviewers independently assessed outcome-specific risk of bias with RoB 2,^25^ and bias due to missing evidence, including selective non-reporting and small-study effects, with ROB-MEN.^26^ Certainty of 52-week placebo comparisons was assessed with CINeMA^27^ and GRADE,^28^ adapted for longitudinal extrapolation, figure-derived data, and indirect populations. A 5-percentage-point threshold was adopted post hoc to define clinical importance based on regulatory benchmarks (eg, US Food and Drug Administration guidance for obesity drug development) within the GRADE minimally contextualised framework.^28^

### Evidence funnel

We mapped eligible trials through prespecified analytical layers: the master evidence base (L1), eligibility for 1-year assessment (L2), usable 48–56-week weight outcomes (L3), the treatment-policy reference network (L4), the longitudinal model (L4′), non-extrapolated strategies eligible for ranking (L5) and decision-specific contrasts (L6). Attrition was quantified using independent randomisation units, with reasons for non-contribution recorded at each stage. Trials excluded from a particular synthesis remained in the evidence map and missing-evidence assessment.

### Statistical analysis

Analyses were organised into four layers: drug strategies under shared lifestyle support, randomised lifestyle-support increments under identical pharmacotherapy, descriptive cross-trial comparisons of background support and randomised titration schedules reaching the same maintenance dose. Disconnected networks were analysed separately. For each outcome, time window and diabetes stratum, we fitted contrast-based frequentist random-effects reference network meta-analyses. Common heterogeneity was estimated by restricted maximum likelihood, and multi-arm correlations were retained through reported, recovered or assumed covariances. Effects were expressed as mean differences or odds ratios with absolute risks, 95% CIs and, where estimable, prediction intervals; sparse binary data used continuity corrections. Inconsistency was assessed globally with the design-by-treatment interaction model^29^ and locally by node-splitting,^30^ and transitivity through the distribution of clinical and design characteristics across comparisons.^31^

The primary Bayesian longitudinal model used a multivariate normal likelihood to jointly synthesise treatment-policy-compatible, placebo-adjusted percentage weight-change contrasts from 12 to 104 weeks. Multi-arm and repeated-measure correlations were retained, with unreported temporal correlations assumed. Trajectories were modelled with restricted cubic splines anchored at randomisation, with internal knots at approximately 12 and 24 weeks. Strategy-specific coefficients were partially pooled within prespecified, mechanistically coherent drug classes, with trial-level random effects. Doses remained distinct strategies, and no monotonic dose–response was imposed. Models were fitted in Stan^32^ with weakly informative priors; specifications, convergence diagnostics and software versions are in the appendix.

Each 52-week estimate was classified as observed, interpolated, borrowing-dependent or extrapolated, and extrapolated strategies were excluded from rankings and decision analyses. We report posterior median mean differences, 95% credible intervals (CrIs), prediction intervals and probabilities of exceeding clinically relevant thresholds; rankings and SUCRA were secondary^33^.

We compared adjacent maintenance-dose steps within the same molecule, route and dosing frequency, estimating posterior probabilities that additional weight reduction exceeded 0·5, 1·0 or 2·0 percentage points; these were weighed against all-cause and adverse-event-related permanent discontinuation without a composite benefit–harm score. Placebo-adjusted between-arm contrasts for maridebart cafraglutide were derived post hoc.

To bound the proportion achieving at least 10% weight reduction without permanent discontinuation, without assuming independence, we applied Fréchet–Hoeffding bounds to compatible arm-level probabilities.^34^ For response probability pR and discontinuation probability pC, the joint probability q satisfies max(0, pR−pC) ≤ q ≤ min(pR, 1−pC). For diabetes-related attenuation, we compared separately randomised populations with and without diabetes receiving the same strategy at matched follow-up times. Log ratios of their placebo-adjusted mean differences were pooled in a three-level random-effects model,^35^ with doses nested within trial programmes.

Sensitivity analyses varied estimands, time windows, covariance assumptions, trajectory functions, heterogeneity priors and class-level borrowing, excluded trials by risk of bias, data source and trial characteristics, and omitted each parent trial in turn. Small-study effects were assessed with comparison-adjusted funnel plots^36^ and Egger’s regression^37^ for outcomes with at least ten trials.

### Data, code and other materials

The data that support the findings of this review, the node dictionary, the screening and extraction records and the analysis code (R and Stan) are available from the corresponding author on reasonable request. The protocol is available from the PROSPERO record (CRD420261532130).

## Results

### Study selection and evidence availability

Searches identified 48 770 database and registry records and 148 from other sources; after deduplication, 28 914 were screened and 986 assessed for eligibility. We included 180 reports of 178 parent trials (196 independent randomisation units; 81 014 participants; figure 1; appendix tables S2, S24). The evidence base contained 214 network nodes, including 160 target strategies across 13 drug classes. Of the 196 units, 138 enrolled adults without type 2 diabetes, 29 adults with diabetes and 29 mixed or incompletely characterised populations (table 1).

**Figure 1:**
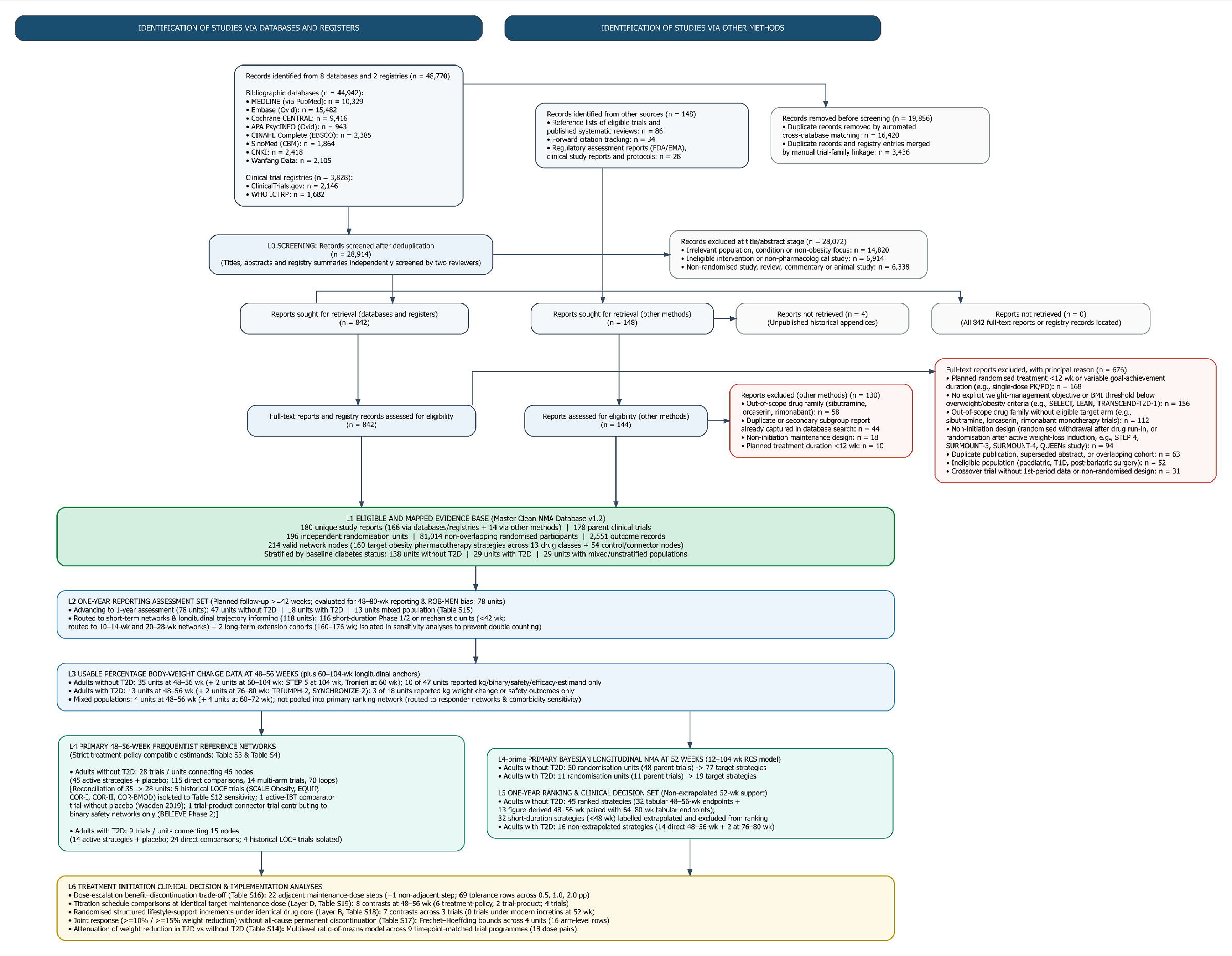
PRISMA 2020 flow diagram and analytical evidence-funnel cascade from eligible mapped trials to clinical decision synthesis layers Top panels display the PRISMA 2020 flow diagram depicting study identification, deduplication, abstract screening, full-text retrieval, and exclusions with primary reasons across eight electronic databases (MEDLINE, Embase, Cochrane CENTRAL, PsycINFO, CINAHL, SinoMed, CNKI, and Wanfang Data), two trial registries (ClinicalTrials.gov and WHO ICTRP), and supplementary searching sources (reference lists, forward citations, regulatory reports, and clinical study reports). Bottom panels display the step-by-step evidence-funnel cascade structured by prespecified protocol gates (L1–L6), tracking independent randomisation units rather than publications as denominators: L1 (Eligible and mapped evidence base): 180 study reports representing 178 parent clinical trials, 196 independent randomisation units, 81,014 non-overlapping randomised participants, 2,551 outcome records, and 214 valid network nodes (160 target obesity pharmacotherapy strategies across 13 drug classes plus 54 control or connector nodes). L2 (One-year reporting assessment set): 78 initiation-stage units with planned treatment follow-up ≥42 weeks assessed for 48–80-week reporting status and ROB-MEN risk of bias due to missing evidence (47 without T2D, 18 with T2D, and 13 mixed/unstratified population); 118 non-advancing units comprise 116 short-duration Phase 1/2 or mechanistic units (<42 weeks, routed to 10–14-week and 20–28-week cross-sectional networks and longitudinal time-course models) and two isolated long-term extension cohorts (160–176 weeks, isolated to prevent double counting). L3 (Usable percentage body-weight change at 48–56 weeks): randomisation units reporting compatible percentage weight change at 48–56 weeks or long-term anchors at 60–104 weeks. L4 (Primary 48–56-week frequentist reference networks): units meeting strict treatment-policy-compatible estimands and connected to the reference networks (28 units connecting 46 nodes in adults without T2D; 9 units connecting 15 nodes in adults with T2D). Five historical LOCF trials in non-diabetic cohorts (SCALE Obesity, EQUIP, COR-I, COR-II, and COR-BMOD) and four in diabetic cohorts (OB-202/DM-230, COR-Diabetes, Hollander 1998, and Miles 2002) were partitioned into sensitivity analyses. L4′ (Primary Bayesian longitudinal model): 50 units (48 parent trials; 77 target strategies) without T2D and 11 units (11 trials; 19 target strategies) with T2D informing the restricted cubic spline longitudinal model across 12–104 weeks. L5 (One-year ranking and clinical decision set): 45 ranked strategies in adults without T2D with non-extrapolated 52-week support (32 tabular 48–56-week endpoints and 13 figure-derived 48–56-week trajectories paired with 64–80-week tabular endpoints); 32 short-duration strategies (<48 weeks) were designated extrapolated and excluded from ranking. L6 (Treatment-initiation clinical decision and implementation analyses): implementation syntheses encompassing 22 adjacent maintenance-dose escalation steps (69 tolerance evaluations), eight randomised titration contrasts at identical target doses (Layer D), seven randomised structured lifestyle-support increments (Layer B), arm-level Fréchet–Hoeffding joint response-persistence bounds across four units (16 arm-level rows), and 18 timepoint-matched dose pairs across nine programmes evaluating weight-reduction attenuation in T2D. Abbreviations: T1D = type 1 diabetes; T2D = type 2 diabetes; BMI = body mass index; PK/PD = pharmacokinetics/pharmacodynamics; LOCF = last observation carried forward; IBT = intensive behavioural therapy; MTD = maximum tolerated dose; NMA = network meta-analysis; rcs = restricted cubic spline; ROB-MEN = Risk of Bias due to Missing Evidence in Network Meta-Analysis.

**Table 1:**
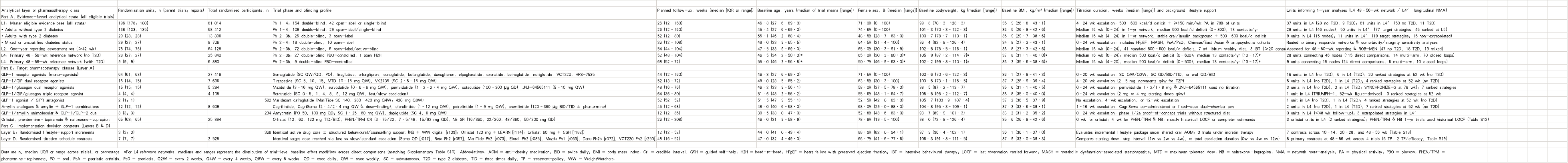
Characteristics of included randomised trials and baseline participant profiles across the evidence-funnel layers and pharmacotherapy classes.

| Analytical layer or pharmacotherapy class | Randomisation units, n (parent trials, reports) | Total randomised participants, n | Trial phase and blinding profile | Planned follow-up, weeks (median [IQR or range]) | Baseline age, years (median of trial means [range]) | Female sex, % (median [range]) | Baseline bodyweight, kg (median [range]) | Baseline BMI, kg/m² (median [range]) | Titration duration, weeks (median [range]) and background lifestyle support | Units informing 1-year analyses (L4 48–56-wk network / L4′ longitudinal NMA) |
|---|---|---|---|---|---|---|---|---|---|---|
| Part A: Evidence-funnel analytical strata (all eligible trials) |  |  |  |  |  |  |  |  |  |  |
| L1: Master eligible evidence base (all strata) | 196 (178; 180) | 81 014 | Ph 1–4; 154 double-blind, 42 open-label or single-blind | 26 (12–160) | 46·8 (27·6–69·0) | 71·0% (0–100) | 99·8 (70·3–128·3) | 35·9 (26·8–43·1) | 4–24 wk escalation; 500–600 kcal/d deficit + ≥150 min/wk PA in 78% of units | 37 units in L4 (28 no T2D, 9 T2D); 61 units in L4′ (50 no T2D, 11 T2D) |
| • Adults without type 2 diabetes | 138 (133; 135) | 58 412 | Ph 1–4; 109 double-blind, 29 open-label/single-blind | 26 (12–160) | 45·4 (27·6–69·0) | 74·6% (0–100) | 101·3 (70·3–122·3) | 36·5 (26·8–42·6) | Median 16 wk (0–24) in 1-yr network; median 500 kcal/d deficit (0–600); 13 contacts/yr | 28 units in L4 (46 nodes); 50 units in L4′ (77 target strategies, 45 ranked at L5) |
| • Adults with type 2 diabetes | 29 (28; 28) | 13 896 | Ph 2–3b; 26 double-blind, 3 open-label | 52 (12–80) | 55·1 (46·2–68·4) | 48·5% (28·7–63·0) | 100·7 (78·7–110·1) | 35·9 (29·7–38·6) | Median 16 wk (4–24) in 1-yr network; stable oral/insulin background + 500–600 kcal/d deficit | 9 units in L4 (15 nodes); 11 units in L4′ (19 target strategies, 16 non-extrapolated) |
| • Mixed or unstratified diabetes status | 29 (27; 27) | 8 706 | Ph 2–4; 19 double-blind, 10 open-label | 36 (12–108) | 45·0 (33·9–65·5) | 64·5% (21·4–100) | 96·4 (88·8–126·4) | 34·9 (27·3–43·1) | 0–24 wk escalation; includes HFpEF, MASH, PsA/PsO, Chinese/East Asian & antipsychotic cohorts | Routed to binary responder networks & comorbidity/integrity sensitivity analyses |
| L2: One-year reporting assessment set (≥42 wk) | 78 (74; 76) | 64 128 | Ph 2–3b; 72 double-blind, 6 open-label/active-blind | 54 (44–104) | 47·5 (33·9–69·0) | 65·0% (30·3–91·9) | 102·5 (78·5–116·1) | 36·8 (27·3–42·6) | Median 16 wk (0–24); 41 standard 500–600 kcal/d deficit, 7 ad libitum healthy diet, 3 IBT (≥20 conta[illegible] | Assessed for 48–80-wk reporting & ROB-MEN (47 no T2D, 18 T2D, 13 mixed) |
| L4: Primary 48–56-wk reference network (no T2D) | 28 (27; 27) | 25 840 | Ph 2–3b; 27 double-blind PBO-controlled, 1 open H2H | 52 (48–104) | 45·5 (34·2–50·0)* | 65·0% (30·3–80·0)* | 105·9 (87·2–114·7)* | 37·8 (31·1–40·0)* | Median 16 wk (0–24); median 500 kcal/d deficit (0–600); median 13 contacts/yr (13–17)* | 28 units connecting 46 nodes (115 direct comparisons, 14 multi-arm, 70 closed loops) |
| L4: Primary 48–56-wk reference network (with T2D) | 9 (9; 9) | 6 880 | Ph 2–3b; 9 double-blind PBO-controlled | 68 (52–72) | 55·0 (46·2–56·8)* | 50·7% (46·9–63·0)* | 102·2 (99·8–110·1)* | 36·2 (35·6–38·6)* | Median 16 wk (4–20); median 500 kcal/d deficit (0–500); median 13 contacts/yr (13–17)* | 9 units connecting 15 nodes (24 direct comparisons, 6 multi-arm, 10 closed loops) |
| Part B: Target pharmacotherapy classes (Layer A) |  |  |  |  |  |  |  |  |  |  |
| GLP-1 receptor agonists (mono-agonists) | 64 (61; 63) | 27 418 | Semaglutide (SC QW/QD, PO), liraglutide, orforglipron, ecnoglutide, bofanglutide, danuglipron, efpeglenatide, exenatide, beinaglutide, noiiglutide, VCT220, HRS-7535 | 44 (12–160) | 46·3 (27·6–69·0) | 71·5% (0–100) | 100·6 (70·6–122·3) | 36·1 (27·9–41·3) | 0–20 wk escalation; SC QW/Q2W, SC QD/BID/TID, or oral QD/BID | 16 units in L4 (no T2D), 6 in L4 (T2D); 20 ranked strategies at 52 wk (no T2D) |
| GLP-1/GIP dual receptor agonists | 16 (14; 15) | 7 696 | Tirzepatide (SC 5, 10, 15, MTD 10–15 mg QW), VK2735 (SC 2·5–15 mg QW) | 52 (13–72) | 48·0 (28·5–65·2) | 63·5% (30·3–100) | 103·5 (73·1–115·5) | 37·3 (28·9–39·4) | 4–20 wk escalation (2·5 mg increments q4w for TZP) | 5 units in L4 (no T2D), 1 in L4 (T2D); 4 ranked strategies at 52 wk (no T2D) |
| GLP-1/glucagon dual receptor agonists | 15 (15; 15) | 5 294 | Mazdutide (3–16 mg QW), survodutide (0·6–6 mg QW), pemvidutide (1·2–2·4 mg QW), cotadutide (100–300 µg QD), JNJ-64565111 (5–10 mg QW) | 48 (16–76) | 48·3 (33·9–56·1) | 58·0% (37·5–78·0) | 98·5 (87·2–113·7) | 35·6 (31·1–40·5) | 0–24 wk escalation; pemvidutide 1·2/1·8 mg & JNJ-64565111 used no titration | 3 units in L4 (no T2D), 0 in L4 (T2D; SYNCHRONIZE-2 at 76 wk); 7 ranked strategies |
| GLP-1/GIP/glucagon triple receptor agonist | 4 (4; 4) | 4 108 | Retatrutide (SC 0·5, 1, 4, 8, 9, 12 mg QW; fast/slow escalation) | 64 (36–80) | 51·6 (48·2–56·2) | 55·6% (48·1–64·7) | 105·6 (98·2–112·7) | 38·0 (35·0–40·0) | 0–24 wk escalation (2 mg or 4 mg starting doses q4w) | 1 unit in L4 (TRIUMPH-1, 52-wk figure-derived); 3 ranked strategies at 52 wk |
| GLP-1 agonist / GIPR antagonist | 2 (1; 1) | 592 | Maridebart cafraglutide (MariTide SC 140, 280, 420 mg Q4W; 420 mg Q8W) | 52 (52–52) | 51·5 (47·9–55·1) | 52·5% (42·0–63·0) | 105·7 (103·9–107·4) | 37·2 (36·5–37·9) | No escalation, 4-wk escalation, or 12-wk escalation | 1 unit in L4 (no T2D), 1 in L4 (T2D); 4 ranked strategies at 52 wk (no T2D) |
| Amylin analogues & amylin + GLP-1 combinations | 12 (12; 12) | 8 609 | Cagrilintide, CagriSema (2·4/2·4 mg QW & dose-finding), eloralintide (1–12 mg QW), petrelintide (1–9 mg QW), pramlintide (120–360 µg BID/TID ± phentermine) | 45 (12–68) | 48·0 (40·6–58·0) | 69·0% (29·0–88·0) | 104·8 (85·3–109·1) | 37·2 (32·6–39·1) | 1–16 wk escalation; CagriSema co-administered or fixed-dose dual-chamber pen | 2 units in L4 (no T2D), 1 in L4 (T2D); 7 ranked strategies at 52 wk (no T2D) |
| GLP-1/amylin unimolecular & GLP-1/GLP-2 dual | 3 (3; 3) | 234 | Amycretin (PO 50, 100 mg QD; SC 1·25–60 mg QW), dapiglutide (SC 4, 6 mg QW) | 12 (12–36) | 39·5 (38·0–47·0) | 52·8% (43·6–63·0) | 93·7 (89·9–101·3) | 33·2 (31·2–35·2) | 0–24 wk escalation; phase 1/2a proof-of-concept trials without structured diet | 0 units in L4 (<48 wk follow-up); 3 extrapolated strategies in L4′ |
| Orlistat, phentermine–topiramate & naltrexone–bupropion | 65 (63; 65) | 25 894 | Orlistat (60, 120 mg TID/BID), PHEN/TPM CR (3·75/23, 7·5/46, 15/92 mg QD), NB SR (16/360, 32/360, 48/360, 50/300 mg QD) | 26 (12–208) | 46·0 (31·9–58·9) | 78·8% (19·5–100) | 98·0 (72·8–126·4) | 35·6 (26·8–42·6) | 0 wk for orlistat; 4 wk for PHEN/TPM & NB; mostly historical LOCF or completer estimands | 3 orlistat units in L4 (2 ranked strategies); PHEN/TPM & NB 1-yr trials used historical LOCF (Table S12) |
| Part C: Implementation decision contrasts (Layers B & D) |  |  |  |  |  |  |  |  |  |  |
| Layer B: Randomised lifestyle-support increments | 3 (3; 3) | 368 | Identical active drug core ± structured behavioural/counselling support (NB + WW digital [r109], Orlistat 120 mg + LEARN [r114], Orlistat 60 mg + GSH [r182]) | 12 (12–52) | 44·0 (41·0–49·4) | 88·9% (82·0–94·1) | 97·9 (96·4–102·1) | 36·1 (36·1–37·0) | Evaluates incremental lifestyle package under shared oral AOM; 0 trials under incretin therapy | 7 contrasts across 10–14, 20–28, and 48–56 wk (Table S18) |
| Layer D: Randomised titration schedule contrasts | 7 (7; 7) | 2 528 | Identical target dose reached via fast vs slow/standard escalation (Sema QD [r017], Reta Ph2 [r057], MariTide Ph2 [r073], Eloral Ph2 [r085], Mazdu Ph1 [r063], Danu Ph2b [r072], VCT220 Ph2 [r250]) | 48 (16–52) | 47·0 (32·4–49·0) | 66·7% (41·6–77·6) | 106·3 (91·8–111·5) | 37·9 (32·0–39·3) | Compares starting dose, step interval (1w vs 2w vs 4w), or total escalation duration (0w vs 4w vs 12w) | 8 primary contrasts at 48–56 wk across 4 trials (6 TP, 2 TP/efficacy; Table S19) |

Data are n, median (IQR or range across trials), or percentage. \*For L4 reference networks, medians and ranges represent the distribution of trial-level baseline effect modifiers across direct comparisons (matching Supplementary Table S10). Abbreviations: AOM = anti-obesity medication; BID = twice daily; BMI = body mass index; CrI = credible interval; GSH = guided self-help; H2H = head-to-head; HFpEF = heart failure with preserved ejection fraction; IBT = intensive behavioural therapy; LOCF = last observation carried forward; MASH = metabolic dysfunction-associated steatohepatitis; MTD = maximum tolerated dose; NB = naltrexone–bupropion; NMA = network meta-analysis; PA = physical activity; PBO = placebo; PHEN/TPM = phentermine–topiramate; PO = oral; PsA = psoriatic arthritis; PsO = psoriasis; Q2W = every 2 weeks; Q4W = every 4 weeks; Q8W = every 8 weeks; QD = once daily; QW = once weekly; SC = subcutaneous; T2D = type 2 diabetes; TID = three times daily; TP = treatment-policy; WW = WeightWatchers.

**Table 1B:**
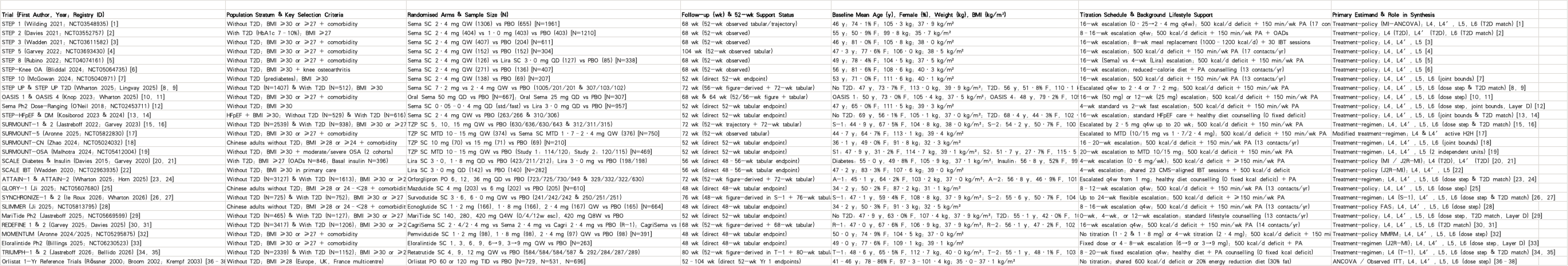
Key randomised controlled trials informing the primary 1 - year comparative efficacy and decision networks (adults without and with type 2 diabetes)

Across the funnel (figure 1), only 78 units met criteria for 1-year assessment (L2). Of 47 such units without diabetes, 35 reported usable 48–56-week percentage weight change, and 28 of these formed the treatment-policy reference network (46 nodes, 115 direct comparisons, 70 closed loops; figure 3). The primary longitudinal model (L4′) was not nested within L2: it incorporated all 50 units without diabetes (from 48 parent trials; 28 with 48–56-week observations and 22 short-duration or long-term anchors) contributing treatment-policy contrasts between 12 and 104 weeks, informing 77 target strategies (table 1).

### Risk of bias and missing evidence

No outcome assessment was at low overall risk of bias, mainly because of missing outcome data and selection of reported results (figure 2). High overall risk applied to 18% of continuous efficacy assessments, 6% of binary weight-response assessments and 2% of safety or discontinuation assessments, but none of the 28 reference-network trials was at high overall risk for percentage weight change. ROB-MEN rated 27 of 71 drug–placebo comparisons at high risk of bias due to missing evidence (appendix table S15).

**Figure 2.**
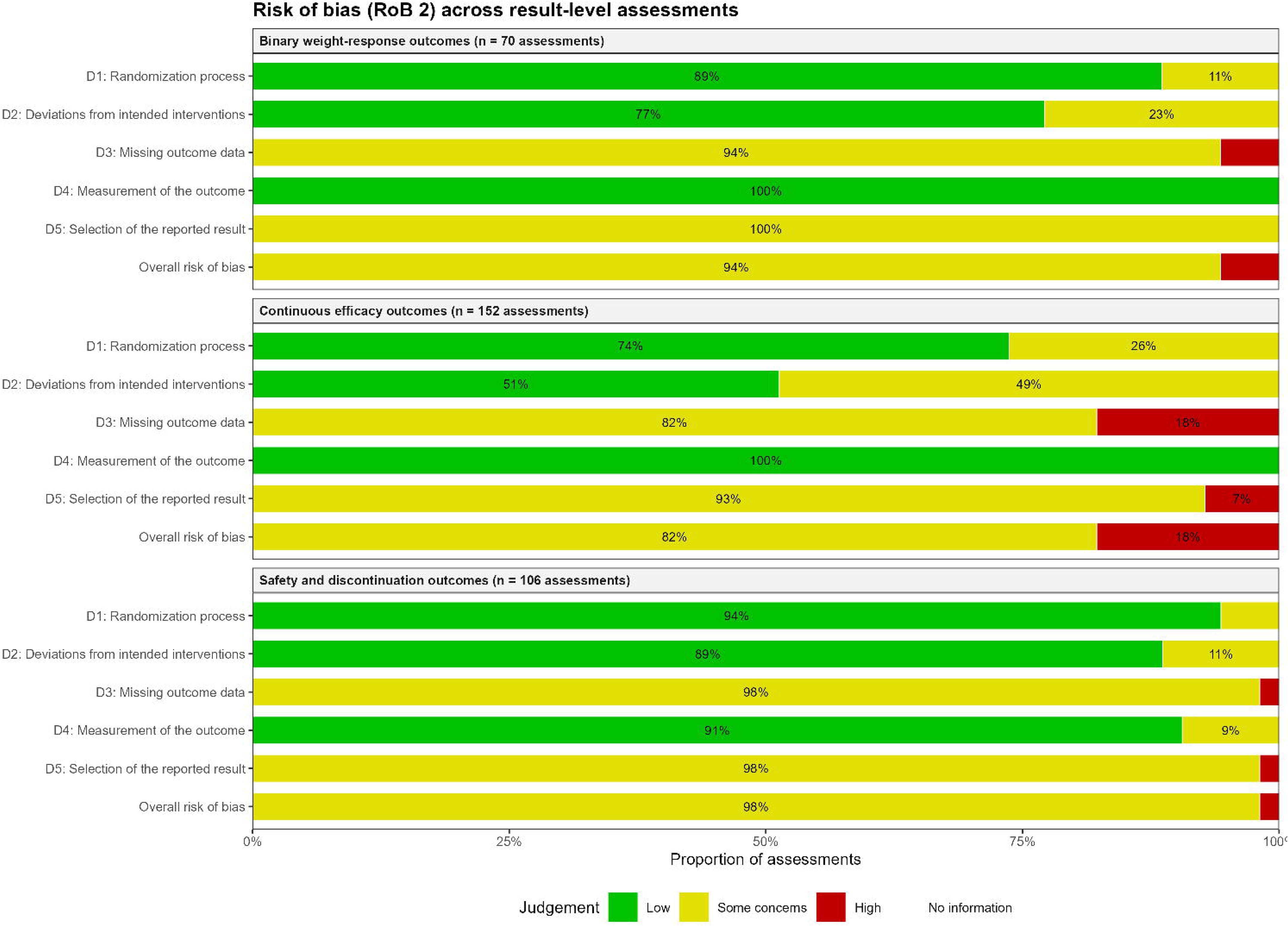
Risk-of-bias summary (RoB 2). Bars show the proportion of result-level assessments judged at low risk, some concerns or high risk in each domain and overall, separately for Binary weight-response outcomes; Continuous efficacy outcomes; Safety and discontinuation outcomes. Study-by-study judgements are shown in the Supplementary Material.

**Figure 3.**
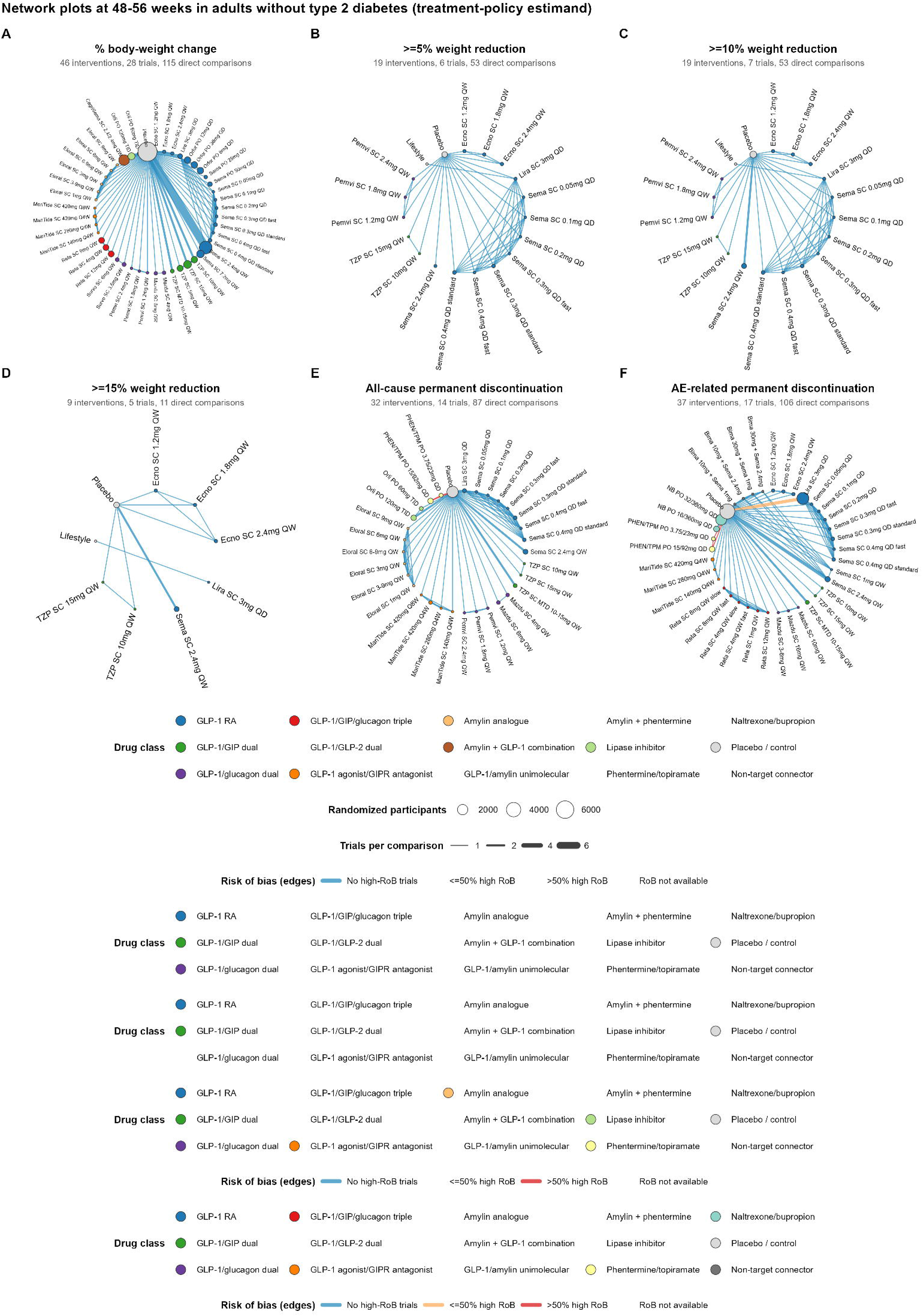
Network plots of the six key outcomes at 48-56 weeks in adults without type 2 diabetes. Each node is an intervention (drug, dose and regimen), sized by the number of randomized participants and coloured by drug class; each line is a direct comparison, with width proportional to the number of trials and colour showing the proportion of trials at high risk of bias. % body-weight change: 46 interventions, 28 trials; >=5% weight reduction: 19 interventions, 6 trials; >=10% weight reduction: 19 interventions, 7 trials; >=15% weight reduction: 9 interventions, 5 trials; All-cause permanent discontinuation: 32 interventions, 14 trials; AE-related permanent discontinuation: 37 interventions, 17 trials. Abbreviations are listed in Supplementary Table S1.

Of the 47 one-year units without diabetes, only 14 reported usable all-cause permanent discontinuation at 48–56 weeks; 12 reported it at 60–80 weeks and 21 provided no extractable data. Only seven reported usable achievement of at least 10% weight reduction, and no unit in adults with diabetes reported usable all-cause permanent discontinuation in this window.

### Weight reduction at 52 weeks

In adults without diabetes, placebo-adjusted 52-week weight reductions were largest with retatrutide 12 mg (−19·80 percentage points, 95% CrI −21·81 to −17·78)^38^ and 9 mg (−18·64, −20·62 to −16·67), followed by tirzepatide 15 mg (−16·87, −18·07 to −15·51) and cagrilintide–semaglutide 2·4/2·4 mg (−16·37, −18·10 to −14·70; figure 4A; appendix table S5). Estimates were −14·26 (−15·76 to −12·75) for semaglutide 7·2 mg and −11·60 (−12·32 to −10·89) for semaglutide 2·4 mg, with smaller reductions for liraglutide 3·0 mg (−4·20, −5·49 to −2·89) and orlistat 120 mg (−3·11, −4·23 to −1·93). Seventeen strategies had a posterior probability of at least 0·95 of exceeding a 10-percentage-point reduction, and 41 of 45 had at least 0·80 probability of exceeding 5 percentage points. Certainty was high only for tirzepatide 15 mg and semaglutide 2·4 mg, moderate for 29 strategies and low for 14. All retatrutide estimates were low certainty because their 52-week values were figure-derived and became unidentifiable when these were excluded (appendix tables S12, S23).

**Figure 4.**
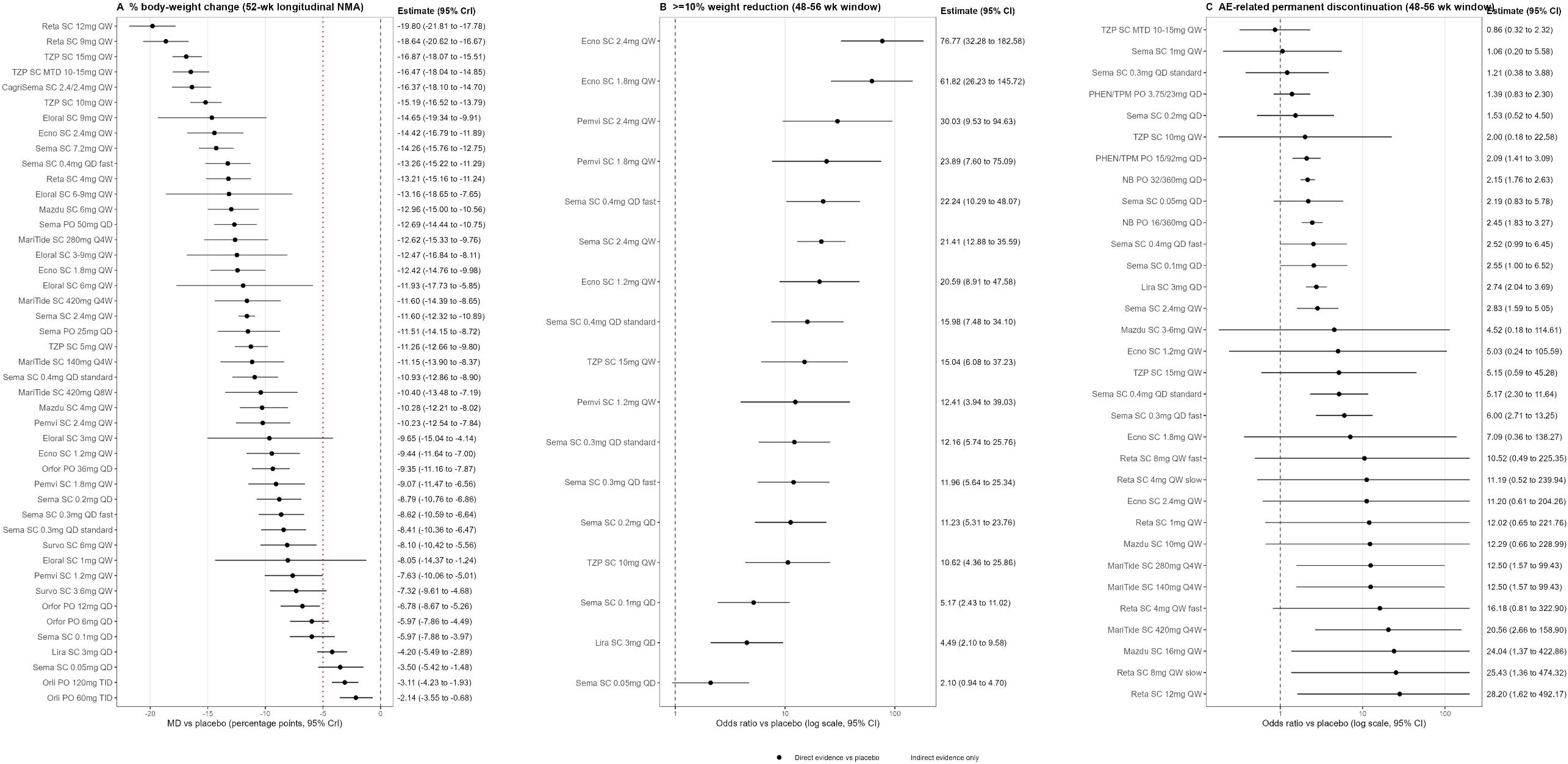
Primary efficacy and tolerability estimates versus placebo in adults without type 2 diabetes. (A) Mean difference (95% CrI) in % body-weight change at 52 weeks from the primary Bayesian longitudinal network meta-analysis (Table S5), with the dotted red line marking an illustrative clinical benchmark of −5 percentage points (prespecified decision-sensitivity tolerances of 0.5, 1.0 and 2.0 percentage points for dose escalation are reported in Supplementary Table S16); (B) odds ratio (95% CI) for achieving ≥10% weight reduction at 48–56 weeks; and (C) odds ratio (95% CI) for adverse-event-related permanent discontinuation at 48–56 weeks across the 32 target pharmacotherapy strategies versus placebo (excluding the four non-target connector arms shown in the 37-node network in Figure 3F).

**Figure 5.**
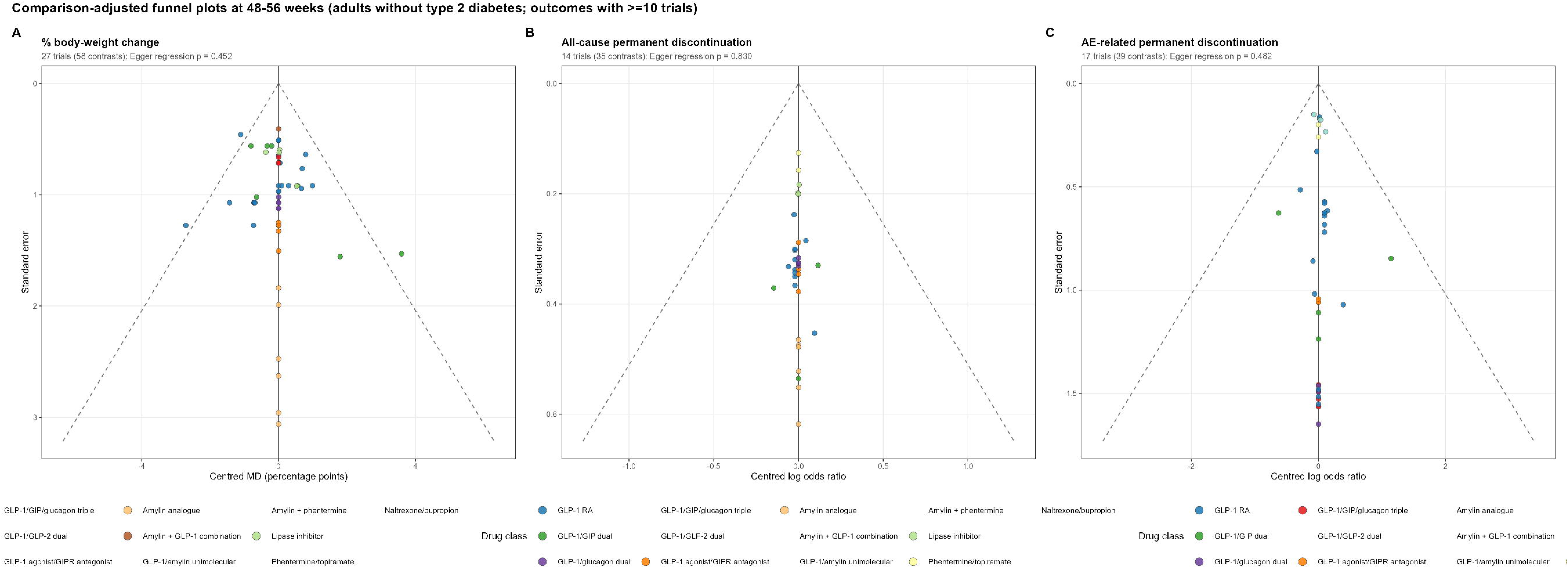
Comparison-adjusted funnel plots for drug-versus-control contrasts in outcomes with at least 10 trials at 48-56 weeks. Each point is a trial-level contrast centred on its network estimate; dashed lines show pseudo-95% confidence limits. (A) % body-weight change (27 trials): Egger regression p = 0.452; (B) All-cause permanent discontinuation (14 trials): Egger regression p = 0.830; (C) AE-related permanent discontinuation (17 trials): Egger regression p = 0.482. Funnel plots for the remaining scenarios with ≥10 trials (serious adverse events and nausea at 48–56 weeks, and AE-related discontinuation at 20–28 weeks) are presented in Supplementary Material (Table S9).

### Reference-network consistency and robustness

The 48–56-week reference network closely reproduced the longitudinal estimates (appendix tables S4, S11). Tirzepatide 15 mg produced 5·41 percentage points greater weight reduction than semaglutide 2·4 mg (95% CI 4·09–6·73) but did not differ clearly from cagrilintide–semaglutide 2·4/2·4 mg. Heterogeneity was moderate (τ=0·68 percentage points; I²=45·5%) and global inconsistency was not significant (p=0·062), but node-splitting identified local disagreement for liraglutide 3·0 mg versus placebo and tirzepatide 10 mg versus 5 mg (appendix tables S6, S7). Background energy deficit, age, sex distribution and titration duration differed across comparisons (appendix table S10).

Results were generally robust to alternative eligibility, covariance and time-window assumptions, although excluding figure-derived values also substantially increased uncertainty for some other strategies. In leave-one-parent-trial-out analyses, the largest change was 2·97 percentage points for tirzepatide 10 mg after excluding SURMOUNT-1;^4^ all other changes were 0·65 percentage points or less (appendix tables S12, S13). Hypothetical or trial-product estimands generally yielded larger weight reductions than treatment-policy-compatible estimates.

### Incremental effects of dose escalation

Of 22 adjacent maintenance-dose steps in adults without diabetes, nine had a posterior probability of at least 0·80 of adding more than 1 percentage point of weight reduction (appendix table S16). Five had at least 0·80 probability of adding more than 2 percentage points: retatrutide 4–9 mg (−5·42 percentage points, 95% CrI −6·70 to −4·14), tirzepatide 5–10 mg (−3·92, −4·97 to −2·87), mazdutide 4–6 mg (−2·67, −4·07 to −1·27), semaglutide 2·4–7·2 mg (−2·67, −4·14 to −1·21) and orforglipron 12–36 mg (−2·59, −3·57 to −1·60). None of these significantly increased adverse-event-related permanent discontinuation, although estimates were imprecise for tirzepatide (odds ratio 1·60, 95% CI 0·98–2·60) and retatrutide (1·63, 0·96–2·75) and unavailable for mazdutide.

Other steps added less weight reduction. Increasing tirzepatide from 10 mg to 15 mg added 1·67 percentage points (95% CrI 0·69–2·66), with only 0·26 probability of exceeding 2 points, and its adverse-event-related discontinuation estimate was highly uncertain (odds ratio 2·58, 95% CI 0·48–13·74). Increasing retatrutide from 9 mg to 12 mg added 1·16 percentage points (−0·15 to 2·47; based on figure-derived values) but increased adverse-event-related discontinuation over longer-term follow-up (odds ratio 1·84, 95% CI 1·21–2·79 at 80 weeks in TRIUMPH-1; 48 additional events per 1000 participants, 13–98; appendix table S16). Similarly, increasing orforglipron from 6 mg to 12 mg added 0·79 percentage points (−0·20 to 1·78) while increasing adverse-event-related discontinuation over 72 weeks (1·54, 1·01–2·35; 26 additional events per 1000, 0–63). Maridebart cafraglutide 420 mg every 4 weeks added no demonstrable weight reduction over 280 mg, whichever randomised 420 mg titration arm was used.

### Effects of titration schedules

Only eight randomised titration contrasts reaching the same maintenance dose were available at 48–56 weeks, from four trials (six treatment-policy, two trial-product; appendix table S19). Faster (2-week rather than 4-week) escalation of daily subcutaneous semaglutide to 0·4 mg produced greater weight reduction (−2·45 percentage points, 95% CI −4·77 to −0·13), with lower but inconclusive estimates for both permanent-discontinuation outcomes. The other five treatment-policy contrasts (semaglutide 0·3 mg, eloralintide 9 mg and three maridebart cafraglutide 420 mg schedules) showed no clear differences in weight reduction, and the two retatrutide trial-product contrasts were also inconclusive^39^.

### Incremental effects of lifestyle support

Seven outcome records from three trials compared additional lifestyle support under identical pharmacotherapy (appendix table S18). In a 12-week naltrexone–bupropion trial, adding a structured commercial behavioural programme increased weight reduction (mean difference −1·64 percentage points, 95% CI −3·24 to −0·04; LOCF analysis).^40^ Two trials of additional behavioural support with orlistat, one with 52-week data, found no clear weight benefit. No trial randomised support intensity under an identical incretin-based regimen or provided a 52-week treatment-policy estimate of an incremental support effect.

Among 51 units with descriptive support data, 41 prescribed a 500–600 kcal per day energy deficit with 150 min per week of physical activity, and only three provided intensive behavioural programmes (≥20 contacts per year; appendix table S20).

### Permanent discontinuation and safety

At 48–56 weeks in adults without diabetes, no target strategy significantly increased all-cause permanent discontinuation versus placebo, and ten of 32 target strategies significantly reduced it (appendix table S4). These included maximum-tolerated-dose tirzepatide 10–15 mg in patients with moderate-to-severe obstructive sleep apnoea (SURMOUNT-OSA; odds ratio 0·36, 95% CI 0·22–0·59; 125 vs 281 events per 1000 participants) and phentermine–topiramate 15/92 mg (0·62, 0·48–0·79). Semaglutide 2·4 mg did not differ significantly from placebo (1·04, 0·69–1·57).

By contrast, 13 of 32 target strategies significantly increased adverse-event-related permanent discontinuation (figures 3F, 4C), including semaglutide 2·4 mg (odds ratio 2·83, 95% CI 1·59–5·05; 114 vs 44 events per 1000), liraglutide 3·0 mg (2·74, 2·04–3·69; 97 vs 38 per 1000) and naltrexone–bupropion 32/360 mg (2·15, 1·76–2·63; 223 vs 118 per 1000). Three significant estimates arose from comparisons with no events in the placebo arm, including retatrutide 12 mg (28·20, 1·62–492·17), for which the observed active-arm risk was 161 per 1000 (90–272).

No strategy significantly increased serious adverse events in adults without diabetes. Gastrointestinal adverse events generally increased with dose, with particularly large estimates for vomiting with maridebart cafraglutide. In adults with diabetes, adverse-event-related discontinuation was significantly increased only with cotadutide 300 µg among the reported comparisons (9·41, 2·54–34·84), whereas semaglutide 2·4 mg reduced serious adverse events (0·53, 0·36–0·78).

### Weight-loss response without permanent discontinuation

Matched arm-level data for achieving at least 10% weight reduction without all-cause permanent discontinuation were available from only four randomisation units (appendix table S17). Fréchet–Hoeffding bounds were 0·55–0·72 for tirzepatide 15 mg (SURMOUNT-CN)^41^, 0·53–0·68 for semaglutide 2·4 mg and 0·17–0·34 for liraglutide 3·0 mg, compared with 0·002–0·10 for placebo.

### Weight reduction in adults with type 2 diabetes

In the 48–56-week reference network, placebo-adjusted reductions in adults with type 2 diabetes were −11·10 percentage points (95% CI −12·20 to −10·00) for tirzepatide 15 mg, −10·80 (−11·60 to −10·00) for cagrilintide–semaglutide 2·4/2·4 mg and −6·35 (−7·07 to −5·63) for semaglutide 2·4 mg, all smaller than in adults without diabetes (appendix table S4).

Nine trial programmes provided 18 matched dose pairs across seven molecules in separately randomised populations with and without diabetes (appendix table S14). At matched follow-up of 52–80 weeks, placebo-adjusted weight reduction with diabetes was 62% of that without diabetes (ratio 0·62, 95% CI 0·58–0·66), a relative attenuation of 37·7% (33·6–41·5). The ratio was similar with one representative dose per programme (0·63, 0·58–0·69).

## Discussion

Far less evidence informs 1-year obesity pharmacotherapy initiation decisions than the number of eligible trials suggests. Only about a quarter of randomisation units informed the primary model, and only 28% of target strategies had non-extrapolated 52-week estimates. Retatrutide, higher-dose tirzepatide and cagrilintide–semaglutide produced the largest weight reductions, but certainty was high only for tirzepatide 15 mg and semaglutide 2·4 mg.

Previous network meta-analyses mainly compared drugs or classes, often pooling final reported outcomes across different follow-up durations and estimands.^11,13,42^ By aligning treatment stage, follow-up and estimand, our evidence funnel showed that short follow-up, incompatible reporting, trial-product estimands and historical LOCF analyses accounted for much of the attrition. These exclusions reflect trial design and reporting, not ineffectiveness of the excluded drugs. The larger reductions under hypothetical and trial-product estimands should not be interpreted as causal effects of discontinuation;^17^ the treatment-policy estimand better reflects the consequences of initiating a strategy, including subsequent treatment changes. Older agents such as phentermine–topiramate and naltrexone–bupropion lacked treatment-policy-compatible 1-year weight evidence, as historical registration trials relied on last-observation-carried-forward estimands. Because modelling cannot replace missing evidence, rankings should be interpreted alongside effect sizes, certainty and tolerability.^28^ Greater weight reduction with tirzepatide than with semaglutide was consistent with their head-to-head trial,^43^ whereas tirzepatide and cagrilintide–semaglutide could not be clearly distinguished.

Adverse-event-related discontinuation reflects tolerability, whereas all-cause discontinuation also captures stopping for inadequate benefit and other reasons, so their divergence is plausible. Neither measures persistence in routine care, where cost, access and patient preferences influence continuation,^10,44,45^ and weight regain after cessation underscores the importance of persistence.^8,9,46^ Yet fewer than a third of eligible 1-year units without diabetes, and none with diabetes, reported usable all-cause permanent discontinuation at 48–56 weeks. Our Fréchet–Hoeffding bounds drew on only four units, did not incorporate sampling uncertainty and cannot rank strategies.

Marginal benefits of dose escalation differed substantially: whereas five adjacent steps had high probability of adding more than 2 percentage points, retatrutide 9–12 mg and orforglipron 6–12 mg added little weight reduction while increasing adverse-event-related discontinuation. These exploratory findings suggest that the highest tested maintenance dose need not offer the most favourable benefit–tolerability balance. Lower all-cause discontinuation with maximum-tolerated-dose tirzepatide cannot be attributed to dosing flexibility, given differences between trial programmes.^47^ The dose-step thresholds were decision aids, not non-inferiority margins, and individual preferences remain central to judging acceptable trade-offs.

Evidence on how to initiate treatment was especially limited. One phase 2 comparison favouring faster escalation of daily semaglutide is hypothesis-generating,^7^ and the maridebart cafraglutide comparisons were imprecise.^6^ WHO conditionally recommends intensive behavioural therapy alongside GLP-1 treatment,^48^ yet its incremental value under incretin therapy is untested: trials providing intensive support to both groups establish drug efficacy under that background, not the benefit of support.^19^

Placebo-adjusted weight reduction in type 2 diabetes was approximately 62% of that without diabetes, consistent with comparisons across the STEP and SURMOUNT programmes.^3,4,20,21^ However, comparisons between separately randomised populations cannot identify individual-level effect modification or distinguish physiological explanations from differences in baseline characteristics and concomitant treatment, and only two programmes provided comparisons matched at 52 weeks. Expected weight reduction should therefore be considered separately by diabetes status, alongside glycaemic and cardiometabolic benefits.

Strengths of this review include the treatment-initiation focus, explicit estimand classification, trial-family linkage, preservation of actual observation times and transparent labelling of extrapolated estimates. Limitations include the post-extraction addition of exploratory analyses and post-hoc decisions, which introduce potential data-driven bias. Modelling relied on assumed covariance structures and class-level borrowing, and figure-derived values materially influenced estimates for some leading strategies. Direct evidence was sparse for some comparisons, two local inconsistencies were detected and potential effect modifiers differed across comparisons, raising transitivity concerns.^31^ Restricting analyses to treatment-policy-compatible estimates reduced coverage, binary safety outcomes were sparse and continuity corrections influenced estimates with zero-event placebo arms. Our CINeMA adaptations for longitudinal estimates have not been formally validated. Finally, trial populations were selected and closely monitored, and long-term clinical outcomes, costs and safety were not assessed.

In conclusion, the evidence supporting 1-year obesity pharmacotherapy initiation decisions is considerably narrower than the trial literature suggests. Newer multi-receptor and higher-dose incretin strategies achieve substantial weight reduction, but certainty and discontinuation trade-offs vary. Better reporting and direct randomised evidence on dose escalation, titration and lifestyle support are needed to guide treatment initiation.

These findings support interpreting 1-year efficacy together with evidence certainty and permanent discontinuation, and assessing each dose increment rather than escalating automatically. Future trials should randomise titration schedules and lifestyle-support intensity under active incretin-based treatment, particularly in type 2 diabetes. Consistent with CONSORT 2025 and the estimands framework, ^17,49^ reports should include 52-week treatment-policy estimates, tabulated interim outcomes, clearly distinguished reasons for permanent discontinuation and arm-level counts of participants achieving weight-loss thresholds without stopping treatment.

## Data Availability

All data analysed in this systematic review and network meta-analysis were extracted from published randomised controlled trials, clinical trial registries, and publicly available study reports. The extracted study-level data and supporting information are provided in the manuscript and supplementary materials. Additional details, including treatment-strategy coding rules and statistical analysis code, are available from the corresponding author upon reasonable request.

---

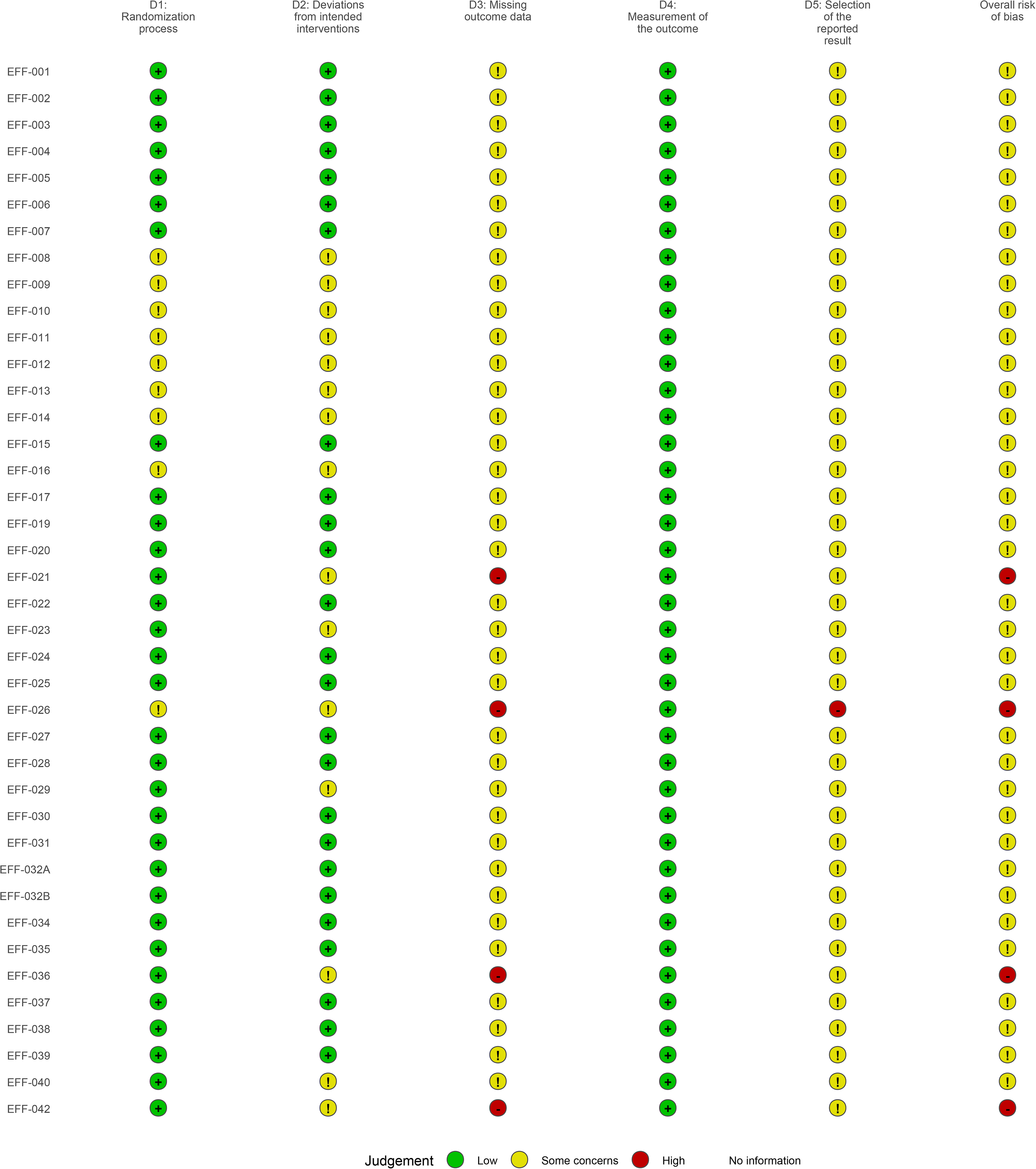

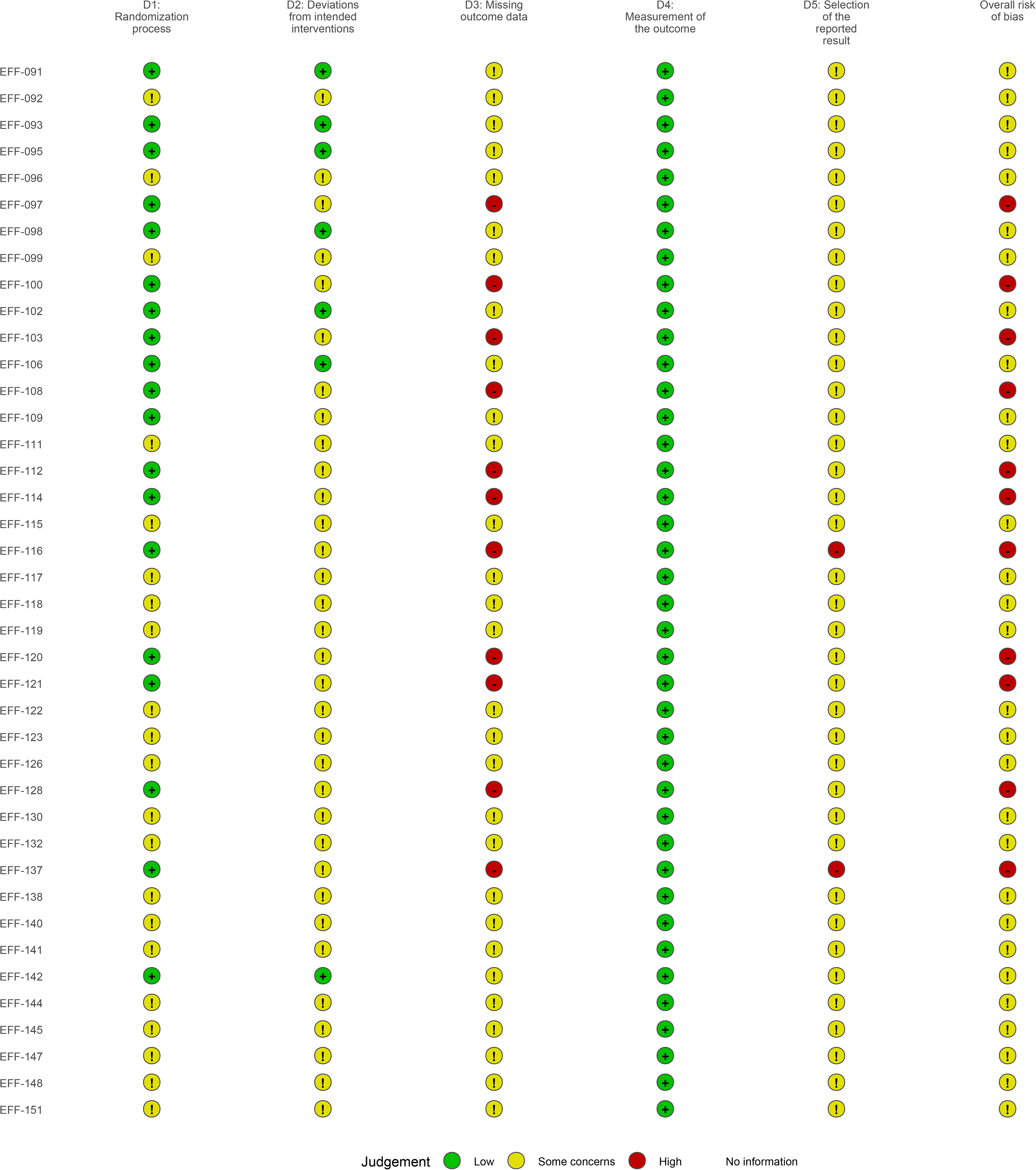

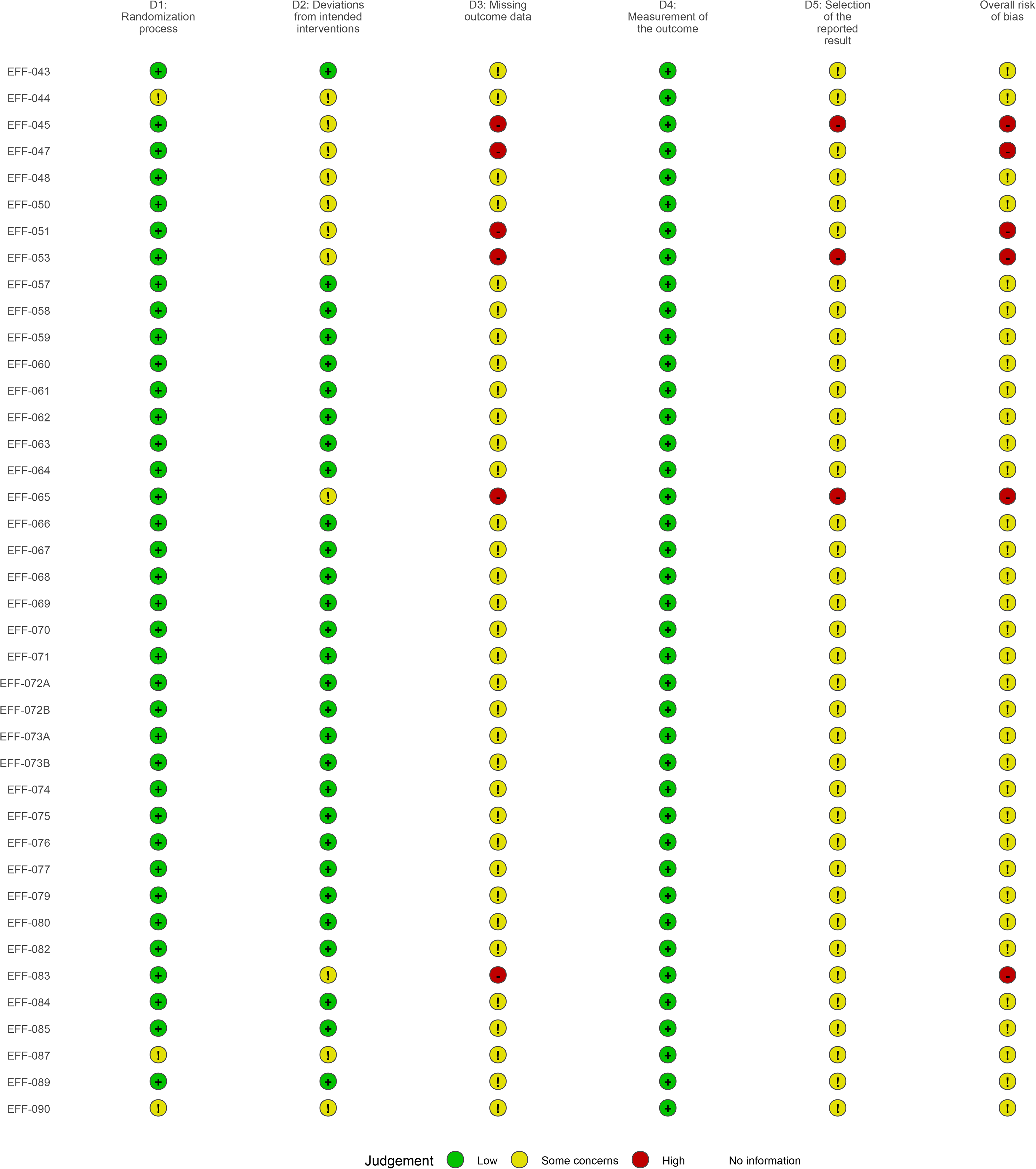

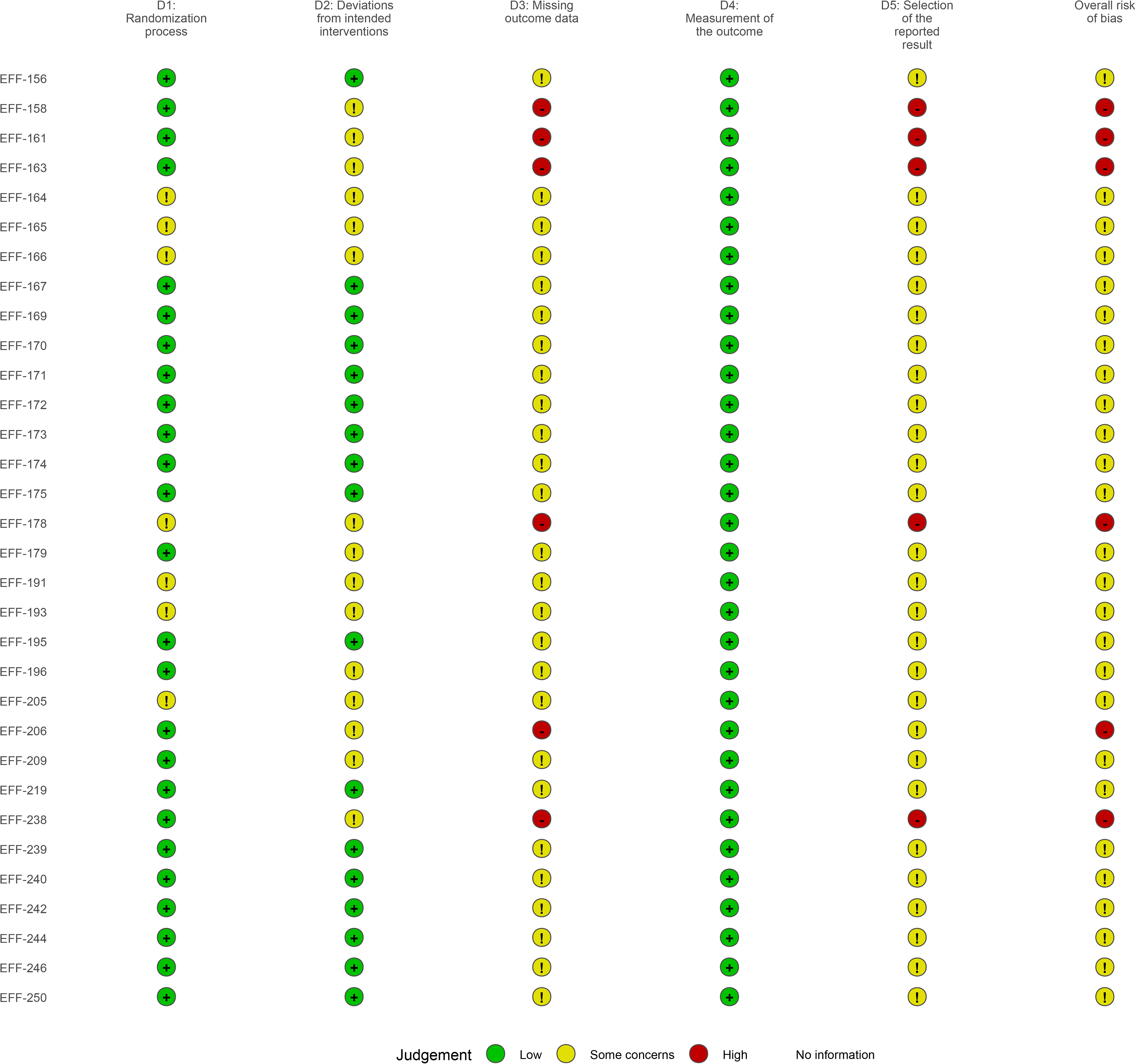

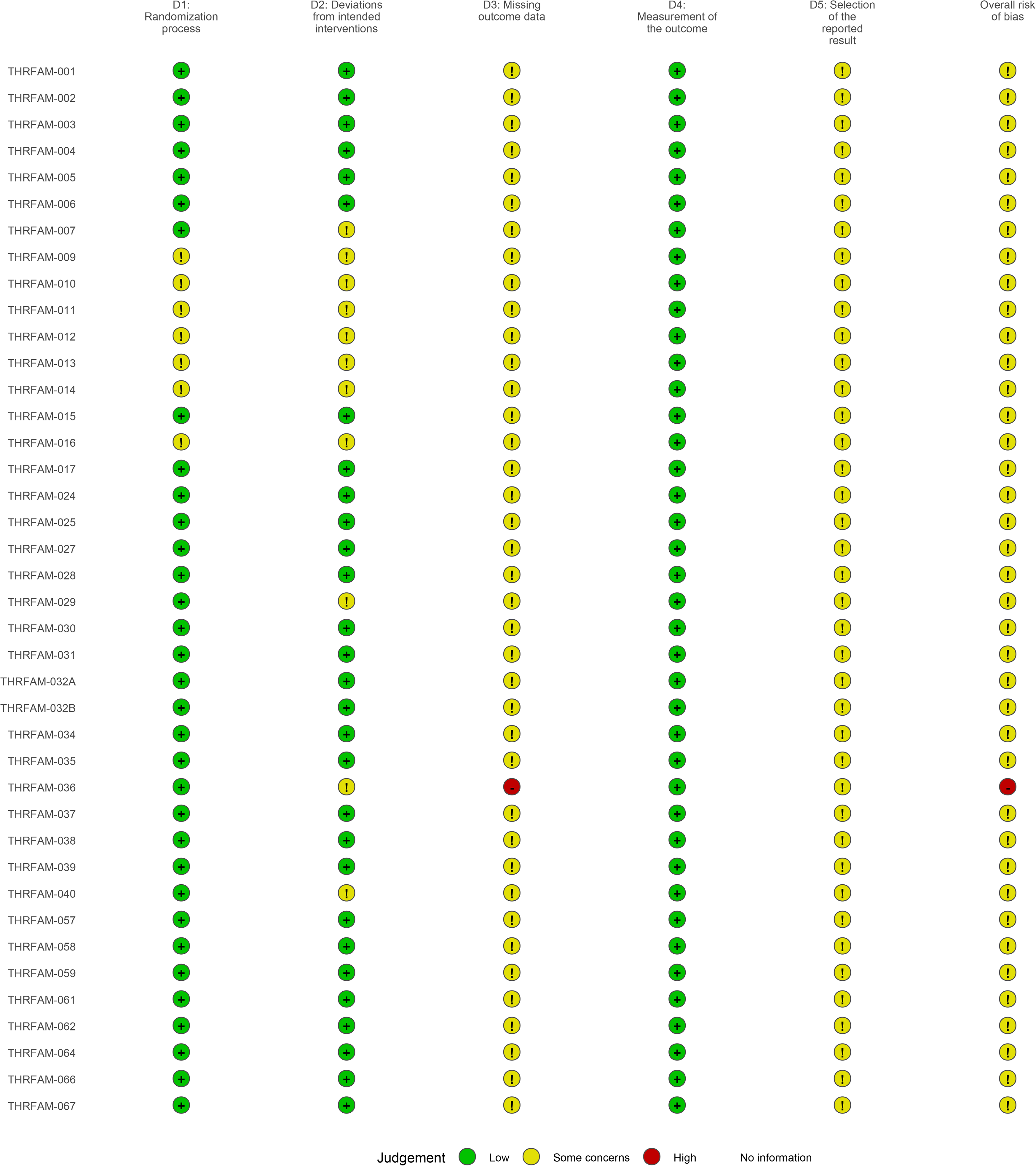

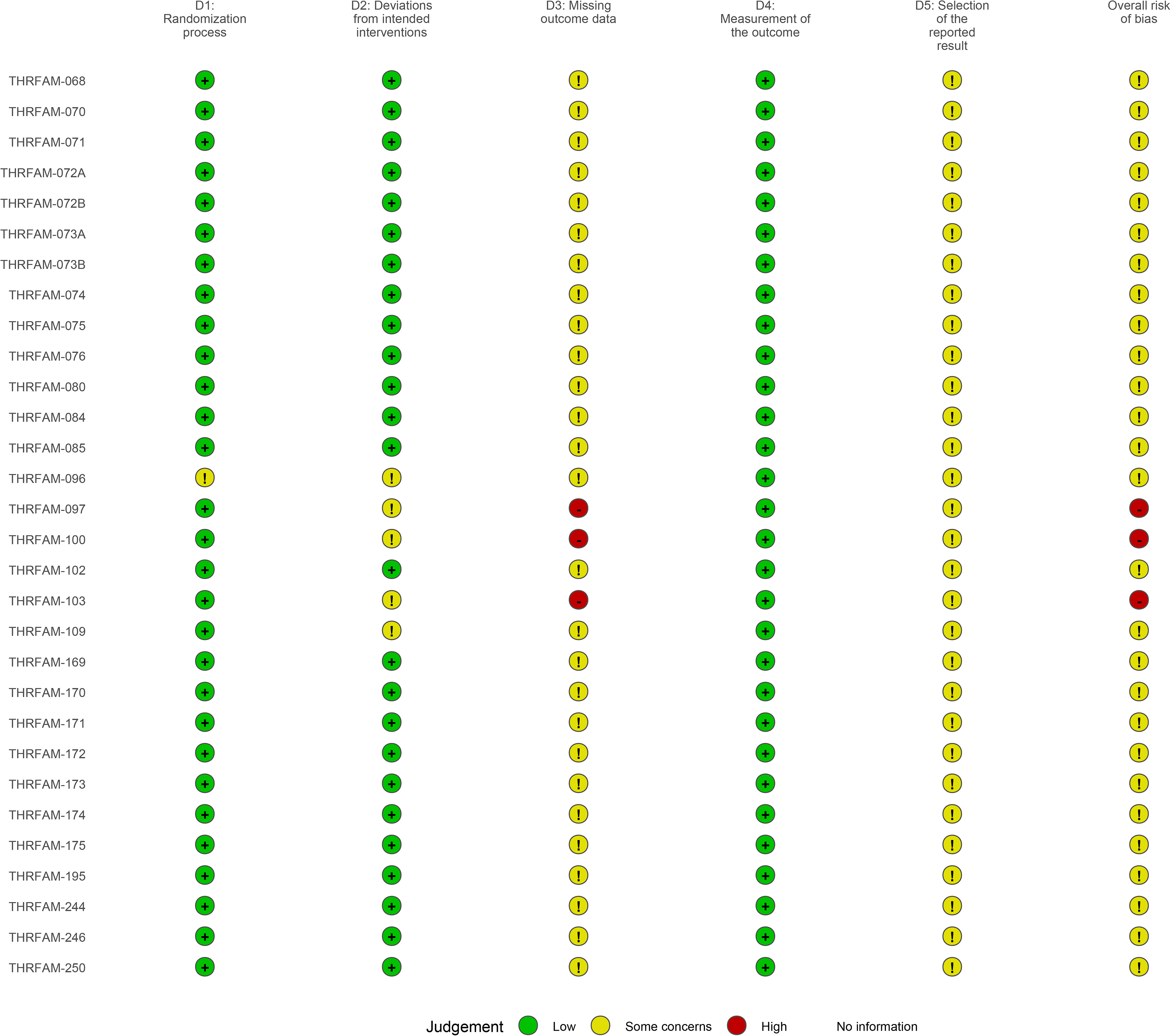

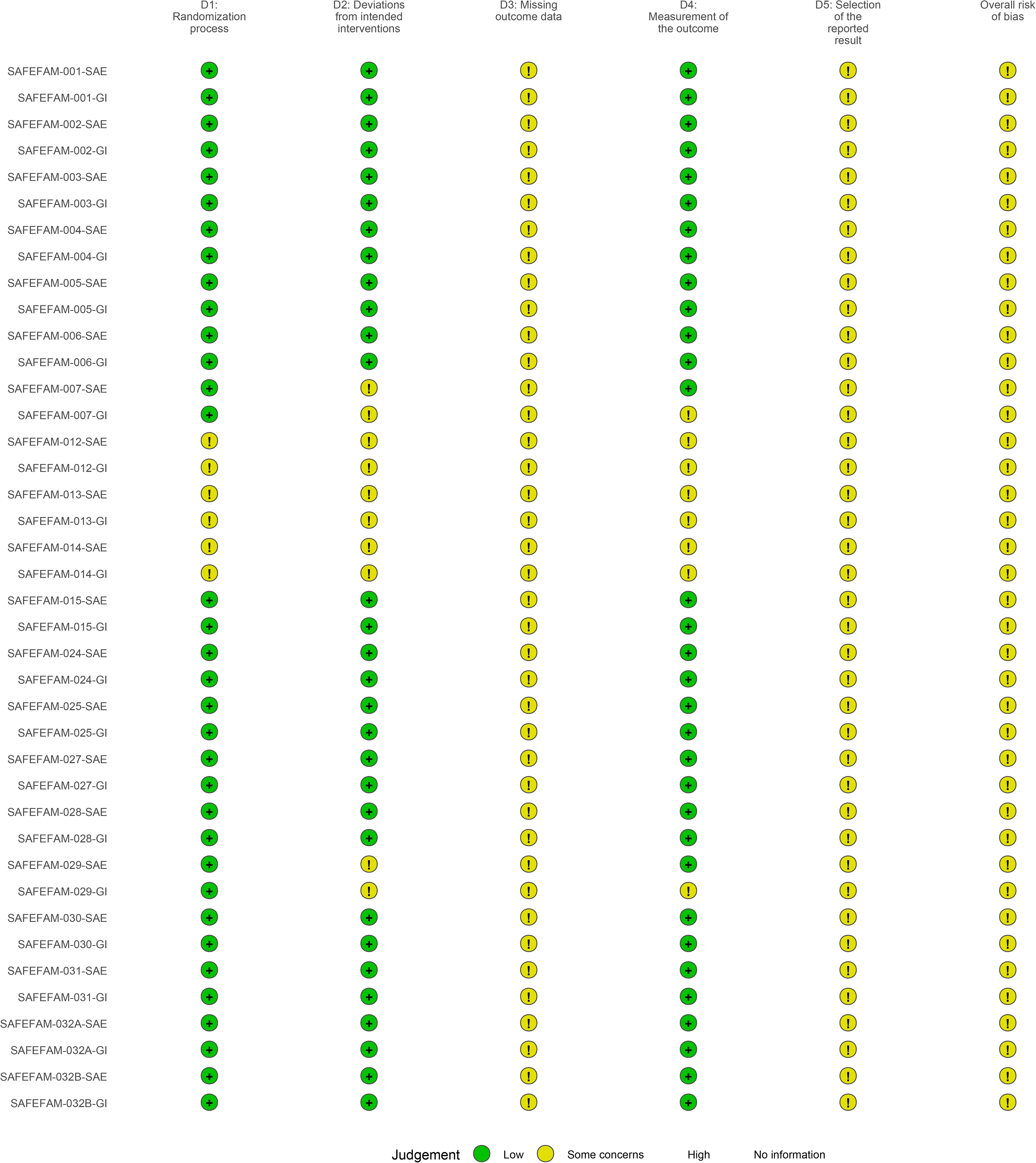

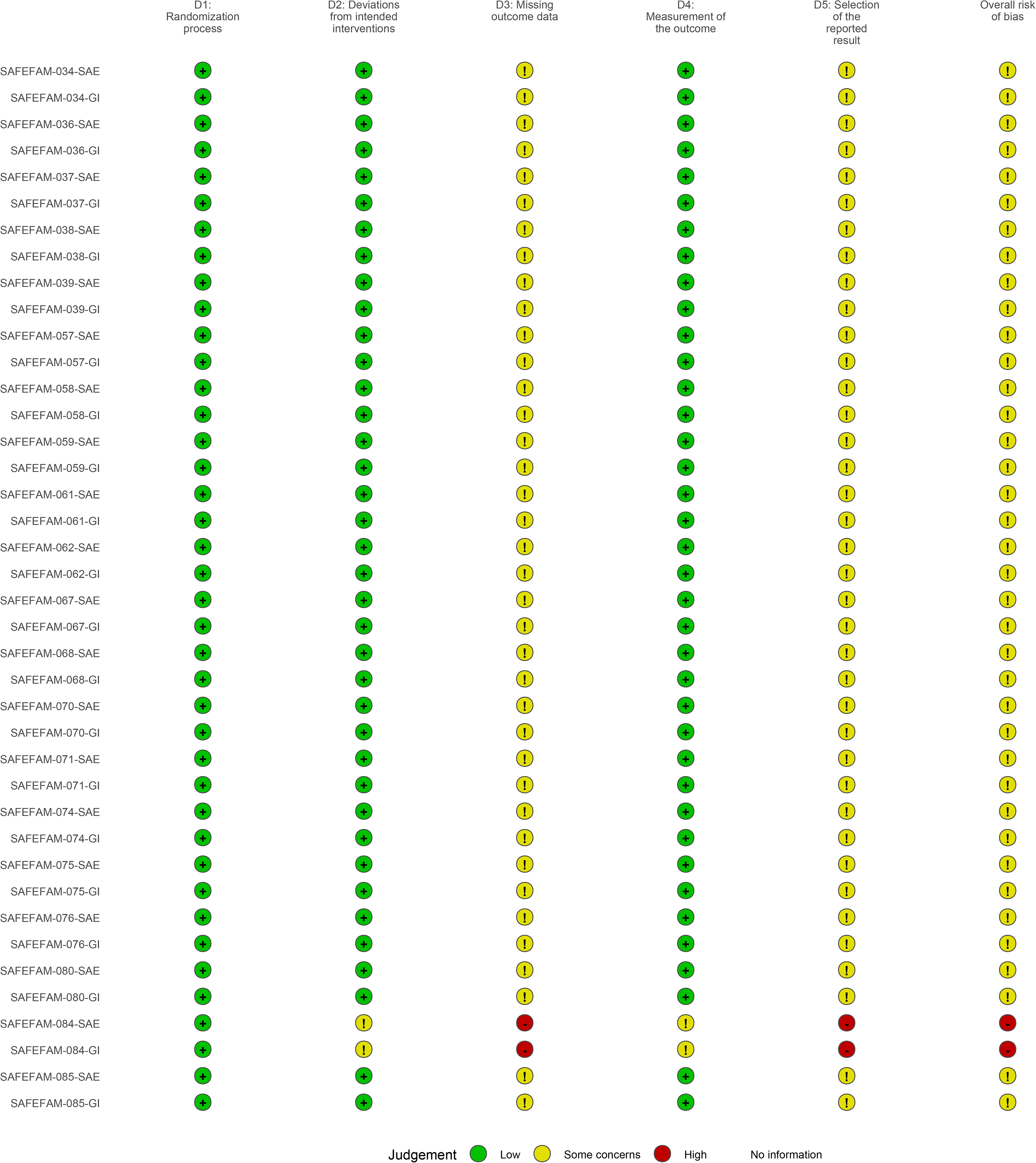

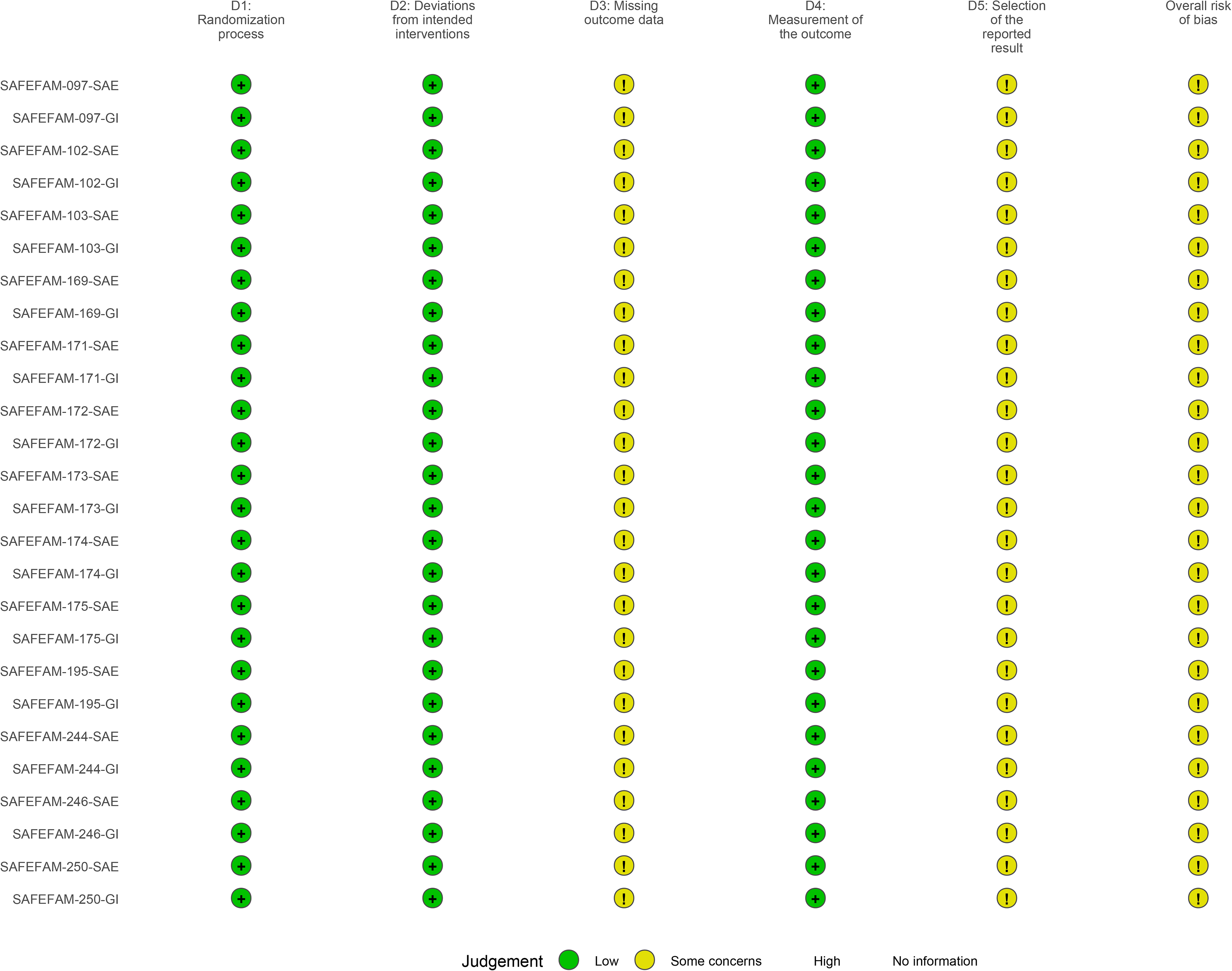

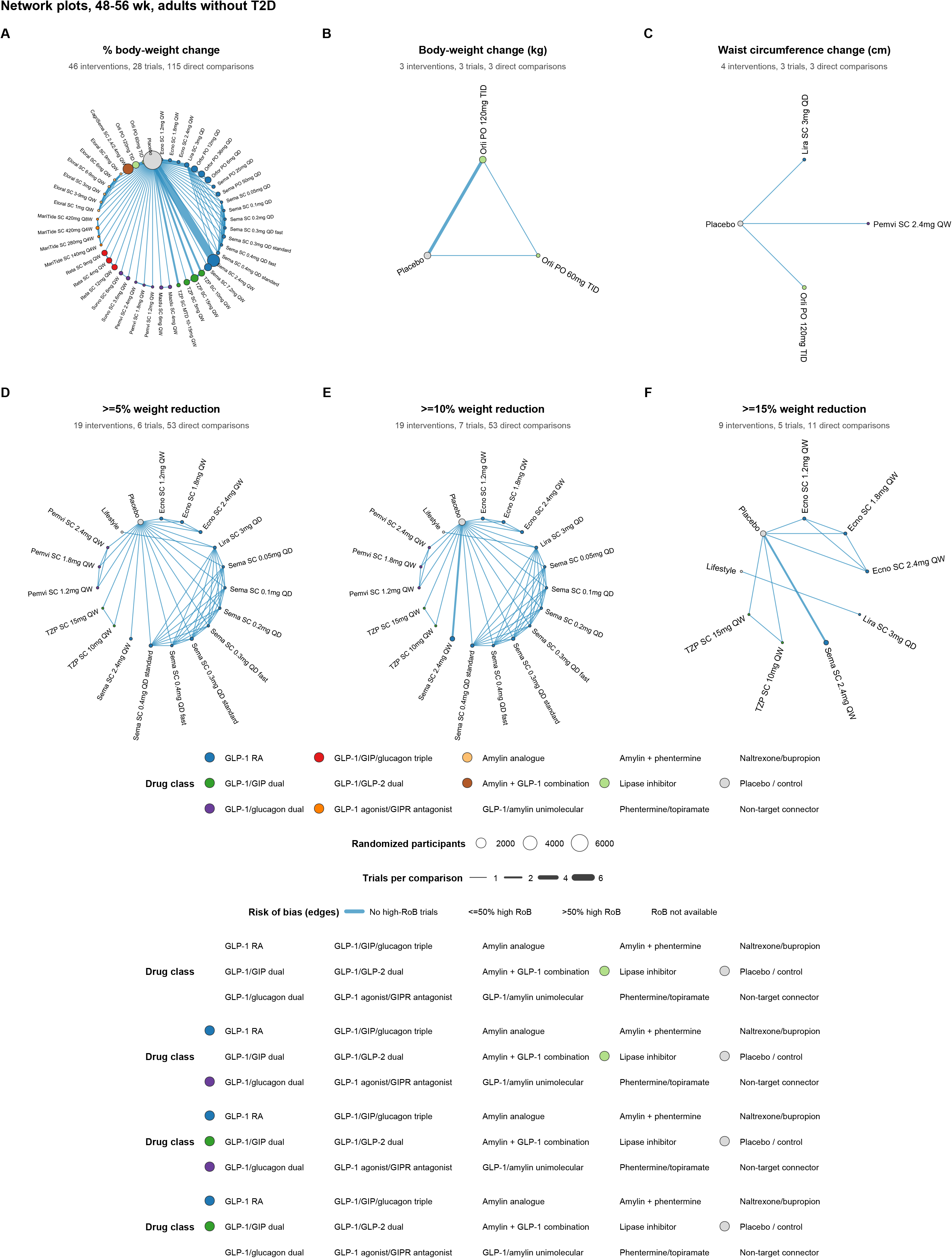

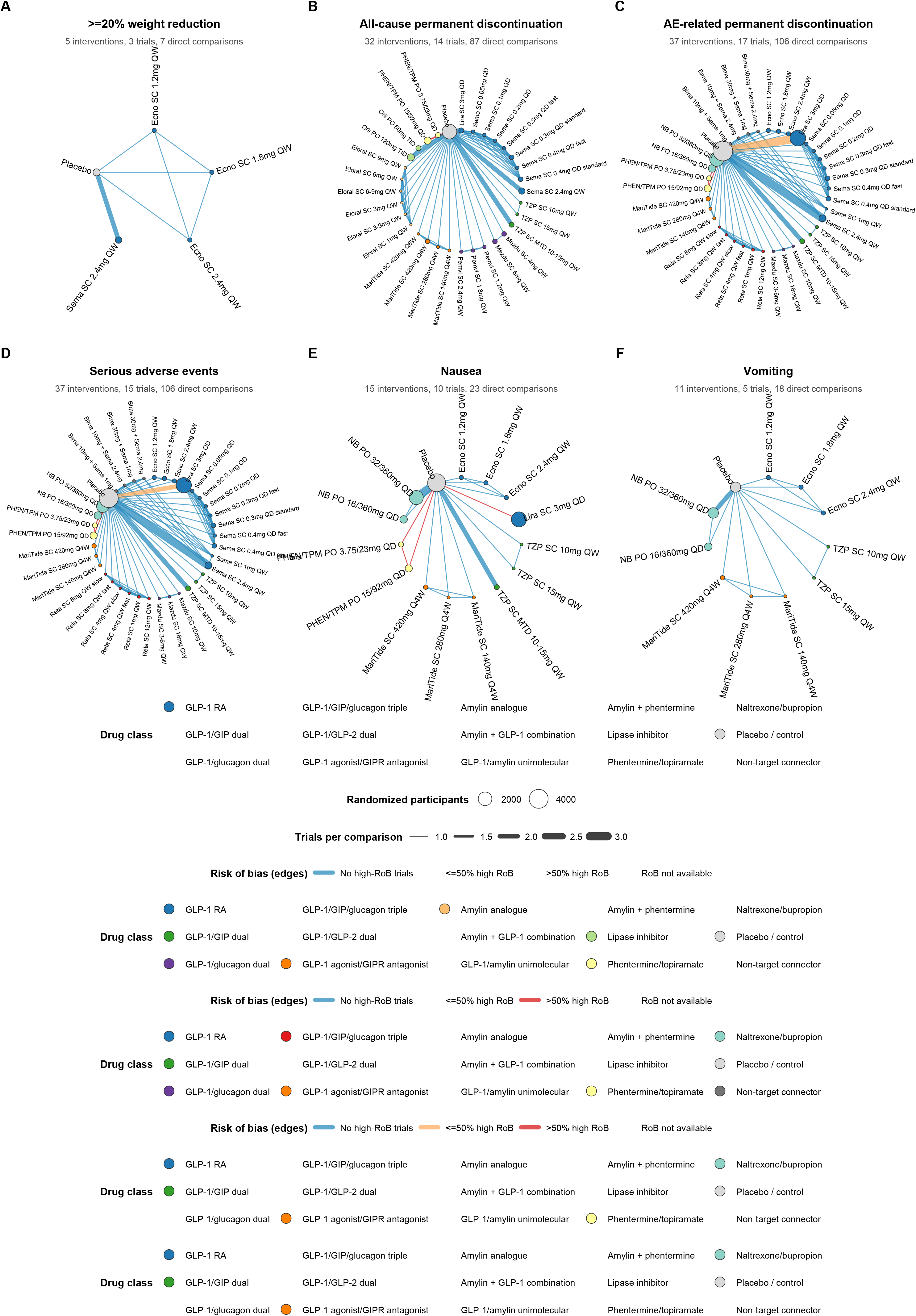

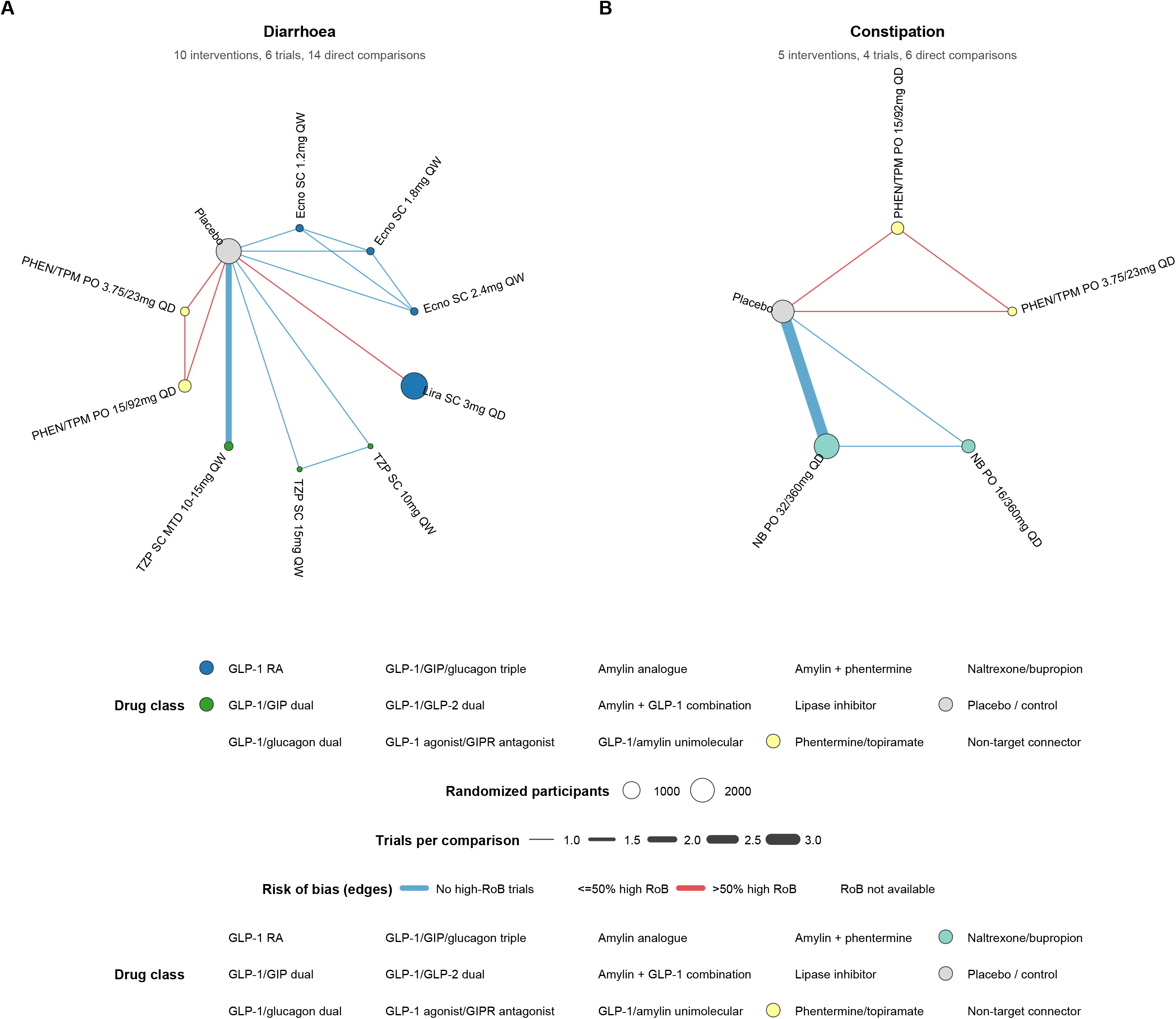

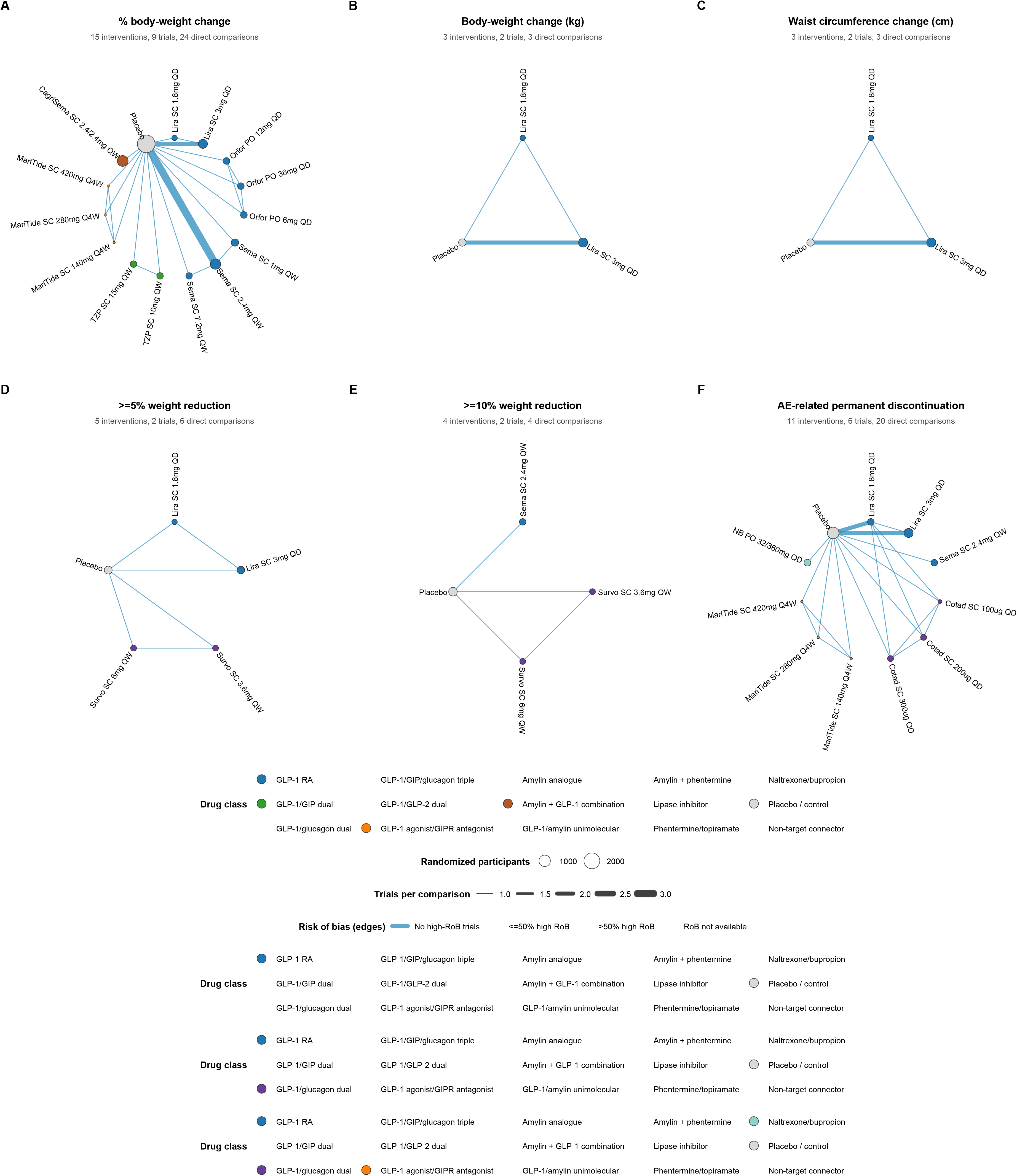

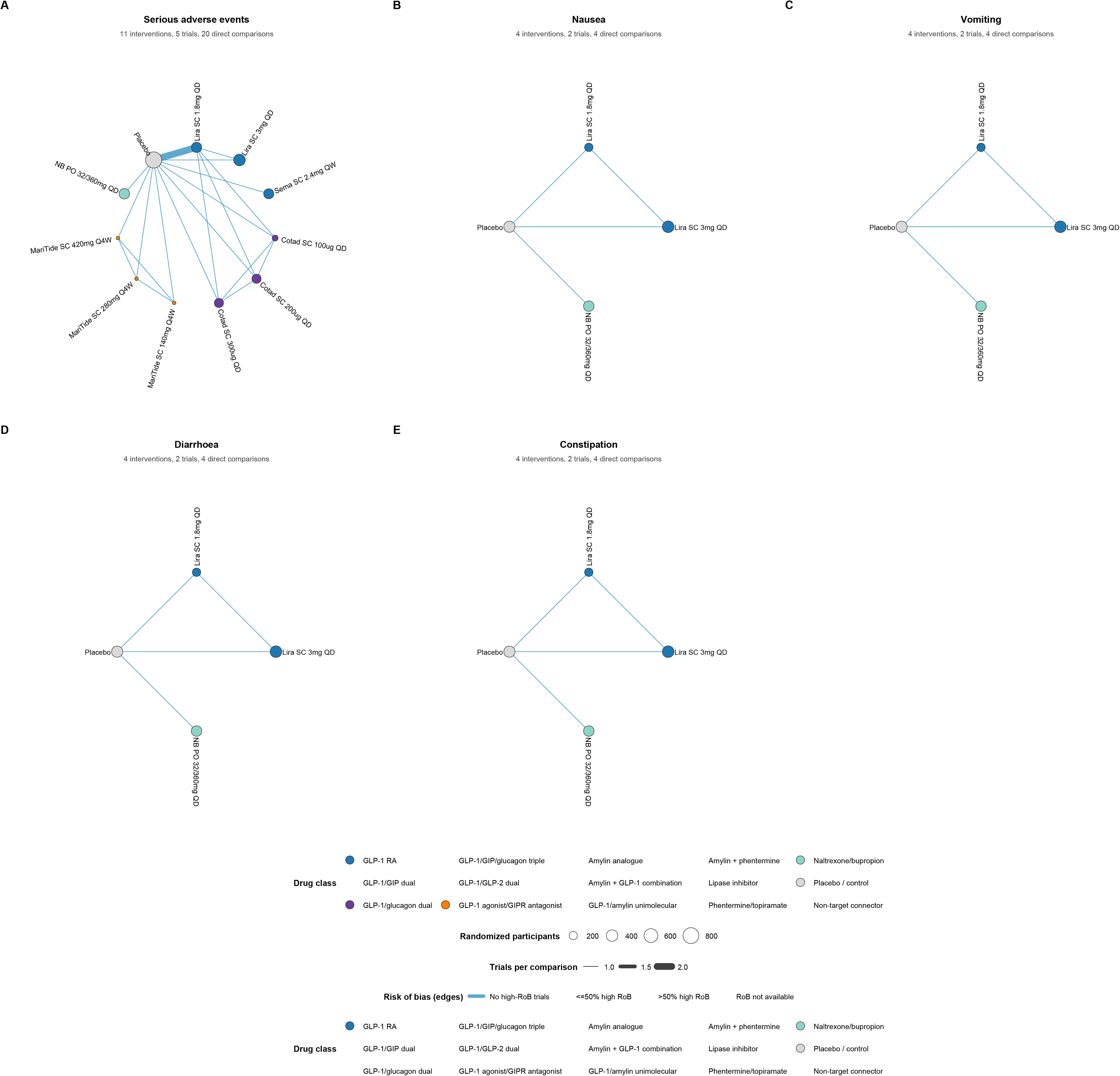

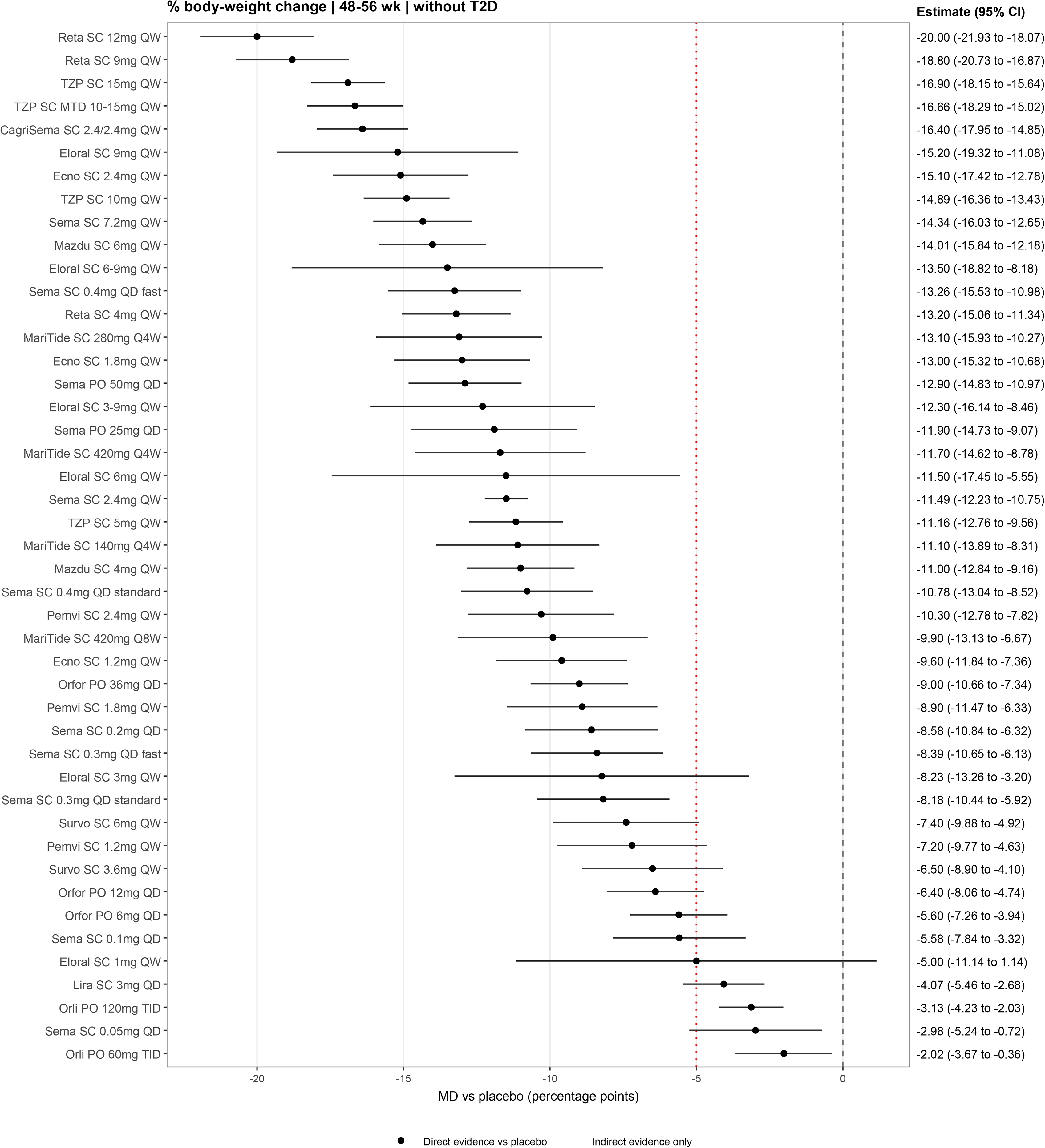

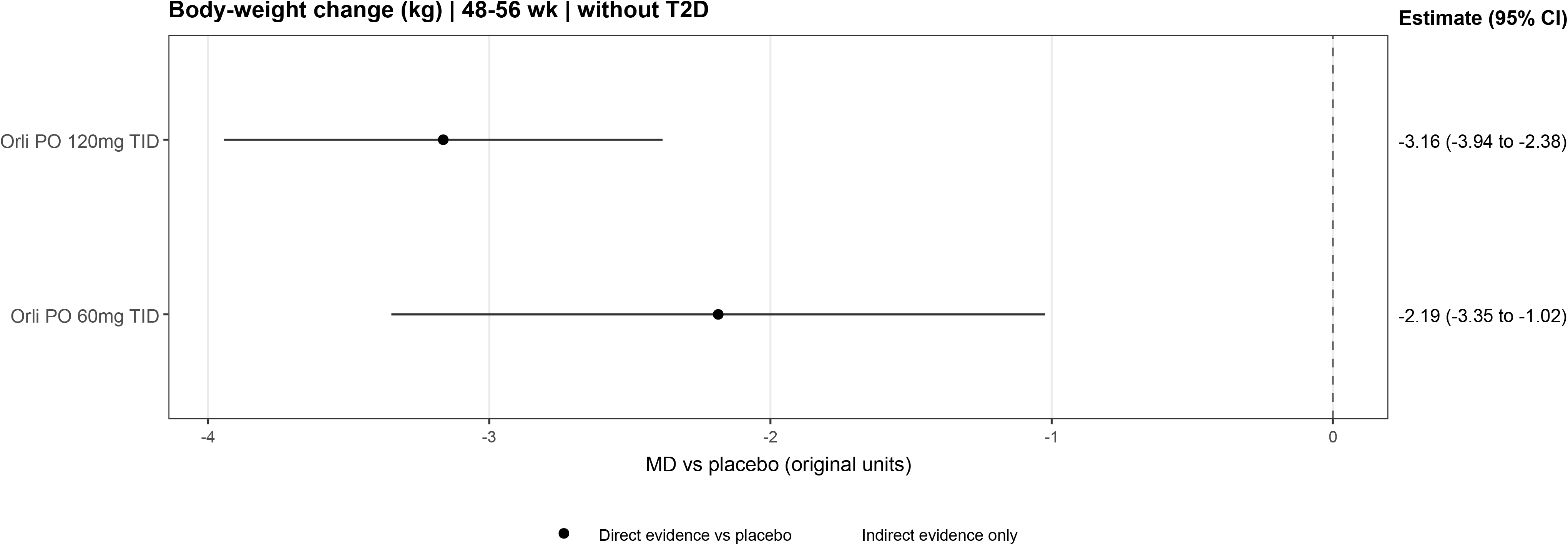

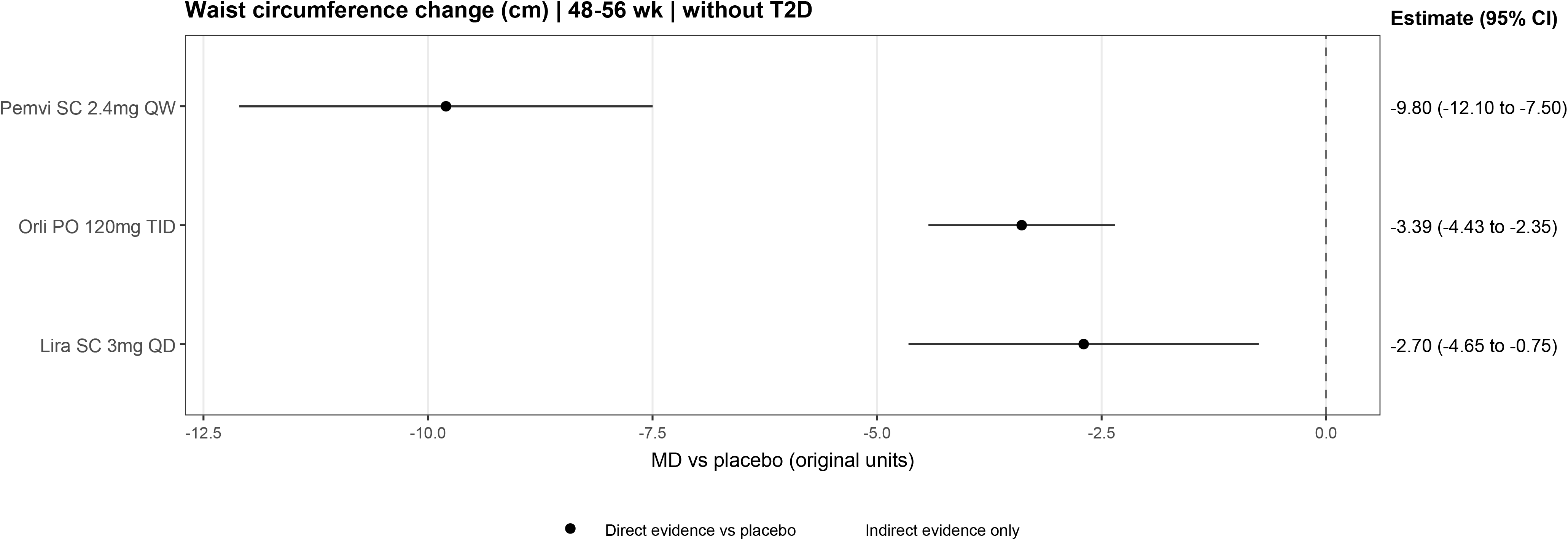

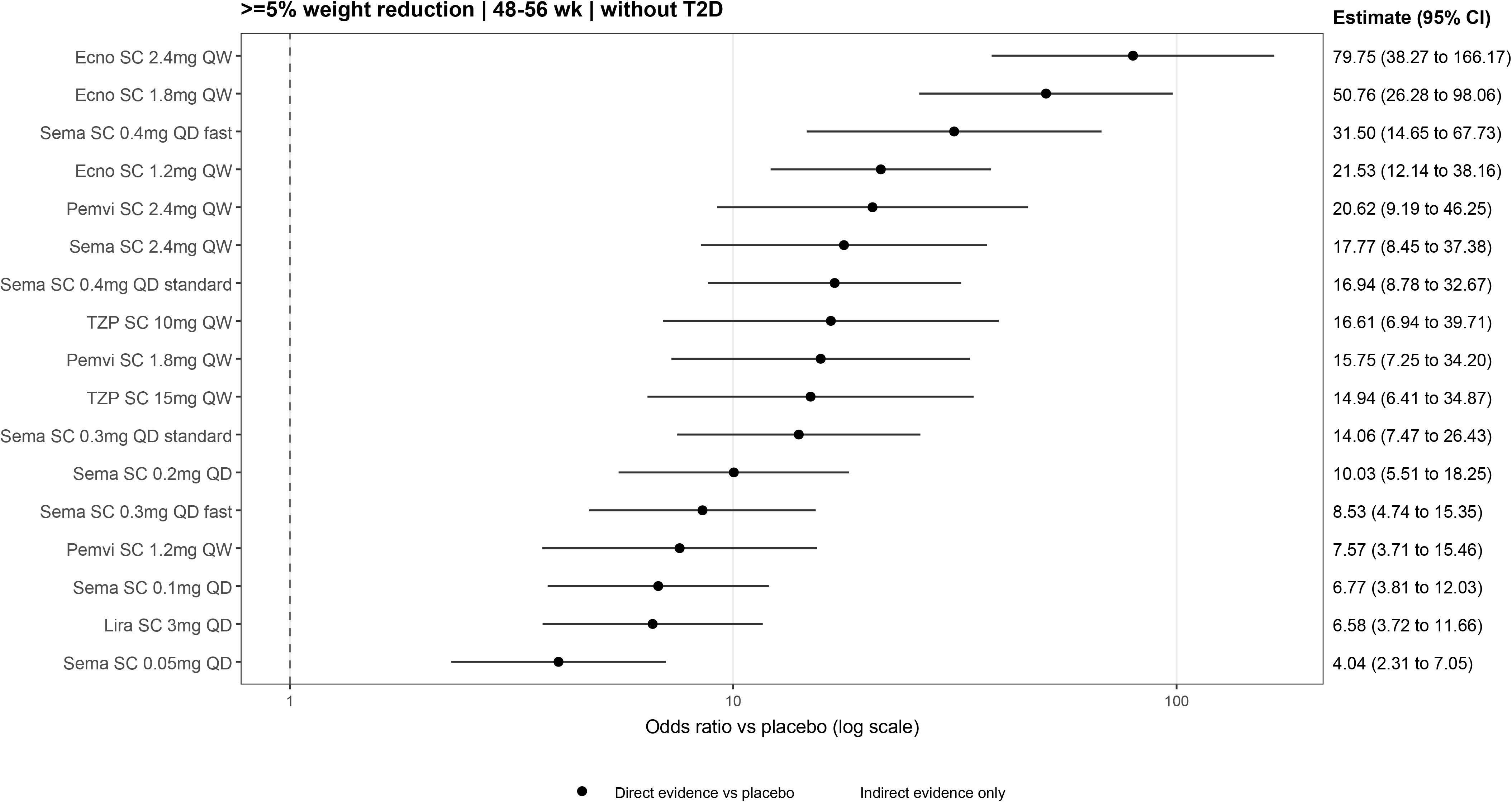

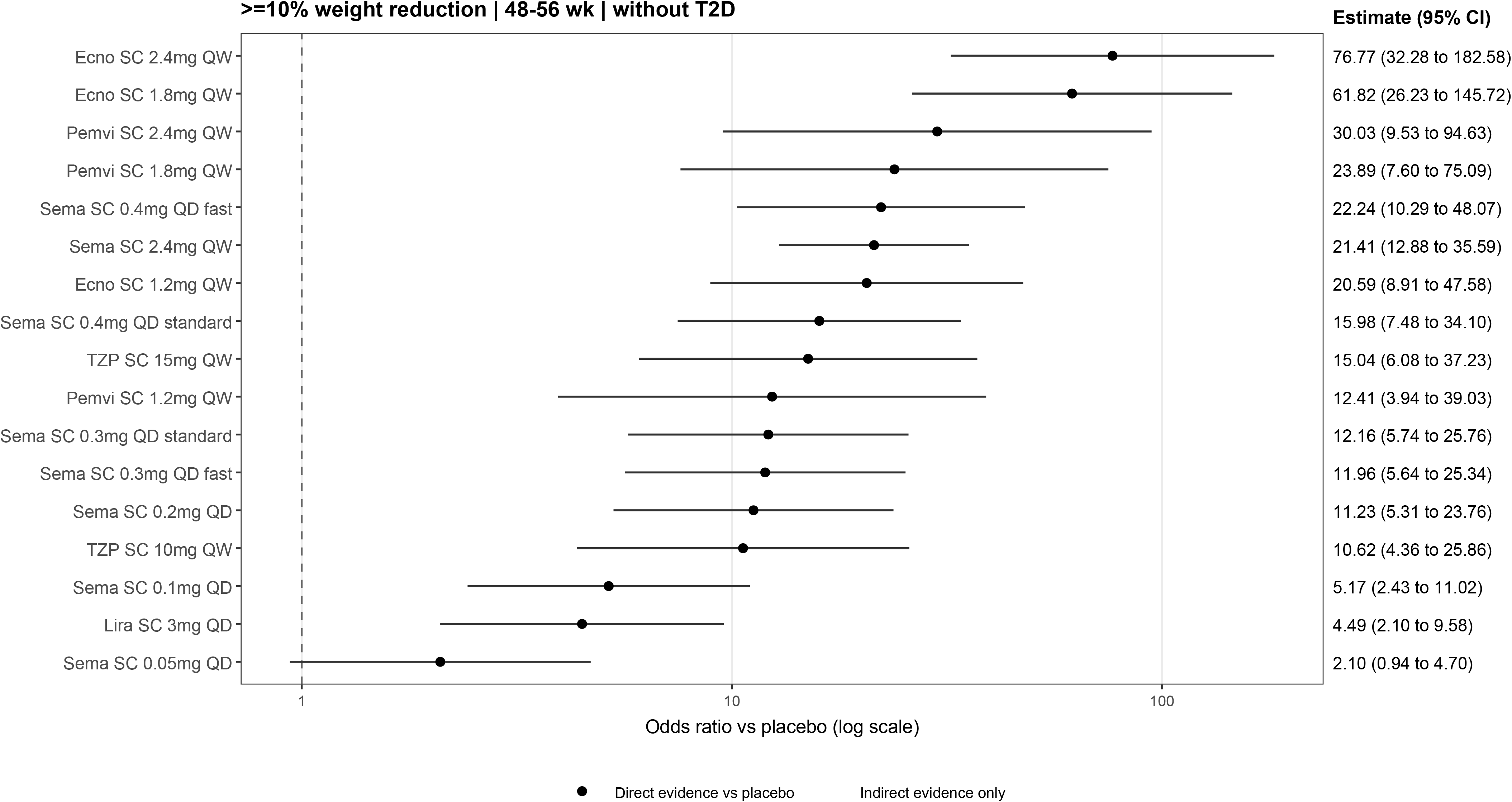

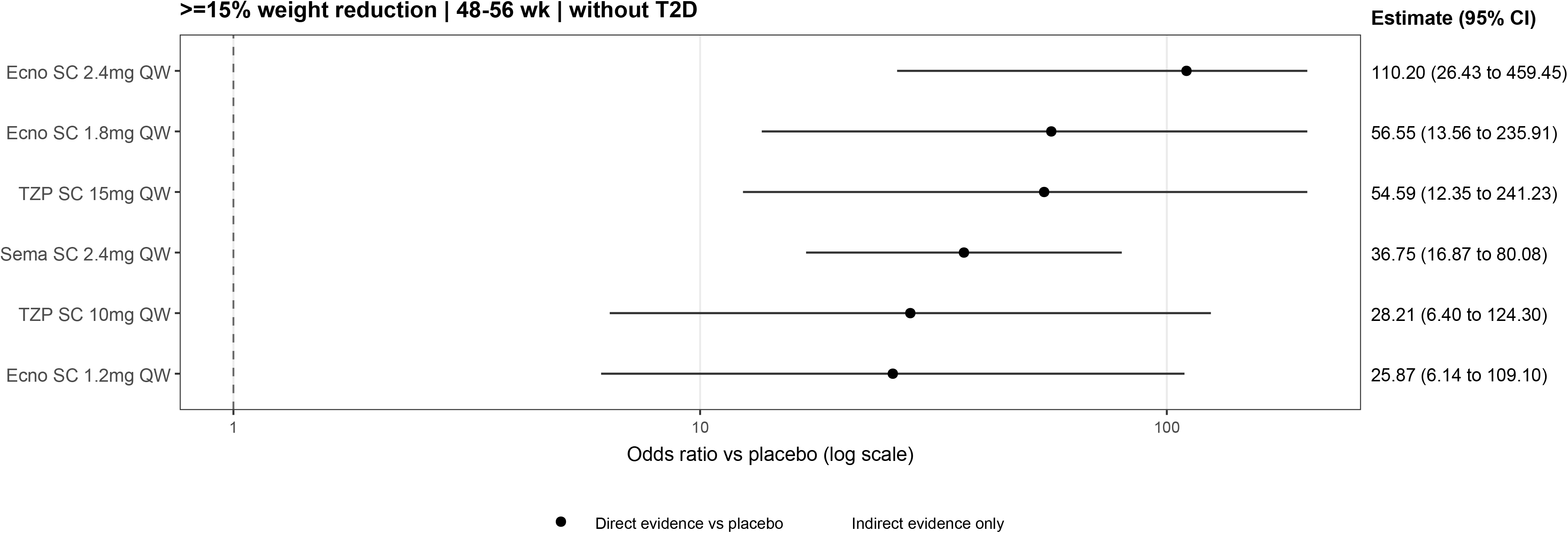

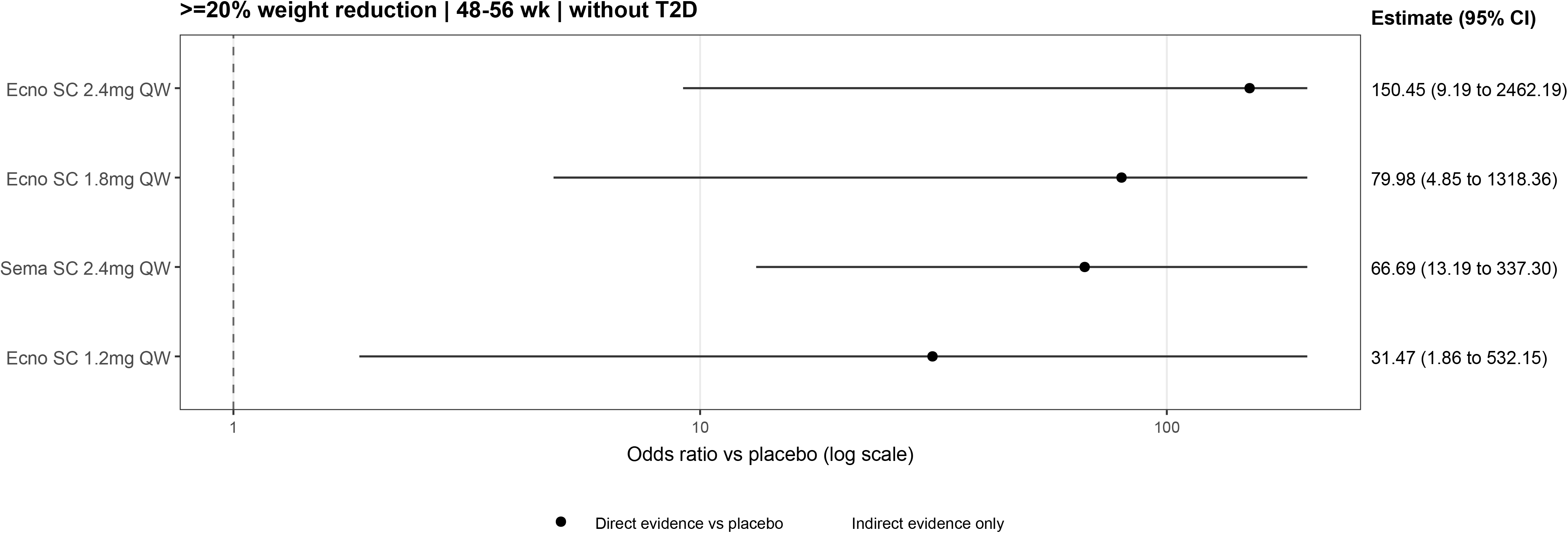

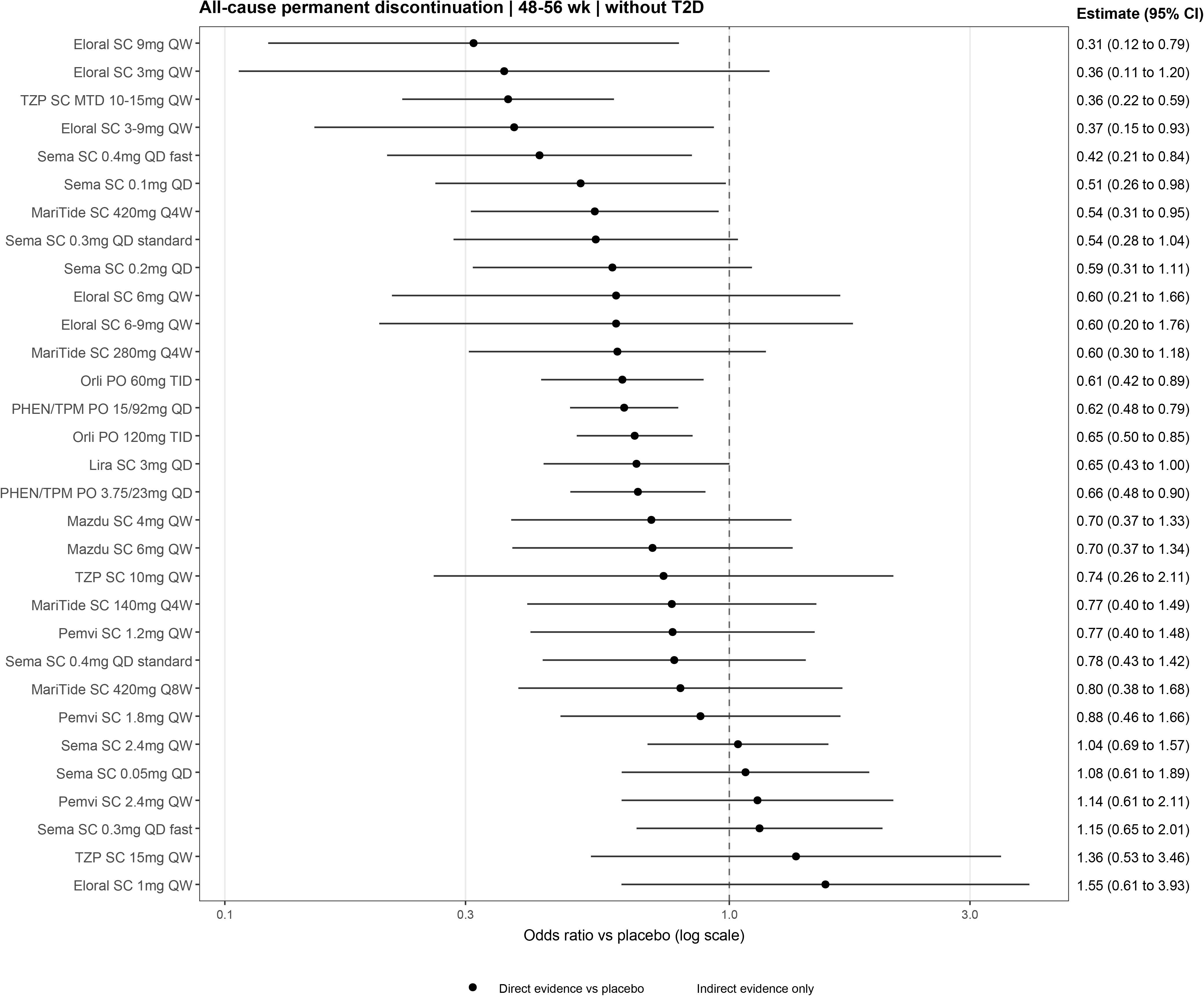

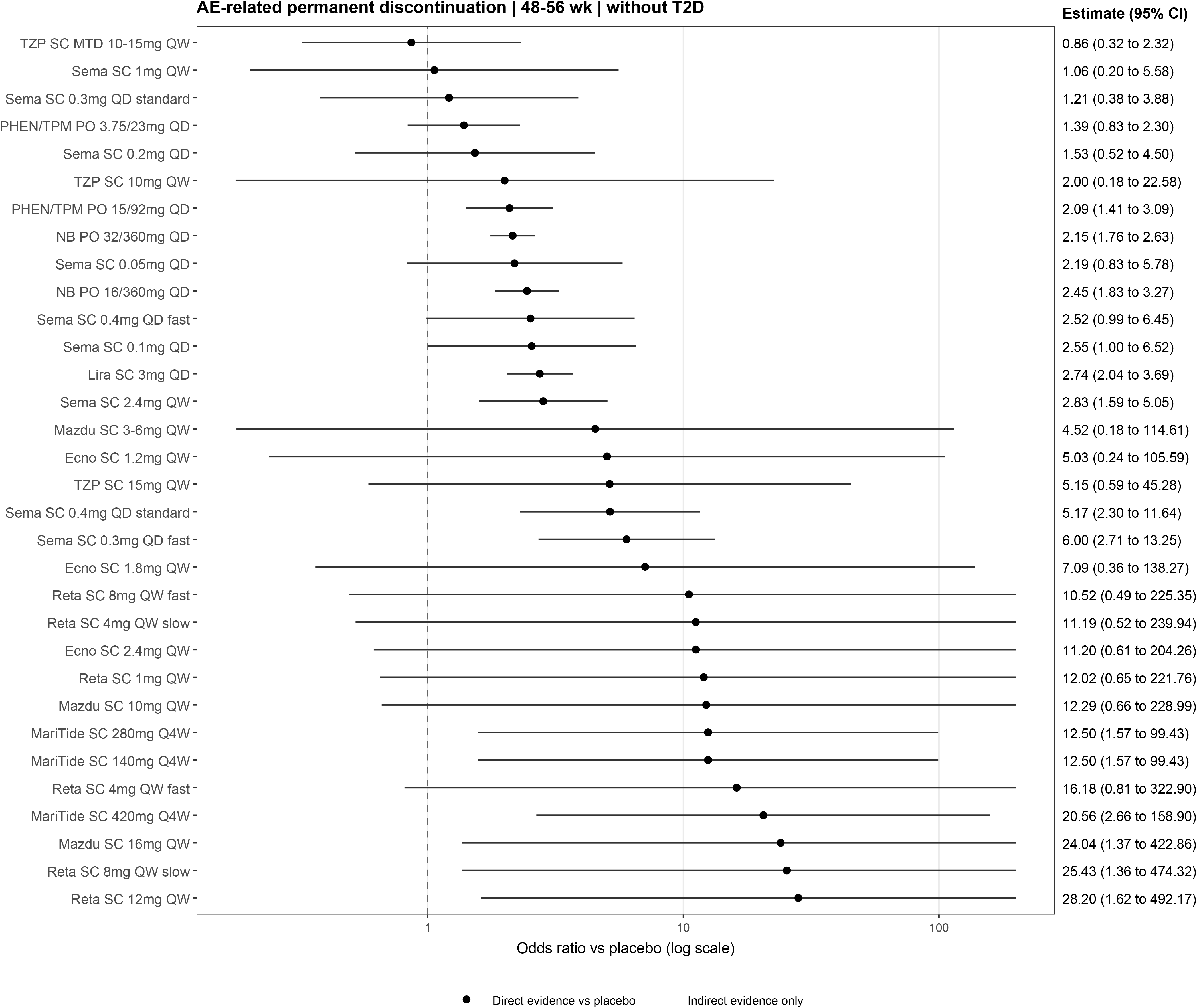

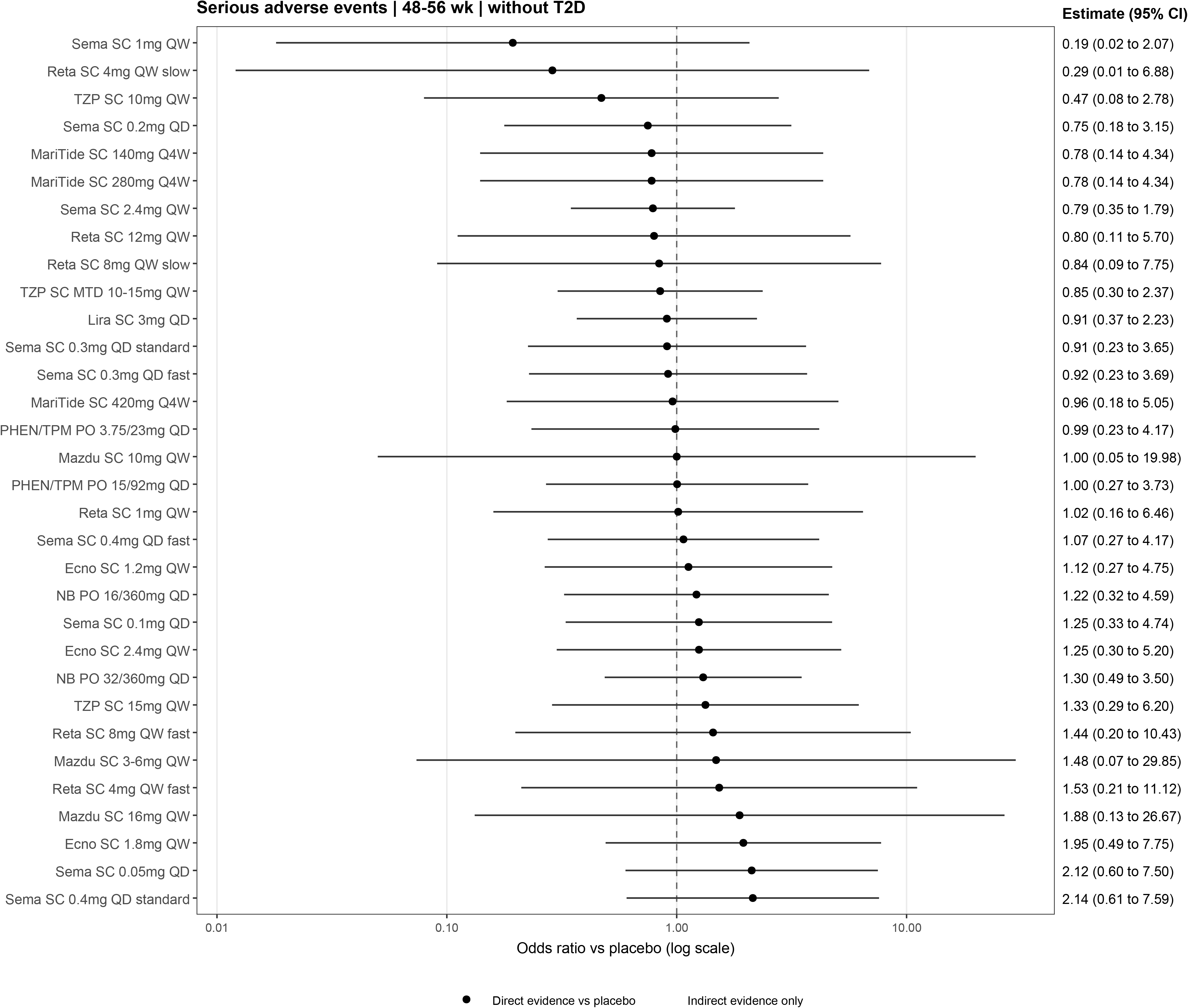

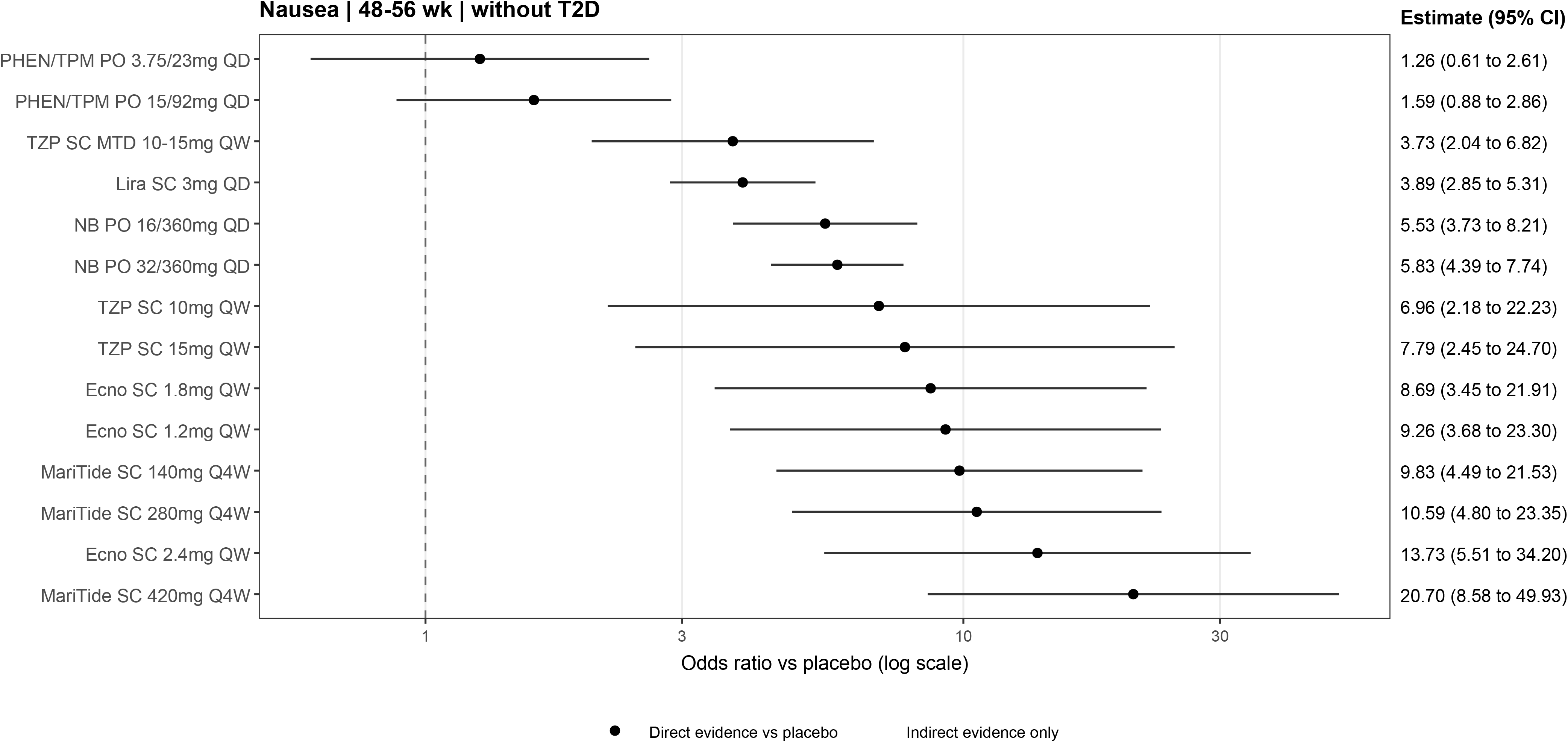

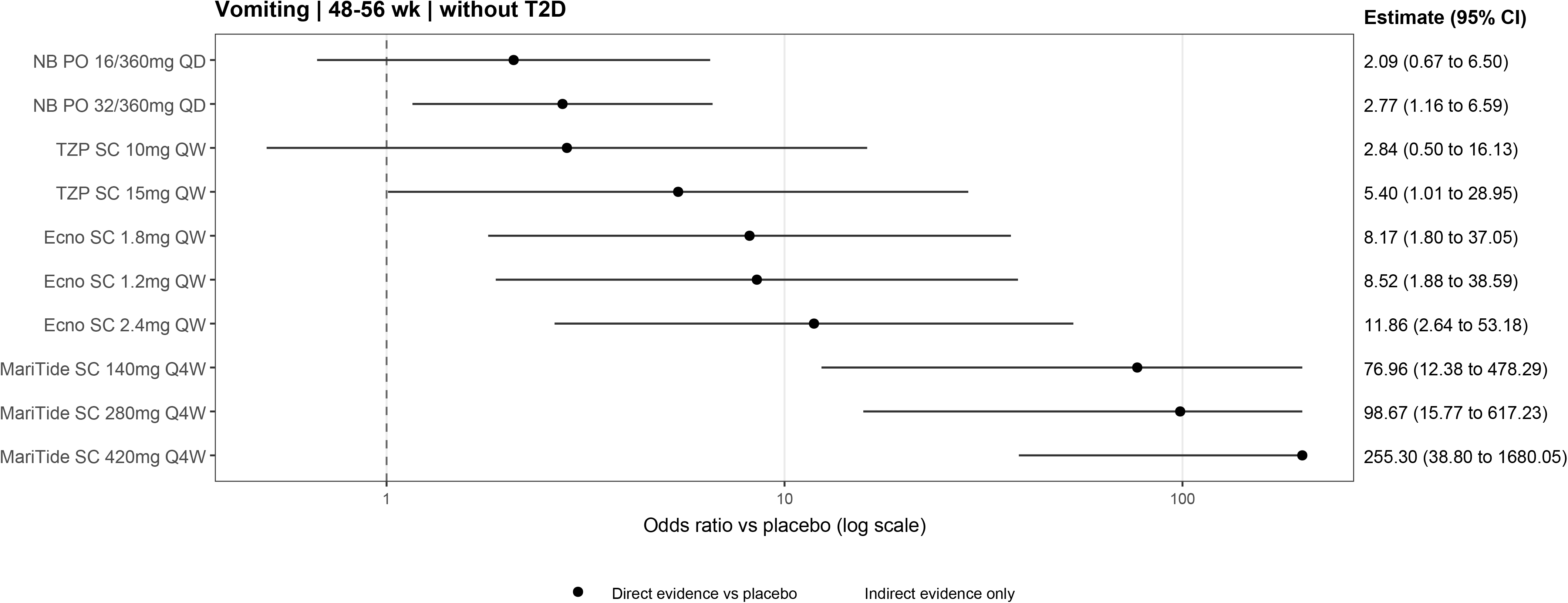

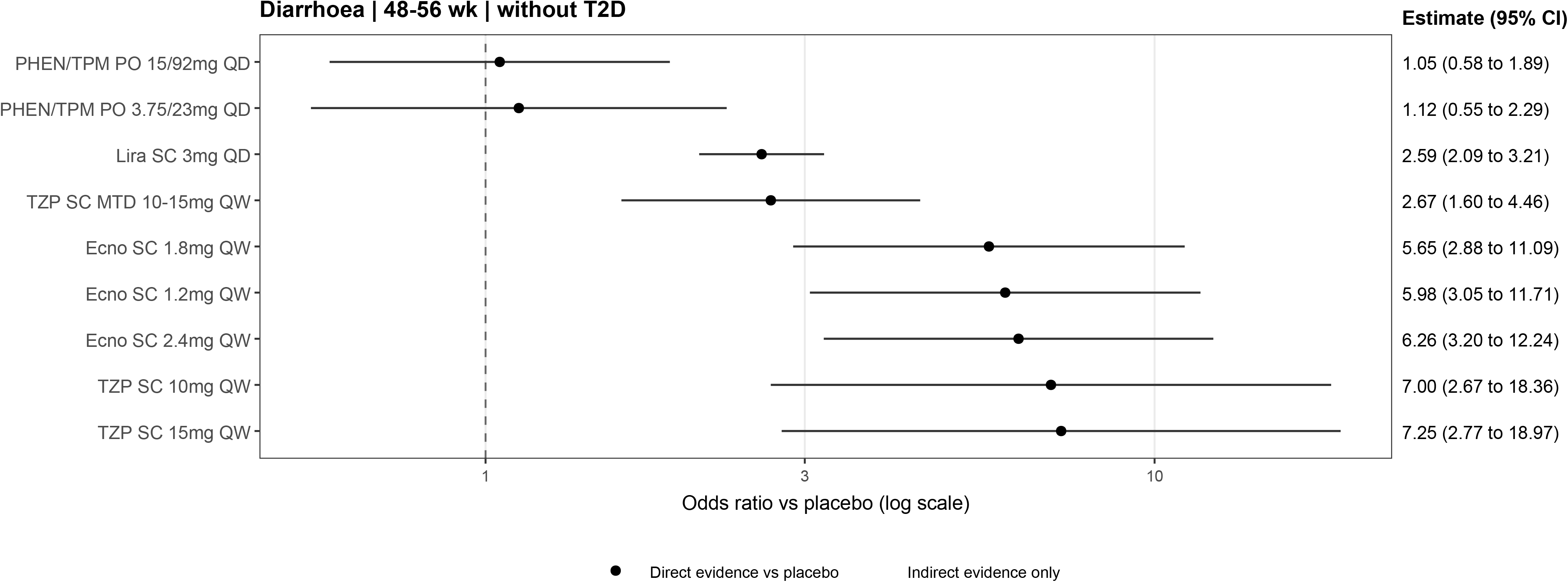

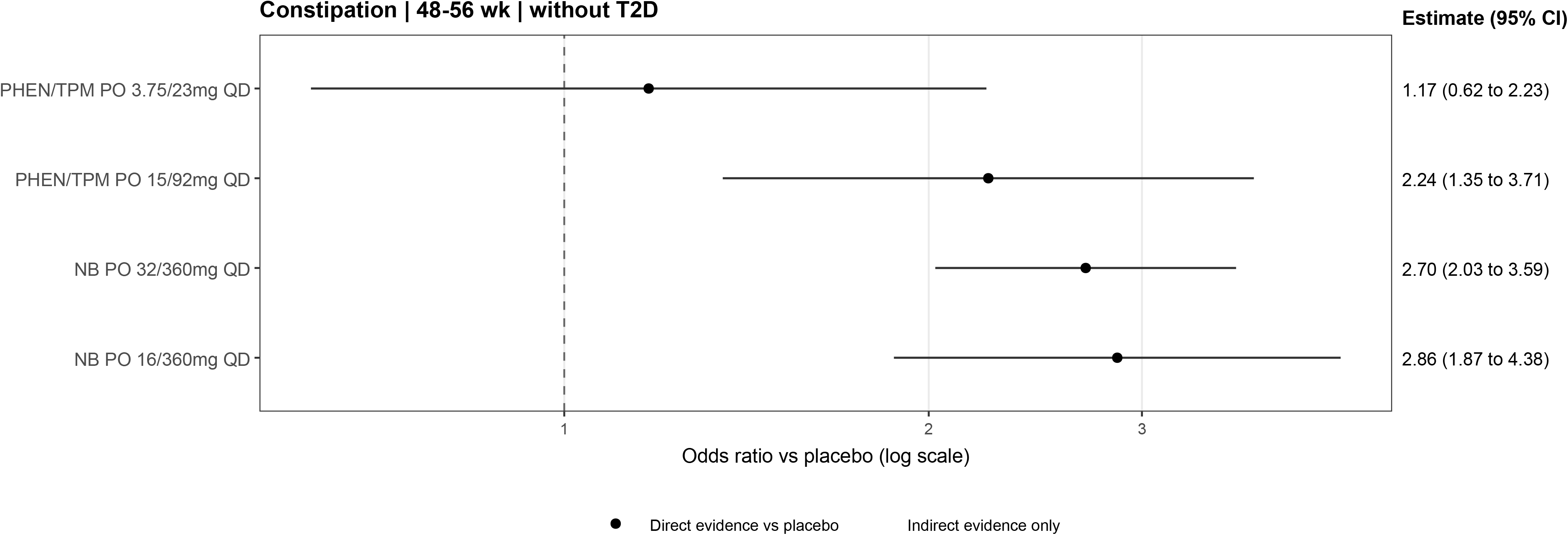

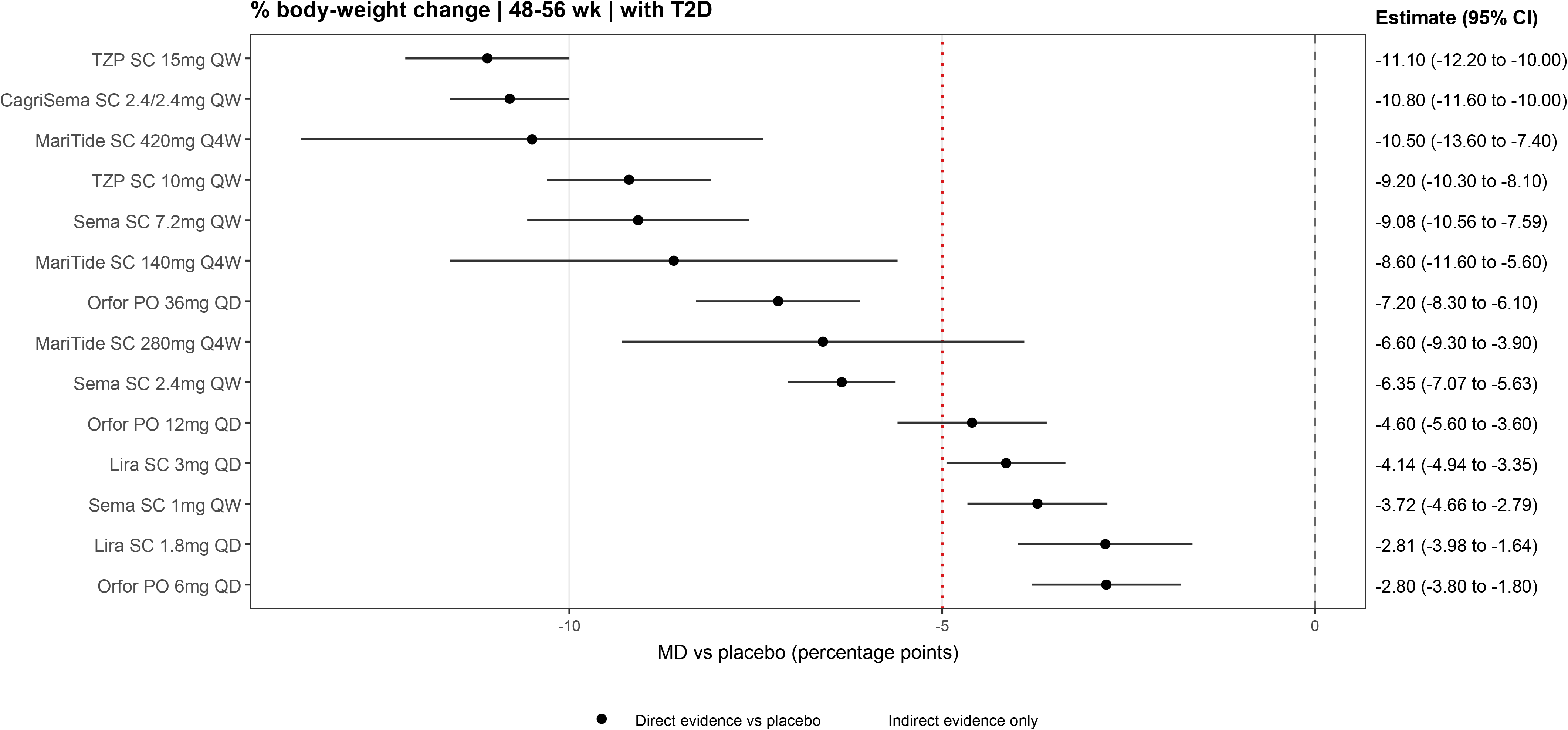

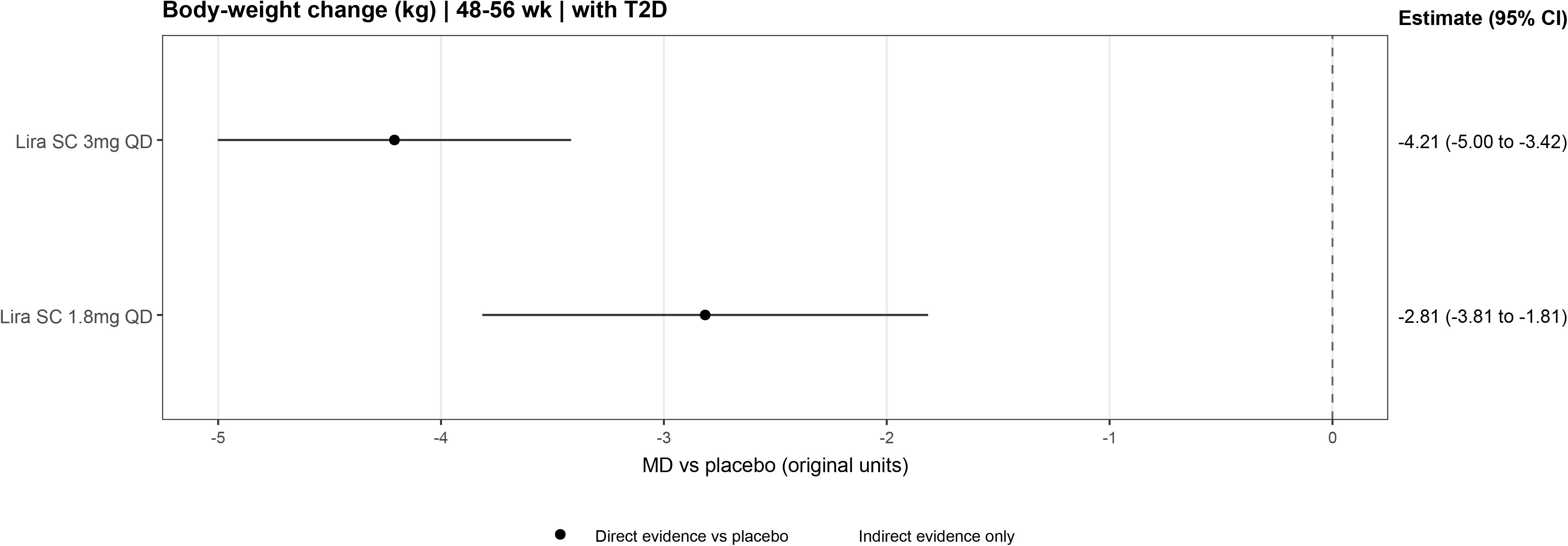

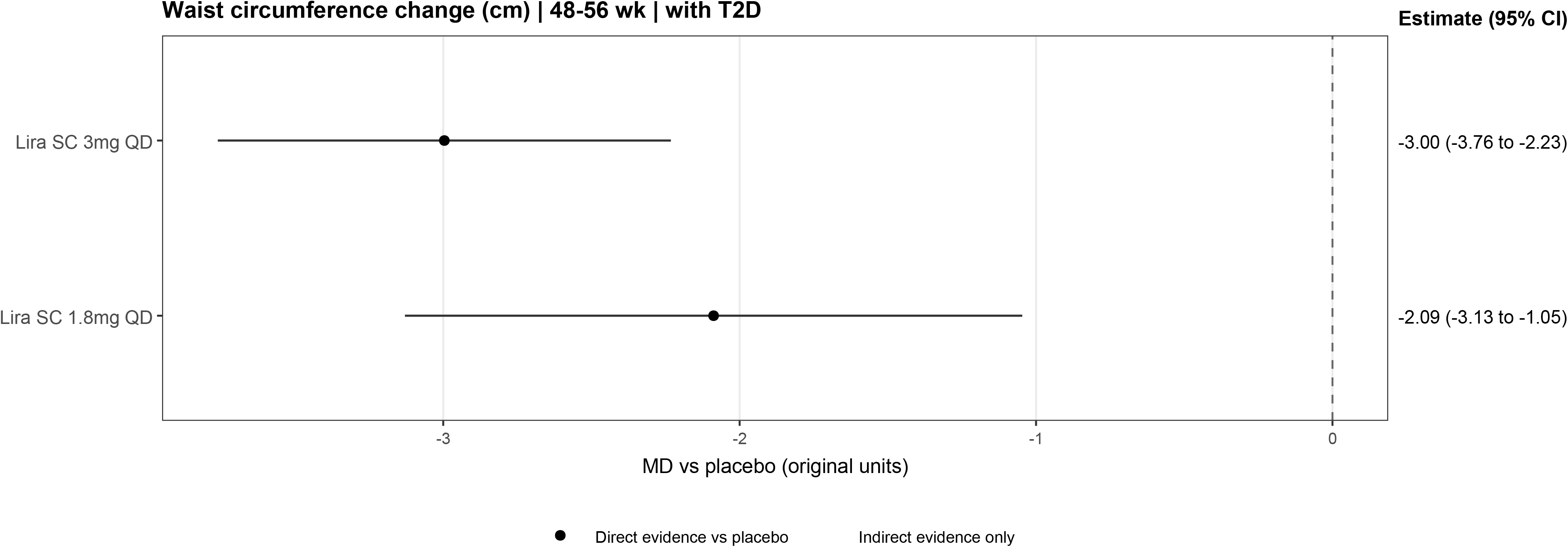

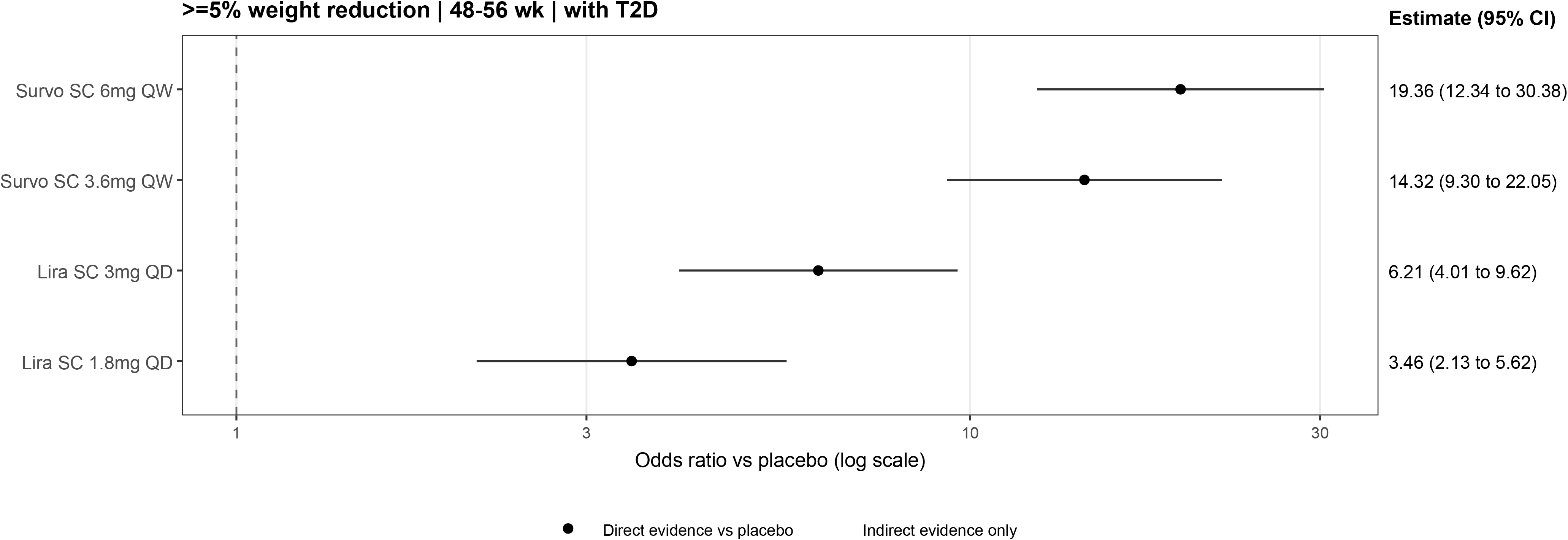

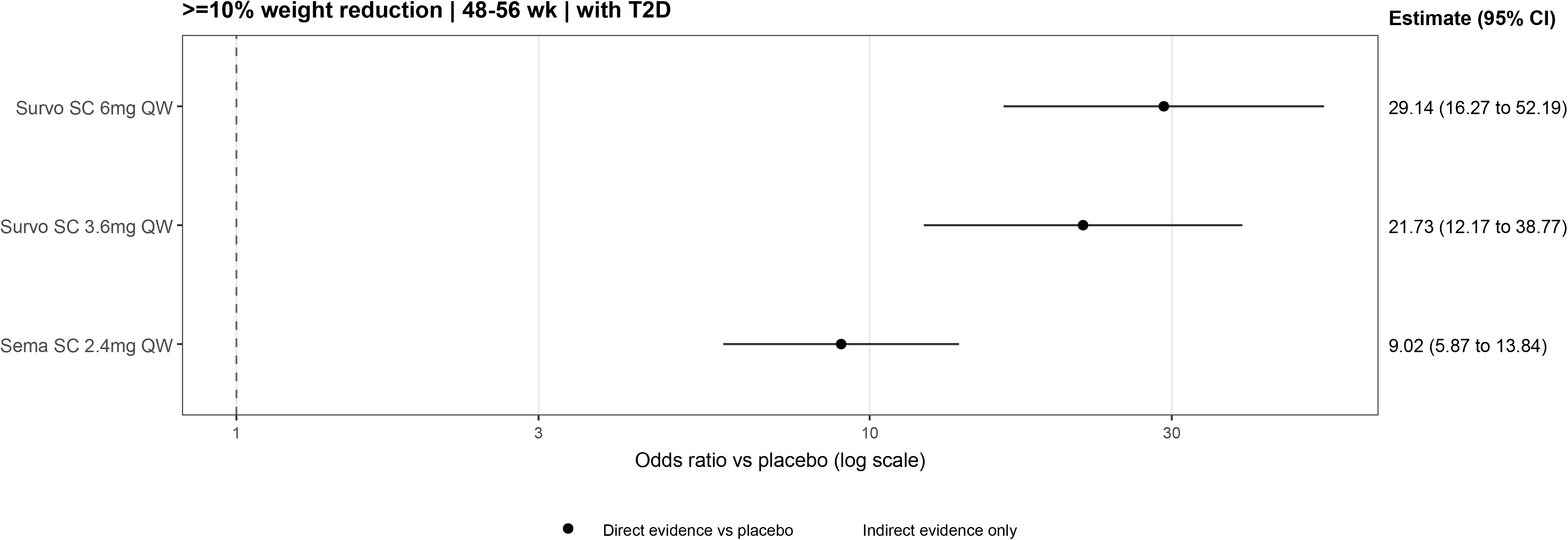

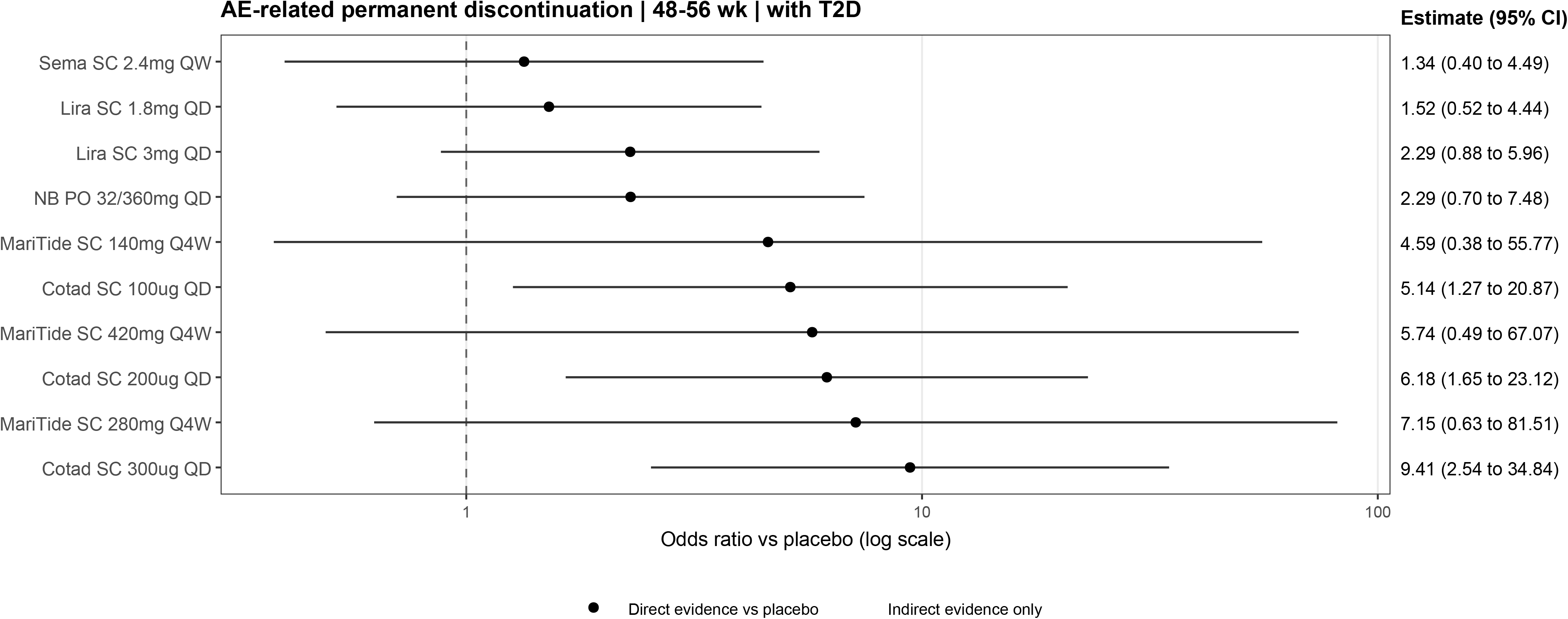

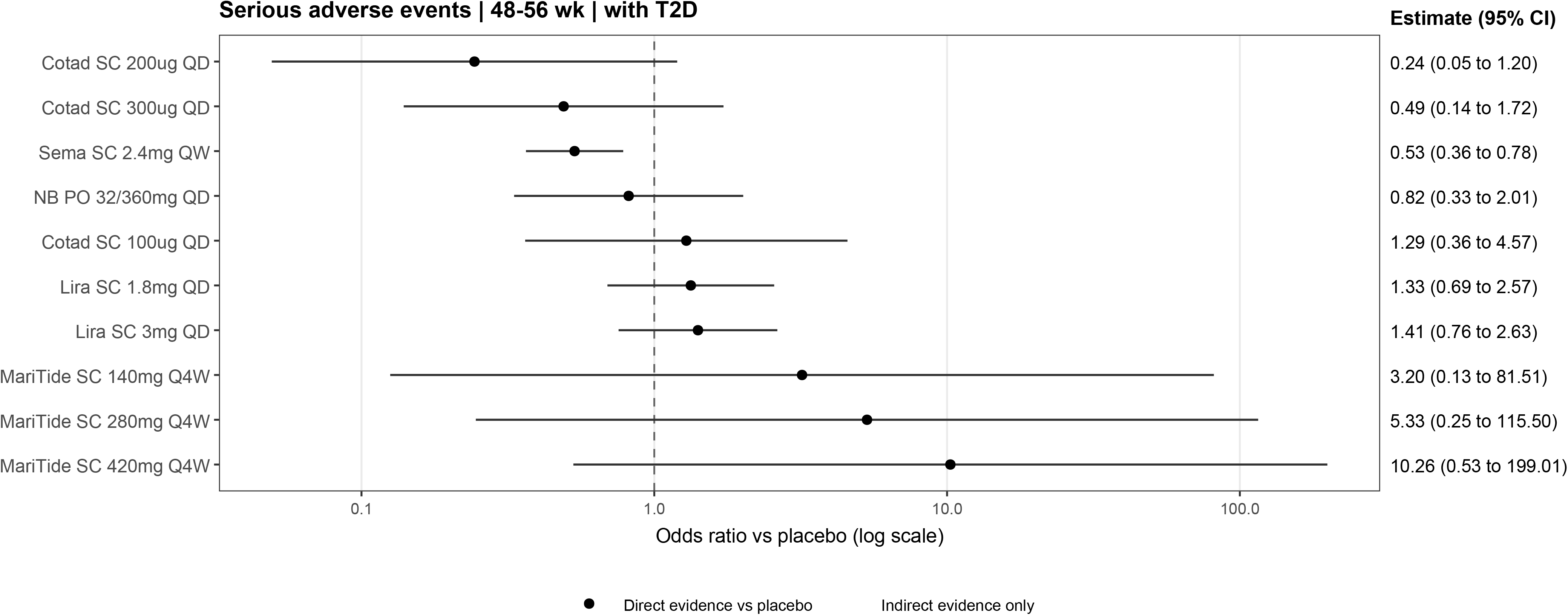

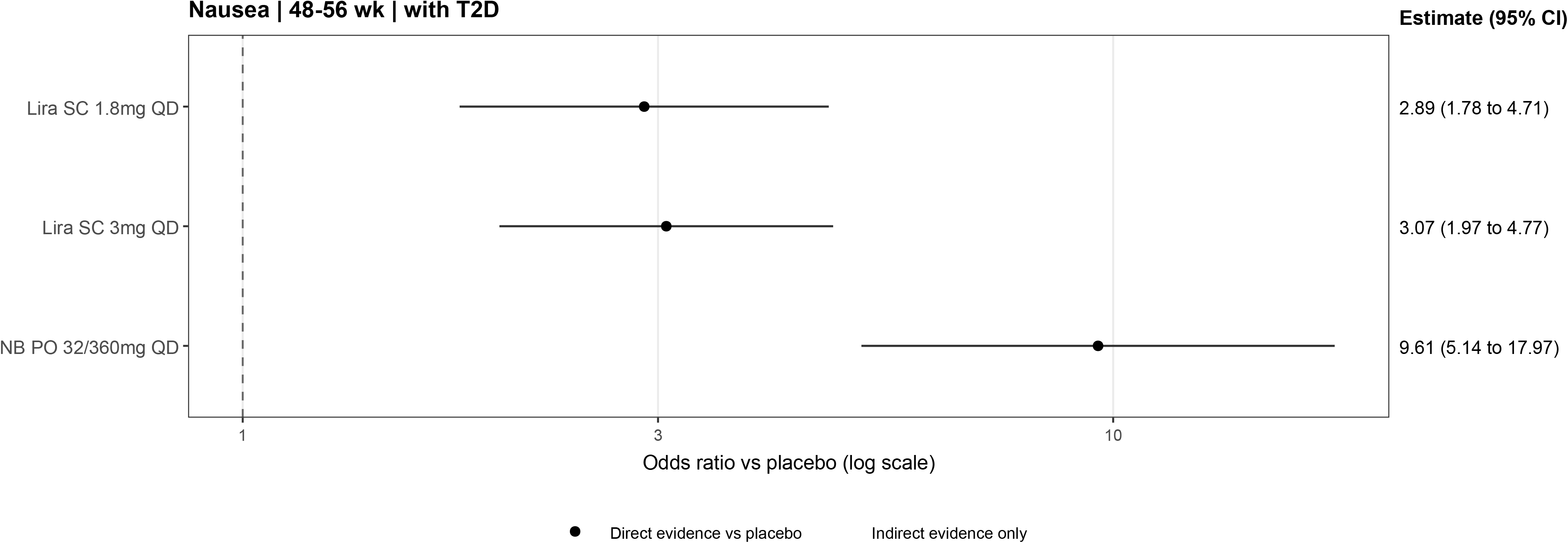

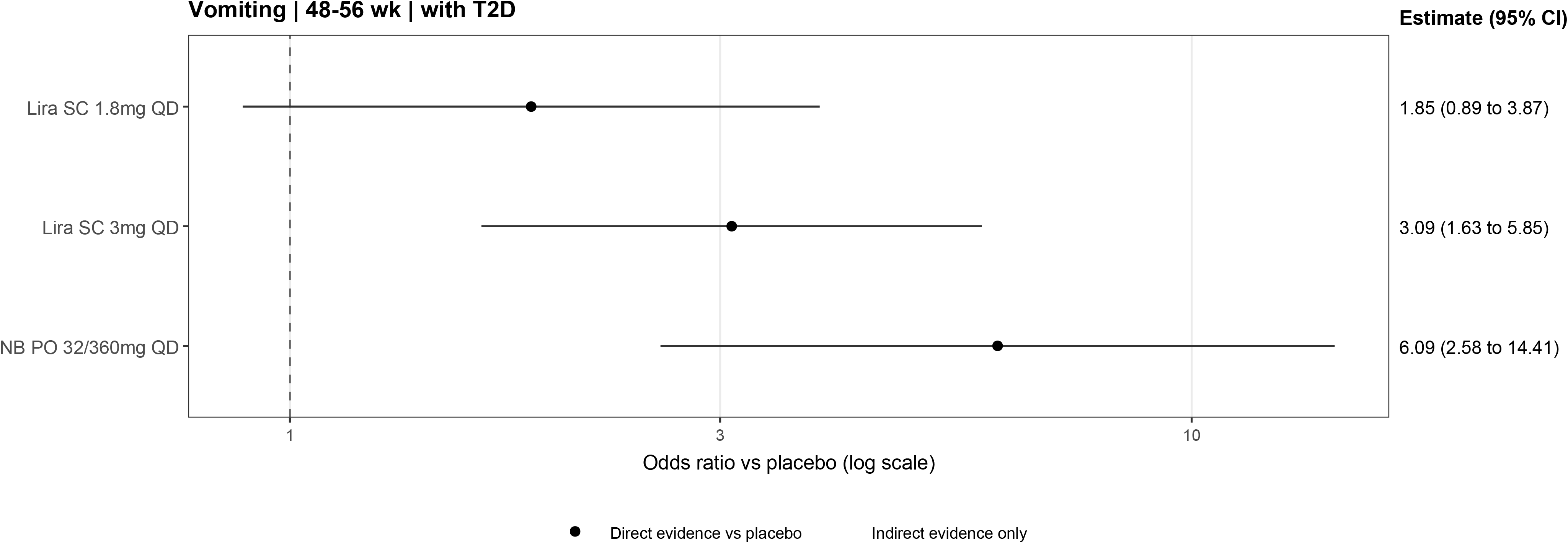

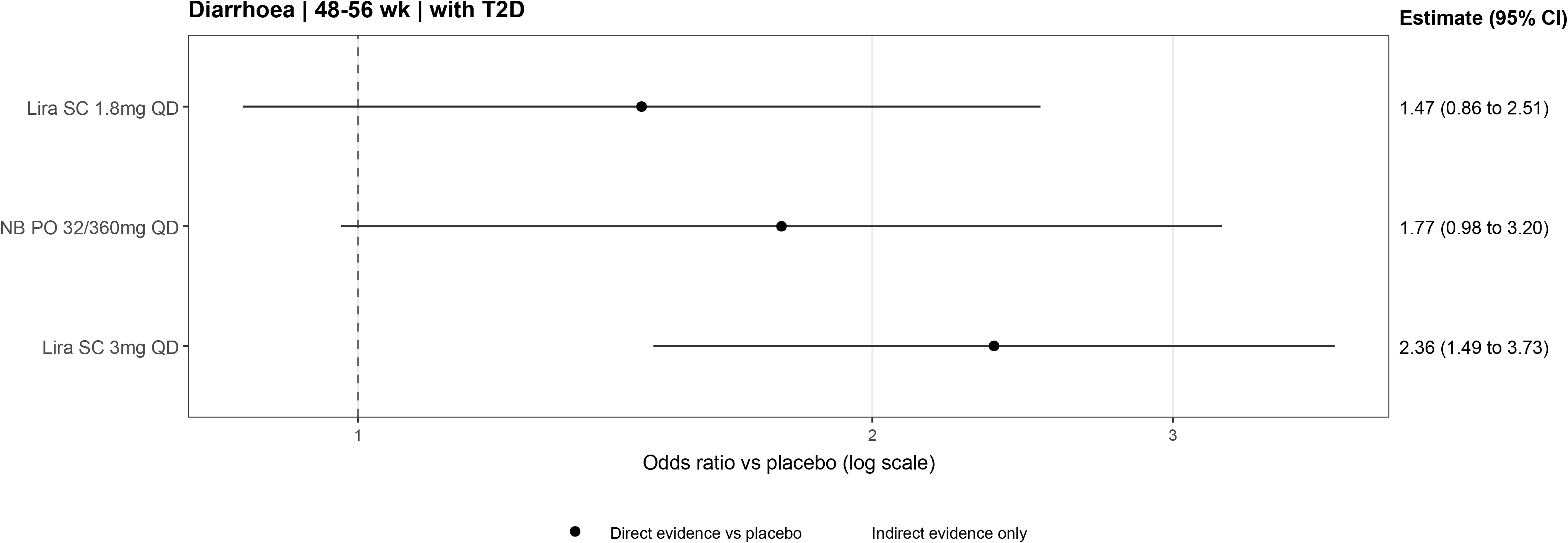

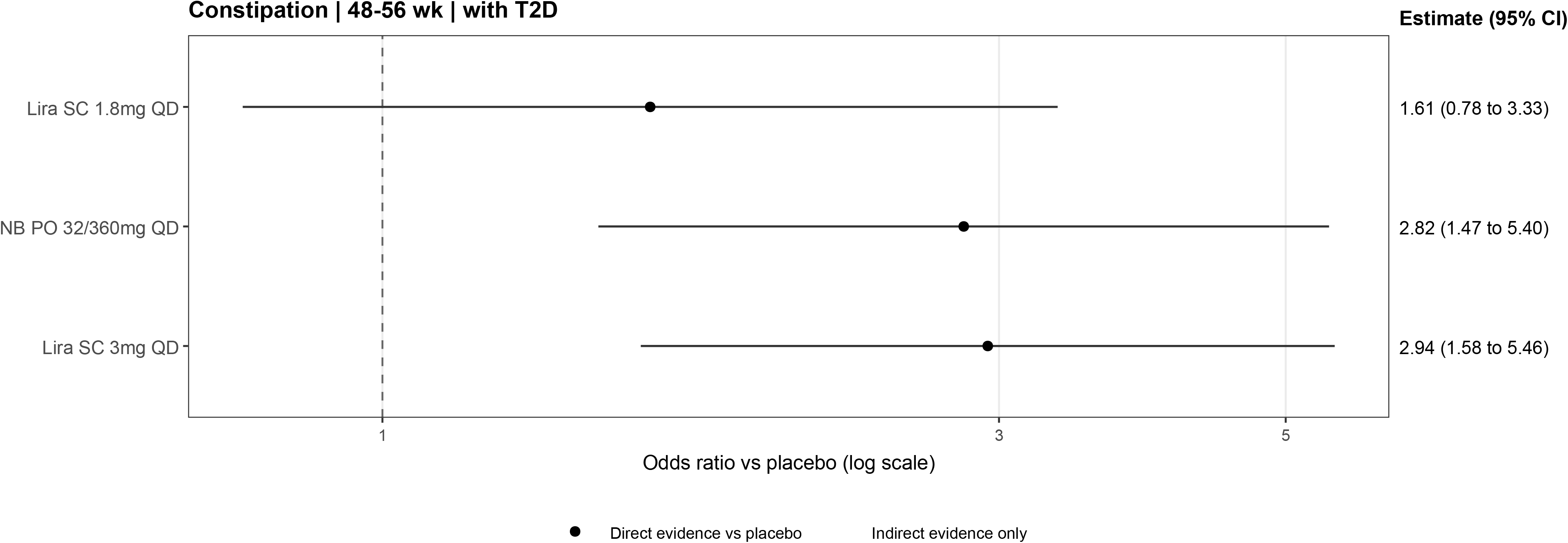

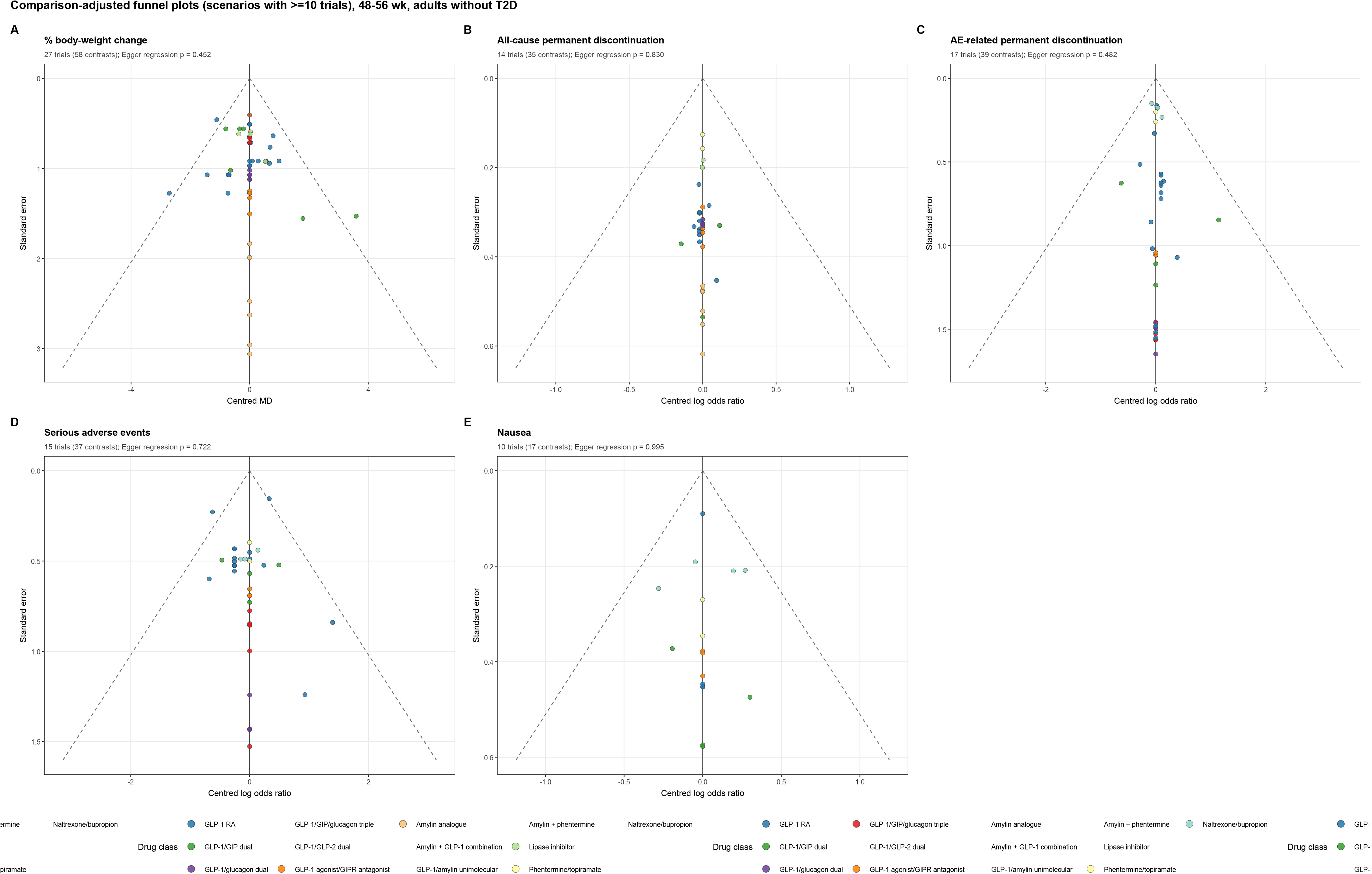

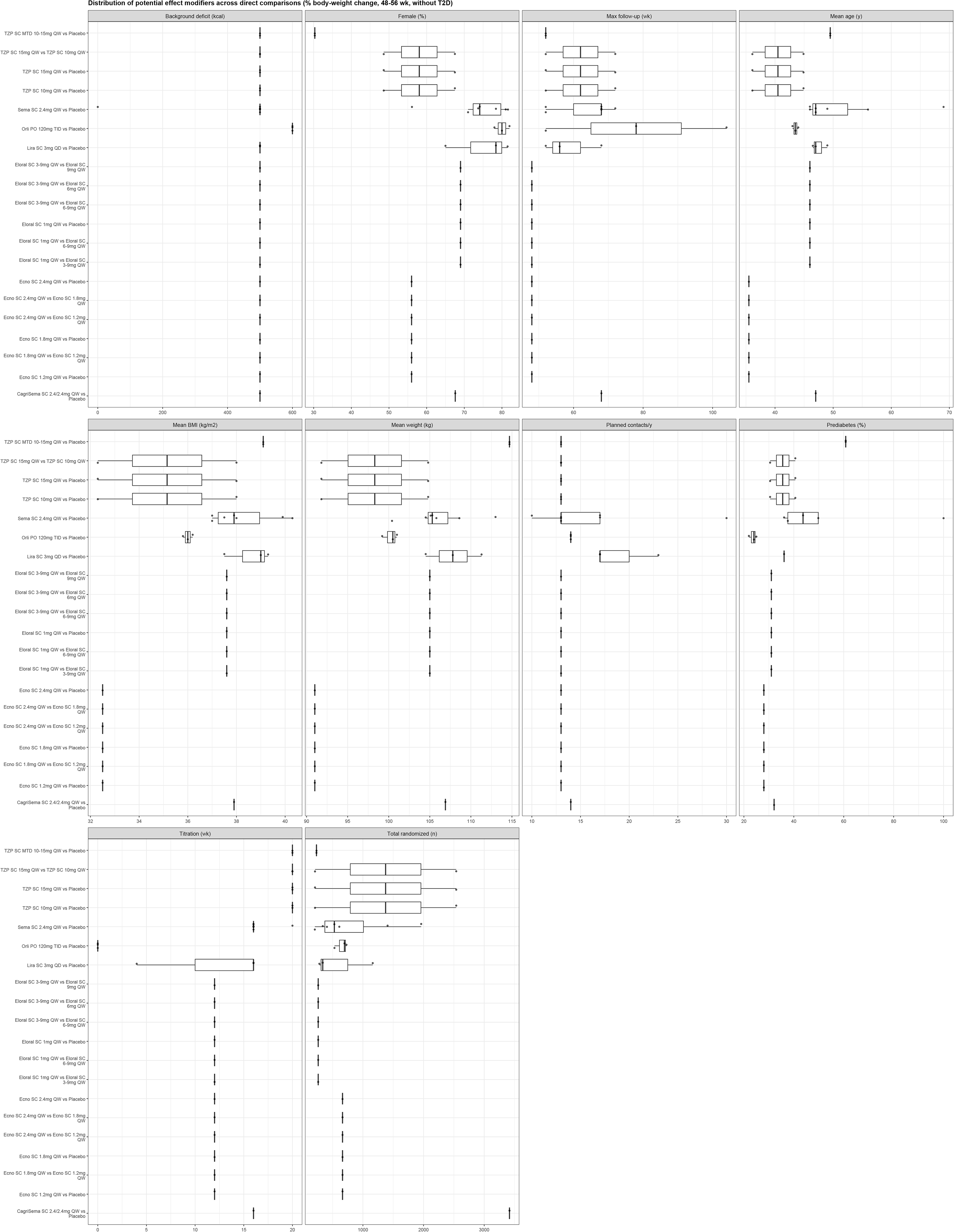

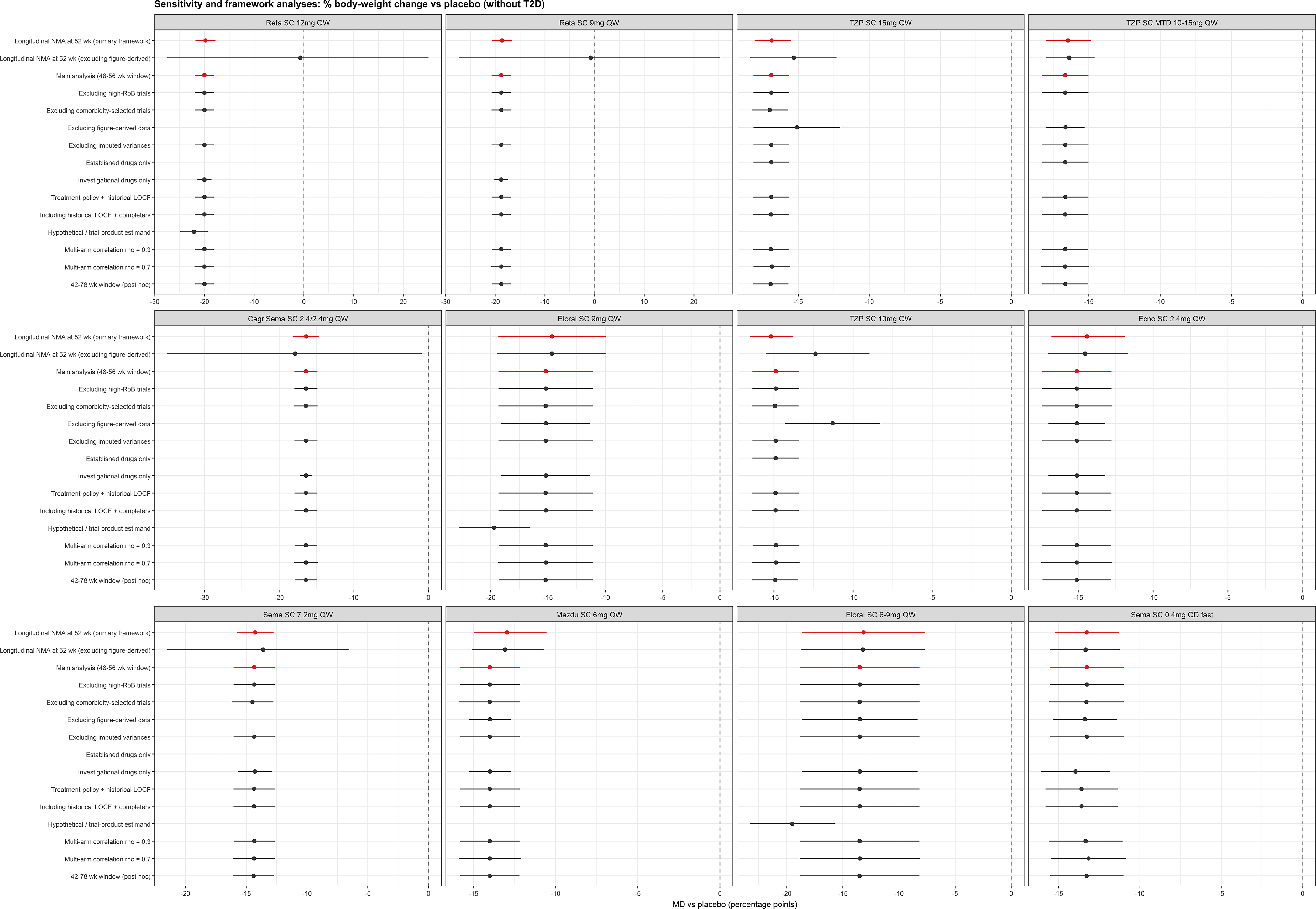

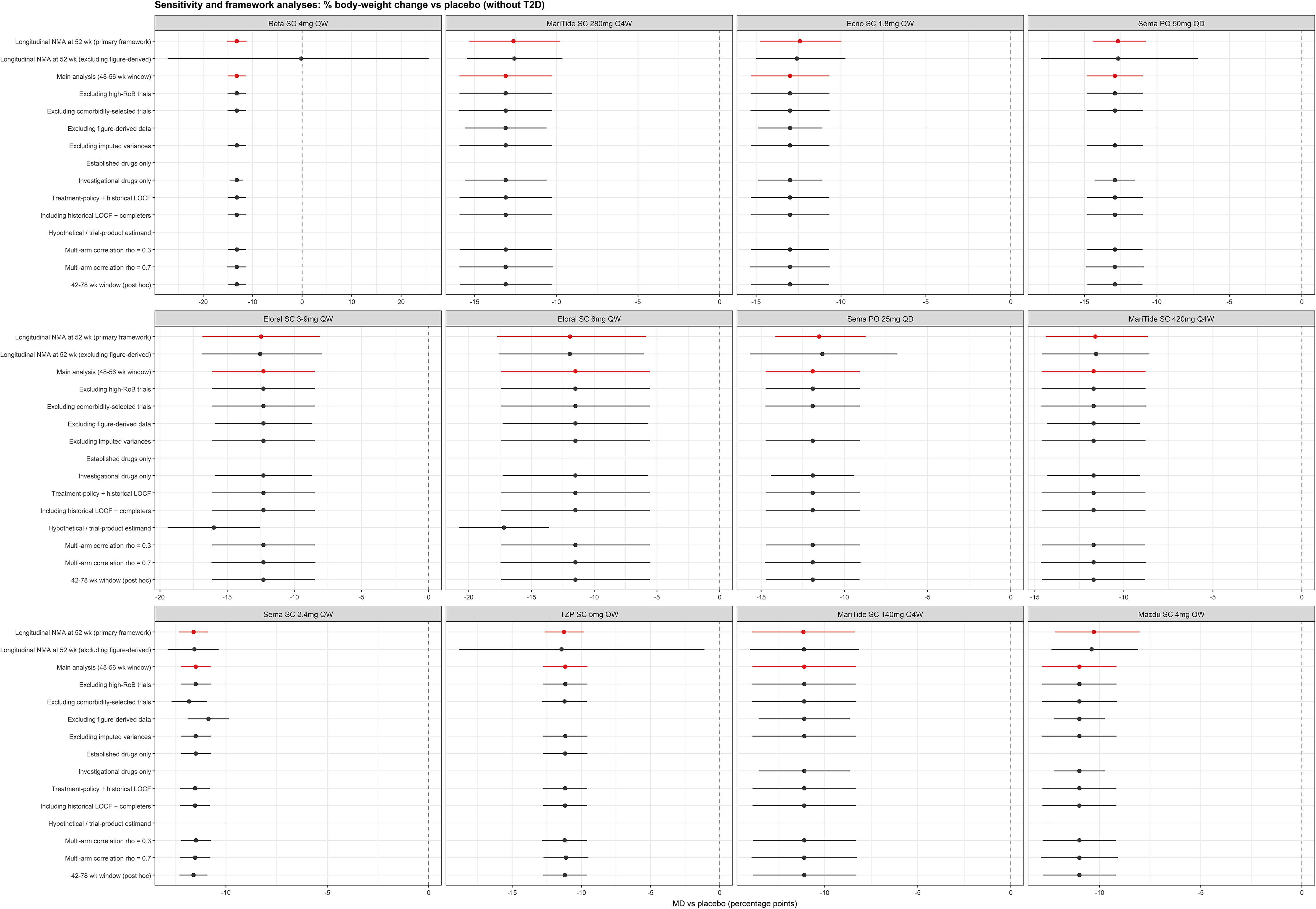

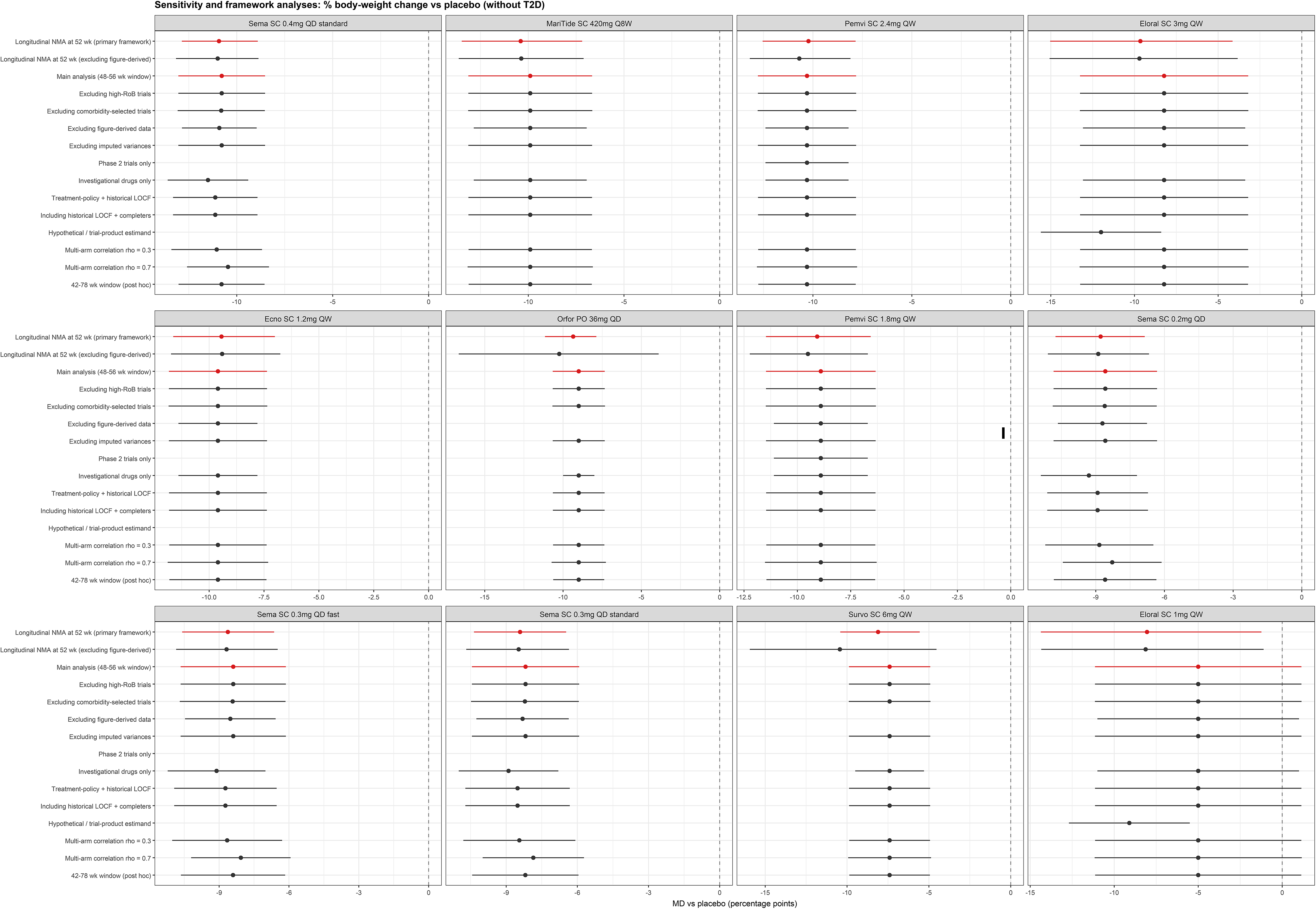

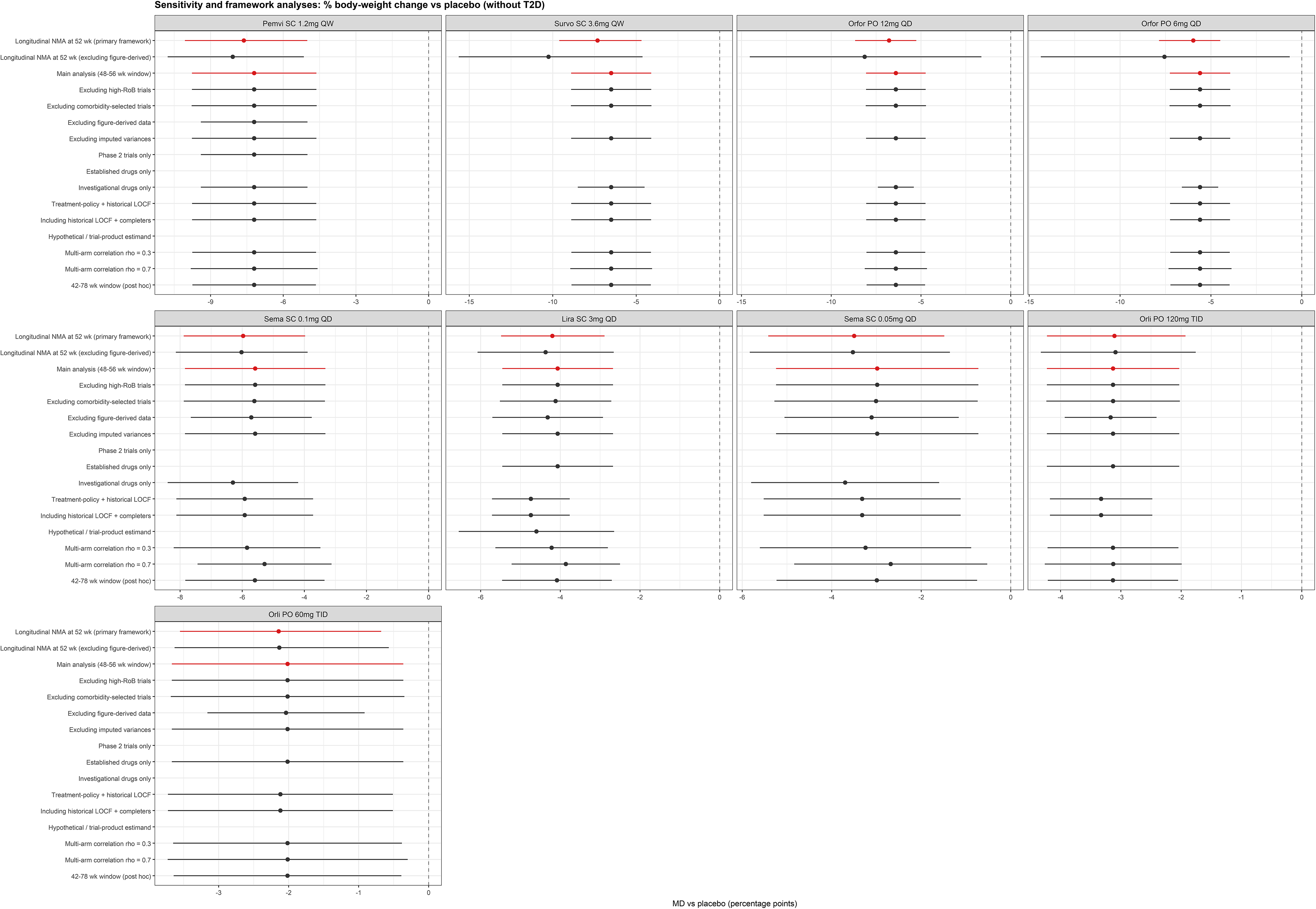

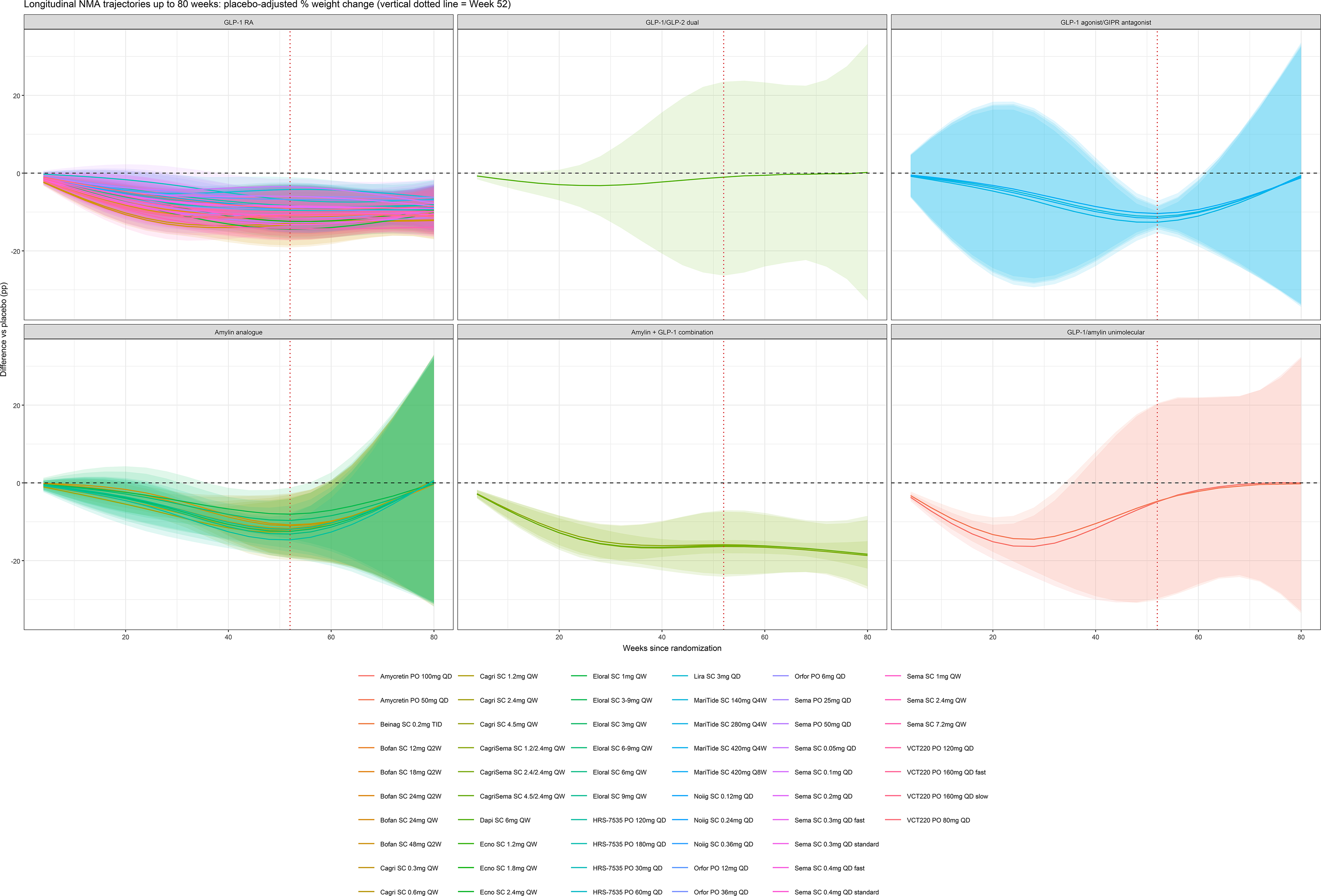

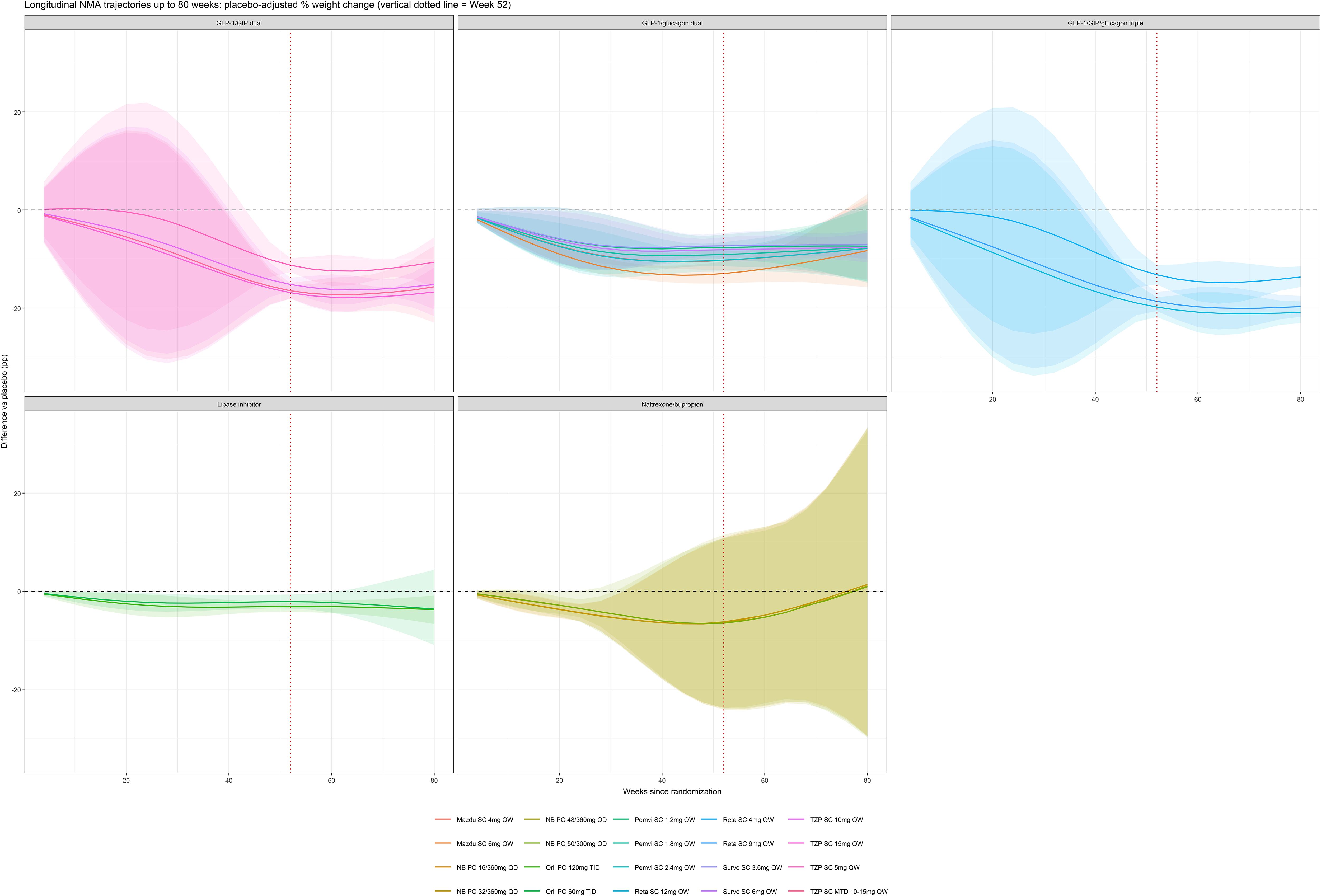

## Supplementary Appendix

## Appendix 1. Literature search strategies

Weight loss and permanent discontinuation across obesity treatment-initiation strategies: Bayesian longitudinal network meta-analysis

### 1.1 Search execution, sources and deduplication

All sources were searched on 7 October 2026, from database inception to that date. No language restriction and no publication-date restriction were applied; Chinese-language databases were searched in Chinese and English-language databases in English. Trial registries were searched on the same day. Reference lists of all included trials and of previous systematic reviews on the same question were hand-searched, forward citation tracking was performed, and regulatory documents (FDA and EMA review reports, clinical study reports) and trial protocols were retrieved for trials without a full publication.

| Source | Type | Records retrieved | Date searched |
|---|---|---|---|
| MEDLINE (via PubMed) | Bibliographic database | 10 329 | 7 October 2026 |
| Embase (via Ovid) | Bibliographic database | 15 482 | 7 October 2026 |
| Cochrane Central Register of Controlled Trials (CENTRAL) | Bibliographic database | 9 416 | 7 October 2026 |
| APA PsycINFO (via Ovid) | Bibliographic database | 943 | 7 October 2026 |
| CINAHL Complete (via EBSCOhost) | Bibliographic database | 2 385 | 7 October 2026 |
| SinoMed / CBM | Bibliographic database (Chinese) | 1 864 | 7 October 2026 |
| CNKI | Bibliographic database (Chinese) | 2 418 | 7 October 2026 |
| Wanfang Data | Bibliographic database (Chinese) | 2 105 | 7 October 2026 |
| ClinicalTrials.gov | Trial registry | 2 146 | 7 October 2026 |
| WHO International Clinical Trials Registry Platform (ICTRP) | Trial registry | 1 682 | 7 October 2026 |
| **Total, bibliographic databases** | **8 sources** | **44 942** |  |
| **Total, trial registries** | **2 sources** | **3 828** |  |

### 1.2 Records and deduplication

Searches of the eight bibliographic databases returned 44 942 records and searches of the two trial registries returned 3 828 records, giving 48 770 records from these sources; a further 148 records were identified through reference lists, forward citation tracking, regulatory documents and trial protocols. Automated cross-database matching on title, DOI and author-year removed 16 420 duplicate records, and manual trial-family linkage merged a further 3 436 associated reports of already identified trials, leaving 28 914 records for screening. Records were screened independently by two reviewers at title and abstract level, and 986 reports were assessed in full text.

No search filter was applied to restrict study design in any database except the highly sensitive randomised-trial filter shown in the strategy lines below; for MEDLINE, Embase, PsycINFO, CINAHL, SinoMed, CNKI and Wanfang the design terms are combined with the population and intervention blocks rather than used as a limiting filter. Records indexed exclusively as animal studies were removed by dedicated NOT lines (MEDLINE lines #9-#10, Embase lines #11-#12). CENTRAL was restricted to trials, and the Chinese databases were restricted to records containing randomisation or trial terminology, as shown in the corresponding lines.

Searches of ClinicalTrials.gov and of the WHO ICTRP used five complementary query sets (10A-10E) covering approved, investigational, regional, oral and amylin-related agents plus the older anti-obesity drugs, because a single registry query could not retrieve all molecule names within the platform’s query length limits.

### 1.3 Search strategies, source by source

#### 1. MEDLINE (via PubMed)

#1 “Obesity“[Mesh] OR “Overweight“[Mesh] OR “Weight Loss“[Mesh] OR “Body Weight Changes“[Mesh] OR “Anti-Obesity Agents“[Mesh]

#2 obes*[tiab] OR overweight*[tiab] OR “over weight*“[tiab] OR adiposit*[tiab] OR corpulen*[tiab] OR “weight loss“[tiab] OR “weight reduction*“[tiab] OR “weight management“[tiab] OR “weight control“[tiab] OR “body weight change*“[tiab] OR “antiobes*“[tiab] OR “anti-obes*“[tiab]

#3 #1 OR #2

#4 “Glucagon-Like Peptide-1 Receptor Agonists“[Mesh] OR “Glucagon-Like Peptide 1“[Mesh] OR “Liraglutide“[Mesh] OR “Exenatide“[Mesh] OR “Orlistat“[Mesh] OR “Amylin Receptor Agonists“[Mesh]

#5 “glucagon-like peptide-1“[tiab] OR “glucagon like peptide 1“[tiab] OR “GLP-1*“[tiab] OR “GLP1*“[tiab] OR “incretin*“[tiab] OR “GIP/GLP*“[tiab] OR “GLP-1/GIP*“[tiab] OR “GLP-1/glucagon*“[tiab] OR “dual agonist*“[tiab] OR “triple agonist*“[tiab] OR “co-agonist*“[tiab] OR “coagonist*“[tiab] OR “triagonist*“[tiab] OR “semaglutide“[tiab] OR “wegovy“[tiab] OR “ozempic“[tiab] OR “rybelsus“[tiab] OR “NN9535“[tiab] OR “NN-9535“[tiab] OR “liraglutide“[tiab] OR “saxenda“[tiab] OR “victoza“[tiab] OR “NN8022“[tiab] OR “NN-8022“[tiab] OR “tirzepatide“[tiab] OR “zepbound“[tiab] OR “mounjaro“[tiab] OR “LY3298176“[tiab] OR “LY-3298176“[tiab] OR “retatrutide“[tiab] OR “LY3437943“[tiab] OR “LY-3437943“[tiab] OR “orforglipron“[tiab] OR “LY3502970“[tiab] OR “LY-3502970“[tiab] OR “OWL833“[tiab] OR “maridebart cafraglutide“[tiab] OR “maridebart“[tiab] OR “cafraglutide“[tiab] OR “maritide“[tiab] OR “AMG 133“[tiab] OR “AMG133“[tiab] OR “AMG-133“[tiab] OR “mazdutide“[tiab] OR “IBI362“[tiab] OR “IBI-362“[tiab] OR “LY3305677“[tiab] OR “LY-3305677“[tiab] OR “OXM3“[tiab] OR “survodutide“[tiab] OR “BI 456906“[tiab] OR “BI456906“[tiab] OR “BI-456906“[tiab] OR “ecnoglutide“[tiab] OR “XW003“[tiab] OR “XW-003“[tiab] OR “XW004“[tiab] OR “bofanglutide“[tiab] OR “noiiglutide“[tiab] OR “SHR20004“[tiab] OR “SHR-20004“[tiab] OR “VCT220“[tiab] OR “VCT-220“[tiab] OR “HRS-7535“[tiab] OR “HRS7535“[tiab] OR “HRS9531“[tiab] OR “HRS-9531“[tiab] OR “pemvidutide“[tiab] OR “ALT-801“[tiab] OR “ALT801“[tiab] OR “danuglipron“[tiab] OR “PF-06882961“[tiab] OR “PF06882961“[tiab] OR “lotiglipron“[tiab] OR “PF-07081532“[tiab] OR “cotadutide“[tiab] OR “MEDI0382“[tiab] OR “MEDI-0382“[tiab] OR “efinopegdutide“[tiab] OR “MK-6024“[tiab] OR “HM12525A“[tiab] OR “efpeglenatide“[tiab] OR “albiglutide“[tiab] OR “dulaglutide“[tiab] OR “exenatide“[tiab] OR “lixisenatide“[tiab] OR “beinaglutide“[tiab] OR “benaglutide“[tiab] OR “loxenatide“[tiab] OR “pegapamodutide“[tiab] OR “apraglutide“[tiab] OR “GMA106“[tiab] OR “CT-388“[tiab] OR “CT388“[tiab] OR “CT-996“[tiab] OR “CT996“[tiab] OR “VK2735“[tiab] OR “VK-2735“[tiab] OR “GSBR-1290“[tiab] OR “GSBR1290“[tiab] OR “ECC5004“[tiab] OR “AZD5004“[tiab] OR “RGT-075“[tiab] OR “HDM1002“[tiab] OR “TG103“[tiab] OR “GZR18“[tiab] OR “PB-119“[tiab] OR “RAY1225“[tiab] OR “UBT251“[tiab] OR “BGM0504“[tiab]

#6 “amylin*“[tiab] OR “cagrilintide“[tiab] OR “AM833“[tiab] OR “AM-833“[tiab] OR “NNC0174-0833“[tiab] OR “CagriSema“[tiab] OR “cagrilintide-semaglutide“[tiab] OR “cagrilintide/semaglutide“[tiab] OR “amycretin“[tiab] OR “NNC0487-0111“[tiab] OR “eloralintide“[tiab] OR “LY3841136“[tiab] OR “LY-3841136“[tiab] OR “pramlintide“[tiab] OR “symlin“[tiab] OR “davalintide“[tiab] OR “AC2307“[tiab] OR “petrelintide“[tiab] OR “ZP8396“[tiab] OR “ZP-8396“[tiab] OR “AZD6234“[tiab] OR “GUB014295“[tiab] OR “orlistat“[tiab] OR “xenical“[tiab] OR “alli“[tiab] OR “tetrahydrolipstatin“[tiab] OR “RO 18-0647“[tiab] OR (“phentermine“[tiab] AND “topiramate“[tiab]) OR “phentermine-topiramate“[tiab] OR “phentermine/topiramate“[tiab] OR “qsymia“[tiab] OR “qsiva“[tiab] OR “qnexa“[tiab] OR “VI-0521“[tiab] OR (“naltrexone“[tiab] AND “bupropion“[tiab]) OR “naltrexone-bupropion“[tiab] OR “naltrexone/bupropion“[tiab] OR “contrave“[tiab] OR “mysimba“[tiab] OR “NB32“[tiab]

#7 #4 OR #5 OR #6

#8 “randomized controlled trial“[pt] OR “controlled clinical trial“[pt] OR randomized[tiab] OR randomised[tiab] OR placebo[tiab] OR “drug therapy“[sh] OR randomly[tiab] OR trial[tiab] OR groups[tiab]

#9 “Animals“[Mesh] NOT (“Animals“[Mesh] AND “Humans“[Mesh])

#10 #8 NOT #9

#11 #3 AND #7 AND #10

#### 2. Embase (via Ovid)

1 exp obesity/ or exp weight reduction/ or exp body weight change/ or exp antiobesity agent/

2 (obes* or overweight* or “over weight*” or adiposit* or corpulen* or “weight loss” or “weight reduction*” or “weight management” or “weight control” or “body weight change*” or antiobes* or “anti-obes*”).ti,ab,kw.

3 1 or 2

4 exp glucagon like peptide 1 receptor agonist/ or exp incretin mimetic agent/ or exp amylin/ or exp amylin derivative/ or exp tetrahydrolipstatin/ or exp phentermine plus topiramate/ or exp bupropion plus naltrexone/

5 (“glucagon-like peptide-1” or “glucagon like peptide 1” or “GLP-1*” or “GLP1*” or incretin* or “GIP/GLP*” or “GLP-1/GIP*” or “GLP-1/glucagon*” or “dual agonist*” or “triple agonist*” or “co-agonist*” or coagonist* or triagonist* or semaglutide or wegovy or ozempic or rybelsus or NN9535 or “NN-9535” or liraglutide or saxenda or victoza or NN8022 or “NN-8022” or tirzepatide or zepbound or mounjaro or LY3298176 or “LY-3298176” or retatrutide or LY3437943 or “LY-3437943” or orforglipron or LY3502970 or “LY-3502970” or OWL833 or “maridebart cafraglutide” or maridebart or cafraglutide or maritide or “AMG 133” or AMG133 or “AMG-133” or mazdutide or IBI362 or “IBI-362” or LY3305677 or “LY-3305677” or OXM3 or survodutide or “BI 456906” or BI456906 or “BI-456906” or ecnoglutide or XW003 or “XW-003” or XW004 or bofanglutide or noiiglutide or SHR20004 or “SHR-20004” or VCT220 or “VCT-220” or “HRS-7535” or HRS7535 or HRS9531 or “HRS-9531” or pemvidutide or “ALT-801” or ALT801 or danuglipron or “PF-06882961” or PF06882961 or lotiglipron or “PF-07081532” or cotadutide or MEDI0382 or “MEDI-0382” or efinopegdutide or “MK-6024” or HM12525A or efpeglenatide or albiglutide or dulaglutide or exenatide or lixisenatide or beinaglutide or benaglutide or loxenatide or pegapamodutide or apraglutide or GMA106 or “CT-388” or CT388 or “CT-996” or CT996 or VK2735 or “VK-2735” or “GSBR-1290” or GSBR1290 or ECC5004 or AZD5004 or “RGT-075” or HDM1002 or TG103 or GZR18 or “PB-119” or RAY1225 or UBT251 or BGM0504).ti,ab,kw,tn,rn.

6 (amylin* or cagrilintide or AM833 or “AM-833” or “NNC0174-0833” or CagriSema or “cagrilintide-semaglutide” or “cagrilintide/semaglutide” or amycretin or “NNC0487-0111” or eloralintide or LY3841136 or “LY-3841136” or pramlintide or symlin or davalintide or AC2307 or petrelintide or ZP8396 or “ZP-8396” or AZD6234 or GUB014295 or orlistat or xenical or alli or tetrahydrolipstatin or “RO 18-0647” or (phentermine and topiramate) or “phentermine-topiramate” or “phentermine/topiramate” or qsymia or qsiva or qnexa or “VI-0521” or (naltrexone and bupropion) or “naltrexone-bupropion” or “naltrexone/bupropion” or contrave or mysimba or NB32).ti,ab,kw,tn,rn.

7 4 or 5 or 6

8 randomized controlled trial/ or controlled clinical trial/ or single blind procedure/ or double blind procedure/ or triple blind procedure/ or crossover procedure/

9 (random* or factorial* or crossover* or “cross over*” or placebo* or ((doubl* or singl* or tripl* or trebl*) adj3 (blind* or mask*)) or assign* or allocat* or volunteer*).ti,ab,kw.

10 8 or 9

11 (exp animal/ or animal.hw. or nonhuman/) not (exp human/ or human cell/ or (human or humans).ti,ab.)

12 10 not 11

13 3 and 7 and 12

3. Cochrane Central Register of Controlled Trials (CENTRAL)

#1 [mh “Obesity”] OR [mh “Overweight”] OR [mh “Weight Loss”] OR [mh “Body Weight Changes”] OR [mh “Anti-Obesity Agents”]

#2 (obes* OR overweight* OR “over weight*” OR adiposit* OR corpulen* OR “weight loss” OR “weight reduction*” OR “weight management” OR “weight control” OR “body weight change*” OR antiobes* OR “anti-obes*”):ti,ab,kw

#3 #1 OR #2

#4 [mh “Glucagon-Like Peptide-1 Receptor Agonists”] OR [mh “Glucagon-Like Peptide 1”] OR [mh “Liraglutide”] OR [mh “Exenatide”] OR [mh “Orlistat”] OR [mh “Amylin Receptor Agonists”]

#5 (“glucagon-like peptide-1” OR “glucagon like peptide 1” OR “GLP-1*” OR “GLP1*” OR incretin* OR “GIP/GLP*” OR “GLP-1/GIP*” OR “GLP-1/glucagon*” OR “dual agonist*” OR “triple agonist*” OR “co-agonist*” OR coagonist* OR triagonist* OR semaglutide OR wegovy OR ozempic OR rybelsus OR NN9535 OR “NN-9535” OR liraglutide OR saxenda OR victoza OR NN8022 OR “NN-8022” OR tirzepatide OR zepbound OR mounjaro OR LY3298176 OR “LY-3298176” OR retatrutide OR LY3437943 OR “LY-3437943” OR orforglipron OR LY3502970 OR “LY-3502970” OR OWL833 OR “maridebart cafraglutide” OR maridebart OR cafraglutide OR maritide OR “AMG 133” OR AMG133 OR “AMG-133” OR mazdutide OR IBI362 OR “IBI-362” OR LY3305677 OR “LY-3305677” OR OXM3 OR survodutide OR “BI 456906” OR BI456906 OR “BI-456906” OR ecnoglutide OR XW003 OR “XW-003” OR XW004 OR bofanglutide OR noiiglutide OR SHR20004 OR “SHR-20004” OR VCT220 OR “VCT-220” OR “HRS-7535” OR HRS7535 OR HRS9531 OR “HRS-9531” OR pemvidutide OR “ALT-801” OR ALT801 OR danuglipron OR “PF-06882961” OR PF06882961 OR lotiglipron OR “PF-07081532” OR cotadutide OR MEDI0382 OR “MEDI-0382” OR efinopegdutide OR “MK-6024” OR HM12525A OR efpeglenatide OR albiglutide OR dulaglutide OR exenatide OR lixisenatide OR beinaglutide OR benaglutide OR loxenatide OR pegapamodutide OR apraglutide OR GMA106 OR “CT-388” OR CT388 OR “CT-996” OR CT996 OR VK2735 OR “VK-2735” OR “GSBR-1290” OR GSBR1290 OR ECC5004 OR AZD5004 OR “RGT-075” OR HDM1002 OR TG103 OR GZR18 OR “PB-119” OR RAY1225 OR UBT251 OR BGM0504):ti,ab,kw

#6 (amylin* OR cagrilintide OR AM833 OR “AM-833” OR “NNC0174-0833” OR CagriSema OR “cagrilintide-semaglutide” OR “cagrilintide/semaglutide” OR amycretin OR “NNC0487-0111” OR eloralintide OR LY3841136 OR “LY-3841136” OR pramlintide OR symlin OR davalintide OR AC2307 OR petrelintide OR ZP8396 OR “ZP-8396” OR AZD6234 OR GUB014295 OR orlistat OR xenical OR alli OR tetrahydrolipstatin OR “RO 18-0647” OR (phentermine AND topiramate) OR “phentermine-topiramate” OR “phentermine/topiramate” OR qsymia OR qsiva OR qnexa OR “VI-0521” OR (naltrexone AND bupropion) OR “naltrexone-bupropion” OR “naltrexone/bupropion” OR contrave OR mysimba OR NB32):ti,ab,kw

#7 #4 OR #5 OR #6

#8 #3 AND #7 in Trials

#### 4. APA PsycINFO (via Ovid)

1 exp Obesity/ or exp Overweight/ or exp Weight Loss/ or expWeight Control/

2 (obes* or overweight* or “over weight*” or adiposit* or corpulen* or “weight loss” or “weight reduction*” or “weight management” or “weight control” or “body weight change*” or antiobes* or “anti-obes*”).ti,ab,id.

3 1 or 2

4 (“glucagon-like peptide-1” or “glucagon like peptide 1” or “GLP-1*” or “GLP1*” or incretin* or “GIP/GLP*” or “GLP-1/GIP*” or “GLP-1/glucagon*” or “dual agonist*” or “triple agonist*” or “co-agonist*” or coagonist* or triagonist* or semaglutide or wegovy or ozempic or rybelsus or NN9535 or liraglutide or saxenda or victoza or NN8022 or tirzepatide or zepbound or mounjaro or LY3298176 or retatrutide or LY3437943 or orforglipron or LY3502970 or OWL833 or “maridebart cafraglutide” or maridebart or cafraglutide or maritide or “AMG 133” or AMG133 or mazdutide or IBI362 or LY3305677 or survodutide or “BI 456906” or BI456906 or ecnoglutide or XW003 or bofanglutide or noiiglutide or SHR20004 or VCT220 or “HRS-7535” or HRS7535 or pemvidutide or “ALT-801” or ALT801 or danuglipron or “PF-06882961” or lotiglipron or cotadutide or MEDI0382 or efinopegdutide or efpeglenatide or albiglutide or dulaglutide or exenatide or lixisenatide or beinaglutide or benaglutide or loxenatide or amycretin or eloralintide or cagrilintide or AM833 or CagriSema or pramlintide or symlin or davalintide or petrelintide or orlistat or xenical or alli or tetrahydrolipstatin or (phentermine and topiramate) or “phentermine-topiramate” or qsymia or qsiva or qnexa or (naltrexone and bupropion) or “naltrexone-bupropion” or contrave or mysimba or NB32).ti,ab,id,hw.

5 exp Clinical Trials/ or exp Randomized Controlled Trials/

6 (random* or placebo* or ((doubl* or singl* or tripl*) adj3 (blind* or mask*)) or “controlled trial*” or “clinical trial*” or allocat* or assign*).ti,ab,id.

7 5 or 6

8 3 and 4 and 7

#### 5. CINAHL Complete (via EBSCOhost)

S1 (MH “Obesity+”) OR (MH “Weight Loss+”) OR (MH “Body Weight Changes+”) OR (MH “Antiobesity Agents+”)

S2 TI (obes* OR overweight* OR “over weight*” OR adiposit* OR corpulen* OR “weight loss” OR “weight reduction*” OR “weight management” OR “weight control” OR “body weight change*” OR antiobes* OR “anti-obes*”) OR AB (obes* OR overweight* OR “over weight*” OR adiposit* OR corpulen* OR “weight loss” OR “weight reduction*” OR “weight management” OR “weight control” OR “body weight change*” OR antiobes* OR “anti-obes*”)

S3 S1 OR S2

S4 (MH “Glucagon-Like Peptide-1 Agonists+”) OR (MH “Orlistat”)

S5 TI (“glucagon-like peptide-1” OR “GLP-1*” OR “GLP1*” OR incretin* OR semaglutide OR wegovy OR ozempic OR rybelsus OR liraglutide OR saxenda OR victoza OR tirzepatide OR zepbound OR mounjaro OR LY3298176 OR retatrutide OR LY3437943 OR orforglipron OR LY3502970 OR “maridebart cafraglutide” OR maridebart OR maritide OR “AMG 133” OR AMG133 OR mazdutide OR IBI362 OR survodutide OR “BI 456906” OR ecnoglutide OR XW003 OR bofanglutide OR noiiglutide OR SHR20004 OR VCT220 OR “HRS-7535” OR pemvidutide OR “ALT-801” OR danuglipron OR cotadutide OR efinopegdutide OR efpeglenatide OR albiglutide OR dulaglutide OR exenatide OR lixisenatide OR beinaglutide OR loxenatide OR amylin* OR cagrilintide OR AM833 OR CagriSema OR amycretin OR eloralintide OR pramlintide OR symlin OR petrelintide OR orlistat OR xenical OR alli OR “phentermine-topiramate” OR (phentermine AND topiramate) OR qsymia OR qsiva OR qnexa OR “naltrexone-bupropion” OR (naltrexone AND bupropion) OR contrave OR mysimba) OR AB (“glucagon-like peptide-1” OR “GLP-1*” OR “GLP1*” OR incretin* OR semaglutide OR wegovy OR ozempic OR rybelsus OR liraglutide OR saxenda OR victoza OR tirzepatide OR zepbound OR mounjaro OR LY3298176 OR retatrutide OR LY3437943 OR orforglipron OR LY3502970 OR “maridebart cafraglutide” OR maridebart OR maritide OR “AMG 133” OR AMG133 OR mazdutide OR IBI362 OR survodutide OR “BI 456906” OR ecnoglutide OR XW003 OR bofanglutide OR noiiglutide OR SHR20004 OR VCT220 OR “HRS-7535” OR pemvidutide OR “ALT-801” OR danuglipron OR cotadutide OR efinopegdutide OR efpeglenatide OR albiglutide OR dulaglutide OR exenatide OR lixisenatide OR beinaglutide OR loxenatide OR amylin* OR cagrilintide OR AM833 OR CagriSema OR amycretin OR eloralintide OR pramlintide OR symlin OR petrelintide OR orlistat OR xenical OR alli OR “phentermine-topiramate” OR (phentermine AND topiramate) OR qsymia OR qsiva OR qnexa OR “naltrexone-bupropion” OR (naltrexone AND bupropion) OR contrave OR mysimba)

S6 S4 OR S5

S7 (MH “Randomized Controlled Trials+”) OR (MH “Clinical Trials+”) OR PT “Randomized Controlled Trial” OR PT “Clinical Trial“

S8 TI (random* OR placebo* OR “double-blind*” OR “single-blind*” OR trial OR groups OR allocat* OR assign*) OR AB (random* OR placebo* OR “double-blind*” OR “single-blind*” OR trial OR groups OR allocat* OR assign*)

S9 S7 OR S8

S10 S3 AND S6 AND S9

#### 6. SinoMed / CBM

#1 “肥胖症“[加权:扩展] OR “超重“[加权:扩展] OR “体重减轻“[加权:扩展] OR “抗肥胖药“[加权:扩展]

#2 (“肥胖” OR “超重” OR “减重” OR “减肥” OR “体重管理” OR “体重控制” OR “体重下降” OR “体重减轻” OR “体质量下降” OR “体质量减轻”)[常用字段:智能]

#3 #1 OR #2

#4 (“胰高血糖素样肽-1” OR “GLP-1” OR “肠促胰素” OR “双靶点激动剂” OR “三靶点激动剂” OR “司美格鲁肽” OR “索马鲁肽” OR “诺和盈” OR “诺和泰” OR “诺和忻” OR “利拉鲁肽” OR “萨克森达” OR “诺和力” OR “替尔泊肽” OR “穆峰达” OR “瑞他鲁肽” OR “LY3437943” OR “奥福格列隆” OR “orforglipron” OR “LY3502970” OR “玛仕度肽” OR “IBI362” OR “IBI-362” OR “苏沃度肽” OR “survodutide” OR “BI456906” OR “伊诺格鲁肽” OR “ecnoglutide” OR “XW003” OR “博凡格鲁肽” OR “bofanglutide” OR “诺利格鲁肽” OR “noiiglutide” OR “SHR20004” OR “SHR-20004” OR “VCT220” OR “HRS-7535” OR “HRS9531” OR “贝那鲁肽” OR “菲塑美” OR “艾塞那肽” OR “度拉糖肽” OR “利司那肽” OR “洛塞那肽” OR “阿必鲁肽” OR “卡格列肽” OR “cagrilintide” OR “CagriSema” OR “普兰林肽” OR “胰淀素” OR “amycretin” OR “eloralintide” OR “maridebart” OR “AMG133” OR “pemvidutide” OR “danuglipron” OR “cotadutide” OR “TG103” OR “GZR18” OR “PB-119” OR “RAY1225” OR “UBT251” OR “BGM0504” OR “HDM1002” OR “奥利司他” OR “尼可” OR “芬特明” OR “托吡酯” OR “纳曲酮” OR “安 非他酮”)[常用字段:智能]

#5 “随机对照试验“[加权:扩展] OR “临床对照试验“[加权:扩展] OR (“随机” OR “对照” OR “安慰剂” OR “双盲” OR “单盲” OR “临床试验” OR “RCT”)[常用字段:智能]

#6 #3 AND #4 AND #5

#### 7. CNKI (China National Knowledge Infrastructure)

SU=(’肥胖’ + ‘超重’ + ‘减重’ + ‘减肥’ + ‘体重管理’ + ‘体重控制’ + ‘体重减轻’ + ‘体重下降’ + ‘体质量减轻’ + ‘体质量下降’) AND SU=(’GLP-1’ + ‘胰高血糖素样肽’ + ‘肠促胰素’ + ‘司美格鲁肽’ + ‘索马鲁肽’ + ‘利拉鲁肽’ + ‘替尔泊肽’ + ‘瑞他鲁肽’ + ‘retatrutide’ + ‘奥福格列隆’ + ‘orforglipron’ + ‘玛仕度肽’ + ‘mazdutide’ + ‘IBI362’ + ‘survodutide’ + ‘BI456906’ + ‘伊诺格鲁肽’ + ‘ecnoglutide’ + ‘XW003’ + ‘博凡格鲁肽’ + ‘bofanglutide’ + ‘诺利格鲁肽’ + ‘noiiglutide’ + ‘SHR20004’ + ‘VCT220’ + ‘HRS-7535’ + ‘HRS9531’ + ‘贝 那鲁肽’ + ‘艾塞那肽’ + ‘度拉糖肽’ + ‘利司那肽’ + ‘洛塞那肽’ + ‘卡格列肽’ + ‘cagrilintide’ + ‘CagriSema’ + ‘普兰林肽’ + ‘胰淀素’ + ‘amycretin’ + ‘eloralintide’ + ‘maridebart’ + ‘AMG133’ + ‘pemvidutide’ + ‘danuglipron’ + ‘cotadutide’ + ‘TG103’ + ‘GZR18’ + ‘RAY1225’ + ‘UBT251’ + ‘BGM0504’ + ‘HDM1002’ + ‘奥利司他’ + ‘芬特明’ + ‘托吡酯’ + ‘纳曲酮’ + ‘安非他酮’) AND FT=(’随机’ + ‘随机对照’ + ‘安慰剂’ + ‘双盲’ + ‘临床试验’ + ‘RCT’)

#### 8. Wanfang Data

题名或关键词:(“肥胖” OR “超重” OR “减重” OR “减肥” OR “体重管理” OR “体重控制” OR “体重减轻” OR “体重下降” OR “体质量减轻” OR “体质量下降”) AND 题名或关键词:(“GLP-1” OR “胰高血糖素样肽” OR “肠促胰素” OR “司美格鲁肽” OR “索马鲁肽” OR “利拉鲁肽” OR “替尔泊肽” OR “瑞他鲁肽” OR “retatrutide” OR “奥福格列隆” OR “orforglipron” OR “玛仕度 肽” OR “mazdutide” OR “IBI362” OR “survodutide” OR “BI456906” OR “伊诺格鲁肽” OR “ecnoglutide” OR “XW003” OR “博凡格鲁肽” OR “bofanglutide” OR “诺利格鲁肽” OR “noiiglutide” OR “SHR20004” OR “VCT220” OR “HRS-7535” OR “HRS9531” OR “贝那鲁肽” OR “艾塞那肽” OR “度拉糖肽” OR “利司那肽” OR “洛塞那肽” OR “卡格列肽” OR “cagrilintide” OR “CagriSema” OR “普兰林肽” OR “胰淀素” OR “amycretin” OR “eloralintide” OR “maridebart” OR “AMG133” OR “pemvidutide” OR “danuglipron” OR “cotadutide” OR “TG103” OR “GZR18” OR “RAY1225” OR “UBT251” OR “BGM0504” OR “HDM1002” OR “奥利司他” OR “芬特明” OR “托吡酯” OR “纳曲酮” OR “安非他酮”) AND 全部:(“随机” OR “安慰剂” OR “双盲” OR “临床试验” OR “RCT”)

#### 9. ClinicalTrials.gov

AREA[StudyType] EXPAND[None] COVER[FullMatch] “Interventional” AND AREA[ConditionSearch] (obesity OR overweight OR “weight loss” OR “weight reduction” OR “weight management” OR “body weight”) AND AREA[InterventionSearch] (“GLP-1” OR incretin OR semaglutide OR NN9535 OR liraglutide OR NN8022 OR tirzepatide OR LY3298176 OR retatrutide OR LY3437943 OR orforglipron OR LY3502970 OR “maridebart cafraglutide” OR maridebart OR cafraglutide OR “AMG 133” OR AMG133 OR mazdutide OR IBI362 OR LY3305677 OR survodutide OR “BI 456906” OR BI456906 OR ecnoglutide OR XW003 OR bofanglutide OR noiiglutide OR SHR20004 OR VCT220 OR “HRS-7535” OR HRS7535 OR HRS9531 OR pemvidutide OR “ALT-801” OR danuglipron OR “PF-06882961” OR lotiglipron OR cotadutide OR MEDI0382 OR efinopegdutide OR efpeglenatide OR albiglutide OR dulaglutide OR exenatide OR lixisenatide OR beinaglutide OR loxenatide OR amylin OR cagrilintide OR AM833 OR CagriSema OR amycretin OR “NNC0487-0111” OR eloralintide OR LY3841136 OR pramlintide OR davalintide OR petrelintide OR ZP8396 OR AZD6234 OR orlistat OR xenical OR alli OR phentermine OR topiramate OR qsymia OR qnexa OR “VI-0521” OR naltrexone OR bupropion OR contrave OR mysimba OR NB32 OR GMA106 OR “CT-388” OR “CT-996” OR VK2735 OR “GSBR-1290” OR ECC5004 OR AZD5004 OR HDM1002 OR TG103 OR GZR18 OR RAY1225 OR UBT251 OR BGM0504)

#### 10. WHO International Clinical Trials Registry Platform (WHO ICTRP)

Query 10A (Approved & Late-Stage GLP-1 / Dual / Triple Agonists):

Condition: obesity OR overweight OR weight loss OR weight management
Intervention: semaglutide OR liraglutide OR tirzepatide OR retatrutide OR LY3437943 OR orforglipron OR LY3502970 OR maridebart OR cafraglutide OR AMG 133 OR AMG133 OR mazdutide OR IBI362 OR survodutide OR BI 456906 OR BI456906

Query 10B (Investigational & Regional GLP-1-Related Agents):

Condition: obesity OR overweight OR weight loss OR weight management
Intervention: ecnoglutide OR XW003 OR bofanglutide OR noiiglutide OR SHR20004 OR VCT220 OR HRS-7535 OR HRS7535 OR HRS9531 OR pemvidutide OR ALT-801 OR danuglipron OR PF-06882961 OR lotiglipron OR cotadutide OR MEDI0382 OR efinopegdutide OR efpeglenatide OR beinaglutide OR loxenatide OR exenatide OR dulaglutide OR lixisenatide OR albiglutide

Query 10C (Emerging Oral/Multimodal Incretins):

Condition: obesity OR overweight OR weight loss OR weight management
Intervention: GMA106 OR CT-388 OR CT-996 OR VK2735 OR GSBR-1290 OR ECC5004 OR AZD5004 OR HDM1002 OR TG103 OR GZR18 OR PB-119 OR RAY1225 OR UBT251 OR BGM0504

Query 10D (Amylin-Related Agents & Co-formulations):

Condition: obesity OR overweight OR weight loss OR weight management
Intervention: amylin OR cagrilintide OR AM833 OR CagriSema OR amycretin OR NNC0487-0111 OR eloralintide OR LY3841136 OR pramlintide OR davalintide OR petrelintide OR ZP8396 OR AZD6234

Query 10E (Orlistat, Phentermine–Topiramate & Naltrexone–Bupropion):

Condition: obesity OR overweight OR weight loss OR weight management
Intervention: orlistat OR xenical OR alli OR tetrahydrolipstatin OR phentermine OR topiramate OR qsymia OR qsiva OR qnexa OR VI-0521 OR naltrexone OR bupropion OR contrave OR mysimba OR NB32

## Appendices 2-5. Completed PRISMA checklists

Four reporting checklists are provided: PRISMA 2020 (appendix 2), PRISMA 2020 for Abstracts (appendix 3), the PRISMA extension for network meta-analyses (appendix 4) and PRISMA-S, the extension for reporting literature searches (appendix 5). Each row gives the item, the requirement and the location in this submission where the item is reported. Locations refer to sections of the main manuscript, to Figure 1, and to the appendices: appendix 1 (search strategies), appendix 6 (data extraction form) and appendix 7 (eligibility criteria and screening outcomes).

Supplementary tables are cited as Table S1 to Table S25 and are provided in Supplementary_Tables.xlsx.

Registration: PROSPERO CRD420261532130, registered 7 October 2026. Searches were executed on 7 October 2026. The review was reported in accordance with PRISMA 2020, the PRISMA extension for network meta-analyses, and PRISMA-S.

*Where an item could not be reported because the corresponding step was not performed, the row says so explicitly rather than leaving the item blank*.

## Appendix 2. PRISMA 2020 checklist

| Section/Topic | # | Checklist Item | Location Where Item is Reported |
|---|---|---|---|
| **TITLE** |  |  |  |
| Title | 1 | Identify the report as a systematic review. | Partially reported: the title identifies the report as a network meta-analysis; the review is described as a systematic review in the Abstract and in Methods |
| **ABSTRACT** |  |  |  |
| Abstract | 2 | See PRISMA 2020 for Abstracts checklist. | Abstract (see Appendix Checklist 2) |
| **INTRODUCTION** |  |  |  |
| Rationale | 3 | Describe the rationale for the review in the context of existing knowledge. | Introduction P1–P2 |
| Objectives | 4 | Provide an explicit statement of the objective(s) or question(s) the review addresses. | Introduction P3 |
| **METHODS** |  |  |  |
| Eligibility criteria | 5 | Specify the inclusion and exclusion criteria for the review and how studies were grouped for the syntheses. | Methods › Eligibility criteria and treatment strategies P1–P2; Outcomes and estimands P3 (stratified by T2D status); Statistical analysis P1 (four-tier analysis); Evidence funnel P1; Table S1 (node dictionary) |
| Information sources | 6 | Specify all databases, registers, websites, organisations, reference lists and other sources searched or consulted to identify studies. Specify the date when each source was last searched or consulted. | Abstract › Methods (8 databases + 2 trial registries, from inception to October 2026); Figure 1 and legend (MEDLINE, Embase, CENTRAL, PsycINFO, CINAHL, SinoMed, CNKI, Wanfang, ClinicalTrials.gov, WHO ICTRP; reference lists, forward citation tracking, FDA/EMA regulatory reports, CSRs, and study protocols); Appendix 1 (sections 1.1-1.3) |
| Search strategy | 7 | Present the full search strategies for all databases, registers and websites, including any filters and limits used. | Appendix 1 (sections 1.1-1.3) |
| Selection process | 8 | Specify the methods used to decide whether a study met the inclusion criteria of the review, including how many reviewers screened each record and each report retrieved, whether they worked independently, and if applicable, details of automation tools used in the process. | Methods › Eligibility P1 (two reviewers independently screened records; discrepancies resolved via consensus or consultation with a third reviewer); Figure 1 (Level-0 dual independent screening; deduplication via automated cross-database matching and manual trial-family linkage); No automation tools were used for record screening or eligibility assessment |
| Data collection process | 9 | Specify the methods used to collect data from reports, including how many reviewers collected data from each report, whether they worked independently, any processes for obtaining or confirming data from study investigators, and if applicable, details of automation tools used in the process. | Methods › Outcomes and estimands P3 (two reviewers independently extracted or verified data; prioritised hierarchically by prespecification, degree of adjustment, and source reliability; digitised from plots when necessary); Figure 1 (CSRs and regulatory filings incorporated); Corresponding authors were written to for missing or unusable data; no additional data were provided in reply (Methods › Outcomes and estimands P3) |
| Data items | 10a | List and define all outcomes for which data were sought. Specify whether all results that were compatible with each outcome domain in each study were sought, and if not, the methods used to decide which results to collect. | Methods › Outcomes and estimands P1–P3 (time windows: 48–56, 10–14, and 20–28 weeks; estimand classification and hierarchical prioritisation) |
|  | 10b | List and define all other variables for which data were sought. Describe any assumptions made about any missing or unclear information. | Methods › Eligibility P2 (node definition components, lifestyle support components and intensity); Table S25 (country, study sites, trial registration identifier, study phase, population, baseline demographics, interventions, dose titration regimens, estimands, funding sources); Table S20 |
| Study risk of bias assessment | 11 | Specify the tool(s) used to assess risk of bias in included studies, how many reviewers assessed each study and whether they worked independently, and if applicable, details of automation tools used in the process. | Methods › Risk of bias and certainty of evidence P1 (Cochrane RoB 2 tool, dual independent review, evaluated by specific outcome) |
| Effect measures | 12 | Specify for each outcome the effect measure(s) used in the synthesis or presentation of results. | Methods › Outcomes and estimands P1 (MD in percentage points); Statistical analysis P2 (MD, OR, absolute risk, 95% CI, PI), P3–P4 (posterior median MD, 95% CrI, PrI, posterior probability of exceeding clinical threshold), P6 (ratio of MDs) |
| Synthesis methods | 13a | Describe the processes used to decide which studies were eligible for each synthesis. | Methods › Evidence funnel P1; Statistical analysis P1, P4 (observed/interpolated/borrowing-dependent/extrapolated classification); Table S24 |
|  | 13b | Describe any methods required to prepare the data for presentation or synthesis, such as handling of missing summary statistics, or data conversions. | Methods › Outcomes P3 (digitised from graphical plots); Statistical analysis P2 (reported, algebraic recovery, or imputed values of covariances; continuity corrections), P3 (assumed temporal correlation structures); Table S22 |
|  | 13c | Describe any methods used to tabulate or visually display results of individual studies and syntheses. | Figure 1-5 legends; Table 1 and Table 1B; Supplementary Figures S2-S8; Appendix 6 defines the record structure from which every table is generated |
|  | 13d | Describe any methods used to synthesize results and provide a rationale for the choice(s). If meta-analysis was performed, describe the model(s), method(s) to identify the presence and extent of statistical heterogeneity, and software package(s) used. | Methods › Statistical analysis P2 (frequentist contrast-based random-effects NMA, REML), P3 (multivariate normal likelihood, restricted cubic splines, partial within-class pooling, Stan, weakly informative priors), P5–P6; Table S21 (convergence diagnostics) |
|  | 13e | Describe any methods used to explore possible causes of heterogeneity among study results. | Methods › Outcomes P3 (subgroup stratification by T2D); Statistical analysis P2 (design-by-treatment interaction model, node-splitting, transitivity assessment), P6 (three-level hierarchical model); Table S10 |
|  | 13f | Describe any sensitivity analyses conducted to assess robustness of the synthesized results. | Methods › Statistical analysis P7; Tables S12, S13; Supplementary Figure S7 |
| Reporting bias assessment | 14 | Describe any methods used to assess risk of bias due to missing results in a synthesis. | Methods › Risk of bias P1 (ROB-MEN framework); Statistical analysis P7 (comparison-adjusted funnel plots and Egger's test for comparisons with ≥10 trials) |
| Certainty assessment | 15 | Describe any methods used to assess certainty (or confidence) in the body of evidence for an outcome. | Methods › Risk of bias P1 (CINeMA, GRADE minimally contextualised framework, 5 percentage-point clinical threshold); Table S23 |
| **RESULTS** |  |  |  |
| Study selection | 16a | Describe the results of the search and selection process, from the number of records identified in the search to the number of studies included in the review, ideally using a flow diagram. | Results › Study selection P1; Figure 1; Table S24; Appendix 7 |
|  | 16b | Cite studies that might appear to meet the inclusion criteria, but which were excluded, and explain why they were excluded. | Figure 1 (676 full-text reports and 130 records from other sources excluded with specific reasons, citing e.g., SELECT, LEAN, TRANSCEND-T2D-1, STEP 4, SURMOUNT-3, SURMOUNT-4, QUEENs); Table S24, row L4 (individual LOCF trials listed by name) |
| Study characteristics | 17 | Cite each included study and present its characteristics. | Table 1; Table 1B; Table S25; Table S2 |
| Risk of bias in studies | 18 | Present assessments of risk of bias for each included study. | Results › Risk of bias P1; Figure 2; Supplementary Figure S1 |
| Results of individual studies | 19 | For all outcomes, present, for each study: (a) summary statistics for each group and (b) an effect estimate and its precision. | Table S8 (direct pairwise comparative estimates by study); Table S14 (paired results per project/trial); Table S17 (results by treatment arm) |
| Results of syntheses | 20a | For each synthesis, briefly summarise the characteristics and risk of bias among contributing studies. | Results › Study selection P2; Risk of bias P1; Table 1 (row L4); Table S3 |
|  | 20b | Present results of all statistical syntheses conducted. | Results › Weight reduction at 52 weeks P1–P2; Incremental effects of dose escalation; Effects of titration schedules; Incremental effects of lifestyle support; Permanent discontinuation and safety; Weight-loss response without permanent discontinuation; Weight reduction in T2D; Figure 4; Tables S4, S5, S11, S14, S16–S19; Supplementary Figures S4, S8 |
|  | 20c | Present results of all investigations of possible causes of heterogeneity among study results. | Results › Reference-network consistency P1; Weight reduction in T2D P2; Tables S6, S7, S10; Supplementary Figure S6 |
|  | 20d | Present results of all sensitivity analyses conducted. | Results › Reference-network consistency P2; Tables S12, S13; Supplementary Figure S7 |
| Reporting biases | 21 | Present assessments of risk of bias due to missing results for each synthesis assessed. | Results › Risk of bias P1–P2 (ROB-MEN 27/71; missing results reported); Table S15; Figure 5; Table S9; Supplementary Figure S5 |
| Certainty of evidence | 22 | Present assessments of certainty in the body of evidence for each outcome assessed. | Results › Weight reduction at 52 weeks P2; Tables S5, S23 |
| **DISCUSSION** |  |  |  |
| Discussion | 23a | Provide a general interpretation of the results in the context of other evidence. | Discussion P1–P2 |
|  | 23b | Discuss any limitations of the evidence included in the review. | Discussion P3, P6, P7 |
|  | 23c | Discuss any limitations of the review processes used. | Discussion P7 |
|  | 23d | Discuss implications of the results for practice, policy, and future research. | Discussion P2, P4, P8–P9 |
| **OTHER INFORMATION** |  |  |  |
| Registration and protocol | 24a | Provide registration information for the review, including register name and registration number, or state that the review was not registered. | Methods › Study design P1 (PROSPERO CRD420261532130, registered 7 October 2026) |
|  | 24b | Indicate where the review protocol can be accessed, or state that a protocol was not prepared. | Registration record available at PROSPERO CRD420261532130 |
|  | 24c | Describe and explain any amendments to information provided at registration or in the protocol. | Methods › Study design P1 (new analyses added in v1.1, post-hoc analytical choices, binary outcome methodology deviating from protocol §50); Statistical analysis P4 (MariTide comparisons derived post hoc); Table S12 (42–78-week window designated as post hoc); Table S19 footnote |
| Support | 25 | Describe sources of financial or non-financial support for the review, and the role of the funders. | Funding; Role of the funding source |
| Competing interests | 26 | Declare any competing interests of review authors. | Declaration of interests (no competing interests declared) |
| Availability of data, code and other materials | 27 | Report which of the following are publicly available and where they can be found: data, code, and other materials. | Data, code and other materials (data, node dictionary, screening and extraction records and R/Stan analysis code available from the corresponding author on reasonable request; protocol available from PROSPERO CRD420261532130) |

## Appendix 3. PRISMA 2020 for Abstracts checklist

| Section | # | Checklist Item | Reported? / Location |
|---|---|---|---|
| **TITLE** | 1 | Identify the report as a systematic review. | Partially reported: the title identifies a network meta-analysis but does not name the review as a systematic review |
| **BACKGROUND** | 2 | Provide an explicit statement of the main objective(s) or question(s) the review addresses. | Yes: Background |
| **METHODS** | 3 | Specify the inclusion and exclusion criteria for the review. | Yes: Methods, sentences 1–2 (RCTs ≥12 weeks, overweight or obese adults, treatment initiation; maintenance, withdrawal, and switch trials excluded) |
|  | 4 | Specify the information sources searched and the date when each was last searched. | Yes: Methods, sentence 1 (8 databases, 2 trial registries, searched from inception through October 2026) |
|  | 5 | Specify the tool(s) used to assess risk of bias. | Partially reported: Methods, sentence 3 (states "assessed bias and certainty", without specifying individual tool names) |
|  | 6 | Specify the methods used to synthesize results. | Yes: Methods, sentence 4 (Bayesian longitudinal NMA, reference networks) |
| **RESULTS** | 7 | Provide the number and type of included studies and participants. | Yes: Findings, sentence 1 (180 reports, 178 trials, 196 randomised units, 81,014 participants) |
|  | 8 | For synthesised results, present the summary estimate and its precision for the main outcome(s). | Yes: Findings (MDs with 95% CrIs; permanent treatment discontinuation; dose escalation; T2D ratio 0·62) |
| **DISCUSSION** | 9 | Provide a brief summary of the limitations of the evidence included in the review. | Yes: Findings (only 2 strategies achieved high certainty; permanent discontinuation effects uncertain or missing); Interpretation, sentence 1 |
|  | 10 | Provide a general interpretation of the results and important implications. | Yes: Interpretation |
| **OTHER** | 11 | Specify the primary source of funding for the review. | Yes: Funding: None |
|  | 12 | Provide the register name and registration number. | Yes: Abstract › Methods (PROSPERO CRD420261532130, registered 7 October 2026) |

## Appendix 4. PRISMA-NMA checklist

| **Section/Topic** | **#** | **Checklist Item** | **Location Where Item is Reported** |
|---|---|---|---|
| **TITLE** | 1 | Identify the report as a systematic review incorporating a network meta-analysis. | Partially reported: the title identifies the report as a network meta-analysis and omits the systematic review descriptor |
| **ABSTRACT** | 2 | Provide a structured summary including network meta-analysis methods and findings. | Abstract |
| **INTRODUCTION** |  |  |  |
| Rationale | 3 | Describe the rationale for the review in the context of existing knowledge, including why a network meta-analysis is needed. | Introduction P1–P2 |
| Objectives | 4 | Provide an explicit statement of the questions addressed, structured according to PICOS. | Introduction P3 |
| **METHODS** |  |  |  |
| Protocol and registration | 5 | Indicate if a review protocol exists, whether and where it can be accessed, and provide registration information. | Methods › Study design P1 (PROSPERO CRD420261532130, registered 7 October 2026); registration record accessible at PROSPERO |
| Eligibility criteria | 6 | Specify the study characteristics and report characteristics used as criteria for eligibility, including how interventions were grouped into nodes. | Methods › Eligibility P1–P2 (nodes categorised by molecule, formulation, route, frequency, target maintenance dose, and titration schedule; MTD arms analysed by allocated intent; connector nodes); Table S1 |
| Information sources | 7 | Describe all information sources in the search and date of the last search. | Abstract › Methods; Figure 1; Appendix 1 |
| Search | 8 | Present the full electronic search strategy for at least one major database. | Appendix 1 (sections 1.1-1.3) |
| Study selection | 9 | State the process for selecting studies. | Methods › Eligibility P1; Figure 1; Appendix 7 |
| Data collection process | 10 | Describe the method of data extraction from reports. | Methods › Outcomes and estimands P3 |
| Data items | 11 | List and define all variables for which data were sought, including intervention details. | Methods › Eligibility P2; Outcomes P1–P3; Tables S1, S25 |
| Geometry of the network | S1 | Describe methods used to explore the geometry of the treatment network under study and potential biases related to it. | Methods › Statistical analysis P1 (disconnected networks evaluated separately); Figure 3 legend (node sizes scaled to sample size, edges weighted by trial count and overall RoB) |
| Risk of bias within studies | 12 | Describe methods used for assessing risk of bias in the included studies. | Methods › Risk of bias P1 (Cochrane RoB 2 tool) |
| Summary measures | 13 | State the principal summary measures, including any rankings. | Methods › Statistical analysis P2 (MD, OR, absolute risk), P4 (posterior median MD, CrI, PrI, posterior threshold probabilities; treatment rankings and SUCRA evaluated as secondary outcomes) |
| Planned methods of analysis | 14 | Describe the methods of handling multi-arm trials, evaluation of heterogeneity, choice of priors, and assessment of model fit. | Methods › Statistical analysis P2 (REML common heterogeneity, adjustment for multi-arm correlation), P3 (multivariate normal likelihood, spline interpolation, within-class pooling, trial-level random effects, weakly informative priors, Stan implementation); Tables S21, S22 |
| Assessment of inconsistency | S2 | Describe statistical methods used to evaluate the presence of inconsistency within the network. | Methods › Statistical analysis P2 (design-by-treatment interaction model, node-splitting method) |
| Risk of bias across studies | 15 | Specify any assessment of risk of bias that may affect the cumulative evidence. | Methods › Risk of bias P1 (ROB-MEN); Statistical analysis P7 (comparison-adjusted funnel plots, Egger's regression test) |
| Additional analyses | 16 | Describe methods of additional analyses (e.g., sensitivity, meta-regression, alternative models). | Methods › Statistical analysis P5 (dose escalation steps), P6 (Fréchet–Hoeffding bounds; three-level meta-regression), P7 (sensitivity analyses) |
| **RESULTS** |  |  |  |
| Study selection | 17 | Provide numbers of studies screened, assessed for eligibility, and included; flow diagram. | Results › Study selection P1; Figure 1 |
| Network structure | S3 | Present network graph of the included study interventions. | Figure 3; Supplementary Figures S2, S3 |
| Summary of network geometry | S4 | Provide a brief overview of network characteristics. | Results › Study selection P2 (46 nodes, 115 direct comparisons, 70 closed loops); Table S3 (nodes, trials, direct comparisons, multi-arm trials, subnetworks, density, and loop counts by outcome); Table 1 |
| Study characteristics | 18 | For each study, present characteristics for which data were extracted. | Table 1; Table 1B; Table S25 |
| Risk of bias within studies | 19 | Present data on risk of bias for each study. | Figure 2; Supplementary Figure S1 |
| Results of individual studies | 20 | For all outcomes, present the study-level summary data and effect estimates with precision. | Table S8; Supplementary Figure S4 |
| Synthesis of results | 21 | Present results of each network meta-analysis, including credible intervals and rankings. | Results › Weight reduction at 52 weeks; Permanent discontinuation and safety; Weight reduction in T2D; Figure 4; Tables S4, S5 (median rank, SUCRA), S11 (league table); Supplementary Figure S4 |
| Exploration for inconsistency | S5 | Present results from the assessment of inconsistency. | Results › Reference-network consistency P1 (global inconsistency test p=0·062; local loop inconsistency: liraglutide 3·0 mg vs placebo, tirzepatide 10 mg vs 5 mg); Tables S6, S7 |
| Risk of bias across studies | 22 | Present results of any assessment of risk of bias across studies. | Results › Risk of bias P1–P2; Table S15; Figure 5; Table S9; Supplementary Figure S5 |
| Results of additional analyses | 23 | Present results of additional analyses (sensitivity, meta-regression, dose-escalation steps, etc.). | Results › Reference-network consistency P2; Incremental effects of dose escalation; Effects of titration schedules; Incremental effects of lifestyle support; Weight-loss response without permanent discontinuation; Weight reduction in T2D P2; Tables S12–S14, S16–S20; Supplementary Figures S7, S8 |
| **DISCUSSION** |  |  |  |
| Summary of evidence | 24 | Summarize the main findings, including the strength and validity of the evidence in the network. | Discussion P1–P2 |
| Limitations | 25 | Discuss the limitations of the evidence, including issues of transitivity and inconsistency. | Discussion P7 |
| Conclusions | 26 | Provide a general interpretation of findings and implications for practice, policy, and research. | Discussion P8–P9 |
| **FUNDING** | 27 | Describe sources of funding and other support, and the role of funders. | Funding; Role of the funding source |

## Appendix 5. PRISMA-S checklist

| **#** | **Checklist Item** | **Reported Location / Content** |
|---|---|---|
| **1** | **Database name** | Appendix 1, sections 1.1 and 1.3; Figure 1: MEDLINE, Embase, Cochrane CENTRAL, APA PsycINFO, CINAHL Complete, SinoMed (CBM), CNKI, Wanfang Data |
| **2** | **Multi-database searching** | Appendix 1, section 1.3, sources 1-5: MEDLINE via PubMed; Embase via Ovid; APA PsycINFO via Ovid (Embase and PsycINFO searched independently); CINAHL via EBSCOhost; Cochrane CENTRAL using Cochrane Library syntax |
| **3** | **Study registries** | Appendix 1, section 1.3 (queries 10A-10E); Figure 1 |
| **4** | **Online resources and browsing** | Appendix 1, section 1.1; Figure 1: regulatory evaluation filings (FDA and EMA), clinical study reports and trial protocols (n=28) |
| **5** | **Citation searching** | Appendix 1, section 1.1; Figure 1: reference lists of included trials and prior systematic reviews (n=86); forward citation tracking (n=34) |
| **6** | **Contacts** | Corresponding authors were written to for missing or unusable data; no additional data were provided in reply (Methods › Outcomes and estimands P3) |
| **7** | **Other methods** | Reference lists of included trials and prior systematic reviews, forward citation tracking, and regulatory documents including clinical study reports and trial protocols (Appendix 1, section 1.1; Figure 1) |
| **8** | **Full search strategies** | Appendix 1, section 1.3 (complete line-by-line strategies for all ten sources, including registry queries 10A-10E) |
| **9** | **Limits and restrictions** | Appendix 1, sections 1.1-1.2; no language or publication-date restriction; MEDLINE lines #9-#10 and Embase lines 11-12 excluded non-human animal studies |
| **10** | **Search filters** | Appendix 1, section 1.3: MEDLINE line #8 (Cochrane highly sensitive search filter for RCTs); Embase lines 8-10; PsycINFO lines 5-7; CINAHL lines S7-S9; CENTRAL restricted to 'in Trials'; SinoMed line #5; CNKI restricted by 'FT' field; Wanfang restricted by 'All fields' randomisation and trial terms |
| **11** | **Prior work** | No separate prior-work search was undertaken |
| **12** | **Updates** | Not applicable: the search was executed once, on 7 October 2026 |
| **13** | **Dates of searches** | Appendix 1, section 1.1 (all sources searched on 7 October 2026, from database inception to that date) |
| **14** | **Peer review** | The search strategy was not independently peer reviewed |
| **15** | **Total records** | Appendix 1, section 1.2; Figure 1: bibliographic databases 44 942 (MEDLINE 10 329; Embase 15 482; CENTRAL 9 416; PsycINFO 943; CINAHL 2 385; SinoMed 1 864; CNKI 2 418; Wanfang 2 105); trial registries 3 828 (ClinicalTrials.gov 2 146; WHO ICTRP 1 682); other sources 148; total 48 770 |
| **16** | **Deduplication** | Appendix 1, section 1.2; Figure 1: automated cross-database matching on title, DOI and author-year removed 16 420 duplicate records (records managed in an EndNote library); manual trial-family linkage consolidated 3 436 associated reports; 28 914 unique records remained |

## Appendix 6. Data extraction form, field definitions and coding rules

This appendix reproduces the data extraction form used in the review, defines every field that was captured, lists the permissible values, and gives the priority order applied when the same item was reported in more than one place. The form was implemented as a relational workbook in which each of the eight record types below forms one table, linked by record_id, report_id, randomization_id, arm_id and rob_id. The completed workbook for all 180 reports, together with the derived analysis database, is available from the corresponding author on reasonable request (Data, code and other materials).

Two reviewers completed the form for every report independently. Disagreements were resolved against the source report and, where the source remained ambiguous, by a third reviewer. Every change made to a value after the first extraction pass was recorded in the reconciliation log (section 6.10), so that the final analysis could be audited field by field.

### 6.1 Structure of the extraction form

| **Record type** | **One row per** | **Primary key** | **Section** |
|---|---|---|---|
| Study / report | report (publication, registry entry, CSR or regulatory document) | record_id | 6.2 |
| Randomisation unit and arm | randomised arm within an independent randomisation unit | arm_id | 6.3 |
| Strategy (network node) | distinct treatment-initiation strategy | analysis_node_id | 6.4 |
| Continuous outcome record | arm x outcome x time point | record_id | 6.5 |
| Binary / safety outcome record | arm x event x time window | record_id | 6.6 |
| Contrast | treatment arm x comparator arm x outcome x window | record_id | 6.7 |
| Risk of bias | outcome-specific assessment | rob_id | 6.8 |
| Reconciliation log | each value changed or verified after extraction | entry_id | 6.10 |

### 6.2 Study and report level (one row per report)

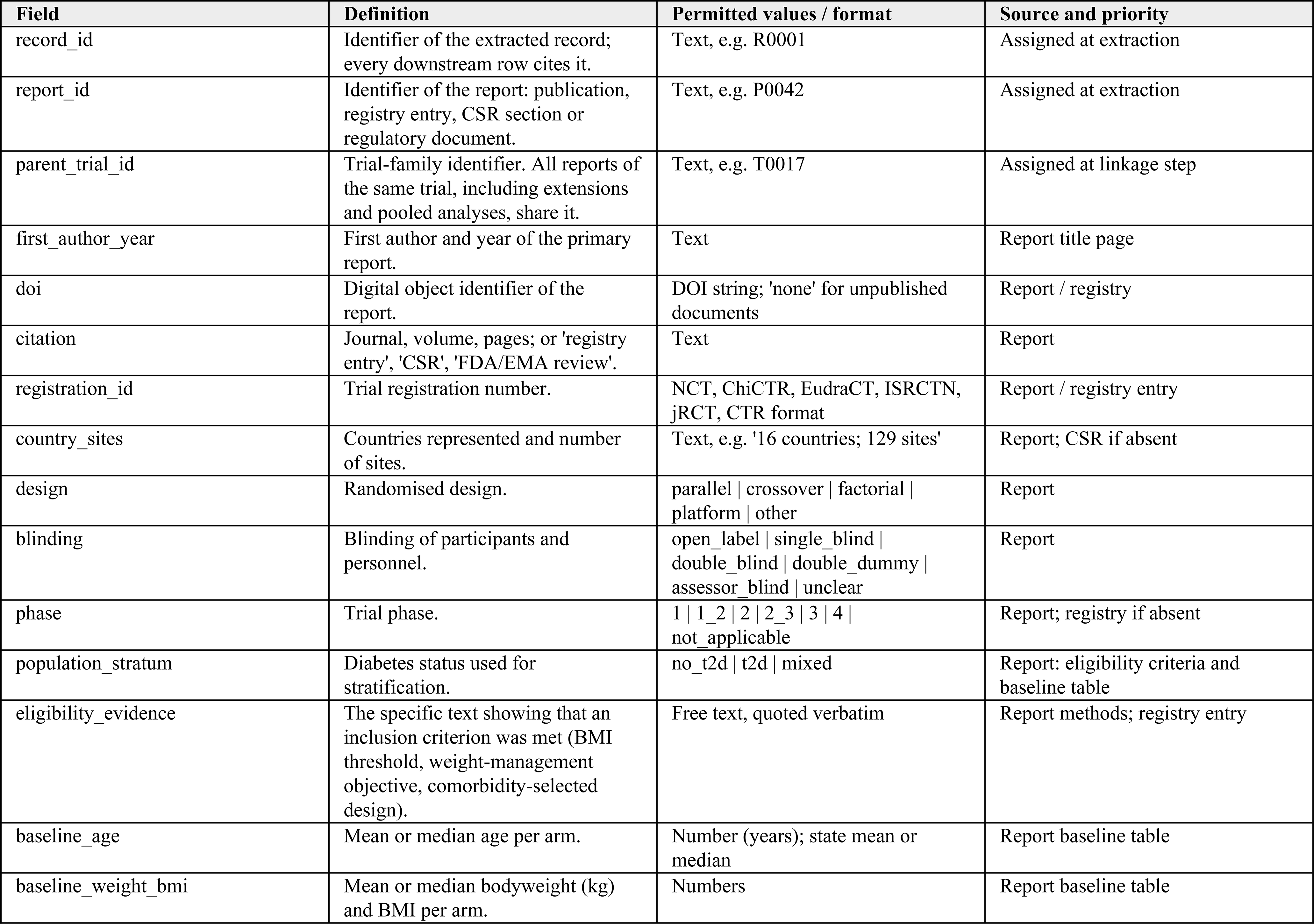
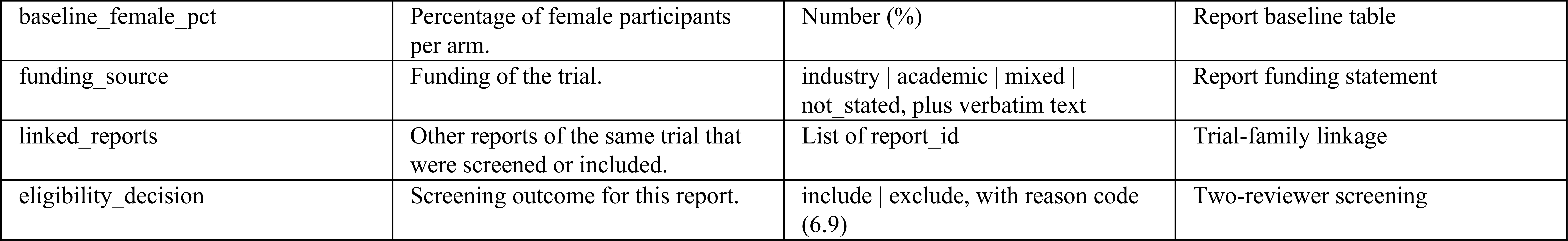

### 6.3 Randomisation unit and arm level (one row per arm)

| Field | Definition | Permitted values / format | Source and priority |
|---|---|---|---|
| arm_id | Identifier of the arm. | Text, e.g. A0311 | Assigned at extraction |
| randomization_id | Independent randomisation unit. Independently randomised cohorts within one report receive separate identifiers. | Text, e.g. Z0031 | Assigned at linkage step |
| origin_randomization_id | For extension, withdrawal or pooled analyses: the randomisation unit in which participants were originally allocated. | Text or blank | Trial-family linkage |
| dependency_group_id | Groups arms and records that must never enter the same synthesis (shared participants). | Text or blank | Trial-family linkage |
| arm_label_original | Arm label exactly as printed, including dose and regimen wording. | Text, verbatim | Report table or figure |
| arm_order | Position of the arm as reported. | Integer | Report |
| arm_type | Nature of the arm. | randomized_arm \| active_control_arm \| withdrawn_agent_comparator \| published_pooled_analysis_arm | Judgement against eligibility criteria |
| randomized_n | Number randomised. | Integer | Report; registry if absent |
| efficacy_n | Number in the efficacy analysis set used for the reported estimate. | Integer | Report, defined by the trial's own analysis set |
| safety_n | Number in the safety set. | Integer | Report |
| completion_n | Number completing the planned follow-up. | Integer | Report or participant flow figure |
| treatment_id | Link to the strategy node (6.4). | analysis_node_id | Node coding rules (Table S1) |
| layer | Analysis layer to which the arm contributes. | Layer_A \| Layer_B \| Layer_C \| Layer_D | Protocol section on layers |
| is_target_intervention | Whether the strategy is a target initiation strategy rather than a connector. | yes \| no | Eligibility criteria |

### 6.4 Strategy (network node) level (one row per strategy)

A strategy, not a molecule, was the unit of analysis. Each node was defined by the seven components below; two arms entered the same node only if all applicable components matched.

| Field | Definition | Permitted values / format | Source and priority |
|---|---|---|---|
| analysis_node_id | Identifier of the strategy node. | Text, e.g. tirzepatide_sc_15mg_qw | Node coding |
| node_label | Display label. | Text, e.g. 'TZP SC 15mg QW' | Node coding |
| drug_component | Molecule or fixed combination, including companion components. | Text | Report |
| dose | Maintenance dose with unit. | Number + unit | Report |
| target_dose | Target maintenance dose the titration aimed at. | Number + unit | Report; titration schedule |
| route | Route of administration. | subcutaneous \| oral \| intramuscular \| intravenous \| transdermal \| other | Report |
| frequency | Dosing frequency. | qd \| bid \| tiw \| qw \| q2w \| q4w \| monthly \| other | Report |
| titration_policy | Escalation schedule to the maintenance dose, described by step size and interval. | Free text plus controlled label, e.g. standard_4w_escalation, fixed_maintenance, 2w_step_escalation, aggregated_maintenance | Report; titration figure or text |
| support_condition | Background lifestyle support that applied to the arm. | shared_background_support \| randomized_support_increment \| standard_advice_support \| counselling_support \| meal_replacement_support \| routine_care | Report; Table S20 |
| exchangeability_basis | The clinical rationale for treating these arms as exchangeable with the rest of the node. | Text, e.g. Path_B_molecule_plus_target_dose, Titration_Regimen_Layer_D, Monotherapy_component_comparator | Adjudicated by two reviewers |
| is_network_connector | Whether the node only connects otherwise disconnected strategies. | yes \| no | Node coding |
| active | Whether the node is retained in the final network. | yes \| no | Node coding |

### 6.5 Continuous outcome record (one row per arm, outcome and time point)

| Field | Definition | Permitted values / format | Source and priority |
|---|---|---|---|
| record_id | Identifier of the record. | Text | Assigned at extraction |
| outcome_code | Protocol outcome. | weight_change_pct \| weight_change_kg \| weight_endpoint_kg \| waist_change_cm \| waist_change_pct \| bmi_change \| iwqol_lite_ct_pf_change \| sf36v2_pf_change \| sf36_pf_change \| walk6min_change_m \| other | Protocol outcome list |
| outcome_definition_verbatim | The outcome as the trial defined it, quoted. | Text, verbatim | Report |
| time_original | Time point as reported. | Text, e.g. 'week 68' | Report |
| time_weeks | Time point converted to weeks from randomisation. | Number (weeks) | Converted; conversion recorded in log |
| data_role | Whether the value is a change from baseline or an absolute value. | change \| endpoint | Report |
| analysis_population | Analysis set used for the reported value. | full_analysis_set \| intent_to_treat \| itt_all_randomized \| modified_intent_to_treat \| mitt_population \| completers \| in_trial_observed \| on_treatment_observed \| observed_data \| evaluable \| per_protocol \| safety_analysis_set \| other | Report, defined by the trial |
| estimand_class | Estimand implied by the trial's handling of discontinuation, rescue, switching and missing data. | treatment_policy \| treatment_regimen \| modified_treatment_regimen \| hypothetical \| trial_product \| completer \| mitt_mmrm \| historical_imputation \| locf \| observed \| per_protocol \| md | Adjudicated from methods, not from terminology |
| n | Number analysed for this value. | Integer | Report |
| mean | Mean change or mean value. | Number | Report |
| sd | Standard deviation. | Number | Report |
| se | Standard error, where SD was not reported. | Number | Report; converted if needed |
| unit | Unit of the value. | percentage_point \| kg \| cm \| kg/m2 \| mmHg \| points \| m | Report |
| value_source | Where the number came from. | table \| text \| figure_digitised \| csr \| regulatory_document \| derived | Source hierarchy (6.9) |
| analysis_role | Whether the record enters the main, sensitivity or descriptive analysis. | main_ready \| sensitivity_ready \| descriptive_ready \| quarantined | Analysis selection |

### 6.6 Binary and safety outcome record (one row per arm, event and window)

| Field | Definition | Permitted values / format | Source and priority |
|---|---|---|---|
| record_id | Identifier of the record. | Text | Assigned at extraction |
| outcome_code | Binary or safety outcome. | weight_response_ge5 \| weight_response_ge10 \| weight_response_ge15 \| weight_response_ge20 \| weight_response_ge25 \| ae_permanent_discontinuation \| allcause_permanent_discontinuation \| sae \| nausea \| vomiting \| diarrhoea \| constipation \| gi_any \| other | Protocol outcome list |
| outcome_definition_verbatim | The event as the trial defined it, quoted, including whether it refers to the drug, the treatment or the study. | Text, verbatim | Report |
| action_type | The action the event describes. | none \| permanent_treatment_discontinuation \| treatment_discontinuation | Report |
| permanent_status | Whether permanence could be established from the report. | yes \| no \| not_established \| not_applicable | Report; 'not_established' recorded rather than assumed |
| event_n | Number with the event. | Integer | Report |
| denominator_n | Denominator used by the trial for this event. | Integer | Report |
| analysis_population | Analysis set. | safety_set \| full_analysis_set \| full_randomized_flow \| full_randomized_treatment_flow \| completer_safety_set \| treated_set \| other | Report |
| missing_data_rule | Rule used by the trial for missing binary data. | observed_only \| multiple_imputation \| logistic_regression_mi \| logistic_regression_j2r_mi \| locf \| hybrid_imputation \| nonresponder_imputation \| nri \| g_computation \| cochran_mantel_haenszel \| rubin_rule_imputed \| other | Report |
| estimand_class | As in 6.5. | As in 6.5 | Adjudicated from methods |
| time_window_original | Window as reported. | Text | Report |
| time_weeks | Window converted to weeks. | Number | Converted; recorded in log |
| safety_window_category | Standardised window. | w52_strict \| w52_broad \| w24 \| w12 \| long_term \| other_intermediate | Protocol windows |
| dependency_group_id | As in 6.3; prevents double counting of overlapping event sets. | Text or blank | Trial-family linkage |
| analysis_role | As in 6.5. | main_ready \| sensitivity_ready \| descriptive_ready \| quarantined | Analysis selection |

### 6.7 Contrast record (one row per comparison, outcome and window)

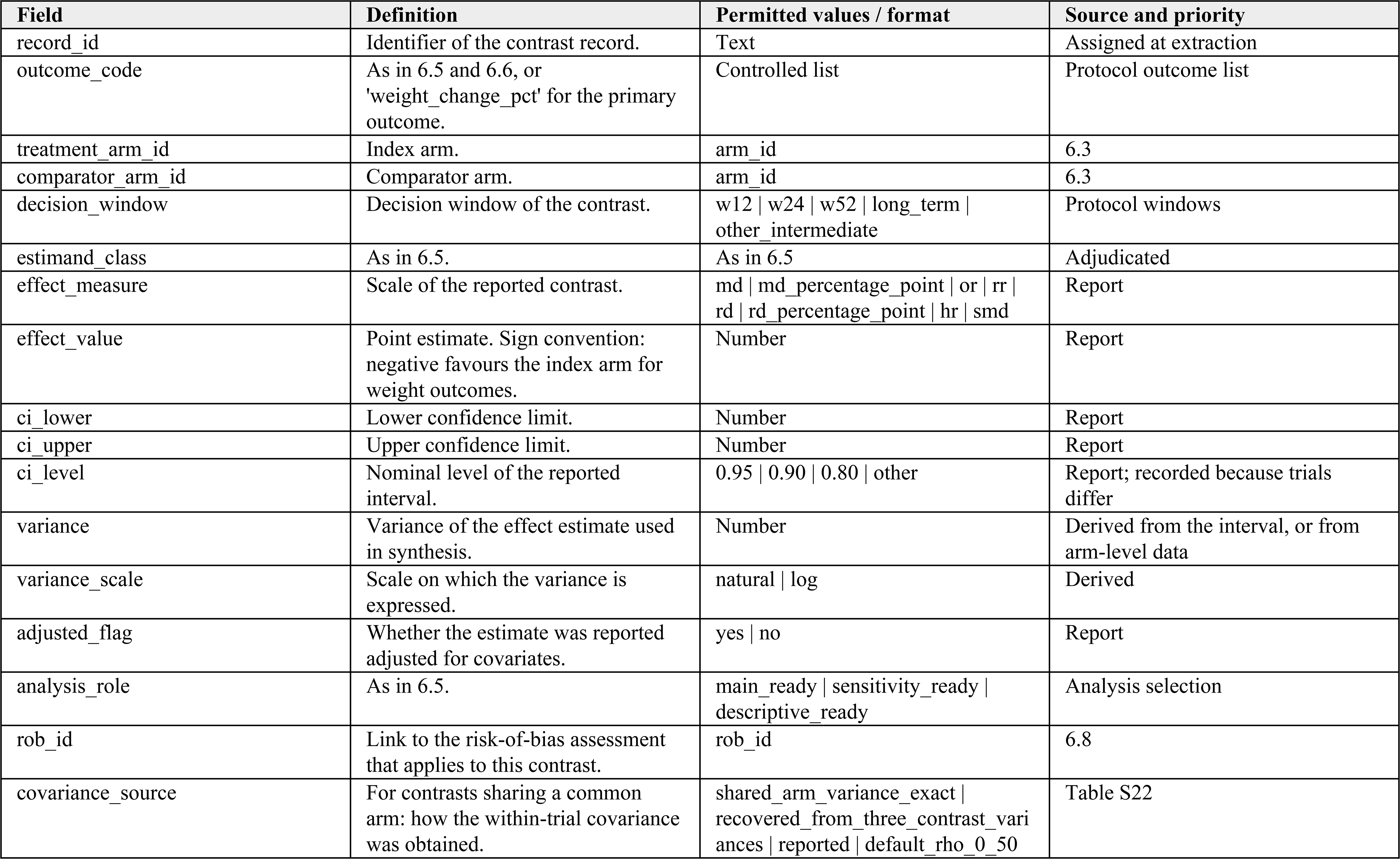

| Field | Definition | Permitted values / format | Source and priority |
|---|---|---|---|
| record_id | Identifier of the contrast record. | Text | Assigned at extraction |
| outcome_code | As in 6.5 and 6.6, or 'weight_change_pct' for the primary outcome. | Controlled list | Protocol outcome list |
| treatment_arm_id | Index arm. | arm_id | 6.3 |
| comparator_arm_id | Comparator arm. | arm_id | 6.3 |
| decision_window | Decision window of the contrast. | w12 \| w24 \| w52 \| long_term \| other_intermediate | Protocol windows |
| estimand_class | As in 6.5. | As in 6.5 | Adjudicated |
| effect_measure | Scale of the reported contrast. | md \| md_percentage_point \| or \| rr \| rd \| rd_percentage_point \| hr \| smd | Report |
| effect_value | Point estimate. Sign convention: negative favours the index arm for weight outcomes. | Number | Report |
| ci_lower | Lower confidence limit. | Number | Report |
| ci_upper | Upper confidence limit. | Number | Report |
| ci_level | Nominal level of the reported interval. | 0.95 \| 0.90 \| 0.80 \| other | Report; recorded because trials differ |
| variance | Variance of the effect estimate used in synthesis. | Number | Derived from the interval, or from arm-level data |
| variance_scale | Scale on which the variance is expressed. | natural \| log | Derived |
| adjusted_flag | Whether the estimate was reported adjusted for covariates. | yes \| no | Report |
| analysis_role | As in 6.5. | main_ready \| sensitivity_ready \| descriptive_ready | Analysis selection |
| rob_id | Link to the risk-of-bias assessment that applies to this contrast. | rob_id | 6.8 |
| covariance_source | For contrasts sharing a common arm: how the within-trial covariance was obtained. | shared_arm_variance_exact \| recovered_from_three_contrast_variances \| reported \| default_rho_0_50 | Table S22 |

### 6.8 Risk-of-bias record (RoB 2, outcome-specific)

| Field | Definition | Permitted values / format | Source and priority |
|---|---|---|---|
| rob_id | Identifier of the assessment. | Text | Assigned at assessment |
| scope | Outcome family to which the judgement applies. | continuous_efficacy \| binary_weight_response \| safety_or_discontinuation | RoB 2 guidance |
| result_ref | The result assessed, identified by record_id. | Text | 6.5 / 6.6 / 6.7 |
| D1_randomisation | Bias arising from the randomisation process. | low \| some_concerns \| high | RoB 2 signalling questions |
| D2_deviations | Bias due to deviations from intended interventions. | low \| some_concerns \| high | RoB 2 signalling questions |
| D3_missing_data | Bias due to missing outcome data. | low \| some_concerns \| high | RoB 2 signalling questions |
| D4_measurement | Bias in measurement of the outcome. | low \| some_concerns \| high | RoB 2 signalling questions |
| D5_selection | Bias in selection of the reported result. | low \| some_concerns \| high | RoB 2 signalling questions |
| overall | Overall risk of bias for this result. | low \| some_concerns \| high | RoB 2 algorithm |
| basis_codes | Coded justification for each domain judgement. | Codes from the RoB 2 basis codebook (8.ROB2.xlsx, sheet RoB2_BasisCodebook) | Free text plus code |

### 6.9 Coding rules applied during extraction

- Estimand class was assigned from what the trial did, not from the words it used: the handling of treatment discontinuation, rescue medication, switching and missing data determined the class. ‘treatment_policy’ was assigned when participants who stopped the assigned strategy remained in the analysis under that strategy.
- A value was assigned to ‘trial_product’ when the reported estimate reflected the effect of taking the drug as dispensed rather than the effect of being assigned to the strategy, as in trial-product estimands used in some phase 2 and head-to-head programmes.
- Analysis sets were taken from the trial and were never re-defined by the reviewers. Where a trial reported several sets for the same outcome, all were extracted and ‘analysis_role’ determined which entered the main analysis.
- Time points were converted to weeks from randomisation and were never rounded into a window. Observations falling outside 48-56 weeks retained their actual time and entered the longitudinal model, but not the 48-56-week reference network.
- Figure-derived values were used only when no tabular or textual value existed. Two reviewers digitised the same plot independently; a difference of more than 0.2 percentage points triggered re-measurement, and every digitised value was flagged as ‘figure_digitised’ and re-examined in the sensitivity analysis that excluded figure-derived records.
- Missing data were recorded as missing. Denominators, windows and permanence of discontinuation were never inferred from a related publication or from another arm.
- Source hierarchy, highest first: (1) tabulated results in the primary report; (2) results stated in the primary report text; (3) clinical study report or regulatory review; (4) conference abstract or registry results posting; (5) figure-derived value; (6) value algebraically recovered from other reported quantities, with the derivation recorded.

#### Controlled vocabulary for exclusion and isolation codes

Each excluded report and each isolated record carried one of the following codes, so that the number of records at every stage of Figure 1 can be reconstructed from the form itself.

| **Field** | **Definition** | **Permitted values / format** | **Source and priority** |
|---|---|---|---|
| estimand_trial_product | Estimand incompatible with the reference network | Code | Screening / analysis selection |
| estimand_completer | Completer or per-protocol analysis only | Code | Analysis selection |
| estimand_historical_imputation | Historical LOCF or similar imputation | Code | Analysis selection |
| withdrawn_agent_sibutramine | Agent withdrawn from the market | Code | Analysis selection |
| withdrawn_agent_rimonabant | Agent withdrawn from the market | Code | Analysis selection |
| pooled_phase2_isolated | Pooled phase 2 analysis isolated to avoid double counting | Code | Analysis selection |
| extension_cohort_isolation | Extension cohort isolated to avoid double counting | Code | Analysis selection |
| study_selection_mash_population | Population outside the review's eligibility | Code | Screening |
| lifestyle_comparator_unshared | Support comparator not shared across arms | Code | Analysis selection |
| estimand_efficacy_mmrm | Efficacy-type MMRM estimand | Code | Analysis selection |
| descriptive_ready_isolated | Descriptive only; isolated from synthesis | Code | Analysis selection |

### 6.10 Reconciliation and conversion log (one row per change)

The log records every value that was changed after the first extraction pass, every online verification, and every conversion. It is the audit trail for section 6.9.

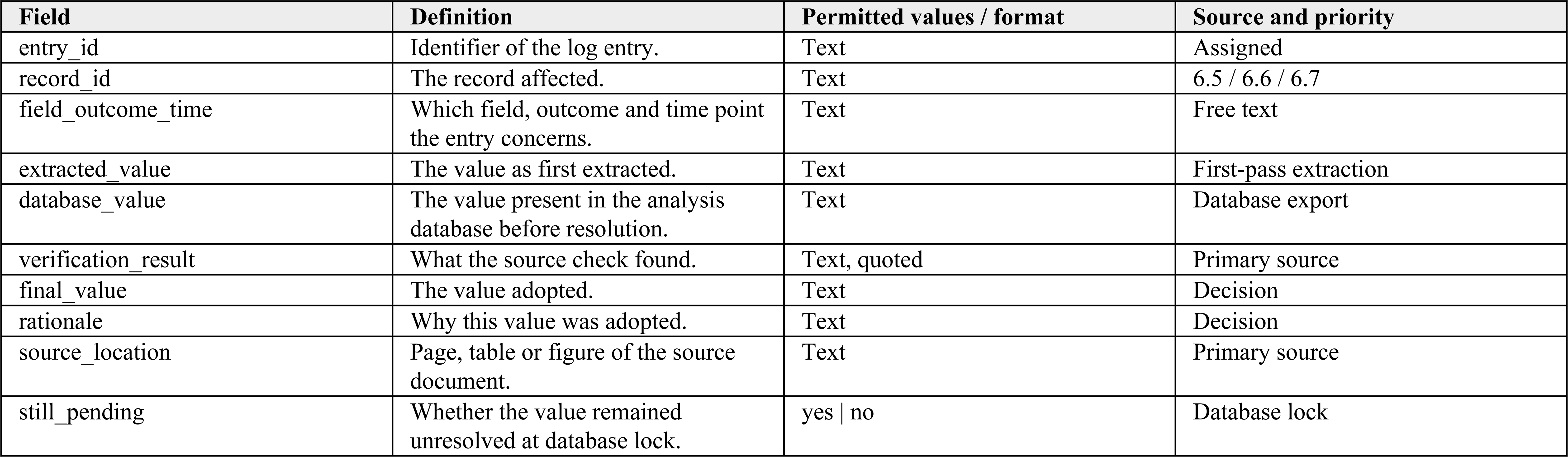

| Field | Definition | Permitted values / format | Source and priority |
|---|---|---|---|
| entry_id | Identifier of the log entry. | Text | Assigned |
| record_id | The record affected. | Text | 6.5 / 6.6 / 6.7 |
| field_outcome_time | Which field, outcome and time point the entry concerns. | Text | Free text |
| extracted_value | The value as first extracted. | Text | First-pass extraction |
| database_value | The value present in the analysis database before resolution. | Text | Database export |
| verification_result | What the source check found. | Text, quoted | Primary source |
| final_value | The value adopted. | Text | Decision |
| rationale | Why this value was adopted. | Text | Decision |
| source_location | Page, table or figure of the source document. | Text | Primary source |
| still_pending | Whether the value remained unresolved at database lock. | yes \| no | Database lock |

### 6.11 Worked example

The example below shows one strategy node and one contrast, completed from the SURMOUNT-1 report of tirzepatide in adults with obesity. Values are those used in the review and can be checked against Table S1 and Table S5. Arm-level rows (6.3, 6.5, 6.6) follow the same pattern and are omitted here for brevity.

| Field | Definition | Permitted values / format | Source and priority |
|---|---|---|---|
| analysis_node_id | Strategy node | tirzepatide_sc_15mg_qw | Table S1 |
| node_label | Display label | TZP SC 15mg QW | Table S1 |
| drug_component | Molecule | Tirzepatide (GIP/GLP-1 dual agonist) | Report |
| dose / target_dose | Maintenance dose | 15 mg | Report |
| route / frequency | Administration | subcutaneous / qw | Report |
| titration_policy | Escalation schedule | Escalated in 4-week steps to the target dose | Report |
| support_condition | Background support | shared_background_support | Table S20 |
| exchangeability_basis | Grouping rationale | Path_B_molecule_plus_target_dose | Node coding |
| is_target_intervention | Target strategy | yes | Table S1 |
| effect_measure | Contrast measure | md (percentage points) | Table S5 |
| effect_value (95% CrI) | 52-week placebo-adjusted mean difference | -16.87 (-18.07 to -15.51) | Table S5 |
| P(MD < -5 pp) / P(MD < -10 pp) | Posterior probability of clinical importance | 1.0000 / 1 | Table S5 |
| support_status | Week 52 support classification | observed or interpolated at 52 weeks (non-extrapolated) | Table S5 |
| certainty | CINeMA/GRADE certainty | High | Tables S5, S23 |

### 6.12 Relationship to the supplementary tables

Each analysis-ready table in the appendix is a view of this form rather than a separate dataset. Table S1 is the strategy dictionary of section 6.4; Tables S2, S3, S24 and S25 summarise sections 6.2 and 6.3; Tables S4, S5, S11 and S12 are built from section 6.7; Tables S8 and S14 from sections 6.7 and 6.5; Tables S15 to S19 from section 6.6; Table S22 from the covariance field of section 6.7; and Table S23 from section 6.8. Any value in a supplementary table can therefore be traced back to a source location through the record identifiers in this form and the log in section 6.10.

## Appendix 7. Eligibility criteria, screening process and exclusion decisions

### 7.1 How the criteria were applied

Two reviewers independently screened every record at title and abstract level against the criteria in sections 7.2 and 7.3. Records not excluded at that stage were retrieved in full text and assessed independently by the same two reviewers against the same criteria, together with the treatment-initiation criteria that determine which stage of treatment a trial addresses. Disagreements were resolved by discussion against the source record and, where the disagreement persisted, by a third reviewer. Each decision was recorded in the screening log with a decision (include or exclude), one or more criterion identifiers, and the principal reason shown in Figure 1, so that every number in the flow diagram can be traced to individual records.

Reports were then linked to parent trials. Independently randomised cohorts within one trial programme were counted as separate randomisation units, and extension cohorts, pooled analyses and secondary reports were identified as dependent records so that the same participants could not enter a synthesis twice.

### 7.2 Inclusion criteria

| # | Criterion | Specification | How it was verified |
|---|---|---|---|
| I1 | Study design | Randomised controlled trial, including open-label and multi-arm designs | Design statements in the report and registry entry; non-randomised designs, crossover trials without first-period data, reviews and commentaries excluded |
| I2 | Duration | Planned randomised treatment of at least 12 weeks in the randomised comparison of interest | Planned treatment duration in the methods or registry entry; run-in and follow-up periods after planned treatment were not counted |
| I3 | Population | Adults (aged 18 years or older) with overweight or obesity | Eligibility text and baseline table of the report; body-mass index criteria recorded verbatim for each trial |
| I4 | Objective | An explicit weight-management objective | Stated primary or key secondary objective; trials in which weight was measured but not a stated objective were excluded |
| I5 | Comorbidity-selected trials | Eligible when weight management was prespecified as an objective | Prespecified weight endpoint in the trial protocol or registry entry, not a post hoc analysis |
| I6 | Interventions | GLP-1-related agents, amylin-related agents, orlistat, phentermine-topiramate or naltrexone-bupropion, including investigational agents | Intervention description; agents outside these families excluded irrespective of weight effect |
| I7 | Treatment stage | Initiation of a pharmacotherapy strategy | Whether participants were randomised at the start of treatment rather than after induction, during maintenance or at withdrawal |

### 7.3 Exclusion criteria

| **#** | **Exclusion criterion** | **What it covers** | **Stage applied** |
|---|---|---|---|
| E1 | Randomised withdrawal | Trials randomising participants who had already lost weight to continued drug versus placebo or withdrawal (for example STEP 4, SURMOUNT-4) | Title/abstract and full text |
| E2 | Switching or de-escalation | Trials comparing a switch between agents, or a reduction in dose, as the randomised comparison | Title/abstract and full text |
| E3 | Post-bariatric management | Trials enrolling participants after bariatric surgery | Title/abstract and full text |
| E4 | Randomisation after active weight-loss induction | Trials in which all participants first completed a lead-in weight-loss phase (for example SURMOUNT-3) | Full text |
| E5 | Lifestyle-only trials | Trials without a pharmacological arm | Title/abstract |
| E6 | Ineligible population | Children and adolescents, type 1 diabetes, syndromic or monogenic obesity, populations defined by a non-weight condition without a weight-management objective | Title/abstract and full text |
| E7 | Planned randomised treatment shorter than 12 weeks | Trials whose randomised comparison was planned to last less than 12 weeks | Full text |
| E8 | Out-of-scope drug family | Agents outside the eligible families, including withdrawn agents for which no eligible target arm existed | Full text |
| E9 | Non-initiation or non-randomised design | Crossover trials without first-period data, single-arm studies, pooled analyses without a randomised comparison | Full text |
| E10 | Duplicate or dependent report | Duplicate publications, superseded abstracts and reports overlapping an already included cohort; retained in the trial family but not counted as a separate randomisation unit | Full text |

### 7.4 Screening stages and numbers

| **Stage** | **Records or reports** | **Derivation** |
|---|---|---|
| Identified in bibliographic databases | 44 942 | Eight databases, searched 7 October 2026 (appendix 1) |
| Identified in trial registries | 3 828 | ClinicalTrials.gov and WHO ICTRP |
| Identified through other sources | 148 | Reference lists (86), forward citation tracking (34), regulatory documents and protocols (28) |
| Removed before screening | 19 856 | 16 420 duplicates removed by automated cross-database matching; 3 436 associated reports consolidated by manual trial-family linkage |
| Records screened at title and abstract | 28 914 | After deduplication |
| Excluded at title and abstract | 28 072 | Table 7.1 |
| Reports sought for retrieval | 990 | 842 from databases and registries; 148 from other sources |
| Reports not retrieved | 4 | Unpublished historical appendices |
| Reports assessed for eligibility | 986 | 842 from databases and registries; 144 from other sources |
| Excluded at full-text stage | 806 | 676 from databases and registries (Table 7.2); 130 from other sources |
| **Included** | **180 reports of 178 parent trials** | **196 independent randomisation units; 81 014 participants** |

**Table 7.1.** Reasons for exclusion at title and abstract stage (n = 28 072)

| **Reason** | **n** |
|---|---|
| Irrelevant population, condition or non-obesity focus | 14 820 |
| Ineligible intervention or non-pharmacological study | 6 914 |
| Non-randomised study, review, commentary or animal study | 6 338 |
| **Total** | **28 072** |

**Table 7.2.** Reasons for exclusion at full-text stage, databases and registries (n = 676)

| **Principal reason** | **Examples** | **n** |
|---|---|---|
| Planned randomised treatment shorter than 12 weeks |  | 109 |
| No explicit weight-management objective, or population below overweight/obesity criteria | SELECT, LEAN, TRANSCEND-T2D-1 | 156 |
| Out-of-scope drug family without an eligible target arm | sibutramine, lorcaserin, rimonabant monotherapy | 58 |
| Non-initiation design: randomised withdrawal after a drug run-in | SURMOUNT-3, SURMOUNT-4, QUEENs | 94 |
| Duplicate publication, superseded abstract or overlapping cohort |  | 63 |
| Ineligible population | paediatric, type 1 diabetes, post-bariatric surgery | 52 |
| Crossover trial without first-period data, or non-randomised design |  | 31 |
| Other reasons, recorded individually in the screening log and not displayed separately in Figure 1 |  | 113 |
| **Total** |  | **676** |

Table 7.1 lists the reasons displayed in Figure 1. Table 7.2 lists the reasons displayed in Figure 1 together with the reports excluded for other reasons recorded individually in the screening log, giving the 676 full-text exclusions shown in the figure. The 130 reports excluded at the same stage but identified through other sources (regulatory documents, protocols and citation searching) were excluded for the same reasons, recorded with the same criterion identifiers in the screening log.

### 7.5 Reproducing Figure 1

Every number in Figure 1 can be reconstructed from the screening log, in which each record carries: a stable record identifier; the source in which it was found; the screening decision at each stage; the criterion identifiers from sections 7.2 and 7.3 that applied; the principal reason as listed in Tables 7.1 and 7.2; the parent trial to which the record was linked; and, for included records, the randomisation unit it belongs to. The same identifiers appear in the data extraction form (appendix 6), so that any value in any supplementary table can be traced to the report, page and table from which it was extracted.

---

**Table S1.**
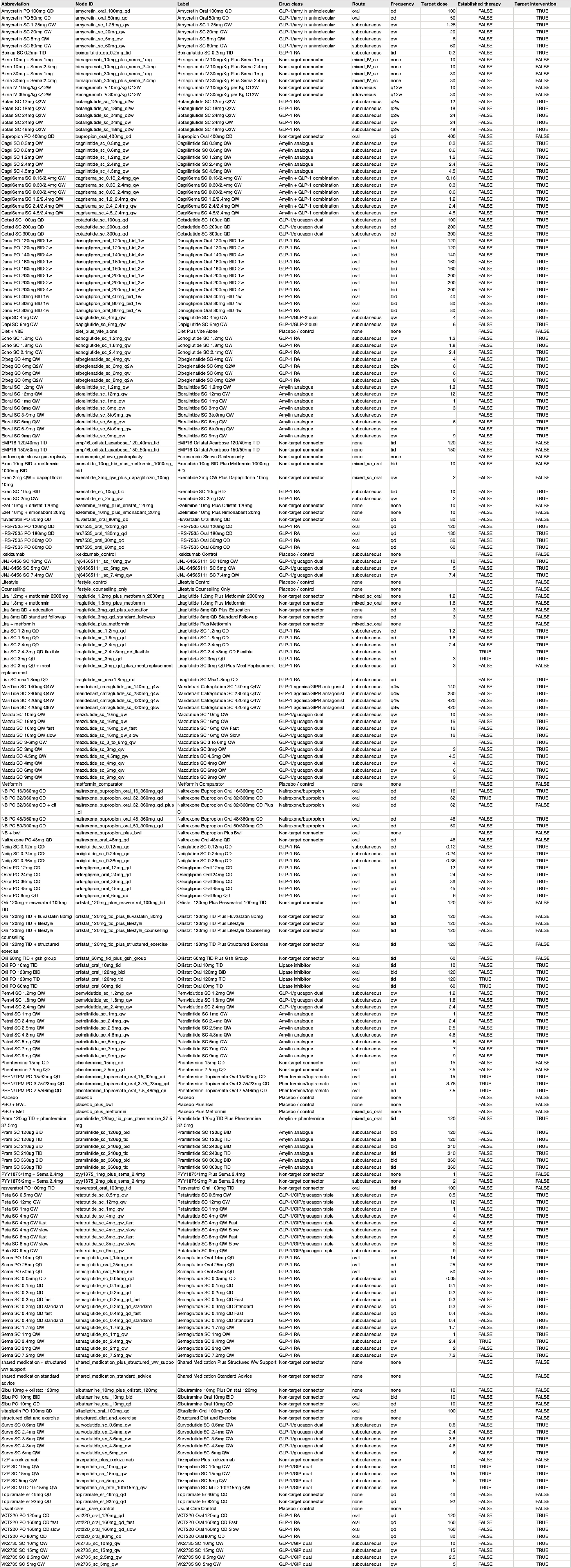
Network node dictionary and abbreviations.

| Abbreviation | Node ID | Label | Drug class | Route | Frequency | Target dose | Established therapy | Target intervention |
|---|---|---|---|---|---|---|---|---|
| Amycretin PO 100mg QD | amycretin_oral_100mg_qd | Amycretin Oral 100mg QD | GLP-1/amylin unimolecular | oral | qd | 100 | FALSE | TRUE |
| Amycretin PO 50mg QD | amycretin_oral_50mg_qd | Amycretin Oral 50mg QD | GLP-1/amylin unimolecular | oral | qd | 50 | FALSE | TRUE |
| Amycretin SC 1.25mg QW | amycretin_sc_1.25mg_qw | Amycretin SC 1.25mg QW | GLP-1/amylin unimolecular | subcutaneous | qw | 1.25 | FALSE | TRUE |
| Amycretin SC 20mg QW | amycretin_sc_20mg_qw | Amycretin SC 20mg QW | GLP-1/amylin unimolecular | subcutaneous | qw | 20 | FALSE | TRUE |
| Amycretin SC 5mg QW | amycretin_sc_5mg_qw | Amycretin SC 5mg QW | GLP-1/amylin unimolecular | subcutaneous | qw | 5 | FALSE | TRUE |
| Amycretin SC 60mg QW | amycretin_sc_60mg_qw | Amycretin SC 60mg QW | GLP-1/amylin unimolecular | subcutaneous | qw | 60 | FALSE | TRUE |
| Beinag SC 0.2mg TID | beinaglutide_sc_0.2mg_tid | Beinaglutide SC 0.2mg TID | GLP-1 RA | subcutaneous | tid | 0.2 | FALSE | TRUE |
| Bima 10mg + Sema 1mg | bimagrumab_10mg_plus_sema_1mg | Bimagrumab IV 10mg/Kg Plus Sema 1mg | Non-target connector | mixed_IV_sc | none | 10 | FALSE | FALSE |
| Bima 10mg + Sema 2.4mg | bimagrumab_10mg_plus_sema_2.4mg | Bimagrumab IV 10mg/Kg Plus Sema 2.4mg | Non-target connector | mixed_IV_sc | none | 10 | FALSE | FALSE |
| Bima 30mg + Sema 1mg | bimagrumab_30mg_plus_sema_1mg | Bimagrumab IV 30mg/Kg Plus Sema 1mg | Non-target connector | mixed_IV_sc | none | 30 | FALSE | FALSE |
| Bima 30mg + Sema 2.4mg | bimagrumab_30mg_plus_sema_2.4mg | Bimagrumab IV 30mg/Kg Plus Sema 2.4mg | Non-target connector | mixed_IV_sc | none | 30 | FALSE | FALSE |
| Bima IV 10mg/kg Q12W | Bimagrumab IV 10mg/kg Q12W | Bimagrumab IV 10mg/Kg per Kg Q12W | Non-target connector | intravenous | q12w | 10 | FALSE | FALSE |
| Bima IV 30mg/kg Q12W | Bimagrumab IV 30mg/kg Q12W | Bimagrumab IV 30mg/Kg per Kg Q12W | Non-target connector | intravenous | q12w | 30 | FALSE | FALSE |
| Bofan SC 12mg Q2W | bofanglutide_sc_12mg_q2w | Bofanglutide SC 12mg Q2W | GLP-1 RA | subcutaneous | q2w | 12 | FALSE | TRUE |
| Bofan SC 18mg Q2W | bofanglutide_sc_18mg_q2w | Bofanglutide SC 18mg Q2W | GLP-1 RA | subcutaneous | q2w | 18 | FALSE | TRUE |
| Bofan SC 24mg Q2W | bofanglutide_sc_24mg_q2w | Bofanglutide SC 24mg Q2W | GLP-1 RA | subcutaneous | q2w | 24 | FALSE | TRUE |
| Bofan SC 24mg QW | bofanglutide_sc_24mg_qw | Bofanglutide SC 24mg QW | GLP-1 RA | subcutaneous | qw | 24 | FALSE | TRUE |
| Bofan SC 48mg Q2W | bofanglutide_sc_48mg_q2w | Bofanglutide SC 48mg Q2W | GLP-1 RA | subcutaneous | q2w | 48 | FALSE | TRUE |
| Bupropion PO 400mg QD | bupropion_oral_400mg_qd | Bupropion Oral 400mg QD | Non-target connector | oral | qd | 400 | FALSE | FALSE |
| Cagri SC 0.3mg QW | cagrilintide_sc_0.3mg_qw | Cagrilintide SC 0.3mg QW | Amylin analogue | subcutaneous | qw | 0.3 | FALSE | TRUE |
| Cagri SC 0.6mg QW | cagrilintide_sc_0.6mg_qw | Cagrilintide SC 0.6mg QW | Amylin analogue | subcutaneous | qw | 0.6 | FALSE | TRUE |
| Cagri SC 1.2mg QW | cagrilintide_sc_1.2mg_qw | Cagrilintide SC 1.2mg QW | Amylin analogue | subcutaneous | qw | 1.2 | FALSE | TRUE |
| Cagri SC 2.4mg QW | cagrilintide_sc_2.4mg_qw | Cagrilintide SC 2.4mg QW | Amylin analogue | subcutaneous | qw | 2.4 | FALSE | TRUE |
| Cagri SC 4.5mg QW | cagrilintide_sc_4.5mg_qw | Cagrilintide SC 4.5mg QW | Amylin analogue | subcutaneous | qw | 4.5 | FALSE | TRUE |
| CagriSema SC 0.16/2.4mg QW | cagrisema_sc_0.16_2.4mg_qw | CagriSema SC 0.16/2.4mg QW | Amylin + GLP-1 combination | subcutaneous | qw | 0.16 | FALSE | TRUE |
| CagriSema SC 0.30/2.4mg QW | cagrisema_sc_0.30_2.4mg_qw | CagriSema SC 0.30/2.4mg QW | Amylin + GLP-1 combination | subcutaneous | qw | 0.3 | FALSE | TRUE |
| CagriSema SC 0.60/2.4mg QW | cagrisema_sc_0.60_2.4mg_qw | CagriSema SC 0.60/2.4mg QW | Amylin + GLP-1 combination | subcutaneous | qw | 0.6 | FALSE | TRUE |
| CagriSema SC 1.2/2.4mg QW | cagrisema_sc_1.2_2.4mg_qw | CagriSema SC 1.2/2.4mg QW | Amylin + GLP-1 combination | subcutaneous | qw | 1.2 | FALSE | TRUE |
| CagriSema SC 2.4/2.4mg QW | cagrisema_sc_2.4_2.4mg_qw | CagriSema SC 2.4/2.4mg QW | Amylin + GLP-1 combination | subcutaneous | qw | 2.4 | FALSE | TRUE |
| CagriSema SC 4.5/2.4mg QW | cagrisema_sc_4.5_2.4mg_qw | CagriSema SC 4.5/2.4mg QW | Amylin + GLP-1 combination | subcutaneous | qw | 4.5 | FALSE | TRUE |
| Cotad SC 100ug QD | cotadutide_sc_100ug_qd | Cotadutide SC 100ug QD | GLP-1/glucagon dual | subcutaneous | qd | 100 | FALSE | TRUE |
| Cotad SC 200ug QD | cotadutide_sc_200ug_qd | Cotadutide SC 200ug QD | GLP-1/glucagon dual | subcutaneous | qd | 200 | FALSE | TRUE |
| Cotad SC 300ug QD | cotadutide_sc_300ug_qd | Cotadutide SC 300ug QD | GLP-1/glucagon dual | subcutaneous | qd | 300 | FALSE | TRUE |
| Danu PO 120mg BID 1w | danuglipron_oral_120mg_bid_1w | Danuglipron Oral 120mg BID 1w | GLP-1 RA | oral | bid | 120 | FALSE | TRUE |
| Danu PO 120mg BID 2w | danuglipron_oral_120mg_bid_2w | Danuglipron Oral 120mg BID 2w | GLP-1 RA | oral | bid | 120 | FALSE | TRUE |
| Danu PO 140mg BID 4w | danuglipron_oral_140mg_bid_4w | Danuglipron Oral 140mg BID 4w | GLP-1 RA | oral | bid | 140 | FALSE | TRUE |
| Danu PO 160mg BID 1w | danuglipron_oral_160mg_bid_1w | Danuglipron Oral 160mg BID 1w | GLP-1 RA | oral | bid | 160 | FALSE | TRUE |
| Danu PO 160mg BID 2w | danuglipron_oral_160mg_bid_2w | Danuglipron Oral 160mg BID 2w | GLP-1 RA | oral | bid | 160 | FALSE | TRUE |
| Danu PO 200mg BID 1w | danuglipron_oral_200mg_bid_1w | Danuglipron Oral 200mg BID 1w | GLP-1 RA | oral | bid | 200 | FALSE | TRUE |
| Danu PO 200mg BID 2w | danuglipron_oral_200mg_bid_2w | Danuglipron Oral 200mg BID 2w | GLP-1 RA | oral | bid | 200 | FALSE | TRUE |
| Danu PO 200mg BID 4w | danuglipron_oral_200mg_bid_4w | Danuglipron Oral 200mg BID 4w | GLP-1 RA | oral | bid | 200 | FALSE | TRUE |
| Danu PO 40mg BID 1w | danuglipron_oral_40mg_bid_1w | Danuglipron Oral 40mg BID 1w | GLP-1 RA | oral | bid | 40 | FALSE | TRUE |
| Danu PO 80mg BID 1w | danuglipron_oral_80mg_bid_1w | Danuglipron Oral 80mg BID 1w | GLP-1 RA | oral | bid | 80 | FALSE | TRUE |
| Danu PO 80mg BID 4w | danuglipron_oral_80mg_bid_4w | Danuglipron Oral 80mg BID 4w | GLP-1 RA | oral | bid | 80 | FALSE | TRUE |
| Dapi SC 4mg QW | dapiglutide_sc_4mg_qw | Dapiglutide SC 4mg QW | GLP-1/GLP-2 dual | subcutaneous | qw | 4 | FALSE | TRUE |
| Dapi SC 6mg QW | dapiglutide_sc_6mg_qw | Dapiglutide SC 6mg QW | GLP-1/GLP-2 dual | subcutaneous | qw | 6 | FALSE | TRUE |
| Diet + VitE | diet_plus_vite_alone | Diet Plus Vite Alone | Placebo / control | none | none |  | FALSE | FALSE |
| Ecno SC 1.2mg QW | ecnoglutide_sc_1.2mg_qw | Ecnoglutide SC 1.2mg QW | GLP-1 RA | subcutaneous | qw | 1.2 | FALSE | TRUE |
| Ecno SC 1.8mg QW | ecnoglutide_sc_1.8mg_qw | Ecnoglutide SC 1.8mg QW | GLP-1 RA | subcutaneous | qw | 1.8 | FALSE | TRUE |
| Ecno SC 2.4mg QW | ecnoglutide_sc_2.4mg_qw | Ecnoglutide SC 2.4mg QW | GLP-1 RA | subcutaneous | qw | 2.4 | FALSE | TRUE |
| Efpeg SC 4mg QW | efpeglenatide_sc_4mg_qw | Efpeglenatide SC 4mg QW | GLP-1 RA | subcutaneous | qw | 4 | FALSE | TRUE |
| Efpeg SC 6mg Q2W | efpeglenatide_sc_6mg_q2w | Efpeglenatide SC 6mg Q2W | GLP-1 RA | subcutaneous | q2w | 6 | FALSE | TRUE |
| Efpeg SC 6mg QW | efpeglenatide_sc_6mg_qw | Efpeglenatide SC 6mg QW | GLP-1 RA | subcutaneous | qw | 6 | FALSE | TRUE |
| Efpeg SC 8mg Q2W | efpeglenatide_sc_8mg_q2w | Efpeglenatide SC 8mg Q2W | GLP-1 RA | subcutaneous | q2w | 8 | FALSE | TRUE |
| Eloral SC 1.2mg QW | eloralintide_sc_1.2mg_qw | Eloralintide SC 1.2mg QW | Amylin analogue | subcutaneous | qw | 1.2 | FALSE | TRUE |
| Eloral SC 12mg QW | eloralintide_sc_12mg_qw | Eloralintide SC 12mg QW | Amylin analogue | subcutaneous | qw | 12 | FALSE | TRUE |
| Eloral SC 1mg QW | eloralintide_sc_1mg_qw | Eloralintide SC 1mg QW | Amylin analogue | subcutaneous | qw | 1 | FALSE | TRUE |
| Eloral SC 3mg QW | eloralintide_sc_3mg_qw | Eloralintide SC 3mg QW | Amylin analogue | subcutaneous | qw | 3 | FALSE | TRUE |
| Eloral SC 3-9mg QW | eloralintide_sc_3to9mg_qw | Eloralintide SC 3to9mg QW | Amylin analogue | subcutaneous | qw |  | FALSE | TRUE |
| Eloral SC 6mg QW | eloralintide_sc_6mg_qw | Eloralintide SC 6mg QW | Amylin analogue | subcutaneous | qw | 6 | FALSE | TRUE |
| Eloral SC 6-9mg QW | eloralintide_sc_6to9mg_qw | Eloralintide SC 6to9mg QW | Amylin analogue | subcutaneous | qw |  | FALSE | TRUE |
| Eloral SC 9mg QW | eloralintide_sc_9mg_qw | Eloralintide SC 9mg QW | Amylin analogue | subcutaneous | qw | 9 | FALSE | TRUE |
| EMP16 120/40mg TID | emp16_orlistat_acarbose_120_40mg_tid | EMP16 Orlistat Acarbose 120/40mg TID | Non-target connector | none | tid | 120 | FALSE | FALSE |
| EMP16 150/50mg TID | emp16_orlistat_acarbose_150_50mg_tid | EMP16 Orlistat Acarbose 150/50mg TID | Non-target connector | none | tid | 150 | FALSE | FALSE |
| endoscopic sleeve gastroplasty | endoscopic_sleeve_gastroplasty | Endoscopic Sleeve Gastroplasty | Non-target connector | none | none |  | FALSE | FALSE |
| Exen 10ug BID + metformin 1000mg BID | exenatide_10ug_bid_plus_metformin_1000mg_bid | Exenatide 10ug BID Plus Metformin 1000mg BID | Non-target connector | mixed_sc_oral | bid | 10 | FALSE | FALSE |
| Exen 2mg QW + dapagliflozin 10mg | exenatide_2mg_qw_plus_dapagliflozin_10mg | Exenatide 2mg QW Plus Dapagliflozin 10mg | Non-target connector | mixed_sc_oral | qw | 2 | FALSE | FALSE |
| Exen SC 10ug BID | exenatide_sc_10ug_bid | Exenatide SC 10ug BID | GLP-1 RA | subcutaneous | bid | 10 | FALSE | TRUE |
| Exen SC 2mg QW | exenatide_sc_2mg_qw | Exenatide SC 2mg QW | GLP-1 RA | subcutaneous | qw | 2 | FALSE | TRUE |
| Ezet 10mg + orlistat 120mg | ezetimibe_10mg_plus_orlistat_120mg | Ezetimibe 10mg Plus Orlistat 120mg | Non-target connector | none | none | 10 | FALSE | FALSE |
| Ezet 10mg + rimonabant 20mg | ezetimibe_10mg_plus_rimonabant_20mg | Ezetimibe 10mg Plus Rimonabant 20mg | Non-target connector | none | none | 10 | FALSE | FALSE |
| fluvastatin PO 80mg QD | fluvastatin_oral_80mg_qd | Fluvastatin Oral 80mg QD | Non-target connector | oral | qd | 80 | FALSE | FALSE |
| HRS-7535 PO 120mg QD | hrs7535_oral_120mg_qd | HRS-7535 Oral 120mg QD | GLP-1 RA | oral | qd | 120 | FALSE | TRUE |
| HRS-7535 PO 180mg QD | hrs7535_oral_180mg_qd | HRS-7535 Oral 180mg QD | GLP-1 RA | oral | qd | 180 | FALSE | TRUE |
| HRS-7535 PO 30mg QD | hrs7535_oral_30mg_qd | HRS-7535 Oral 30mg QD | GLP-1 RA | oral | qd | 30 | FALSE | TRUE |
| HRS-7535 PO 60mg QD | hrs7535_oral_60mg_qd | HRS-7535 Oral 60mg QD | GLP-1 RA | oral | qd | 60 | FALSE | TRUE |
| Ixekizumab | ixekizumab_control | Ixekizumab Control | Placebo / control | subcutaneous | none |  | FALSE | FALSE |
| JNJ-6456 SC 10mg QW | jnj64565111_sc_10mg_qw | JNJ-64565111 SC 10mg QW | GLP-1/glucagon dual | subcutaneous | qw | 10 | FALSE | TRUE |
| JNJ-6456 SC 5mg QW | jnj64565111_sc_5mg_qw | JNJ-64565111 SC 5mg QW | GLP-1/glucagon dual | subcutaneous | qw | 5 | FALSE | TRUE |
| JNJ-6456 SC 7.4mg QW | jnj64565111_sc_7.4mg_qw | JNJ-64565111 SC 7.4mg QW | GLP-1/glucagon dual | subcutaneous | qw | 7.4 | FALSE | TRUE |
| Lifestyle | lifestyle_control | Lifestyle Control | Placebo / control | none | none |  | FALSE | FALSE |
| Counselling | lifestyle_counselling_only | Lifestyle Counselling Only | Placebo / control | none | none |  | FALSE | FALSE |
| Lira 1.2mg + metformin 2000mg | liraglutide_1.2mg_plus_metformin_2000mg | Liraglutide 1.2mg Plus Metformin 2000mg | Non-target connector | mixed_sc_oral | none | 1.2 | FALSE | FALSE |
| Lira 1.8mg + metformin | liraglutide_1.8mg_plus_metformin | Liraglutide 1.8mg Plus Metformin | Non-target connector | mixed_sc_oral | none | 1.8 | FALSE | FALSE |
| Lira 3mg QD + education | liraglutide_3mg_qd_plus_education | Liraglutide 3mg QD Plus Education | Non-target connector | none | qd | 3 | FALSE | FALSE |
| Lira 3mg QD standard followup | liraglutide_3mg_qd_standard_followup | Liraglutide 3mg QD Standard Followup | Non-target connector | none | qd | 3 | FALSE | FALSE |
| Lira + metformin | liraglutide_plus_metformin | Liraglutide Plus Metformin | Non-target connector | mixed_sc_oral | none |  | FALSE | FALSE |
| Lira SC 1.2mg QD | liraglutide_sc_1.2mg_qd | Liraglutide SC 1.2mg QD | GLP-1 RA | subcutaneous | qd | 1.2 | FALSE | TRUE |
| Lira SC 1.8mg QD | liraglutide_sc_1.8mg_qd | Liraglutide SC 1.8mg QD | GLP-1 RA | subcutaneous | qd | 1.8 | FALSE | TRUE |
| Lira SC 2.4mg QD | liraglutide_sc_2.4mg_qd | Liraglutide SC 2.4mg QD | GLP-1 RA | subcutaneous | qd | 2.4 | FALSE | TRUE |
| Lira SC 2.4-3mg QD flexible | liraglutide_sc_2.4to3mg_qd_flexible | Liraglutide SC 2.4to3mg QD Flexible | GLP-1 RA | subcutaneous | qd |  | TRUE | TRUE |
| Lira SC 3mg QD | liraglutide_sc_3mg_qd | Liraglutide SC 3mg QD | GLP-1 RA | subcutaneous | qd | 3 | TRUE | TRUE |
| Lira SC 3mg QD + meal replacement | liraglutide_sc_3mg_qd_plus_meal_replacement | Liraglutide SC 3mg QD Plus Meal Replacement | GLP-1 RA | subcutaneous | qd | 3 | FALSE | FALSE |
| Lira SC max1.8mg QD | liraglutide_sc_max1.8mg_qd | Liraglutide SC Max1.8mg QD | GLP-1 RA | subcutaneous | qd |  | FALSE | TRUE |
| MariTide SC 140mg Q4W | maridebart_cafraglutide_sc_140mg_q4w | Maridebart Cafraglutide SC 140mg Q4W | GLP-1 agonist/GIPR antagonist | subcutaneous | q4w | 140 | FALSE | TRUE |
| MariTide SC 280mg Q4W | maridebart_cafraglutide_sc_280mg_q4w | Maridebart Cafraglutide SC 280mg Q4W | GLP-1 agonist/GIPR antagonist | subcutaneous | q4w | 280 | FALSE | TRUE |
| MariTide SC 420mg Q4W | maridebart_cafraglutide_sc_420mg_q4w | Maridebart Cafraglutide SC 420mg Q4W | GLP-1 agonist/GIPR antagonist | subcutaneous | q4w | 420 | FALSE | TRUE |
| MariTide SC 420mg Q8W | maridebart_cafraglutide_sc_420mg_q8w | Maridebart Cafraglutide SC 420mg Q8W | GLP-1 agonist/GIPR antagonist | subcutaneous | q8w | 420 | FALSE | TRUE |
| Mazdu SC 10mg QW | mazdutide_sc_10mg_qw | Mazdutide SC 10mg QW | GLP-1/glucagon dual | subcutaneous | qw | 10 | FALSE | TRUE |
| Mazdu SC 16mg QW | mazdutide_sc_16mg_qw | Mazdutide SC 16mg QW | GLP-1/glucagon dual | subcutaneous | qw | 16 | FALSE | TRUE |
| Mazdu SC 16mg QW fast | mazdutide_sc_16mg_qw_fast | Mazdutide SC 16mg QW Fast | GLP-1/glucagon dual | subcutaneous | qw | 16 | FALSE | TRUE |
| Mazdu SC 16mg QW slow | mazdutide_sc_16mg_qw_slow | Mazdutide SC 16mg QW Slow | GLP-1/glucagon dual | subcutaneous | qw | 16 | FALSE | TRUE |
| Mazdu SC 3-6mg QW | mazdutide_sc_3_to_6mg_qw | Mazdutide SC 3 to 6mg QW | GLP-1/glucagon dual | subcutaneous | qw |  | FALSE | TRUE |
| Mazdu SC 3mg QW | mazdutide_sc_3mg_qw | Mazdutide SC 3mg QW | GLP-1/glucagon dual | subcutaneous | qw | 3 | FALSE | TRUE |
| Mazdu SC 4.5mg QW | mazdutide_sc_4.5mg_qw | Mazdutide SC 4.5mg QW | GLP-1/glucagon dual | subcutaneous | qw | 4.5 | FALSE | TRUE |
| Mazdu SC 4mg QW | mazdutide_sc_4mg_qw | Mazdutide SC 4mg QW | GLP-1/glucagon dual | subcutaneous | qw | 4 | FALSE | TRUE |
| Mazdu SC 6mg QW | mazdutide_sc_6mg_qw | Mazdutide SC 6mg QW | GLP-1/glucagon dual | subcutaneous | qw | 6 | FALSE | TRUE |
| Mazdu SC 9mg QW | mazdutide_sc_9mg_qw | Mazdutide SC 9mg QW | GLP-1/glucagon dual | subcutaneous | qw | 9 | FALSE | TRUE |
| Metformin | metformin_comparator | Metformin Comparator | Placebo / control | none | none |  | FALSE | FALSE |
| NB PO 16/360mg QD | naltrexone_bupropion_oral_16_360mg_qd | Naltrexone Bupropion Oral 16/360mg QD | Naltrexone/bupropion | oral | qd | 16 | FALSE | TRUE |
| NB PO 32/360mg QD | naltrexone_bupropion_oral_32_360mg_qd | Naltrexone Bupropion Oral 32/360mg QD | Naltrexone/bupropion | oral | qd | 32 | TRUE | TRUE |
| NB PO 32/360mg QD + cli | naltrexone_bupropion_oral_32_360mg_qd_plus_cli | Naltrexone Bupropion Oral 32/360mg QD Plus Cli | Naltrexone/bupropion | oral | qd | 32 | FALSE | FALSE |
| NB PO 48/360mg QD | naltrexone_bupropion_oral_48_360mg_qd | Naltrexone Bupropion Oral 48/360mg QD | Naltrexone/bupropion | oral | qd | 48 | FALSE | TRUE |
| NB PO 50/300mg QD | naltrexone_bupropion_oral_50_300mg_qd | Naltrexone Bupropion Oral 50/300mg QD | Naltrexone/bupropion | oral | qd | 50 | FALSE | TRUE |
| NB + bwl | naltrexone_bupropion_plus_bwl | Naltrexone Bupropion Plus Bwl | Non-target connector | none | none |  | FALSE | FALSE |
| Naltrexone PO 48mg QD | naltrexone_oral_48mg_qd | Naltrexone Oral 48mg QD | Non-target connector | oral | qd | 48 | FALSE | FALSE |
| Noiig SC 0.12mg QD | noiiglutide_sc_0.12mg_qd | Noiiglutide SC 0.12mg QD | GLP-1 RA | subcutaneous | qd | 0.12 | FALSE | TRUE |
| Noiig SC 0.24mg QD | noiiglutide_sc_0.24mg_qd | Noiiglutide SC 0.24mg QD | GLP-1 RA | subcutaneous | qd | 0.24 | FALSE | TRUE |
| Noiig SC 0.36mg QD | noiiglutide_sc_0.36mg_qd | Noiiglutide SC 0.36mg QD | GLP-1 RA | subcutaneous | qd | 0.36 | FALSE | TRUE |
| Orfor PO 12mg QD | orforglipron_oral_12mg_qd | Orforglipron Oral 12mg QD | GLP-1 RA | oral | qd | 12 | FALSE | TRUE |
| Orfor PO 24mg QD | orforglipron_oral_24mg_qd | Orforglipron Oral 24mg QD | GLP-1 RA | oral | qd | 24 | FALSE | TRUE |
| Orfor PO 36mg QD | orforglipron_oral_36mg_qd | Orforglipron Oral 36mg QD | GLP-1 RA | oral | qd | 36 | FALSE | TRUE |
| Orfor PO 45mg QD | orforglipron_oral_45mg_qd | Orforglipron Oral 45mg QD | GLP-1 RA | oral | qd | 45 | FALSE | TRUE |
| Orfor PO 6mg QD | orforglipron_oral_6mg_qd | Orforglipron Oral 6mg QD | GLP-1 RA | oral | qd | 6 | FALSE | TRUE |
| Orli 120mg TID + resveratrol 100mg TID | orlistat_120mg_tid_plus_resveratrol_100mg_tid | Orlistat 120mg TID Plus Resveratrol 100mg TID | Non-target connector | oral | tid | 120 | FALSE | FALSE |
| Orli 120mg TID + fluvastatin 80mg | orlistat_120mg_tid_plus_fluvastatin_80mg | Orlistat 120mg TID Plus Fluvastatin 80mg | Non-target connector | oral | tid | 120 | FALSE | FALSE |
| Orli 120mg TID + lifestyle | orlistat_120mg_tid_plus_lifestyle | Orlistat 120mg TID Plus Lifestyle | Non-target connector | oral | tid | 120 | FALSE | FALSE |
| Orli 120mg TID + lifestyle counselling | orlistat_120mg_tid_plus_lifestyle_counselling | Orlistat 120mg TID Plus Lifestyle Counselling | Non-target connector | oral | tid | 120 | FALSE | FALSE |
| Orli 120mg TID + structured exercise | orlistat_120mg_tid_plus_structured_exercise | Orlistat 120mg TID Plus Structured Exercise | Non-target connector | oral | tid | 120 | FALSE | FALSE |
| Orli 60mg TID + gsh group | orlistat_60mg_tid_plus_gsh_group | Orlistat 60mg TID Plus Gsh Group | Non-target connector | oral | tid | 60 | FALSE | FALSE |
| Orli PO 10mg TID | orlistat_oral_10mg_tid | Orlistat Oral 10mg TID | Lipase inhibitor | oral | tid | 10 | FALSE | TRUE |
| Orli PO 120mg BID | orlistat_oral_120mg_bid | Orlistat Oral 120mg BID | Lipase inhibitor | oral | bid | 120 | FALSE | TRUE |
| Orli PO 120mg TID | orlistat_oral_120mg_tid | Orlistat Oral 120mg TID | Lipase inhibitor | oral | tid | 120 | TRUE | TRUE |
| Orli PO 60mg TID | orlistat_oral_60mg_tid | Orlistat Oral 60mg TID | Lipase inhibitor | oral | tid | 60 | TRUE | TRUE |
| Pemvi SC 1.2mg QW | pemvidutide_sc_1.2mg_qw | Pemvidutide SC 1.2mg QW | GLP-1/glucagon dual | subcutaneous | qw | 1.2 | FALSE | TRUE |
| Pemvi SC 1.8mg QW | pemvidutide_sc_1.8mg_qw | Pemvidutide SC 1.8mg QW | GLP-1/glucagon dual | subcutaneous | qw | 1.8 | FALSE | TRUE |
| Pemvi SC 2.4mg QW | pemvidutide_sc_2.4mg_qw | Pemvidutide SC 2.4mg QW | GLP-1/glucagon dual | subcutaneous | qw | 2.4 | FALSE | TRUE |
| Petrel SC 1mg QW | petrelintide_sc_1mg_qw | Petrelintide SC 1mg QW | Amylin analogue | subcutaneous | qw | 1 | FALSE | TRUE |
| Petrel SC 2.4mg QW | petrelintide_sc_2.4mg_qw | Petrelintide SC 2.4mg QW | Amylin analogue | subcutaneous | qw | 2.4 | FALSE | TRUE |
| Petrel SC 2.5mg QW | petrelintide_sc_2.5mg_qw | Petrelintide SC 2.5mg QW | Amylin analogue | subcutaneous | qw | 2.5 | FALSE | TRUE |
| Petrel SC 4.8mg QW | petrelintide_sc_4.8mg_qw | Petrelintide SC 4.8mg QW | Amylin analogue | subcutaneous | qw | 4.8 | FALSE | TRUE |
| Petrel SC 5mg QW | petrelintide_sc_5mg_qw | Petrelintide SC 5mg QW | Amylin analogue | subcutaneous | qw | 5 | FALSE | TRUE |
| Petrel SC 7mg QW | petrelintide_sc_7mg_qw | Petrelintide SC 7mg QW | Amylin analogue | subcutaneous | qw | 7 | FALSE | TRUE |
| Petrel SC 9mg QW | petrelintide_sc_9mg_qw | Petrelintide SC 9mg QW | Amylin analogue | subcutaneous | qw | 9 | FALSE | TRUE |
| Phentermine 15mg QD | phentermine_15mg_qd | Phentermine 15mg QD | Non-target connector | oral | qd | 15 | FALSE | FALSE |
| Phentermine 7.5mg QD | phentermine_7.5mg_qd | Phentermine 7.5mg QD | Non-target connector | oral | qd | 7.5 | FALSE | FALSE |
| PHEN/TPM PO 15/92mg QD | phentermine_topiramate_oral_15_92mg_qd | Phentermine Topiramate Oral 15/92mg QD | Phentermine/topiramate | oral | qd | 15 | TRUE | TRUE |
| PHEN/TPM PO 3.75/23mg QD | phentermine_topiramate_oral_3.75_23mg_qd | Phentermine Topiramate Oral 3.75/23mg QD | Phentermine/topiramate | oral | qd | 3.75 | FALSE | TRUE |
| PHEN/TPM PO 7.5/46mg QD | phentermine_topiramate_oral_7.5_46mg_qd | Phentermine Topiramate Oral 7.5/46mg QD | Phentermine/topiramate | oral | qd | 7.5 | TRUE | TRUE |
| Placebo | placebo | Placebo | Placebo / control | none | none |  | FALSE | FALSE |
| PBO + BWL | placebo_plus_bwl | Placebo Plus Bwl | Placebo / control | none | none |  | FALSE | FALSE |
| PBO + Met | placebo_plus_metformin | Placebo Plus Metformin | Placebo / control | mixed_sc_oral | none |  | FALSE | FALSE |
| Pram 120ug TID + phentermine 37.5mg | pramlintide_120ug_tid_plus_phentermine_37.5mg | Pramlintide 120ug TID Plus Phentermine 37.5mg | Amylin + phentermine | mixed_sc_oral | tid | 120 | FALSE | FALSE |
| Pram SC 120ug BID | pramlintide_sc_120ug_bid | Pramlintide SC 120ug BID | Amylin analogue | subcutaneous | bid | 120 | FALSE | TRUE |
| Pram SC 120ug TID | pramlintide_sc_120ug_tid | Pramlintide SC 120ug TID | Amylin analogue | subcutaneous | tid | 120 | FALSE | TRUE |
| Pram SC 240ug BID | pramlintide_sc_240ug_bid | Pramlintide SC 240ug BID | Amylin analogue | subcutaneous | bid | 240 | FALSE | TRUE |
| Pram SC 240ug TID | pramlintide_sc_240ug_tid | Pramlintide SC 240ug TID | Amylin analogue | subcutaneous | tid | 240 | FALSE | TRUE |
| Pram SC 360ug BID | pramlintide_sc_360ug_bid | Pramlintide SC 360ug BID | Amylin analogue | subcutaneous | bid | 360 | FALSE | TRUE |
| Pram SC 360ug TID | pramlintide_sc_360ug_tid | Pramlintide SC 360ug TID | Amylin analogue | subcutaneous | tid | 360 | FALSE | TRUE |
| PYY1875/1mg + Sema 2.4mg | pyy1875_1mg_plus_sema_2.4mg | PYY1875/1mg Plus Sema 2.4mg | Non-target connector | subcutaneous | none | 1 | FALSE | FALSE |
| PYY1875/2mg + Sema 2.4mg | pyy1875_2mg_plus_sema_2.4mg | PYY1875/2mg Plus Sema 2.4mg | Non-target connector | subcutaneous | none | 2 | FALSE | FALSE |
| resveratrol PO 100mg TID | resveratrol_oral_100mg_tid | Resveratrol Oral 100mg TID | Non-target connector | oral | tid | 100 | FALSE | FALSE |
| Reta SC 0.5mg QW | retatrutide_sc_0.5mg_qw | Retatrutide SC 0.5mg QW | GLP-1/GIP/glucagon triple | subcutaneous | qw | 0.5 | FALSE | TRUE |
| Reta SC 12mg QW | retatrutide_sc_12mg_qw | Retatrutide SC 12mg QW | GLP-1/GIP/glucagon triple | subcutaneous | qw | 12 | FALSE | TRUE |
| Reta SC 1mg QW | retatrutide_sc_1mg_qw | Retatrutide SC 1mg QW | GLP-1/GIP/glucagon triple | subcutaneous | qw | 1 | FALSE | TRUE |
| Reta SC 4mg QW | retatrutide_sc_4mg_qw | Retatrutide SC 4mg QW | GLP-1/GIP/glucagon triple | subcutaneous | qw | 4 | FALSE | TRUE |
| Reta SC 4mg QW fast | retatrutide_sc_4mg_qw_fast | Retatrutide SC 4mg QW Fast | GLP-1/GIP/glucagon triple | subcutaneous | qw | 4 | FALSE | TRUE |
| Reta SC 4mg QW slow | retatrutide_sc_4mg_qw_slow | Retatrutide SC 4mg QW Slow | GLP-1/GIP/glucagon triple | subcutaneous | qw | 4 | FALSE | TRUE |
| Reta SC 8mg QW fast | retatrutide_sc_8mg_qw_fast | Retatrutide SC 8mg QW Fast | GLP-1/GIP/glucagon triple | subcutaneous | qw | 8 | FALSE | TRUE |
| Reta SC 8mg QW slow | retatrutide_sc_8mg_qw_slow | Retatrutide SC 8mg QW Slow | GLP-1/GIP/glucagon triple | subcutaneous | qw | 8 | FALSE | TRUE |
| Reta SC 9mg QW | retatrutide_sc_9mg_qw | Retatrutide SC 9mg QW | GLP-1/GIP/glucagon triple | subcutaneous | qw | 9 | FALSE | TRUE |
| Sema PO 14mg QD | semaglutide_oral_14mg_qd | Semaglutide Oral 14mg QD | GLP-1 RA | oral | qd | 14 | FALSE | TRUE |
| Sema PO 25mg QD | semaglutide_oral_25mg_qd | Semaglutide Oral 25mg QD | GLP-1 RA | oral | qd | 25 | FALSE | TRUE |
| Sema PO 50mg QD | semaglutide_oral_50mg_qd | Semaglutide Oral 50mg QD | GLP-1 RA | oral | qd | 50 | FALSE | TRUE |
| Sema SC 0.05mg QD | semaglutide_sc_0.05mg_qd | Semaglutide SC 0.05mg QD | GLP-1 RA | subcutaneous | qd | 0.05 | FALSE | TRUE |
| Sema SC 0.1mg QD | semaglutide_sc_0.1mg_qd | Semaglutide SC 0.1mg QD | GLP-1 RA | subcutaneous | qd | 0.1 | FALSE | TRUE |
| Sema SC 0.2mg QD | semaglutide_sc_0.2mg_qd | Semaglutide SC 0.2mg QD | GLP-1 RA | subcutaneous | qd | 0.2 | FALSE | TRUE |
| Sema SC 0.3mg QD fast | semaglutide_sc_0.3mg_qd_fast | Semaglutide SC 0.3mg QD Fast | GLP-1 RA | subcutaneous | qd | 0.3 | FALSE | TRUE |
| Sema SC 0.3mg QD standard | semaglutide_sc_0.3mg_qd_standard | Semaglutide SC 0.3mg QD Standard | GLP-1 RA | subcutaneous | qd | 0.3 | FALSE | TRUE |
| Sema SC 0.4mg QD fast | semaglutide_sc_0.4mg_qd_fast | Semaglutide SC 0.4mg QD Fast | GLP-1 RA | subcutaneous | qd | 0.4 | FALSE | TRUE |
| Sema SC 0.4mg QD standard | semaglutide_sc_0.4mg_qd_standard | Semaglutide SC 0.4mg QD Standard | GLP-1 RA | subcutaneous | qd | 0.4 | FALSE | TRUE |
| Sema SC 1.7mg QW | semaglutide_sc_1.7mg_qw | Semaglutide SC 1.7mg QW | GLP-1 RA | subcutaneous | qw | 1.7 | FALSE | TRUE |
| Sema SC 1mg QW | semaglutide_sc_1mg_qw | Semaglutide SC 1mg QW | GLP-1 RA | subcutaneous | qw | 1 | FALSE | TRUE |
| Sema SC 2.4mg QW | semaglutide_sc_2.4mg_qw | Semaglutide SC 2.4mg QW | GLP-1 RA | subcutaneous | qw | 2.4 | TRUE | TRUE |
| Sema SC 2mg QW | semaglutide_sc_2mg_qw | Semaglutide SC 2mg QW | GLP-1 RA | subcutaneous | qw | 2 | FALSE | TRUE |
| Sema SC 7.2mg QW | semaglutide_sc_7.2mg_qw | Semaglutide SC 7.2mg QW | GLP-1 RA | subcutaneous | qw | 7.2 | FALSE | TRUE |
| shared medication + structured ww support | shared_medication_plus_structured_ww_support | Shared Medication Plus Structured Ww Support | Non-target connector | none | none |  | FALSE | FALSE |
| shared medication standard advice | shared_medication_standard_advice | Shared Medication Standard Advice | Non-target connector | none | none |  | FALSE | FALSE |
| Sibu 10mg + orlistat 120mg | sibutramine_10mg_plus_orlistat_120mg | Sibutramine 10mg Plus Orlistat 120mg | Non-target connector | none | none | 10 | FALSE | FALSE |
| Sibu PO 10mg BID | sibutramine_oral_10mg_bid | Sibutramine Oral 10mg BID | Non-target connector | oral | bid | 10 | FALSE | FALSE |
| Sibu PO 10mg QD | sibutramine_oral_10mg_qd | Sibutramine Oral 10mg QD | Non-target connector | oral | qd | 10 | FALSE | FALSE |
| sitagliptin PO 100mg QD | sitagliptin_oral_100mg_qd | Sitagliptin Oral 100mg QD | Non-target connector | oral | qd | 100 | FALSE | FALSE |
| structured diet and exercise | structured_diet_and_exercise | Structured Diet and Exercise | Non-target connector | none | none |  | FALSE | FALSE |
| Survo SC 0.6mg QW | survodutide_sc_0.6mg_qw | Survodutide SC 0.6mg QW | GLP-1/glucagon dual | subcutaneous | qw | 0.6 | FALSE | TRUE |
| Survo SC 2.4mg QW | survodutide_sc_2.4mg_qw | Survodutide SC 2.4mg QW | GLP-1/glucagon dual | subcutaneous | qw | 2.4 | FALSE | TRUE |
| Survo SC 3.6mg QW | survodutide_sc_3.6mg_qw | Survodutide SC 3.6mg QW | GLP-1/glucagon dual | subcutaneous | qw | 3.6 | FALSE | TRUE |
| Survo SC 4.8mg QW | survodutide_sc_4.8mg_qw | Survodutide SC 4.8mg QW | GLP-1/glucagon dual | subcutaneous | qw | 4.8 | FALSE | TRUE |
| Survo SC 6mg QW | survodutide_sc_6mg_qw | Survodutide SC 6mg QW | GLP-1/glucagon dual | subcutaneous | qw | 6 | FALSE | TRUE |
| TZP + ixekizumab | tirzepatide_plus_ixekizumab | Tirzepatide Plus Ixekizumab | Non-target connector | subcutaneous | none |  | FALSE | FALSE |
| TZP SC 10mg QW | tirzepatide_sc_10mg_qw | Tirzepatide SC 10mg QW | GLP-1/GIP dual | subcutaneous | qw | 10 | TRUE | TRUE |
| TZP SC 15mg QW | tirzepatide_sc_15mg_qw | Tirzepatide SC 15mg QW | GLP-1/GIP dual | subcutaneous | qw | 15 | TRUE | TRUE |
| TZP SC 5mg QW | tirzepatide_sc_5mg_qw | Tirzepatide SC 5mg QW | GLP-1/GIP dual | subcutaneous | qw | 5 | TRUE | TRUE |
| TZP SC MTD 10-15mg QW | tirzepatide_sc_mtd_10to15mg_qw | Tirzepatide SC MTD 10to15mg QW | GLP-1/GIP dual | subcutaneous | qw |  | TRUE | TRUE |
| Topiramate er 46mg QD | topiramate_er_46mg_qd | Topiramate Er 46mg QD | Non-target connector | none | qd | 46 | FALSE | FALSE |
| Topiramate er 92mg QD | topiramate_er_92mg_qd | Topiramate Er 92mg QD | Non-target connector | none | qd | 92 | FALSE | FALSE |
| Usual care | usual_care_control | Usual Care Control | Placebo / control | none | none |  | FALSE | FALSE |
| VCT220 PO 120mg QD | vct220_oral_120mg_qd | VCT220 Oral 120mg QD | GLP-1 RA | oral | qd | 120 | FALSE | TRUE |
| VCT220 PO 160mg QD fast | vct220_oral_160mg_qd_fast | VCT220 Oral 160mg QD Fast | GLP-1 RA | oral | qd | 160 | FALSE | TRUE |
| VCT220 PO 160mg QD slow | vct220_oral_160mg_qd_slow | VCT220 Oral 160mg QD Slow | GLP-1 RA | oral | qd | 160 | FALSE | TRUE |
| VCT220 PO 80mg QD | vct220_oral_80mg_qd | VCT220 Oral 80mg QD | GLP-1 RA | oral | qd | 80 | FALSE | TRUE |
| VK2735 SC 10mg QW | vk2735_sc_10mg_qw | VK2735 SC 10mg QW | GLP-1/GIP dual | subcutaneous | qw | 10 | FALSE | TRUE |
| VK2735 SC 15mg QW | vk2735_sc_15mg_qw | VK2735 SC 15mg QW | GLP-1/GIP dual | subcutaneous | qw | 15 | FALSE | TRUE |
| VK2735 SC 2.5mg QW | vk2735_sc_2.5mg_qw | VK2735 SC 2.5mg QW | GLP-1/GIP dual | subcutaneous | qw | 2.5 | FALSE | TRUE |
| VK2735 SC 5mg QW | vk2735_sc_5mg_qw | VK2735 SC 5mg QW | GLP-1/GIP dual | subcutaneous | qw | 5 | FALSE | TRUE |

**Table S2.** Summary of included evidence.

| Metric | Count | Description |
|---|---|---|
| records_extracted_total | 2551 | Total outcome records extracted across continuous, binary response and safety tables |
| reports_unique | 180 | Unique study reports included in the linked database (report_id) |
| parent_trials_unique | 178 | Unique parent clinical trials (parent_trial_id <= reports_unique; extension/secondary reports linked to parent trial) |
| randomization_units_unique | 196 | Independent randomization units (randomization_id > reports_unique because multi-cohort trials such as SURMOUNT-OSA [r032], MariTide [r073] and Guy-Grand [r149] contain multiple independent randomized strata) |
| participants_randomized_non_overlapping | 81014 | Total non-overlapping randomized participants (excluding long-term extension cohorts) |
| target_medication_nodes | 160 | Target obesity pharmacotherapy nodes (molecule-route-dose-titration strategies) |
| all_network_nodes | 214 | Total valid network nodes (including placebo, active controls and connectors) |

**Table S3.** Network geometry for every protocol outcome, time window and population.

| Outcome | Window | Population | Interventions | Trials | Direct comparisons | Multi-arm trials | Sub-networks | Density | Closed loops |
|---|---|---|---|---|---|---|---|---|---|
| % body-weight change | 48-56 wk | without T2D | 46 | 28 | 115 | 14 | 1 | 0.11111 | 70 |
| % body-weight change | 48-56 wk | with T2D | 15 | 9 | 24 | 6 | 1 | 0.22857 | 10 |
| % body-weight change | 20-28 wk | without T2D | 24 | 10 | 49 | 4 | 2 | 0.17754 | 27 |
| % body-weight change | 10-14 wk | without T2D | 8 | 5 | 7 | 1 | 2 | 0.25 | 1 |
| Body-weight change (kg) | 48-56 wk | without T2D | 3 | 3 | 3 | 1 | 1 | 1 | 1 |
| Body-weight change (kg) | 48-56 wk | with T2D | 3 | 2 | 3 | 1 | 1 | 1 | 1 |
| Body-weight change (kg) | 20-28 wk | without T2D | 8 | 5 | 11 | 2 | 1 | 0.39286 | 4 |
| Body-weight change (kg) | 10-14 wk | without T2D | 6 | 4 | 8 | 1 | 1 | 0.53333 | 3 |
| Waist circumference change (cm) | 48-56 wk | without T2D | 4 | 3 | 3 | 0 | 1 | 0.5 | 0 |
| Waist circumference change (cm) | 48-56 wk | with T2D | 3 | 2 | 3 | 1 | 1 | 1 | 1 |
| Waist circumference change (cm) | 20-28 wk | without T2D | 3 | 2 | 2 | 0 | 1 | 0.66667 | 0 |
| >=5% weight reduction | 48-56 wk | without T2D | 19 | 6 | 53 | 4 | 1 | 0.30994 | 35 |
| >=5% weight reduction | 48-56 wk | with T2D | 5 | 2 | 6 | 2 | 1 | 0.6 | 2 |
| >=5% weight reduction | 48-56 wk | mixed population | 6 | 3 | 8 | 1 | 1 | 0.53333 | 3 |
| >=5% weight reduction | 20-28 wk | without T2D | 11 | 5 | 19 | 2 | 1 | 0.34545 | 9 |
| >=5% weight reduction | 10-14 wk | without T2D | 3 | 1 | 3 | 1 | 1 | 1 | 1 |
| >=10% weight reduction | 48-56 wk | without T2D | 19 | 7 | 53 | 4 | 1 | 0.30994 | 35 |
| >=10% weight reduction | 48-56 wk | with T2D | 4 | 2 | 4 | 1 | 1 | 0.66667 | 1 |
| >=10% weight reduction | 48-56 wk | mixed population | 3 | 2 | 2 | 0 | 1 | 0.66667 | 0 |
| >=10% weight reduction | 20-28 wk | without T2D | 11 | 5 | 19 | 2 | 1 | 0.34545 | 9 |
| >=10% weight reduction | 10-14 wk | without T2D | 3 | 1 | 3 | 1 | 1 | 1 | 1 |
| >=15% weight reduction | 48-56 wk | without T2D | 9 | 5 | 11 | 2 | 2 | 0.30556 | 4 |
| >=15% weight reduction | 20-28 wk | without T2D | 3 | 2 | 2 | 0 | 1 | 0.66667 | 0 |
| >=20% weight reduction | 48-56 wk | without T2D | 5 | 3 | 7 | 1 | 1 | 0.7 | 3 |
| All-cause permanent discontinuation | 48-56 wk | without T2D | 32 | 14 | 87 | 8 | 1 | 0.1754 | 56 |
| All-cause permanent discontinuation | 20-28 wk | without T2D | 20 | 7 | 59 | 3 | 1 | 0.31053 | 40 |
| AE-related permanent discontinuation | 48-56 wk | without T2D | 37 | 17 | 106 | 9 | 1 | 0.15916 | 70 |
| AE-related permanent discontinuation | 48-56 wk | with T2D | 11 | 6 | 20 | 3 | 1 | 0.36364 | 10 |
| AE-related permanent discontinuation | 48-56 wk | mixed population | 4 | 2 | 4 | 1 | 1 | 0.66667 | 1 |
| AE-related permanent discontinuation | 20-28 wk | without T2D | 40 | 12 | 117 | 8 | 2 | 0.15 | 79 |
| Serious adverse events | 48-56 wk | without T2D | 37 | 15 | 106 | 9 | 1 | 0.15916 | 70 |
| Serious adverse events | 48-56 wk | with T2D | 11 | 5 | 20 | 3 | 1 | 0.36364 | 10 |
| Serious adverse events | 48-56 wk | mixed population | 6 | 3 | 5 | 1 | 2 | 0.33333 | 1 |
| Serious adverse events | 20-28 wk | without T2D | 41 | 10 | 133 | 8 | 1 | 0.1622 | 93 |
| Nausea | 48-56 wk | without T2D | 15 | 10 | 23 | 5 | 1 | 0.21905 | 9 |
| Nausea | 48-56 wk | with T2D | 4 | 2 | 4 | 1 | 1 | 0.66667 | 1 |
| Nausea | 20-28 wk | without T2D | 22 | 6 | 67 | 4 | 1 | 0.29004 | 46 |
| Nausea | 10-14 wk | without T2D | 5 | 3 | 3 | 0 | 2 | 0.3 | 0 |
| Vomiting | 48-56 wk | without T2D | 11 | 5 | 18 | 4 | 1 | 0.32727 | 8 |
| Vomiting | 48-56 wk | with T2D | 4 | 2 | 4 | 1 | 1 | 0.66667 | 1 |
| Vomiting | 20-28 wk | without T2D | 15 | 4 | 45 | 3 | 1 | 0.42857 | 31 |
| Diarrhoea | 48-56 wk | without T2D | 10 | 6 | 14 | 3 | 1 | 0.31111 | 5 |
| Diarrhoea | 48-56 wk | with T2D | 4 | 2 | 4 | 1 | 1 | 0.66667 | 1 |
| Diarrhoea | 20-28 wk | without T2D | 13 | 4 | 37 | 2 | 1 | 0.47436 | 25 |
| Constipation | 48-56 wk | without T2D | 5 | 4 | 6 | 2 | 1 | 0.6 | 2 |
| Constipation | 48-56 wk | with T2D | 4 | 2 | 4 | 1 | 1 | 0.66667 | 1 |
| Constipation | 20-28 wk | without T2D | 13 | 3 | 42 | 2 | 1 | 0.53846 | 30 |

**Table S4:**
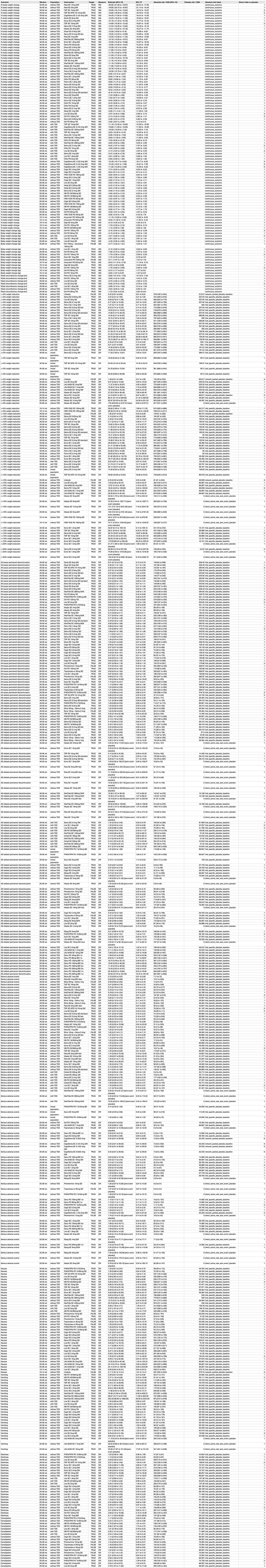
Network meta-analysis estimates versus placebo (all protocol outcomes, windows and populations) MO, mean difference; OR, odds ratio: Pl, prediction interval. Absolute risks use trial-specific placebo baseline risk where available; contrasts with zero placebo events report the direct active-arm rate with 527 Wilson 95% Cl

**Table S5.** Primary longitudinal network meta-analysis at 52 weeks, support classification and certainty of evidence (adults without T2D, treatment-policy) Support status per protocol sections 20, 41 and 56: observed_within_4w (direct observation within 48-56 weeks), interpolated (observations both before and after 52 weeks), borrowing_dependent_single_late_timepoint (single trial endpoint at 64-80 weeks informed by longitudinal trajectory), and extrapolated_beyond_followup (follow-up <48 weeks; excluded from 52-week ranking and decision table). Domain-by-domain CINeMA/GRADE evaluations are reported in Table S23.

| Intervention | Longitudinal trials | Observed weeks | Week 52 support status | MD vs placebo at 52 wk (95% CrI) | 95% PrI | P(MD < -5 pp) | P(MD < -10 pp) | Median rank (95% CrI) | SUCRA | Certainty (CINeMA/GRADE) |
|---|---|---|---|---|---|---|---|---|---|---|
| Reta SC 12mg QW | 1 | 52;80 | observed_within_4w_figure_derived (tabular_80w) | -19.80 (-21.81 to -17.78) | -22.26 to -17.18 | 1.0000 | 1 | 1 (1 to 2) | 0.9975 | Low |
| Reta SC 9mg QW | 1 | 52;80 | observed_within_4w_figure_derived (tabular_80w) | -18.64 (-20.62 to -16.67) | -21.18 to -16.15 | 1.0000 | 1 | 2 (1 to 5) | 0.9728 | Low |
| TZP SC 15mg QW | 3 | 52;72 | observed_within_4w_tabular | -16.87 (-18.07 to -15.51) | -18.75 to -14.77 | 1.0000 | 1 | 4 (2 to 7) | 0.9335 | High |
| TZP SC MTD 10-15mg QW | 2 | 52 | observed_within_4w_tabular | -16.47 (-18.04 to -14.85) | -18.62 to -14.28 | 1.0000 | 1 | 5 (2 to 9) | 0.9153 | Moderate |
| CagriSema SC 2.4/2.4mg QW | 2 | 20;52;68 | observed_within_4w_figure_derived (tabular_20w_68w) | -16.37 (-18.10 to -14.70) | -18.65 to -14.20 | 1.0000 | 1 | 5 (3 to 9) | 0.911 | Moderate |
| CagriSema SC 4.5/2.4mg QW | 1 | 20 | extrapolated_beyond_followup_class_borrowing | -16.08 (-24.14 to -7.04) | -24.11 to -6.96 | 0.9867 | 0.9335 | Not ranked (extrapolated) | NA | Low |
| CagriSema SC 1.2/2.4mg QW | 1 | 20 | extrapolated_beyond_followup_class_borrowing | -15.89 (-23.47 to -7.41) | -23.55 to -7.20 | 0.9893 | 0.9388 | Not ranked (extrapolated) | NA | Low |
| TZP SC 10mg QW | 2 | 52;72 | observed_within_4w_tabular | -15.19 (-16.52 to -13.79) | -17.15 to -13.09 | 1.0000 | 1 | 7 (5 to 12) | 0.8561 | Moderate |
| Eloral SC 9mg QW | 1 | 48 | observed_within_4w_tabular | -14.65 (-19.34 to -9.91) | -19.58 to -9.69 | 1.0000 | 0.9723 | 8 (2 to 25) | 0.8099 | Moderate |
| Ecno SC 2.4mg QW | 1 | 40;48 | observed_within_4w_tabular | -14.42 (-16.79 to -11.89) | -17.10 to -11.38 | 1.0000 | 0.9995 | 9 (4 to 18) | 0.8077 | Moderate |
| Sema SC 7.2mg QW | 1 | 56;72 | observed_within_4w_figure_derived (tabular_72w) | -14.26 (-15.76 to -12.75) | -16.36 to -12.18 | 1.0000 | 1 | 9 (6 to 16) | 0.8052 | Moderate |
| Bofan SC 24mg QW | 1 | 30 | extrapolated_beyond_followup_class_borrowing | -13.30 (-19.12 to -7.58) | -19.22 to -7.40 | 0.9967 | 0.8773 | Not ranked (extrapolated) | NA | Low |
| Sema SC 0.4mg QD fast | 1 | 52 | observed_within_4w_tabular | -13.26 (-15.22 to -11.29) | -15.61 to -10.81 | 1.0000 | 0.999 | 13 (7 to 22) | 0.731 | Moderate |
| Reta SC 4mg QW | 1 | 52;80 | observed_within_4w_figure_derived (tabular_80w) | -13.21 (-15.16 to -11.24) | -15.52 to -10.80 | 1.0000 | 0.9975 | 13 (7 to 22) | 0.7276 | Low |
| Eloral SC 6-9mg QW | 1 | 48 | observed_within_4w_tabular | -13.16 (-18.65 to -7.65) | -18.85 to -7.63 | 0.9985 | 0.876 | 13 (3 to 34) | 0.6968 | Moderate |
| Mazdu SC 6mg QW | 1 | 32;48 | observed_within_4w_tabular | -12.96 (-15.00 to -10.56) | -15.37 to -10.13 | 1.0000 | 0.9898 | 14 (7 to 24) | 0.7024 | Moderate |
| Bofan SC 48mg Q2W | 1 | 30 | extrapolated_beyond_followup_class_borrowing | -12.88 (-18.61 to -7.15) | -18.65 to -7.08 | 0.9968 | 0.8603 | Not ranked (extrapolated) | NA | Low |
| Sema PO 50mg QD | 1 | 52;68 | observed_within_4w_figure_derived (tabular_68w) | -12.69 (-14.44 to -10.75) | -14.99 to -10.20 | 1.0000 | 0.9947 | 15 (8 to 25) | 0.6818 | Moderate |
| MariTide SC 280mg Q4W | 1 | 52 | observed_within_4w_tabular | -12.62 (-15.33 to -9.76) | -15.76 to -9.49 | 1.0000 | 0.9637 | 15 (7 to 26) | 0.678 | Moderate |
| Cagri SC 4.5mg QW | 1 | 26 | extrapolated_beyond_followup_class_borrowing | -12.51 (-19.88 to -3.97) | -20.09 to -3.80 | 0.9593 | 0.7523 | Not ranked (extrapolated) | NA | Very low |
| Eloral SC 3-9mg QW | 1 | 48 | observed_within_4w_tabular | -12.47 (-16.84 to -8.11) | -17.04 to -7.90 | 0.9997 | 0.872 | 16 (5 to 33) | 0.6494 | Moderate |
| Ecno SC 1.8mg QW | 1 | 40;48 | observed_within_4w_tabular | -12.42 (-14.76 to -9.98) | -15.12 to -9.50 | 1.0000 | 0.9742 | 16 (8 to 27) | 0.6552 | Moderate |
| Cagri SC 2.4mg QW | 1 | 26 | extrapolated_beyond_followup_class_borrowing | -12.07 (-19.34 to -3.75) | -19.45 to -3.59 | 0.9573 | 0.7187 | Not ranked (extrapolated) | NA | Very low |
| Eloral SC 6mg QW | 1 | 48 | observed_within_4w_tabular | -11.93 (-17.73 to -5.85) | -17.97 to -5.75 | 0.9885 | 0.7398 | 18 (4 to 38) | 0.5942 | Moderate |
| Cagri SC 1.2mg QW | 1 | 26 | extrapolated_beyond_followup_class_borrowing | -11.71 (-19.33 to -3.06) | -19.48 to -3.02 | 0.9455 | 0.6693 | Not ranked (extrapolated) | NA | Very low |
| MariTide SC 420mg Q4W | 1 | 52 | observed_within_4w_tabular | -11.60 (-14.39 to -8.65) | -14.76 to -8.28 | 1.0000 | 0.8687 | 20 (9 to 32) | 0.579 | Moderate |
| Sema SC 2.4mg QW | 13 | 20;40;44;52;56;60;68;72 | observed_within_4w_tabular | -11.60 (-12.32 to -10.89) | -13.34 to -9.92 | 1.0000 | 1 | 20 (14 to 25) | 0.5823 | High |
| Sema PO 25mg QD | 1 | 56;64 | observed_within_4w_figure_derived (tabular_64w) | -11.51 (-14.15 to -8.72) | -14.51 to -8.37 | 1.0000 | 0.8597 | 20 (9 to 32) | 0.571 | Moderate |
| Bofan SC 24mg Q2W | 1 | 30 | extrapolated_beyond_followup_class_borrowing | -11.40 (-17.19 to -5.77) | -17.27 to -5.62 | 0.9852 | 0.6977 | Not ranked (extrapolated) | NA | Low |
| TZP SC 5mg QW | 1 | 52;72 | observed_within_4w_figure_derived (tabular_72w) | -11.26 (-12.66 to -9.80) | -13.22 to -9.14 | 1.0000 | 0.9592 | 21 (14 to 29) | 0.5473 | Moderate |
| MariTide SC 140mg Q4W | 1 | 52 | observed_within_4w_tabular | -11.15 (-13.90 to -8.37) | -14.23 to -7.99 | 1.0000 | 0.799 | 22 (11 to 33) | 0.5355 | Moderate |
| Cagri SC 0.6mg QW | 1 | 26 | extrapolated_beyond_followup_class_borrowing | -11.04 (-18.68 to -2.90) | -18.82 to -2.61 | 0.9327 | 0.6162 | Not ranked (extrapolated) | NA | Very low |
| VCT220 PO 160mg QD fast | 1 | 16 | extrapolated_beyond_followup_class_borrowing | -10.95 (-17.36 to -4.12) | -17.52 to -4.10 | 0.9603 | 0.6187 | Not ranked (extrapolated) | NA | Very low |
| Sema SC 0.4mg QD standard | 1 | 52 | observed_within_4w_tabular | -10.93 (-12.86 to -8.90) | -13.27 to -8.42 | 1.0000 | 0.8318 | 23 (14 to 31) | 0.5181 | Moderate |
| Bofan SC 18mg Q2W | 1 | 30 | extrapolated_beyond_followup_class_borrowing | -10.81 (-16.34 to -4.90) | -16.61 to -4.87 | 0.9737 | 0.6158 | Not ranked (extrapolated) | NA | Very low |
| VCT220 PO 160mg QD slow | 1 | 16 | extrapolated_beyond_followup_class_borrowing | -10.80 (-17.12 to -3.89) | -17.31 to -3.79 | 0.9547 | 0.6055 | Not ranked (extrapolated) | NA | Very low |
| Cagri SC 0.3mg QW | 1 | 26 | extrapolated_beyond_followup_class_borrowing | -10.79 (-19.04 to -2.69) | -19.04 to -2.84 | 0.9323 | 0.5853 | Not ranked (extrapolated) | NA | Very low |
| Sema SC 1mg QW | 1 | 16 | extrapolated_beyond_followup_class_borrowing | -10.54 (-17.15 to -3.65) | -17.37 to -3.53 | 0.9483 | 0.5712 | Not ranked (extrapolated) | NA | Very low |
| MariTide SC 420mg Q8W | 1 | 52 | observed_within_4w_tabular | -10.40 (-13.48 to -7.19) | -13.69 to -6.91 | 0.9998 | 0.6082 | 25 (13 to 37) | 0.4598 | Moderate |
| Mazdu SC 4mg QW | 1 | 32;48 | observed_within_4w_tabular | -10.28 (-12.21 to -8.02) | -12.64 to -7.46 | 1.0000 | 0.6098 | 26 (16 to 35) | 0.4499 | Moderate |
| Pemvi SC 2.4mg QW | 1 | 48 | observed_within_4w_tabular | -10.23 (-12.54 to -7.84) | -12.97 to -7.52 | 0.9998 | 0.577 | 26 (15 to 34) | 0.4538 | Moderate |
| VCT220 PO 120mg QD | 1 | 16 | extrapolated_beyond_followup_class_borrowing | -9.96 (-16.35 to -3.40) | -16.46 to -3.16 | 0.9363 | 0.4953 | Not ranked (extrapolated) | NA | Very low |
| Eloral SC 3mg QW | 1 | 48 | observed_within_4w_tabular | -9.65 (-15.04 to -4.14) | -15.27 to -4.01 | 0.9515 | 0.4495 | 28 (10 to 41) | 0.4078 | Low |
| HRS-7535 PO 180mg QD | 1 | 26 | extrapolated_beyond_followup_class_borrowing | -9.57 (-15.43 to -3.47) | -15.67 to -3.32 | 0.9342 | 0.4412 | Not ranked (extrapolated) | NA | Very low |
| Ecno SC 1.2mg QW | 1 | 40;48 | observed_within_4w_tabular | -9.44 (-11.64 to -7.00) | -11.95 to -6.44 | 0.9993 | 0.3148 | 29 (19 to 38) | 0.3763 | Moderate |
| Orfor PO 36mg QD | 1 | 52;72 | observed_within_4w_figure_derived (tabular_72w) | -9.35 (-11.16 to -7.87) | -11.85 to -7.38 | 1.0000 | 0.211 | 29 (20 to 36) | 0.3743 | Moderate |
| Noiig SC 0.12mg QD | 1 | 24 | extrapolated_beyond_followup_class_borrowing | -9.22 (-15.27 to -3.17) | -15.42 to -2.98 | 0.9225 | 0.3907 | Not ranked (extrapolated) | NA | Very low |
| VCT220 PO 80mg QD | 1 | 16 | extrapolated_beyond_followup_class_borrowing | -9.10 (-15.71 to -2.89) | -15.88 to -2.73 | 0.9063 | 0.3782 | Not ranked (extrapolated) | NA | Very low |
| Pemvi SC 1.8mg QW | 1 | 48 | observed_within_4w_tabular | -9.07 (-11.47 to -6.56) | -11.86 to -6.19 | 0.9988 | 0.2272 | 31 (20 to 38) | 0.349 | Moderate |
| Noiig SC 0.36mg QD | 1 | 24 | extrapolated_beyond_followup_class_borrowing | -9.04 (-15.32 to -3.07) | -15.51 to -2.86 | 0.9167 | 0.3678 | Not ranked (extrapolated) | NA | Very low |
| Beinag SC 0.2mg TID | 1 | 16 | extrapolated_beyond_followup_class_borrowing | -9.03 (-15.58 to -2.91) | -15.69 to -2.66 | 0.9037 | 0.375 | Not ranked (extrapolated) | NA | Very low |
| Bofan SC 12mg Q2W | 1 | 30 | extrapolated_beyond_followup_class_borrowing | -8.95 (-14.42 to -3.28) | -14.54 to -3.11 | 0.9253 | 0.3493 | Not ranked (extrapolated) | NA | Very low |
| Noiig SC 0.24mg QD | 1 | 24 | extrapolated_beyond_followup_class_borrowing | -8.86 (-15.16 to -3.11) | -15.38 to -2.86 | 0.9103 | 0.3477 | Not ranked (extrapolated) | NA | Very low |
| Sema SC 0.2mg QD | 1 | 52 | observed_within_4w_tabular | -8.79 (-10.76 to -6.86) | -11.11 to -6.37 | 1.0000 | 0.1118 | 32 (23 to 38) | 0.324 | Moderate |
| HRS-7535 PO 60mg QD | 1 | 26 | extrapolated_beyond_followup_class_borrowing | -8.73 (-14.75 to -2.65) | -14.99 to -2.58 | 0.9010 | 0.33 | Not ranked (extrapolated) | NA | Very low |
| Sema SC 0.3mg QD fast | 1 | 52 | observed_within_4w_tabular | -8.62 (-10.59 to -6.64) | -11.09 to -6.12 | 1.0000 | 0.0793 | 32 (24 to 38) | 0.3093 | Moderate |
| Sema SC 0.3mg QD standard | 1 | 52 | observed_within_4w_tabular | -8.41 (-10.36 to -6.47) | -10.78 to -5.95 | 0.9998 | 0.0593 | 33 (25 to 39) | 0.2914 | Moderate |
| HRS-7535 PO 120mg QD | 1 | 26 | extrapolated_beyond_followup_class_borrowing | -8.36 (-14.76 to -2.38) | -14.88 to -2.10 | 0.8788 | 0.2842 | Not ranked (extrapolated) | NA | Very low |
| Survo SC 6mg QW | 1 | 48;76 | observed_within_4w_figure_derived (tabular_76w) | -8.10 (-10.42 to -5.56) | -10.86 to -5.25 | 0.9917 | 0.0575 | 34 (25 to 40) | 0.2749 | Moderate |
| Eloral SC 1mg QW | 1 | 48 | observed_within_4w_tabular | -8.05 (-14.37 to -1.24) | -14.59 to -1.14 | 0.8178 | 0.2813 | 35 (12 to 45) | 0.3022 | Low |
| Pemvi SC 1.2mg QW | 1 | 48 | observed_within_4w_tabular | -7.63 (-10.06 to -5.01) | -10.39 to -4.67 | 0.9752 | 0.0283 | 36 (27 to 41) | 0.236 | Low |
| Survo SC 3.6mg QW | 1 | 48;76 | observed_within_4w_figure_derived (tabular_76w) | -7.32 (-9.61 to -4.68) | -10.09 to -4.34 | 0.9578 | 0.0103 | 37 (28 to 41) | 0.2146 | Low |
| HRS-7535 PO 30mg QD | 1 | 26 | extrapolated_beyond_followup_class_borrowing | -6.89 (-13.28 to -1.05) | -13.44 to -0.86 | 0.7530 | 0.1462 | Not ranked (extrapolated) | NA | Very low |
| Orfor PO 12mg QD | 1 | 52;72 | observed_within_4w_figure_derived (tabular_72w) | -6.78 (-8.67 to -5.26) | -9.26 to -4.70 | 0.9888 | 0.0008 | 38 (32 to 40) | 0.1905 | Low |
| NB PO 50/300mg QD | 1 | 12 | extrapolated_beyond_followup_class_borrowing | -6.53 (-23.80 to 10.82) | -23.78 to 10.94 | 0.5690 | 0.3478 | Not ranked (extrapolated) | NA | Very low |
| NB PO 16/360mg QD | 1 | 24 | extrapolated_beyond_followup_class_borrowing | -6.42 (-23.98 to 10.95) | -24.01 to 11.10 | 0.5612 | 0.3375 | Not ranked (extrapolated) | NA | Very low |
| NB PO 48/360mg QD | 1 | 24 | extrapolated_beyond_followup_class_borrowing | -6.38 (-24.16 to 11.50) | -24.13 to 11.44 | 0.5662 | 0.3447 | Not ranked (extrapolated) | NA | Very low |
| NB PO 32/360mg QD | 2 | 16;24 | extrapolated_beyond_followup_class_borrowing | -6.26 (-23.79 to 10.84) | -23.78 to 10.69 | 0.5585 | 0.3338 | Not ranked (extrapolated) | NA | Very low |
| Orfor PO 6mg QD | 1 | 52;72 | observed_within_4w_figure_derived (tabular_72w) | -5.97 (-7.86 to -4.49) | -8.41 to -3.90 | 0.8937 | 0.0003 | 40 (35 to 42) | 0.1444 | Low |
| Sema SC 0.1mg QD | 1 | 52 | observed_within_4w_tabular | -5.97 (-7.88 to -3.97) | -8.32 to -3.50 | 0.8330 | 0 | 40 (35 to 42) | 0.1473 | Low |
| Amycretin PO 100mg QD | 1 | 12.14 | extrapolated_beyond_followup_class_borrowing | -4.86 (-29.70 to 20.31) | -29.77 to 20.30 | 0.4947 | 0.3472 | Not ranked (extrapolated) | NA | Very low |
| Amycretin PO 50mg QD | 1 | 12.14 | extrapolated_beyond_followup_class_borrowing | -4.71 (-30.18 to 20.49) | -30.35 to 20.61 | 0.4927 | 0.3407 | Not ranked (extrapolated) | NA | Very low |
| Lira SC 3mg QD | 6 | 17;26;40;52;56;68 | observed_within_4w_tabular | -4.20 (-5.49 to -2.89) | -6.18 to -2.21 | 0.1077 | 0 | 42 (40 to 44) | 0.0873 | Low |
| Sema SC 0.05mg QD | 1 | 52 | observed_within_4w_tabular | -3.50 (-5.42 to -1.48) | -5.89 to -1.10 | 0.0615 | 0 | 43 (41 to 45) | 0.0646 | Low |
| Orli PO 120mg TID | 4 | 12;52;78 | observed_within_4w_tabular | -3.11 (-4.23 to -1.93) | -4.99 to -1.21 | 0.0008 | 0 | 44 (42 to 45) | 0.0562 | Low |
| Orli PO 60mg TID | 2 | 24;52 | observed_within_4w_tabular | -2.14 (-3.55 to -0.68) | -4.21 to 0.01 | 0.0002 | 0 | 45 (43 to 45) | 0.0277 | Low |
| Dapi SC 6mg QW | 1 | 12 | extrapolated_beyond_followup_class_borrowing | -1.05 (-26.34 to 23.49) | -26.26 to 23.82 | 0.3755 | 0.238 | Not ranked (extrapolated) | NA | Very low |

**Table S6.** Heterogeneity and global inconsistency. tau on the MD scale (continuous) or log-OR scale (binary), extracted from the between-trial variance component of rma.mv.

| Outcome | Window | Population | Interventions | Trials | Model | tau | Q (total) | df | p (Q) | I2 (%) | Q between designs | df between | p (between designs) |
|---|---|---|---|---|---|---|---|---|---|---|---|---|---|
| % body-weight change | 48-56 wk | without T2D | 46 | 28 | random_effects_consistency_REML_rho0.5 | 0.67649 | 25.681 | 14 | 0.028412 | 45.485 | 14.842 | 8 | 0.062282 |
| % body-weight change | 48-56 wk | with T2D | 15 | 9 | random_effects_consistency_REML_rho0.5 | 0.000076662 | 0.15493 | 3 | 0.98451 | 0 | 0.15493 | 3 | 0.98451 |
| % body-weight change | 20-28 wk | without T2D | 22 | 9 | common_effect_heterogeneity_not_estimable | 0 | 0 | 0 | 1 |  | 3.2585E-29 | 0 |  |
| % body-weight change | 10-14 wk | without T2D | 6 | 4 | common_effect_heterogeneity_not_estimable | 0 | 0 | 0 | 1 |  | 4.2744E-30 | 0 |  |
| Body-weight change (kg) | 48-56 wk | without T2D | 3 | 3 | random_effects_consistency_REML_rho0.5 | 0.000091286 | 0.41798 | 2 | 0.8114 | 0 | 0.13776 | 1 | 0.71052 |
| Body-weight change (kg) | 48-56 wk | with T2D | 3 | 2 | common_effect_heterogeneity_not_estimable | 0 | 1.7281 | 1 | 0.18866 | 42.132 | 1.7281 | 1 | 0.18866 |
| Body-weight change (kg) | 20-28 wk | without T2D | 8 | 5 | random_effects_consistency_REML_rho0.5 | 0.031621 | 0.20263 | 1 | 0.65261 | 0 | 0.20263 | 1 | 0.65261 |
| Body-weight change (kg) | 10-14 wk | without T2D | 6 | 4 | random_effects_consistency_REML_rho0.5 | 0.000092376 | 0.022233 | 1 | 0.88147 | 0 | 0.022233 | 1 | 0.88147 |
| Waist circumference change (cm) | 48-56 wk | without T2D | 4 | 3 | common_effect_heterogeneity_not_estimable | 0 | 0 | 0 | 1 |  |  |  |  |
| Waist circumference change (cm) | 48-56 wk | with T2D | 3 | 2 | common_effect_heterogeneity_not_estimable | 0 | 0.38367 | 1 | 0.53565 | 0 | 0.38367 | 1 | 0.53565 |
| Waist circumference change (cm) | 20-28 wk | without T2D | 3 | 2 | common_effect_heterogeneity_not_estimable | 0 | 0 | 0 | 1 |  |  |  |  |
| >=5% weight reduction | 48-56 wk | without T2D | 19 | 6 | common_effect_heterogeneity_not_estimable | 0 | 0 | 0 | 1 |  | 2.8523E-29 | 0 |  |
| >=5% weight reduction | 48-56 wk | with T2D | 5 | 2 | common_effect_heterogeneity_not_estimable | 0 | 0 | 0 | 1 |  | 3.1981E-30 | 0 |  |
| >=5% weight reduction | 48-56 wk | mixed population | 6 | 3 | common_effect_heterogeneity_not_estimable | 0 | 0 | 0 | 1 |  | 1.336E-29 | 0 |  |
| >=5% weight reduction | 20-28 wk | without T2D | 11 | 5 | common_effect_heterogeneity_not_estimable | 0 | 0 | 0 | 1 |  | 1.9598E-29 | 0 |  |
| >=5% weight reduction | 10-14 wk | without T2D | 3 | 1 | common_effect_heterogeneity_not_estimable | 0 | 0 | 0 | 1 |  | 0 | 0 |  |
| >=10% weight reduction | 48-56 wk | without T2D | 19 | 7 | random_effects_consistency_REML_rho0.5 | 0.16459 | 1.1707 | 1 | 0.27926 | 14.582 | 0 | 0 |  |
| >=10% weight reduction | 48-56 wk | with T2D | 4 | 2 | common_effect_heterogeneity_not_estimable | 0 | 0 | 0 | 1 |  | -8.8022E-30 | 0 |  |
| >=10% weight reduction | 48-56 wk | mixed population | 3 | 2 | common_effect_heterogeneity_not_estimable | 0 | 0 | 0 | 1 |  |  |  |  |
| >=10% weight reduction | 20-28 wk | without T2D | 11 | 5 | common_effect_heterogeneity_not_estimable | 0 | 0 | 0 | 1 |  | 2.6696E-28 | 0 |  |
| >=10% weight reduction | 10-14 wk | without T2D | 3 | 1 | common_effect_heterogeneity_not_estimable | 0 | 0 | 0 | 1 |  | 2.522E-30 | 0 |  |
| >=15% weight reduction | 48-56 wk | without T2D | 7 | 4 | random_effects_consistency_REML_rho0.5 | 0.00010347 | 0.27314 | 1 | 0.60123 | 0 | 0 | 0 |  |
| >=15% weight reduction | 20-28 wk | without T2D | 3 | 2 | common_effect_heterogeneity_not_estimable | 0 | 0 | 0 | 1 |  |  |  |  |
| >=20% weight reduction | 48-56 wk | without T2D | 5 | 3 | random_effects_consistency_REML_rho0.5 | 0.00021644 | 0.1215 | 1 | 0.72742 | 0 | 0 | 0 |  |
| All-cause permanent discontinuation | 48-56 wk | without T2D | 32 | 14 | random_effects_consistency_REML_rho0.5 | 0.00001821 | 0.38895 | 4 | 0.98337 | 0 | 0.055644 | 2 | 0.97256 |
| All-cause permanent discontinuation | 20-28 wk | without T2D | 20 | 7 | random_effects_consistency_REML_rho0.5 | 0.000062299 | 0.4222 | 2 | 0.80969 | 0 | 0.4222 | 2 | 0.80969 |
| AE-related permanent discontinuation | 48-56 wk | without T2D | 37 | 17 | random_effects_consistency_REML_rho0.5 | 0.000032974 | 3.8469 | 7 | 0.79723 | 0 | 0.12549 | 3 | 0.98861 |
| AE-related permanent discontinuation | 48-56 wk | with T2D | 11 | 6 | random_effects_consistency_REML_rho0.5 | 0.55094 | 4.1334 | 2 | 0.1266 | 51.614 | 4.1334 | 2 | 0.1266 |
| AE-related permanent discontinuation | 48-56 wk | mixed population | 4 | 2 | common_effect_heterogeneity_not_estimable | 0 | 0 | 0 | 1 |  | 1.2973E-31 | 0 |  |
| AE-related permanent discontinuation | 20-28 wk | without T2D | 38 | 11 | random_effects_consistency_REML_rho0.5 | 0.00016275 | 1.5724 | 3 | 0.66566 | 0 | 1.5724 | 3 | 0.66566 |
| Serious adverse events | 48-56 wk | without T2D | 37 | 15 | random_effects_consistency_REML_rho0.5 | 0.53941 | 11.645 | 5 | 0.039996 | 57.062 | 7.5847 | 3 | 0.055421 |
| Serious adverse events | 48-56 wk | with T2D | 11 | 5 | random_effects_consistency_REML_rho0.5 | 0.000056331 | 0.1774 | 1 | 0.67362 | 0 | 0.1774 | 1 | 0.67362 |
| Serious adverse events | 48-56 wk | mixed population | 4 | 2 | common_effect_heterogeneity_not_estimable | 0 | 0 | 0 | 1 |  | -3.8799E-32 | 0 |  |
| Serious adverse events | 20-28 wk | without T2D | 41 | 10 | random_effects_consistency_REML_rho0.5 | 0.0003629 | 2.4511 | 3 | 0.48419 | 0 | 2.4511 | 3 | 0.48419 |
| Nausea | 48-56 wk | without T2D | 15 | 10 | random_effects_consistency_REML_rho0.5 | 0.13102 | 3.7017 | 3 | 0.29553 | 18.956 | 2.4707 | 1 | 0.11598 |
| Nausea | 48-56 wk | with T2D | 4 | 2 | common_effect_heterogeneity_not_estimable | 0 | 0 | 0 | 1 |  | 0 | 0 |  |
| Nausea | 20-28 wk | without T2D | 22 | 6 | random_effects_consistency_REML_rho0.5 | 0.80292 | 9.1605 | 2 | 0.010253 | 78.167 | 9.1605 | 2 | 0.010253 |
| Nausea | 10-14 wk | without T2D | 3 | 2 | common_effect_heterogeneity_not_estimable | 0 | 0 | 0 | 1 |  |  |  |  |
| Vomiting | 48-56 wk | without T2D | 11 | 5 | random_effects_consistency_REML_rho0.5 | 0.54413 | 4.0708 | 1 | 0.043632 | 75.435 | 4.0708 | 1 | 0.043632 |
| Vomiting | 48-56 wk | with T2D | 4 | 2 | common_effect_heterogeneity_not_estimable | 0 | 0 | 0 | 1 |  | 2.11E-30 | 0 |  |
| Vomiting | 20-28 wk | without T2D | 15 | 4 | random_effects_consistency_REML_rho0.5 | 0.00016206 | 0.6854 | 2 | 0.70985 | 0 | 0.6854 | 2 | 0.70985 |
| Diarrhoea | 48-56 wk | without T2D | 10 | 6 | random_effects_consistency_REML_rho0.5 | 8.4784E-06 | 0.08137 | 1 | 0.77545 | 0 | 0 | 0 |  |
| Diarrhoea | 48-56 wk | with T2D | 4 | 2 | common_effect_heterogeneity_not_estimable | 0 | 0 | 0 | 1 |  | -1.5931E-30 | 0 |  |
| Diarrhoea | 20-28 wk | without T2D | 13 | 4 | random_effects_consistency_REML_rho0.5 | 0.000029788 | 0.062018 | 1 | 0.80333 | 0 | 0.062018 | 1 | 0.80333 |
| Constipation | 48-56 wk | without T2D | 5 | 4 | random_effects_consistency_REML_rho0.5 | 0.13986 | 2.9614 | 2 | 0.22748 | 32.464 | 0.64179 | 1 | 0.42306 |
| Constipation | 48-56 wk | with T2D | 4 | 2 | common_effect_heterogeneity_not_estimable | 0 | 0 | 0 | 1 |  | 1.8974E-31 | 0 |  |
| Constipation | 20-28 wk | without T2D | 13 | 3 | random_effects_consistency_REML_rho0.5 | 0.00012071 | 0.46514 | 1 | 0.49523 | 0 | 0.46514 | 1 | 0.49523 |

Note: Discrepancies between the total trial counts in descriptive network geometry (Table S3) and analytical inconsistency models (Table S6) are strictly attributable to network connectivity constraints. In disconnected sub-networks (e.g., % body-weight change at 20–28 weeks without T2D, where 2 unconnected intervention arms from 1 trial formed an isolated island; and >=15% weight reduction at 48–56 weeks, where 2 nodes from 1 trial lacked bridging paths to the reference comparator), disconnected arms are computationally non-estimable within global contrast-based variance matrices and were therefore automatically partitioned from global design-by-treatment interaction models. All eligible trials and randomized arms remain fully accounted for in Table S2 and the complete study repository.

**Table S7:**
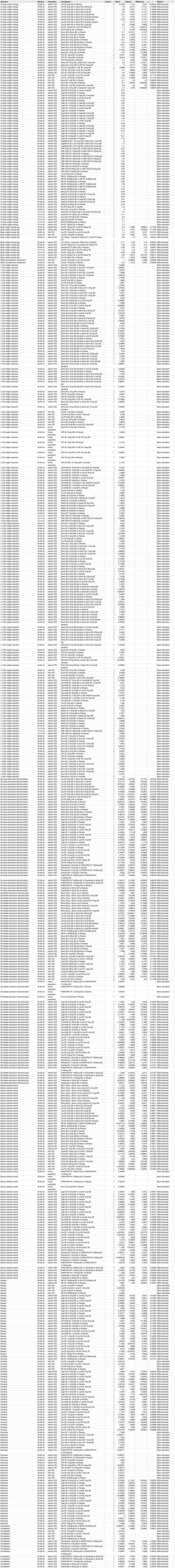
Local inconsistency (node-splitting) Binary outcomes on the log-OR scale.

**Table S8:**
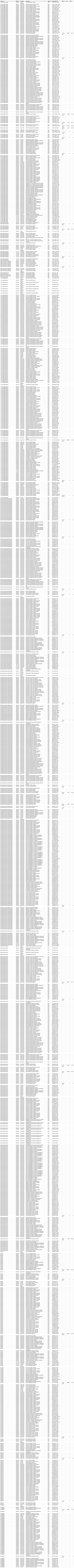
Pairwise meta-analyses of direct comparison in the primary network (Layer A, matching network estimand)

**Table S9.** Small-study effects and publication bias (comparison-adjusted Egger regression for scenarios with >=10 trials) Per protocol section 36. funnel plot asymmetry was tested using Egger regression only when at least 10 trials were available. The remaining 41 scenario(s) had fewer than 10 trials and were not tested.

| Outcome | Window | Population | Trials | Contrasts | Egger z | Egger p |
|---|---|---|---|---|---|---|
| % body-weight change | 48-56 wk | without T2D | 27 | 58 | 0.75769 | 0.45181 |
| All-cause permanent discontinuation | 48-56 wk | without T2D | 14 | 35 | -0.21669 | 0.82978 |
| AE-related permanent discontinuation | 48-56 wk | without T2D | 17 | 39 | 0.71105 | 0.48151 |
| AE-related permanent discontinuation | 20-28 wk | without T2D | 11 | 36 | -0.28839 | 0.7748 |
| Serious adverse events | 48-56 wk | without T2D | 15 | 37 | -0.35815 | 0.72238 |
| Nausea | 48-56 wk | without T2D | 10 | 17 | -0.0061223 | 0.9952 |

**Table S10.**
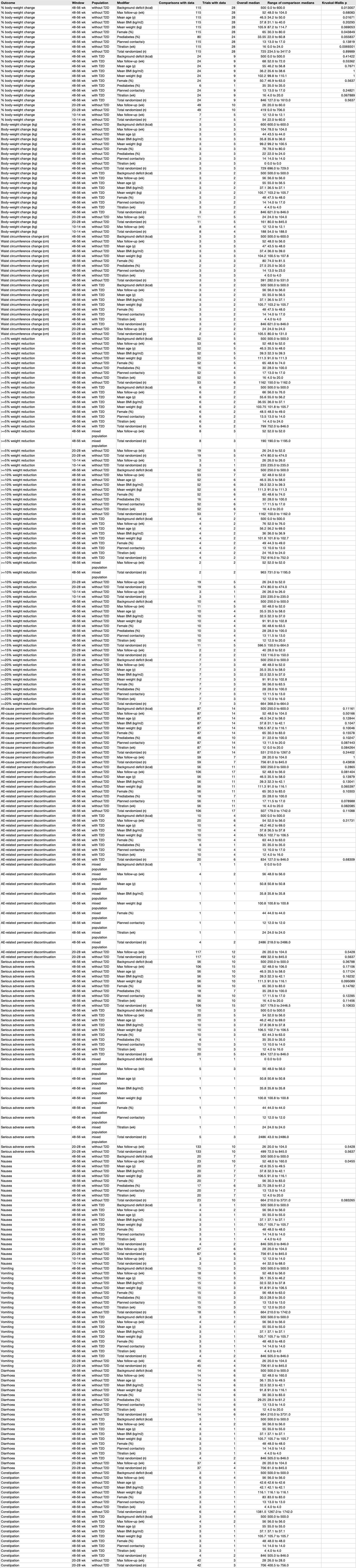
Transitivity: distribution of baseline effect modifiers across comparisons. Per-comparison modifier distributions are provided in per_outcome_transitivity_by_comparison.csv.

| Outcome | Window | Population | Modifier | Comparisons with data | Trials with data | Overall median | Range of comparison medians | Kruskal-Wallis p |
|---|---|---|---|---|---|---|---|---|
| % body-weight change | 48-56 wk | without T2D | Background deficit (kcal) | 115 | 28 | 500 | 0.0 to 600.0 | 0.013001 |
| % body-weight change | 48-56 wk | without T2D | Max follow-up (wk) | 115 | 28 | 52 | 48.0 to 104.0 | 0.68083 |
| % body-weight change | 48-56 wk | without T2D | Mean age (y) | 115 | 28 | 46.5 | 34.2 to 50.0 | 0.01671 |
| % body-weight change | 48-56 wk | without T2D | Mean BMI (kg/m2) | 115 | 28 | 37.8 | 31.1 to 40.0 | 0.20293 |
| % body-weight change | 48-56 wk | without T2D | Mean weight (kg) | 115 | 28 | 105.9 | 87.2 to 114.7 | 0.069053 |
| % body-weight change | 48-56 wk | without T2D | Female (%) | 115 | 28 | 65 | 30.3 to 80.0 | 0.043849 |
| % body-weight change | 48-56 wk | without T2D | Prediabetes (%) | 80 | 24 | 33.55 | 22.0 to 60.8 | 0.055837 |
| % body-weight change | 48-56 wk | without T2D | Planned contacts/y | 115 | 28 | 13 | 13.0 to 17.0 | 0.13819 |
| % body-weight change | 48-56 wk | without T2D | Titration (wk) | 115 | 28 | 16 | 0.0 to 24.0 | 0.0069301 |
| % body-weight change | 48-56 wk | without T2D | Total randomized (n) | 115 | 28 | 725 | 234.5 to 3417.0 | 0.89989 |
| % body-weight change | 48-56 wk | with T2D | Background deficit (kcal) | 24 | 9 | 500 | 0.0 to 500.0 | 0.41422 |
| % body-weight change | 48-56 wk | with T2D | Max follow-up (wk) | 24 | 9 | 68 | 52.0 to 72.0 | 0.55362 |
| % body-weight change | 48-56 wk | with T2D | Mean age (y) | 24 | 9 | 55 | 46.2 to 56.8 | 0.7671 |
| % body-weight change | 48-56 wk | with T2D | Mean BMI (kg/m2) | 24 | 9 | 36.2 | 35.6 to 38.6 | 1 |
| % body-weight change | 48-56 wk | with T2D | Mean weight (kg) | 24 | 9 | 102.2 | 99.6 to 110.1 | 1 |
| % body-weight change | 48-56 wk | with T2D | Female (%) | 24 | 9 | 50.7 | 46.9 to 65.0 | 0.5637 |
| % body-weight change | 48-56 wk | with T2D | Prediabetes (%) | 6 | 1 | 35 | 35.0 to 35.0 |  |
| % body-weight change | 48-56 wk | with T2D | Planned contacts/y | 24 | 9 | 13 | 13.0 to 17.0 | 0.24821 |
| % body-weight change | 48-56 wk | with T2D | Titration (wk) | 24 | 9 | 16 | 4.0 to 20.0 | 0.067889 |
| % body-weight change | 48-56 wk | with T2D | Total randomized (n) | 24 | 9 | 846 | 127.0 to 1613.0 | 0.5637 |
| % body-weight change | 20-28 wk | without T2D | Max follow-up (wk) | 49 | 10 | 26 | 20.0 to 60.0 |  |
| % body-weight change | 20-28 wk | without T2D | Total randomized (n) | 49 | 10 | 419 | 0.0 to 706.0 |  |
| % body-weight change | 10-14 wk | without T2D | Max follow-up (wk) | 7 | 5 | 12 | 12.0 to 12.1 |  |
| % body-weight change | 10-14 wk | without T2D | Total randomized (n) | 7 | 5 | 54 | 22.0 to 60.0 |  |
| Body-weight change (kg) | 48-56 wk | without T2D | Background deficit (kcal) | 3 | 3 | 600 | 600.0 to 600.0 |  |
| Body-weight change (kg) | 48-56 wk | without T2D | Max follow-up (wk) | 3 | 3 | 104 | 78.0 to 104.0 |  |
| Body-weight change (kg) | 48-56 wk | without T2D | Mean age (y) | 3 | 3 | 44 | 43.5 to 44.0 |  |
| Body-weight change (kg) | 48-56 wk | without T2D | Mean BMI (kg/m2) | 3 | 3 | 35.8 | 35.8 to 36.0 |  |
| Body-weight change (kg) | 48-56 wk | without T2D | Mean weight (kg) | 3 | 3 | 99.2 | 99.2 to 100.5 |  |
| Body-weight change (kg) | 48-56 wk | without T2D | Female (%) | 3 | 3 | 78 | 78.0 to 80.0 |  |
| Body-weight change (kg) | 48-56 wk | without T2D | Prediabetes (%) | 3 | 3 | 22 | 22.0 to 24.0 |  |
| Body-weight change (kg) | 48-56 wk | without T2D | Planned contacts/y | 3 | 3 | 14 | 14.0 to 14.0 |  |
| Body-weight change (kg) | 48-56 wk | without T2D | Titration (wk) | 3 | 3 | [illegible] | [illegible] |  |
| Body-weight change (kg) | 48-56 wk | without T2D | Total randomized (n) | 3 | 3 | 729 | 696.0 to 729.0 |  |
| Body-weight change (kg) | 48-56 wk | with T2D | Background deficit (kcal) | 3 | 2 | 500 | 500.0 to 500.0 |  |
| Body-weight change (kg) | 48-56 wk | with T2D | Max follow-up (wk) | 3 | 2 | 56 | 56.0 to 56.0 |  |
| Body-weight change (kg) | 48-56 wk | with T2D | Mean age (y) | 3 | 2 | 55 | 55.0 to 56.5 |  |
| Body-weight change (kg) | 48-56 wk | with T2D | Mean BMI (kg/m2) | 3 | 2 | 37.1 | 36.5 to 37.1 |  |
| Body-weight change (kg) | 48-56 wk | with T2D | Mean weight (kg) | 3 | 2 | 105.7 | 103.2 to 105.7 |  |
| Body-weight change (kg) | 48-56 wk | with T2D | Female (%) | 3 | 2 | 48 | 47.5 to 48.0 |  |
| Body-weight change (kg) | 48-56 wk | with T2D | Planned contacts/y | 3 | 2 | 14 | 14.0 to 17.0 |  |
| Body-weight change (kg) | 48-56 wk | with T2D | Titration (wk) | 3 | 2 | 4 | 4.0 to 4.0 |  |
| Body-weight change (kg) | 48-56 wk | with T2D | Total randomized (n) | 3 | 2 | 846 | 621.0 to 846.0 |  |
| Body-weight change (kg) | 20-28 wk | without T2D | Max follow-up (wk) | 11 | 5 | 24 | 24.0 to 104.0 |  |
| Body-weight change (kg) | 20-28 wk | without T2D | Total randomized (n) | 11 | 5 | 161 | 80.0 to 845.0 |  |
| Body-weight change (kg) | 10-14 wk | without T2D | Max follow-up (wk) | 8 | 4 | 12 | 12.0 to 12.1 |  |
| Body-weight change (kg) | 10-14 wk | without T2D | Total randomized (n) | 8 | 4 | 188 | 54.0 to 188.0 |  |
| Waist circumference change (cm) | 48-56 wk | without T2D | Background deficit (kcal) | 3 | 3 | 500 | 500.0 to 600.0 |  |
| Waist circumference change (cm) | 48-56 wk | without T2D | Max follow-up (wk) | 3 | 3 | 52 | 48.0 to 56.0 |  |
| Waist circumference change (cm) | 48-56 wk | without T2D | Mean age (y) | 3 | 3 | 47 | 43.5 to 48.0 |  |
| Waist circumference change (cm) | 48-56 wk | without T2D | Mean BMI (kg/m2) | 3 | 3 | 37.4 | 36.0 to 39.0 |  |
| Waist circumference change (cm) | 48-56 wk | without T2D | Mean weight (kg) | 3 | 3 | 104.2 | 100.5 to 107.8 |  |
| Waist circumference change (cm) | 48-56 wk | without T2D | Female (%) | 3 | 3 | 80 | 74.0 to 81.5 |  |
| Waist circumference change (cm) | 48-56 wk | without T2D | Prediabetes (%) | 2 | 2 | 27.5 | 25.0 to 30.0 |  |
| Waist circumference change (cm) | 48-56 wk | without T2D | Planned contacts/y | 3 | 3 | 14 | 13.0 to 23.0 |  |
| Waist circumference change (cm) | 48-56 wk | without T2D | Titration (wk) | 3 | 3 | 4 | 0.0 to 4.0 |  |
| Waist circumference change (cm) | 48-56 wk | without T2D | Total randomized (n) | 3 | 3 | 391 | 282.0 to 531.0 |  |
| Waist circumference change (cm) | 48-56 wk | with T2D | Background deficit (kcal) | 3 | 2 | 500 | 500.0 to 500.0 |  |
| Waist circumference change (cm) | 48-56 wk | with T2D | Max follow-up (wk) | 3 | 2 | 56 | 56.0 to 56.0 |  |
| Waist circumference change (cm) | 48-56 wk | with T2D | Mean age (y) | 3 | 2 | 55 | 55.0 to 56.5 |  |
| Waist circumference change (cm) | 48-56 wk | with T2D | Mean BMI (kg/m2) | 3 | 2 | 37.1 | 36.5 to 37.1 |  |
| Waist circumference change (cm) | 48-56 wk | with T2D | Mean weight (kg) | 3 | 2 | 105.7 | 103.2 to 105.7 |  |
| Waist circumference change (cm) | 48-56 wk | with T2D | Female (%) | 3 | 2 | 48 | 47.5 to 48.0 |  |
| Waist circumference change (cm) | 48-56 wk | with T2D | Planned contacts/y | 3 | 2 | 14 | 14.0 to 17.0 |  |
| Waist circumference change (cm) | 48-56 wk | with T2D | Titration (wk) | 3 | 2 | 4 | 4.0 to 4.0 |  |
| Waist circumference change (cm) | 48-56 wk | with T2D | Total randomized (n) | 3 | 2 | 846 | 621.0 to 846.0 |  |
| Waist circumference change (cm) | 20-28 wk | without T2D | Max follow-up (wk) | 2 | 2 | 24 | 24.0 to 24.0 |  |
| Waist circumference change (cm) | 20-28 wk | without T2D | Total randomized (n) | 2 | 2 | 105.5 | 80.0 to 131.0 |  |
| >=5% weight reduction | 48-56 wk | without T2D | Background deficit (kcal) | 52 | 5 | 500 | 500.0 to 500.0 |  |
| >=5% weight reduction | 48-56 wk | without T2D | Max follow-up (wk) | 53 | 6 | 52 | 48.0 to 52.0 |  |
| >=5% weight reduction | 48-56 wk | without T2D | Mean age (y) | 52 | 5 | 46.5 | 35.5 to 46.6 |  |
| >=5% weight reduction | 48-56 wk | without T2D | Mean BMI (kg/m2) | 52 | 5 | 39.3 | 32.3 to 39.3 |  |
| >=5% weight reduction | 48-56 wk | without T2D | Mean weight (kg) | 52 | 5 | 111.3 | 91.0 to 111.3 |  |
| >=5% weight reduction | 48-56 wk | without T2D | Female (%) | 52 | 5 | 65 | 48.6 to 74.0 |  |
| >=5% weight reduction | 48-56 wk | without T2D | Prediabetes (%) | 16 | 4 | 30 | 28.0 to 100.0 |  |
| >=5% weight reduction | 48-56 wk | without T2D | Planned contacts/y | 52 | 5 | 17 | 13.0 to 17.0 |  |
| >=5% weight reduction | 48-56 wk | without T2D | Titration (wk) | 52 | 5 | 16 | 4.0 to 20.0 |  |
| >=5% weight reduction | 48-56 wk | without T2D | Total randomized (n) | 53 | 6 | 1162 | 150.0 to 1162.0 |  |
| >=5% weight reduction | 48-56 wk | with T2D | Background deficit (kcal) | 6 | 2 | 500 | 500.0 to 500.0 |  |
| >=5% weight reduction | 48-56 wk | with T2D | Max follow-up (wk) | 6 | 2 | 66 | 56.0 to 76.0 |  |
| >=5% weight reduction | 48-56 wk | with T2D | Mean age (y) | 6 | 2 | 55.6 | 55.0 to 56.2 |  |
| >=5% weight reduction | 48-56 wk | with T2D | Mean BMI (kg/m2) | 6 | 2 | 36.55 | 36.0 to 37.1 |  |
| >=5% weight reduction | 48-56 wk | with T2D | Mean weight (kg) | 6 | 2 | 103.75 | 101.8 to 105.7 |  |
| >=5% weight reduction | 48-56 wk | with T2D | Female (%) | 6 | 2 | 48.5 | 48.0 to 49.0 |  |
| >=5% weight reduction | 48-56 wk | with T2D | Planned contacts/y | 6 | 2 | 13.5 | 13.0 to 14.0 |  |
| >=5% weight reduction | 48-56 wk | with T2D | Titration (wk) | 6 | 2 | 14 | 4.0 to 24.0 |  |
| >=5% weight reduction | 48-56 wk | with T2D | Total randomized (n) | 6 | 2 | 799 | 752.0 to 846.0 |  |
| >=5% weight reduction | 48-56 wk | mixed population | Max follow-up (wk) | 8 | 3 | 52 | 52.0 to 52.0 |  |
| >=5% weight reduction | 48-56 wk | mixed population | Total randomized (n) | 8 | 3 | 190 | 190.0 to 1195.0 |  |
| >=5% weight reduction | 20-28 wk | without T2D | Max follow-up (wk) | 19 | 5 | 26 | 24.0 to 52.0 |  |
| >=5% weight reduction | 20-28 wk | without T2D | Total randomized (n) | 19 | 5 | 474 | 80.0 to 474.0 |  |
| >=5% weight reduction | 10-14 wk | without T2D | Max follow-up (wk) | 7 | 5 | 26 | 26.0 to 26.0 |  |
| >=5% weight reduction | 10-14 wk | without T2D | Total randomized (n) | 7 | 5 | 235 | 235.0 to 235.0 |  |
| >=10% weight reduction | 48-56 wk | without T2D | Background deficit (kcal) | 52 | 6 | 500 | 250.0 to 500.0 |  |
| >=10% weight reduction | 48-56 wk | without T2D | Max follow-up (wk) | 53 | 7 | 52 | 48.0 to 52.0 |  |
| >=10% weight reduction | 48-56 wk | without T2D | Mean age (y) | 52 | 6 | 46.5 | 35.5 to 58.0 |  |
| >=10% weight reduction | 48-56 wk | without T2D | Mean BMI (kg/m2) | 52 | 6 | 39.3 | 32.3 to 39.3 |  |
| >=10% weight reduction | 48-56 wk | without T2D | Mean weight (kg) | 52 | 6 | 111.3 | 91.0 to 111.3 |  |
| >=10% weight reduction | 48-56 wk | without T2D | Female (%) | 52 | 6 | 65 | 48.6 to 74.0 |  |
| >=10% weight reduction | 48-56 wk | without T2D | Prediabetes (%) | 16 | 4 | 30 | 28.0 to 100.0 |  |
| >=10% weight reduction | 48-56 wk | without T2D | Planned contacts/y | 52 | 6 | 17 | 11.5 to 17.0 |  |
| >=10% weight reduction | 48-56 wk | without T2D | Titration (wk) | 52 | 6 | 16 | 4.0 to 20.0 |  |
| >=10% weight reduction | 48-56 wk | without T2D | Total randomized (n) | 53 | 7 | 1162 | 150.0 to 1162.0 |  |
| >=10% weight reduction | 48-56 wk | with T2D | Background deficit (kcal) | 4 | 2 | 500 | 0.0 to 500.0 |  |
| >=10% weight reduction | 48-56 wk | with T2D | Max follow-up (wk) | 4 | 2 | 76 | 52.0 to 100.0 |  |
| >=10% weight reduction | 48-56 wk | with T2D | Mean age (y) | 4 | 2 | 56.2 | 56.2 to 69.0 |  |
| >=10% weight reduction | 48-56 wk | with T2D | Mean BMI (kg/m2) | 4 | 2 | 36 | 36.0 to 36.9 |  |
| >=10% weight reduction | 48-56 wk | with T2D | Mean weight (kg) | 4 | 2 | 101.8 | 101.8 to 102.7 |  |
| >=10% weight reduction | 48-56 wk | with T2D | Female (%) | 4 | 2 | 49 | 44.9 to 49.0 |  |
| >=10% weight reduction | 48-56 wk | with T2D | Planned contacts/y | 4 | 2 | 13 | 13.0 to 17.0 |  |
| >=10% weight reduction | 48-56 wk | with T2D | Titration (wk) | 4 | 2 | 24 | 16.0 to 24.0 |  |
| >=10% weight reduction | 48-56 wk | with T2D | Total randomized (n) | 4 | 2 | 752 | 616.0 to 752.0 |  |
| >=10% weight reduction | 48-56 wk | mixed population | Max follow-up (wk) | 2 | 2 | 52 | 52.0 to 52.0 |  |
| >=10% weight reduction | 48-56 wk | mixed population | Total randomized (n) | 2 | 2 | 963 | 731.0 to 1195.0 |  |
| >=10% weight reduction | 20-28 wk | without T2D | Max follow-up (wk) | 19 | 5 | 26 | 24.0 to 52.0 |  |
| >=10% weight reduction | 20-28 wk | without T2D | Total randomized (n) | 19 | 5 | 474 | 80.0 to 474.0 |  |
| >=10% weight reduction | 10-14 wk | without T2D | Max follow-up (wk) | 7 | 5 | 26 | 26.0 to 26.0 |  |
| >=10% weight reduction | 10-14 wk | without T2D | Total randomized (n) | 7 | 5 | 235 | 235.0 to 235.0 |  |
| >=15% weight reduction | 48-56 wk | without T2D | Background deficit (kcal) | 10 | 4 | 500 | 250.0 to 500.0 |  |
| >=15% weight reduction | 48-56 wk | without T2D | Max follow-up (wk) | 11 | 5 | 50 | 48.0 to 52.0 |  |
| >=15% weight reduction | 48-56 wk | without T2D | Mean age (y) | 10 | 4 | 35.5 | 35.5 to 58.0 |  |
| >=15% weight reduction | 48-56 wk | without T2D | Mean BMI (kg/m2) | 10 | 4 | 32.5 | 32.3 to 37.0 |  |
| >=15% weight reduction | 48-56 wk | without T2D | Mean weight (kg) | 10 | 4 | 91 | 91.0 to 102.8 |  |
| >=15% weight reduction | 48-56 wk | without T2D | Female (%) | 10 | 4 | 56 | 48.6 to 63.5 |  |
| >=15% weight reduction | 48-56 wk | without T2D | Prediabetes (%) | 10 | 3 | 28 | 28.0 to 100.0 |  |
| >=15% weight reduction | 48-56 wk | without T2D | Planned contacts/y | 10 | 4 | 13 | 11.5 to 13.0 |  |
| >=15% weight reduction | 48-56 wk | without T2D | Titration (wk) | 10 | 4 | 12 | 12.0 to 20.0 |  |
| >=15% weight reduction | 48-56 wk | without T2D | Total randomized (n) | 11 | 5 | 596.5 | 150.0 to 664.0 |  |
| >=15% weight reduction | 20-28 wk | without T2D | Max follow-up (wk) | 2 | 2 | 40 | 28.0 to 52.0 |  |
| >=15% weight reduction | 20-28 wk | without T2D | Total randomized (n) | 2 | 2 | 133 | 116.0 to 150.0 |  |
| >=20% weight reduction | 48-56 wk | without T2D | Background deficit (kcal) | 7 | 3 | 500 | 250.0 to 500.0 |  |
| >=20% weight reduction | 48-56 wk | without T2D | Max follow-up (wk) | 7 | 3 | 48 | 48.0 to 52.0 |  |
| >=20% weight reduction | 48-56 wk | without T2D | Mean age (y) | 7 | 3 | 35.5 | 35.5 to 58.0 |  |
| >=20% weight reduction | 48-56 wk | without T2D | Mean BMI (kg/m2) | 7 | 3 | 32.5 | 32.5 to 37.0 |  |
| >=20% weight reduction | 48-56 wk | without T2D | Mean weight (kg) | 7 | 3 | 91 | 91.0 to 102.8 |  |
| >=20% weight reduction | 48-56 wk | without T2D | Female (%) | 7 | 3 | 56 | 56.0 to 63.5 |  |
| >=20% weight reduction | 48-56 wk | without T2D | Prediabetes (%) | 7 | 2 | 28 | 28.0 to 100.0 |  |
| >=20% weight reduction | 48-56 wk | without T2D | Planned contacts/y | 7 | 3 | 13 | 11.5 to 13.0 |  |
| >=20% weight reduction | 48-56 wk | without T2D | Titration (wk) | 7 | 3 | 12 | 12.0 to 16.0 |  |
| >=20% weight reduction | 48-56 wk | without T2D | Total randomized (n) | 7 | 3 | 664 | 368.0 to 664.0 |  |
| All-cause permanent discontinuation | 48-56 wk | without T2D | Background deficit (kcal) | 87 | 14 | 500 | 250.0 to 600.0 | 0.11161 |
| All-cause permanent discontinuation | 48-56 wk | without T2D | Max follow-up (wk) | 87 | 14 | 52 | 48.0 to 104.0 | 0.50166 |
| All-cause permanent discontinuation | 48-56 wk | without T2D | Mean age (y) | 87 | 14 | 46.5 | 34.2 to 58.0 | 0.12844 |
| All-cause permanent discontinuation | 48-56 wk | without T2D | Mean BMI (kg/m2) | 87 | 14 | 37.8 | 31.1 to 40.1 | 0.1047 |
| All-cause permanent discontinuation | 48-56 wk | without T2D | Mean weight (kg) | 87 | 14 | 106.5 | 87.2 to 116.1 | 0.10046 |
| All-cause permanent discontinuation | 48-56 wk | without T2D | Female (%) | 87 | 14 | 65 | 30.3 to 83.0 | 0.15578 |
| All-cause permanent discontinuation | 48-56 wk | without T2D | Prediabetes (%) | 48 | 10 | 31 | 22.0 to 100.0 | 0.10247 |
| All-cause permanent discontinuation | 48-56 wk | without T2D | Planned contacts/y | 87 | 14 | 13 | 11.5 to 20.0 | 0.087443 |
| All-cause permanent discontinuation | 48-56 wk | without T2D | Titration (wk) | 87 | 14 | 12 | 0.0 to 24.0 | 0.084264 |
| All-cause permanent discontinuation | 48-56 wk | without T2D | Total randomized (n) | 87 | 14 | 506 | 150.0 to 1267.0 | 0.24402 |
| All-cause permanent discontinuation | 20-28 wk | without T2D | Max follow-up (wk) | 59 | 7 | 28 | 20.0 to 104.0 | 1 |
| All-cause permanent discontinuation | 20-28 wk | without T2D | Total randomized (n) | 59 | 7 | 756 | 61.0 to 845.0 | 0.43858 |
| AE-related permanent discontinuation | 48-56 wk | without T2D | Background deficit (kcal) | 56 | 11 | 500 | 250.0 to 500.0 | 0.2865 |
| AE-related permanent discontinuation | 48-56 wk | without T2D | Max follow-up (wk) | 106 | 17 | 52 | 48.0 to 104.0 | 0.081404 |
| AE-related permanent discontinuation | 48-56 wk | without T2D | Mean age (y) | 56 | 11 | 46.5 | 35.5 to 58.0 | 0.13979 |
| AE-related permanent discontinuation | 48-56 wk | without T2D | Mean BMI (kg/m2) | 56 | 11 | 39.3 | 32.3 to 39.5 | 0.13041 |
| AE-related permanent discontinuation | 48-56 wk | without T2D | Mean weight (kg) | 56 | 11 | 111.3 | 91.0 to 116.1 | 0.065397 |
| AE-related permanent discontinuation | 48-56 wk | without T2D | Female (%) | 56 | 11 | 65 | 30.3 to 83.0 | 0.10303 |
| AE-related permanent discontinuation | 48-56 wk | without T2D | Prediabetes (%) | 18 | 7 | 35 | 28.0 to 100.0 |  |
| AE-related permanent discontinuation | 48-56 wk | without T2D | Planned contacts/y | 56 | 11 | 17 | 11.5 to 17.0 | 0.078988 |
| AE-related permanent discontinuation | 48-56 wk | without T2D | Titration (wk) | 56 | 11 | 16 | 4.0 to 20.0 | 0.082085 |
| AE-related permanent discontinuation | 48-56 wk | without T2D | Total randomized (n) | 106 | 17 | 507 | 179.0 to 1742.0 | 0.11088 |
| AE-related permanent discontinuation | 48-56 wk | with T2D | Background deficit (kcal) | 10 | 4 | 500 | 0.0 to 500.0 |  |
| AE-related permanent discontinuation | 48-56 wk | with T2D | Max follow-up (wk) | 20 | 6 | 52 | 52.0 to 56.0 | 0.31731 |
| AE-related permanent discontinuation | 48-56 wk | with T2D | Mean age (y) | 10 | 4 | 46.2 | 46.2 to 69.0 |  |
| AE-related permanent discontinuation | 48-56 wk | with T2D | Mean BMI (kg/m2) | 10 | 4 | 37.8 | 36.5 to 37.8 |  |
| AE-related permanent discontinuation | 48-56 wk | with T2D | Mean weight (kg) | 10 | 4 | 106.5 | 102.7 to 106.5 |  |
| AE-related permanent discontinuation | 48-56 wk | with T2D | Female (%) | 10 | 4 | 83 | 44.3 to 83.0 |  |
| AE-related permanent discontinuation | 48-56 wk | with T2D | Prediabetes (%) | 6 | 1 | 35 | 35.0 to 35.0 |  |
| AE-related permanent discontinuation | 48-56 wk | with T2D | Planned contacts/y | 10 | 4 | 13 | 10.0 to 17.0 |  |
| AE-related permanent discontinuation | 48-56 wk | with T2D | Titration (wk) | 10 | 4 | 12 | 4.0 to 16.0 |  |
| AE-related permanent discontinuation | 48-56 wk | with T2D | Total randomized (n) | 20 | 6 | 834 | 127.0 to 846.0 | 0.68309 |
| AE-related permanent discontinuation | 48-56 wk | mixed population | Background deficit (kcal) | 1 | 1 | 0 | 0.0 to 0.0 |  |
| AE-related permanent discontinuation | 48-56 wk | mixed population | Max follow-up (wk) | 4 | 2 | 56 | 48.0 to 56.0 |  |
| AE-related permanent discontinuation | 48-56 wk | mixed population | Mean age (y) | 1 | 1 | 50.8 | 50.8 to 50.8 |  |
| AE-related permanent discontinuation | 48-56 wk | mixed population | Mean BMI (kg/m2) | 1 | 1 | 35.8 | 35.8 to 35.8 |  |
| AE-related permanent discontinuation | 48-56 wk | mixed population | Mean weight (kg) | 1 | 1 | 100.8 | 100.8 to 100.8 |  |
| AE-related permanent discontinuation | 48-56 wk | mixed population | Female (%) | 1 | 1 | 44 | 44.0 to 44.0 |  |
| AE-related permanent discontinuation | 48-56 wk | mixed population | Planned contacts/y | 1 | 1 | 12 | 12.0 to 12.0 |  |
| AE-related permanent discontinuation | 48-56 wk | mixed population | Titration (wk) | 1 | 1 | 24 | 24.0 to 24.0 |  |
| AE-related permanent discontinuation | 48-56 wk | mixed population | Total randomized (n) | 4 | 2 | 2486 | 218.0 to 2486.0 |  |
| AE-related permanent discontinuation | 20-28 wk | without T2D | Max follow-up (wk) | 117 | 12 | 26 | 20.0 to 104.0 | 0.5428 |
| AE-related permanent discontinuation | 20-28 wk | without T2D | Total randomized (n) | 117 | 12 | 499 | 32.0 to 845.0 | 0.5637 |
| Serious adverse events | 48-56 wk | without T2D | Background deficit (kcal) | 56 | 10 | 500 | 250.0 to 500.0 | 0.36788 |
| Serious adverse events | 48-56 wk | without T2D | Max follow-up (wk) | 106 | 15 | 52 | 48.0 to 104.0 | 0.17106 |
| Serious adverse events | 48-56 wk | without T2D | Mean age (y) | 56 | 10 | 46.5 | 35.5 to 58.0 | 0.17124 |
| Serious adverse events | 48-56 wk | without T2D | Mean BMI (kg/m2) | 56 | 10 | 39.3 | 32.3 to 42.1 | 0.16232 |
| Serious adverse events | 48-56 wk | without T2D | Mean weight (kg) | 56 | 10 | 111.3 | 91.0 to 116.1 | 0.095089 |
| Serious adverse events | 48-56 wk | without T2D | Female (%) | 56 | 10 | 65 | 30.3 to 83.0 | 0.14782 |
| Serious adverse events | 48-56 wk | without T2D | Prediabetes (%) | 18 | 7 | 35 | 28.0 to 100.0 |  |
| Serious adverse events | 48-56 wk | without T2D | Planned contacts/y | 56 | 10 | 17 | 11.5 to 17.0 | 0.12285 |
| Serious adverse events | 48-56 wk | without T2D | Titration (wk) | 56 | 10 | 16 | 4.0 to 20.0 | 0.11456 |
| Serious adverse events | 48-56 wk | without T2D | Total randomized (n) | 106 | 15 | 507 | 179.0 to 2446.5 | 0.10633 |
| Serious adverse events | 48-56 wk | with T2D | Background deficit (kcal) | 10 | 3 | 500 | 0.0 to 500.0 |  |
| Serious adverse events | 48-56 wk | with T2D | Max follow-up (wk) | 20 | 5 | 54 | 52.0 to 56.0 |  |
| Serious adverse events | 48-56 wk | with T2D | Mean age (y) | 10 | 3 | 46.2 | 46.2 to 69.0 |  |
| Serious adverse events | 48-56 wk | with T2D | Mean BMI (kg/m2) | 10 | 3 | 37.8 | 36.9 to 37.8 |  |
| Serious adverse events | 48-56 wk | with T2D | Mean weight (kg) | 10 | 3 | 106.5 | 102.7 to 106.5 |  |
| Serious adverse events | 48-56 wk | with T2D | Female (%) | 10 | 3 | 63 | 44.3 to 63.0 |  |
| Serious adverse events | 48-56 wk | with T2D | Prediabetes (%) | 6 | 1 | 35 | 35.0 to 35.0 |  |
| Serious adverse events | 48-56 wk | with T2D | Planned contacts/y | 10 | 3 | 13 | 10.0 to 14.0 |  |
| Serious adverse events | 48-56 wk | with T2D | Titration (wk) | 10 | 3 | 12 | 4.0 to 12.0 |  |
| Serious adverse events | 48-56 wk | with T2D | Total randomized (n) | 20 | 5 | 834 | 127.0 to 846.0 |  |
| Serious adverse events | 48-56 wk | mixed population | Background deficit (kcal) | 1 | 1 | 0 | 0.0 to 0.0 |  |
| Serious adverse events | 48-56 wk | mixed population | Max follow-up (wk) | 5 | 3 | 56 | 48.0 to 56.0 |  |
| Serious adverse events | 48-56 wk | mixed population | Mean age (y) | 1 | 1 | 50.8 | 50.8 to 50.8 |  |
| Serious adverse events | 48-56 wk | mixed population | Mean BMI (kg/m2) | 1 | 1 | 35.8 | 35.8 to 35.8 |  |
| Serious adverse events | 48-56 wk | mixed population | Mean weight (kg) | 1 | 1 | 100.8 | 100.8 to 100.8 |  |
| Serious adverse events | 48-56 wk | mixed population | Female (%) | 1 | 1 | 44 | 44.0 to 44.0 |  |
| Serious adverse events | 48-56 wk | mixed population | Planned contacts/y | 1 | 1 | 12 | 12.0 to 12.0 |  |
| Serious adverse events | 48-56 wk | mixed population | Titration (wk) | 1 | 1 | 24 | 24.0 to 24.0 |  |
| Serious adverse events | 48-56 wk | mixed population | Total randomized (n) | 5 | 3 | 2486 | 2486.0 to 2486.0 |  |
| Serious adverse events | 20-28 wk | without T2D | Max follow-up (wk) | 133 | 10 | 26 | 20.0 to 104.0 | 0.5428 |
| Serious adverse events | 20-28 wk | without T2D | Total randomized (n) | 133 | 10 | 499 | 72.0 to 845.0 | 0.5637 |
| Nausea | 48-56 wk | without T2D | Background deficit (kcal) | 20 | 7 | 500 | 500.0 to 500.0 |  |
| Nausea | 48-56 wk | without T2D | Max follow-up (wk) | 23 | 10 | 52 | 48.0 to 160.0 | 0.0455 |
| Nausea | 48-56 wk | without T2D | Mean age (y) | 20 | 7 | 42.6 | 35.5 to 49.5 |  |
| Nausea | 48-56 wk | without T2D | Mean BMI (kg/m2) | 20 | 7 | 37.8 | 32.3 to 42.1 |  |
| Nausea | 48-56 wk | without T2D | Mean weight (kg) | 20 | 7 | 106.5 | 91.0 to 116.1 |  |
| Nausea | 48-56 wk | without T2D | Female (%) | 20 | 7 | 56 | 30.3 to 80.0 |  |
| Nausea | 48-56 wk | without T2D | Prediabetes (%) | 17 | 6 | 32.75 | 28.0 to 61.2 |  |
| Nausea | 48-56 wk | without T2D | Planned contacts/y | 20 | 7 | 13 | 13.0 to 14.0 |  |
| Nausea | 48-56 wk | without T2D | Titration (wk) | 20 | 7 | 12 | 4.0 to 20.0 |  |
| Nausea | 48-56 wk | without T2D | Total randomized (n) | 23 | 10 | 664 | 210.0 to 3731.0 | 0.083265 |
| Nausea | 48-56 wk | with T2D | Background deficit (kcal) | 4 | 3 | 500 | 500.0 to 500.0 |  |
| Nausea | 48-56 wk | with T2D | Max follow-up (wk) | 4 | 2 | 56 | 56.0 to 56.0 |  |
| Nausea | 48-56 wk | with T2D | Mean age (y) | 4 | 2 | 55 | 55.0 to 55.0 |  |
| Nausea | 48-56 wk | with T2D | Mean BMI (kg/m2) | 4 | 2 | 37.1 | 37.1 to 37.1 |  |
| Nausea | 48-56 wk | with T2D | Mean weight (kg) | 4 | 2 | 105.7 | 105.7 to 105.7 |  |
| Nausea | 48-56 wk | with T2D | Female (%) | 4 | 2 | 48 | 48.0 to 48.0 |  |
| Nausea | 48-56 wk | with T2D | Planned contacts/y | 4 | 2 | 14 | 14.0 to 14.0 |  |
| Nausea | 48-56 wk | with T2D | Titration (wk) | 4 | 2 | 4 | 4.0 to 4.0 |  |
| Nausea | 48-56 wk | with T2D | Total randomized (n) | 4 | 2 | 846 | 505.0 to 846.0 |  |
| Nausea | 20-28 wk | without T2D | Max follow-up (wk) | 67 | 6 | 28 | 20.0 to 104.0 |  |
| Nausea | 20-28 wk | without T2D | Total randomized (n) | 67 | 6 | 756 | 61.0 to 845.0 |  |
| Nausea | 10-14 wk | without T2D | Max follow-up (wk) | 3 | 3 | 12 | 12.0 to 14.0 |  |
| Nausea | 10-14 wk | without T2D | Total randomized (n) | 3 | 3 | 44 | 32.0 to 68.0 |  |
| Vomiting | 48-56 wk | without T2D | Background deficit (kcal) | 15 | 6 | 500 | 500.0 to 500.0 |  |
| Vomiting | 48-56 wk | without T2D | Max follow-up (wk) | 18 | 9 | 52 | 48.0 to 160.0 |  |
| Vomiting | 48-56 wk | without T2D | Mean age (y) | 15 | 6 | 36.1 | 35.5 to 46.2 |  |
| Vomiting | 48-56 wk | without T2D | Mean BMI (kg/m2) | 15 | 6 | 32.5 | 32.3 to 37.8 |  |
| Vomiting | 48-56 wk | without T2D | Mean weight (kg) | 15 | 6 | 91.8 | 91.0 to 106.5 |  |
| Vomiting | 48-56 wk | without T2D | Female (%) | 15 | 6 | 56 | 48.6 to 63.5 |  |
| Vomiting | 48-56 wk | without T2D | Prediabetes (%) | 15 | 6 | 30.5 | 28.0 to 35.0 |  |
| Vomiting | 48-56 wk | without T2D | Planned contacts/y | 15 | 6 | 13 | 13.0 to 13.0 |  |
| Vomiting | 48-56 wk | without T2D | Titration (wk) | 15 | 6 | 12 | 12.0 to 20.0 |  |
| Vomiting | 48-56 wk | without T2D | Total randomized (n) | 18 | 9 | 664 | 210.0 to 1742.0 |  |
| Vomiting | 48-56 wk | with T2D | Background deficit (kcal) | 4 | 2 | 500 | 500.0 to 500.0 |  |
| Vomiting | 48-56 wk | with T2D | Max follow-up (wk) | 4 | 2 | 56 | 56.0 to 56.0 |  |
| Vomiting | 48-56 wk | with T2D | Mean age (y) | 4 | 2 | 55 | 55.0 to 55.0 |  |
| Vomiting | 48-56 wk | with T2D | Mean BMI (kg/m2) | 4 | 2 | 37.1 | 37.1 to 37.1 |  |
| Vomiting | 48-56 wk | with T2D | Mean weight (kg) | 4 | 2 | 105.7 | 105.7 to 105.7 |  |
| Vomiting | 48-56 wk | with T2D | Female (%) | 4 | 2 | 48 | 48.0 to 48.0 |  |
| Vomiting | 48-56 wk | with T2D | Planned contacts/y | 4 | 2 | 14 | 14.0 to 14.0 |  |
| Vomiting | 48-56 wk | with T2D | Titration (wk) | 4 | 2 | 4 | 4.0 to 4.0 |  |
| Vomiting | 48-56 wk | with T2D | Total randomized (n) | 4 | 2 | 846 | 505.0 to 846.0 |  |
| Vomiting | 20-28 wk | without T2D | Max follow-up (wk) | 45 | 4 | 26 | 20.0 to 104.0 |  |
| Vomiting | 20-28 wk | without T2D | Total randomized (n) | 45 | 4 | 706 | 61.0 to 845.0 |  |
| Diarrhoea | 48-56 wk | without T2D | Background deficit (kcal) | 14 | 6 | 500 | 500.0 to 500.0 |  |
| Diarrhoea | 48-56 wk | without T2D | Max follow-up (wk) | 14 | 6 | 52 | 48.0 to 104.0 |  |
| Diarrhoea | 48-56 wk | without T2D | Mean age (y) | 14 | 6 | 36.1 | 35.5 to 49.5 |  |
| Diarrhoea | 48-56 wk | without T2D | Mean BMI (kg/m2) | 14 | 6 | 32.5 | 32.3 to 42.1 |  |
| Diarrhoea | 48-56 wk | without T2D | Mean weight (kg) | 14 | 6 | 91.8 | 91.0 to 116.1 |  |
| Diarrhoea | 48-56 wk | without T2D | Female (%) | 14 | 6 | 56 | 30.3 to 63.5 |  |
| Diarrhoea | 48-56 wk | without T2D | Prediabetes (%) | 11 | 5 | 29.25 | 28.0 to 61.2 |  |
| Diarrhoea | 48-56 wk | without T2D | Planned contacts/y | 14 | 6 | 13 | 13.0 to 14.0 |  |
| Diarrhoea | 48-56 wk | without T2D | Titration (wk) | 14 | 6 | 12 | 4.0 to 20.0 |  |
| Diarrhoea | 48-56 wk | without T2D | Total randomized (n) | 14 | 6 | 664 | 210.0 to 3731.0 |  |
| Diarrhoea | 48-56 wk | with T2D | Background deficit (kcal) | 3 | 2 | 500 | 500.0 to 500.0 |  |
| Diarrhoea | 48-56 wk | with T2D | Max follow-up (wk) | 3 | 2 | 56 | 56.0 to 56.0 |  |
| Diarrhoea | 48-56 wk | with T2D | Mean age (y) | 3 | 2 | 55 | 55.0 to 55.0 |  |
| Diarrhoea | 48-56 wk | with T2D | Mean BMI (kg/m2) | 3 | 2 | 37.1 | 37.1 to 37.1 |  |
| Diarrhoea | 48-56 wk | with T2D | Mean weight (kg) | 3 | 2 | 105.7 | 105.7 to 105.7 |  |
| Diarrhoea | 48-56 wk | with T2D | Female (%) | 3 | 2 | 48 | 48.0 to 48.0 |  |
| Diarrhoea | 48-56 wk | with T2D | Planned contacts/y | 3 | 2 | 14 | 14.0 to 14.0 |  |
| Diarrhoea | 48-56 wk | with T2D | Titration (wk) | 3 | 2 | 4 | 4.0 to 4.0 |  |
| Diarrhoea | 48-56 wk | with T2D | Total randomized (n) | 4 | 2 | 846 | 505.0 to 846.0 |  |
| Diarrhoea | 20-28 wk | without T2D | Max follow-up (wk) | 37 | 4 | 26 | 20.0 to 104.0 |  |
| Diarrhoea | 20-28 wk | without T2D | Total randomized (n) | 37 | 4 | 706 | 61.0 to 845.0 |  |
| Constipation | 48-56 wk | without T2D | Background deficit (kcal) | 3 | 3 | 500 | 500.0 to 500.0 |  |
| Constipation | 48-56 wk | without T2D | Max follow-up (wk) | 6 | 4 | 56 | 56.0 to 56.0 |  |
| Constipation | 48-56 wk | without T2D | Mean age (y) | 3 | 3 | 42.6 | 42.6 to 42.6 |  |
| Constipation | 48-56 wk | without T2D | Mean BMI (kg/m2) | 3 | 3 | 42.1 | 42.1 to 42.1 |  |
| Constipation | 48-56 wk | without T2D | Mean weight (kg) | 3 | 3 | 116.1 | 116.1 to 116.1 |  |
| Constipation | 48-56 wk | without T2D | Female (%) | 3 | 3 | 83 | 83.0 to 83.0 |  |
| Constipation | 48-56 wk | without T2D | Planned contacts/y | 3 | 3 | 13 | 13.0 to 13.0 |  |
| Constipation | 48-56 wk | without T2D | Titration (wk) | 3 | 3 | 4 | 4.0 to 4.0 |  |
| Constipation | 48-56 wk | without T2D | Total randomized (n) | 6 | 4 | 1381.5 | 1267.0 to 1742.0 |  |
| Constipation | 48-56 wk | with T2D | Background deficit (kcal) | 4 | 2 | 500 | 500.0 to 500.0 |  |
| Constipation | 48-56 wk | with T2D | Max follow-up (wk) | 4 | 2 | 56 | 56.0 to 56.0 |  |
| Constipation | 48-56 wk | with T2D | Mean age (y) | 4 | 2 | 55 | 55.0 to 55.0 |  |
| Constipation | 48-56 wk | with T2D | Mean BMI (kg/m2) | 4 | 2 | 37.1 | 37.1 to 37.1 |  |
| Constipation | 48-56 wk | with T2D | Mean weight (kg) | 4 | 2 | 105.7 | 105.7 to 105.7 |  |
| Constipation | 48-56 wk | with T2D | Female (%) | 4 | 2 | 48 | 48.0 to 48.0 |  |
| Constipation | 48-56 wk | with T2D | Planned contacts/y | 4 | 2 | 14 | 14.0 to 14.0 |  |
| Constipation | 48-56 wk | with T2D | Titration (wk) | 4 | 2 | 4 | 4.0 to 4.0 |  |
| Constipation | 48-56 wk | with T2D | Total randomized (n) | 4 | 2 | 846 | 505.0 to 846.0 |  |
| Constipation | 20-28 wk | without T2D | Max follow-up (wk) | 42 | 3 | 28 | 26.0 to 28.0 |  |
| Constipation | 20-28 wk | without T2D | Total randomized (n) | 42 | 3 | 706 | 400.5 to 756.0 |  |

**Table S11.** League table for the primary outcome (selected interventions) Cells below the diagonal: MD (95% Cl) in% body-weight change for the row intervention versus the column intervention (negative favours the row). Full league table in nma_league_table_main.csv.

| **Placebo** | **Sema SC 2.4mg QW** | **Lira SC 3mg QD** | **TZP SC 5mg QW** | **TZP SC 10mg QW** | **TZP SC 15mg QW** | **Orli PO 120mg TID** | **Orli PO 60mg TID** | **TZP SC MTD 10-15mg QW** | **Sema SC 7.2mg QW** | **CagriSema SC 2.4/2.4mg QW** | **Reta SC 12mg QW** |
|---|---|---|---|---|---|---|---|---|---|---|---|
| Placebo |  |  |  |  |  |  |  |  |  |  |  |
| -11.49<br>(-12.23, -10.75) | Sema SC 2.4mg QW |  |  |  |  |  |  |  |  |  |  |
| -4.07<br>(-5.46, -2.68) | 7.42<br>(5.91, 8.94) | Lira SC 3mg QD |  |  |  |  |  |  |  |  |  |
| -11.16<br>(-12.76, -9.56) | 0.33<br>(-1.38, 2.03) | -7.10<br>(-9.21, -4.98) | TZP SC 5mg QW |  |  |  |  |  |  |  |  |
| -14.89<br>(-16.36, -13.43) | -3.40<br>(-4.99, -1.82) | -10.83<br>(-12.84, -8.81) | -3.73<br>(-5.40, -2.06) | TZP SC 10mg QW |  |  |  |  |  |  |  |
| -16.90<br>(-18.15, -15.64) | -5.41<br>(-6.73, -4.09) | -12.83<br>(-14.69, -10.97) | -5.73<br>(-7.34, -4.12) | -2.00<br>(-3.50, -0.51) | TZP SC 15mg QW |  |  |  |  |  |  |
| -3.13<br>(-4.23, -2.03) | 8.36<br>(7.04, 9.68) | 0.94<br>(-0.83, 2.71) | 8.03<br>(6.09, 9.97) | 11.76<br>(9.93, 13.60) | 13.77<br>(12.10, 15.44) | Orli PO 120mg TID |  |  |  |  |  |
| -2.02<br>(-3.67, -0.36) | 9.48<br>(7.66, 11.29) | 2.05<br>(-0.11, 4.21) | 9.15<br>(6.85, 11.45) | 12.88<br>(10.67, 15.09) | 14.88<br>(12.80, 16.96) | 1.11<br>(-0.53, 2.76) | Orli PO 60mg TID |  |  |  |  |
| -16.66<br>(-18.29, -15.02) | -5.17<br>(-6.96, -3.37) | -12.59<br>(-14.74, -10.44) | -5.49<br>(-7.78, -3.21) | -1.76<br>(-3.96, 0.43) | 0.24<br>(-1.82, 2.30) | -13.53<br>(-15.50, -11.56) | -14.64<br>(-16.97, -12.32) | TZP SC MTD 10-15mg QW |  |  |  |
| -14.34<br>(-16.03, -12.65) | -2.85<br>(-4.56, -1.14) | -10.27<br>(-12.44, -8.10) | -3.18<br>(-5.49, -0.87) | 0.55<br>(-1.67, 2.77) | 2.56<br>(0.49, 4.62) | -11.21<br>(-13.22, -9.20) | -12.32<br>(-14.69, -9.96) | 2.32<br>(-0.03, 4.67) | Sema SC 7.2mg QW |  |  |
| -16.40<br>(-17.95, -14.85) | -4.91<br>(-6.62, -3.19) | -12.33<br>(-14.41, -10.25) | -5.24<br>(-7.46, -3.01) | -1.51<br>(-3.64, 0.63) | 0.50<br>(-1.50, 2.49) | -13.27<br>(-15.17, -11.37) | -14.38<br>(-16.65, -12.12) | 0.26<br>(-1.99, 2.51) | -2.06<br>(-4.35, 0.23) | CagriSema SC 2.4/2.4mg QW |  |
| -20.00<br>(-21.93, -18.07) | -8.51<br>(-10.57, -6.45) | -15.93<br>(-18.31, -13.56) | -8.84<br>(-11.34, -6.33) | -5.11<br>(-7.53, -2.68) | -3.10<br>(-5.41, -0.80) | -16.87<br>(-19.09, -14.65) | -17.98<br>(-20.52, -15.44) | -3.34<br>(-5.87, -0.81) | -5.66<br>(-8.22, -3.10) | -3.60<br>(-6.07, -1.13) | Reta SC 12mg QW |

**Table S12.**
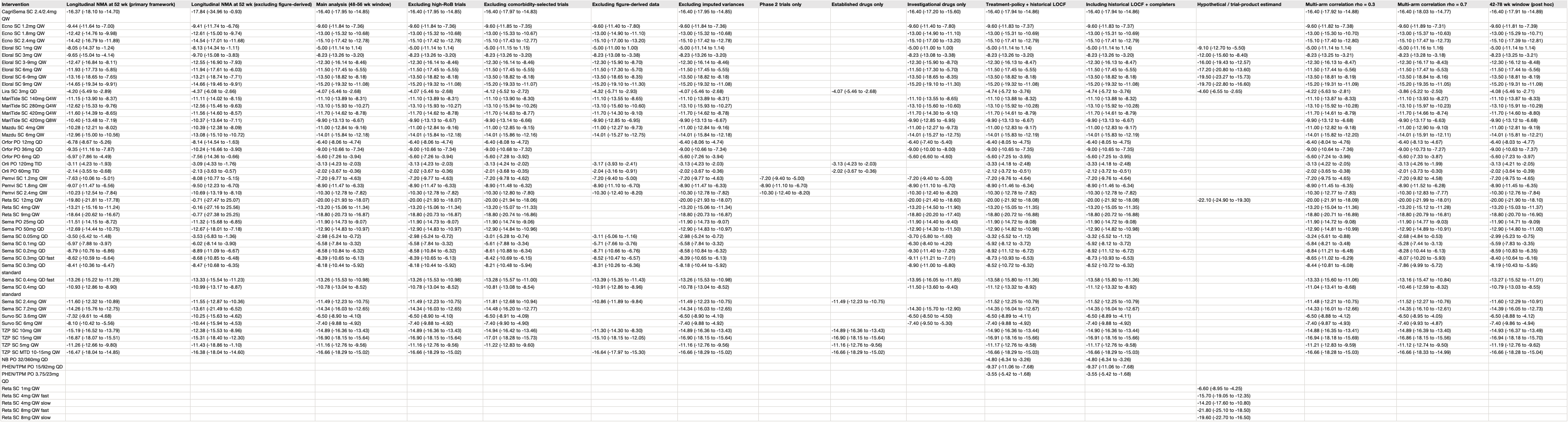
Sensitivity and estimand analyses of the primary outcome. Includes the primary longitudinal model at 52 weeks (with and without figure-derived records), the 48-56 week reference window NMA. exclusion of comorbidity-selected trials (HFpEF, OSA. MASH, OA), and alternative estimands (including historical LOCF and completer analyses for PHEN/TPM CR 3.75123 mg and 15/92 mg from EQUIP and NB 32/360 mg).

| Intervention | Longitudinal NMA at 52 wk (primary framework) | Longitudinal NMA at 52 wk (excluding figure-derived) | Main analysis (48-56 wk window) | Excluding high-RoB trials | Excluding comorbidity-selected trials | Excluding figure-derived data | Excluding imputed variances | Phase 2 trials only | Established drugs only | Investigational drugs only | Treatment-policy + historical LOCF | Including historical LOCF + completers | Hypothetical / trial-product estimand | Multi-arm correlation rho = 0.3 | Multi-arm correlation rho = 0.7 | 42-78 wk window (post hoc) |
|---|---|---|---|---|---|---|---|---|---|---|---|---|---|---|---|---|
| CagriSema SC 2.4/2.4mg QW | -16.37 (-18.10 to -14.70) | -17.84 (-34.96 to -0.93) | -16.40 (-17.95 to -14.85) | -16.40 (-17.95 to -14.85) | -16.40 (-17.97 to -14.83) |  | -16.40 (-17.95 to -14.85) |  |  | -16.40 (-17.20 to -15.60) | -16.40 (-17.94 to -14.86) | -16.40 (-17.94 to -14.86) |  | -16.40 (-17.92 to -14.88) | -16.40 (-18.03 to -14.77) | -16.40 (-17.91 to -14.89) |
| Ecno SC 1.2mg QW | -9.44 (-11.64 to -7.00) | -9.41 (-11.74 to -6.76) | -9.60 (-11.84 to -7.36) | -9.60 (-11.84 to -7.36) | -9.60 (-11.84 to -7.35) | -9.60 (-11.40 to -7.80) | -9.60 (-11.84 to -7.36) |  |  | -9.60 (-11.40 to -7.80) | -9.60 (-11.83 to -7.37) | -9.60 (-11.83 to -7.37) |  | -9.60 (-11.82 to -7.38) | -9.60 (-11.89 to -7.31) | -9.60 (-11.81 to -7.39) |
| Ecno SC 1.8mg QW | -12.42 (-14.76 to -9.98) | -12.61 (-15.01 to -9.74) | -13.00 (-15.32 to -10.68) | -13.00 (-15.32 to -10.68) | -13.00 (-15.33 to -10.67) | -13.00 (-14.90 to -11.10) | -13.00 (-15.32 to -10.68) |  |  | -13.00 (-14.90 to -11.10) | -13.00 (-15.31 to -10.69) | -13.00 (-15.31 to -10.69) |  | -13.00 (-15.30 to -10.70) | -13.00 (-15.37 to -10.63) | -13.00 (-15.29 to -10.71) |
| Ecno SC 2.4mg QW | -14.42 (-16.79 to -11.89) | -14.54 (-17.01 to -11.68) | -15.10 (-17.42 to -12.78) | -15.10 (-17.42 to -12.78) | -15.10 (-17.43 to -12.77) | -15.10 (-17.00 to -13.20) | -15.10 (-17.42 to -12.78) |  |  | -15.10 (-17.00 to -13.20) | -15.10 (-17.41 to -12.79) | -15.10 (-17.41 to -12.79) |  | -15.10 (-17.40 to -12.80) | -15.10 (-17.47 to -12.73) | -15.10 (-17.39 to -12.81) |
| Eloral SC 1mg QW | -8.05 (-14.37 to -1.24) | -8.13 (-14.34 to -1.11) | -5.00 (-11.14 to 1.14) | -5.00 (-11.14 to 1.14) | -5.00 (-11.15 to 1.15) | -5.00 (-11.00 to 1.00) | -5.00 (-11.14 to 1.14) |  |  | -5.00 (-11.00 to 1.00) | -5.00 (-11.14 to 1.14) | -5.00 (-11.14 to 1.14) | -9.10 (-12.70 to -5.50) | -5.00 (-11.14 to 1.14) | -5.00 (-11.16 to 1.16) | -5.00 (-11.14 to 1.14) |
| Eloral SC 3mg QW | -9.65 (-15.04 to -4.14) | -9.70 (-15.08 to -3.83) | -8.23 (-13.26 to -3.20) | -8.23 (-13.26 to -3.20) | -8.23 (-13.26 to -3.20) | -8.23 (-13.08 to -3.38) | -8.23 (-13.26 to -3.20) |  |  | -8.23 (-13.08 to -3.38) | -8.23 (-13.26 to -3.20) | -8.23 (-13.26 to -3.20) | -12.00 (-15.60 to -8.40) | -8.23 (-13.25 to -3.21) | -8.23 (-13.28 to -3.18) | -8.23 (-13.25 to -3.21) |
| Eloral SC 3-9mg QW | -12.47 (-16.84 to -8.11) | -12.55 (-16.90 to -7.93) | -12.30 (-16.14 to -8.46) | -12.30 (-16.14 to -8.46) | -12.30 (-16.14 to -8.46) | -12.30 (-15.90 to -8.70) | -12.30 (-16.14 to -8.46) |  |  | -12.30 (-15.90 to -8.70) | -12.30 (-16.13 to -8.47) | -12.30 (-16.13 to -8.47) | -16.00 (-19.43 to -12.57) | -12.30 (-16.13 to -8.47) | -12.30 (-16.17 to -8.43) | -12.30 (-16.12 to -8.48) |
| Eloral SC 6mg QW | -11.93 (-17.73 to -5.85) | -11.94 (-17.61 to -6.03) | -11.50 (-17.45 to -5.55) | -11.50 (-17.45 to -5.55) | -11.50 (-17.45 to -5.55) | -11.50 (-17.30 to -5.70) | -11.50 (-17.45 to -5.55) |  |  | -11.50 (-17.30 to -5.70) | -11.50 (-17.45 to -5.55) | -11.50 (-17.45 to -5.55) | -17.20 (-20.80 to -13.60) | -11.50 (-17.44 to -5.56) | -11.50 (-17.47 to -5.53) | -11.50 (-17.44 to -5.56) |
| Eloral SC 6-9mg QW | -13.16 (-18.65 to -7.65) | -13.21 (-18.74 to -7.71) | -13.50 (-18.82 to -8.18) | -13.50 (-18.82 to -8.18) | -13.50 (-18.82 to -8.18) | -13.50 (-18.65 to -8.35) | -13.50 (-18.82 to -8.18) |  |  | -13.50 (-18.65 to -8.35) | -13.50 (-18.82 to -8.18) | -13.50 (-18.82 to -8.18) | -19.50 (-23.27 to -15.73) | -13.50 (-18.81 to -8.19) | -13.50 (-18.84 to -8.16) | -13.50 (-18.81 to -8.19) |
| Eloral SC 9mg QW | -14.65 (-19.34 to -9.91) | -14.66 (-19.46 to -9.91) | -15.20 (-19.32 to -11.08) | -15.20 (-19.32 to -11.08) | -15.20 (-19.33 to -11.07) | -15.20 (-19.10 to -11.30) | -15.20 (-19.32 to -11.08) |  |  | -15.20 (-19.10 to -11.30) | -15.20 (-19.32 to -11.08) | -15.20 (-19.32 to -11.08) | -19.70 (-22.80 to -16.60) | -15.20 (-19.31 to -11.09) | -15.20 (-19.35 to -11.05) | -15.20 (-19.31 to -11.09) |
| Lira SC 3mg QD | -4.20 (-5.49 to -2.89) | -4.37 (-6.03 to -2.66) | -4.07 (-5.46 to -2.68) | -4.07 (-5.46 to -2.68) | -4.12 (-5.52 to -2.72) | -4.32 (-5.71 to -2.93) | -4.07 (-5.46 to -2.68) |  | -4.07 (-5.46 to -2.68) |  | -4.74 (-5.72 to -3.76) | -4.74 (-5.72 to -3.76) | -4.60 (-6.55 to -2.65) | -4.22 (-5.63 to -2.81) | -3.86 (-5.22 to -2.50) | -4.08 (-5.46 to -2.71) |
| MariTide SC 140mg Q4W | -11.15 (-13.90 to -8.37) | -11.11 (-14.02 to -8.15) | -11.10 (-13.89 to -8.31) | -11.10 (-13.89 to -8.31) | -11.10 (-13.90 to -8.30) | -11.10 (-13.55 to -8.65) | -11.10 (-13.89 to -8.31) |  |  | -11.10 (-13.55 to -8.65) | -11.10 (-13.88 to -8.32) | -11.10 (-13.88 to -8.32) |  | -11.10 (-13.87 to -8.33) | -11.10 (-13.93 to -8.27) | -11.10 (-13.87 to -8.33) |
| MariTide SC 280mg Q4W | -12.62 (-15.33 to -9.76) | -12.82 (-15.46 to -9.63) | -13.10 (-15.93 to -10.27) | -13.10 (-15.93 to -10.27) | -13.10 (-15.94 to -10.26) | -13.10 (-15.60 to -10.60) | -13.10 (-15.93 to -10.27) |  |  | -13.10 (-15.60 to -10.60) | -13.10 (-15.92 to -10.28) | -13.10 (-15.92 to -10.28) |  | -13.10 (-15.92 to -10.28) | -13.10 (-15.97 to -10.23) | -13.10 (-15.91 to -10.29) |
| MariTide SC 420mg Q4W | -11.60 (-14.39 to -8.65) | -11.56 (-14.60 to -8.57) | -11.70 (-14.62 to -8.78) | -11.70 (-14.62 to -8.78) | -11.70 (-14.63 to -8.77) | -11.70 (-14.30 to -9.10) | -11.70 (-14.62 to -8.78) |  |  | -11.70 (-14.30 to -9.10) | -11.70 (-14.61 to -8.79) | -11.70 (-14.61 to -8.79) |  | -11.70 (-14.61 to -8.79) | -11.70 (-14.66 to -8.74) | -11.70 (-14.60 to -8.80) |
| MariTide SC 420mg Q8W | -10.40 (-13.48 to -7.19) | -10.37 (-13.64 to -7.11) | -9.90 (-13.13 to -6.67) | -9.90 (-13.13 to -6.67) | -9.90 (-13.14 to -6.66) | -9.90 (-12.85 to -6.95) | -9.90 (-13.13 to -6.67) |  |  | -9.90 (-12.85 to -6.95) | -9.90 (-13.13 to -6.67) | -9.90 (-13.13 to -6.67) |  | -9.90 (-13.12 to -6.68) | -9.90 (-13.17 to -6.63) | -9.90 (-13.12 to -6.68) |
| Mazdu SC 4mg QW | -10.28 (-12.21 to -8.02) | -10.39 (-12.38 to -8.09) | -11.00 (-12.84 to -9.16) | -11.00 (-12.84 to -9.16) | -11.00 (-12.85 to -9.15) | -11.00 (-12.27 to -9.73) | -11.00 (-12.84 to -9.16) |  |  | -11.00 (-12.27 to -9.73) | -11.00 (-12.83 to -9.17) | -11.00 (-12.83 to -9.17) |  | -11.00 (-12.82 to -9.18) | -11.00 (-12.90 to -9.10) | -11.00 (-12.81 to -9.19) |
| Mazdu SC 6mg QW | -12.96 (-15.00 to -10.56) | -13.08 (-15.10 to -10.72) | -14.01 (-15.84 to -12.18) | -14.01 (-15.84 to -12.18) | -14.01 (-15.86 to -12.16) | -14.01 (-15.27 to -12.75) | -14.01 (-15.84 to -12.18) |  |  | -14.01 (-15.27 to -12.75) | -14.01 (-15.83 to -12.19) | -14.01 (-15.83 to -12.19) |  | -14.01 (-15.82 to -12.20) | -14.01 (-15.91 to -12.11) | -14.01 (-15.81 to -12.21) |
| Orfor PO 12mg QD | -6.78 (-8.67 to -5.26) | -8.14 (-14.54 to -1.63) | -6.40 (-8.06 to -4.74) | -6.40 (-8.06 to -4.74) | -6.40 (-8.08 to -4.72) |  | -6.40 (-8.06 to -4.74) |  |  | -6.40 (-7.40 to -5.40) | -6.40 (-8.05 to -4.75) | -6.40 (-8.05 to -4.75) |  | -6.40 (-8.04 to -4.76) | -6.40 (-8.13 to -4.67) | -6.40 (-8.03 to -4.77) |
| Orfor PO 36mg QD | -9.35 (-11.16 to -7.87) | -10.24 (-16.66 to -3.90) | -9.00 (-10.66 to -7.34) | -9.00 (-10.66 to -7.34) | -9.00 (-10.68 to -7.32) |  | -9.00 (-10.66 to -7.34) |  |  | -9.00 (-10.00 to -8.00) | -9.00 (-10.65 to -7.35) | -9.00 (-10.65 to -7.35) |  | -9.00 (-10.64 to -7.36) | -9.00 (-10.73 to -7.27) | -9.00 (-10.63 to -7.37) |
| Orfor PO 6mg QD | -5.97 (-7.86 to -4.49) | -7.56 (-14.36 to -0.66) | -5.60 (-7.26 to -3.94) | -5.60 (-7.26 to -3.94) | -5.60 (-7.28 to -3.92) |  | -5.60 (-7.26 to -3.94) |  |  | -5.60 (-6.60 to -4.60) | -5.60 (-7.25 to -3.95) | -5.60 (-7.25 to -3.95) |  | -5.60 (-7.24 to -3.96) | -5.60 (-7.33 to -3.87) | -5.60 (-7.23 to -3.97) |
| Orli PO 120mg TID | -3.11 (-4.23 to -1.93) | -3.09 (-4.33 to -1.76) | -3.13 (-4.23 to -2.03) | -3.13 (-4.23 to -2.03) | -3.13 (-4.24 to -2.02) | -3.17 (-3.93 to -2.41) | -3.13 (-4.23 to -2.03) |  | -3.13 (-4.23 to -2.03) |  | -3.33 (-4.18 to -2.48) | -3.33 (-4.18 to -2.48) |  | -3.13 (-4.22 to -2.05) | -3.13 (-4.26 to -1.99) | -3.13 (-4.21 to -2.05) |
| Orli PO 60mg TID | -2.14 (-3.55 to -0.68) | -2.13 (-3.63 to -0.57) | -2.02 (-3.67 to -0.36) | -2.02 (-3.67 to -0.36) | -2.01 (-3.68 to -0.35) | -2.04 (-3.16 to -0.91) | -2.02 (-3.67 to -0.36) |  | -2.02 (-3.67 to -0.36) |  | -2.12 (-3.72 to -0.51) | -2.12 (-3.72 to -0.51) |  | -2.02 (-3.65 to -0.38) | -2.01 (-3.73 to -0.30) | -2.02 (-3.64 to -0.39) |
| Pemvi SC 1.2mg QW | -7.63 (-10.06 to -5.01) | -8.08 (-10.77 to -5.15) | -7.20 (-9.77 to -4.63) | -7.20 (-9.77 to -4.63) | -7.20 (-9.78 to -4.62) | -7.20 (-9.40 to -5.00) | -7.20 (-9.77 to -4.63) | -7.20 (-9.40 to -5.00) |  | -7.20 (-9.40 to -5.00) | -7.20 (-9.76 to -4.64) | -7.20 (-9.76 to -4.64) |  | -7.20 (-9.75 to -4.65) | -7.20 (-9.82 to -4.58) | -7.20 (-9.75 to -4.65) |
| Pemvi SC 1.8mg QW | -9.07 (-11.47 to -6.56) | -9.50 (-12.23 to -6.70) | -8.90 (-11.47 to -6.33) | -8.90 (-11.47 to -6.33) | -8.90 (-11.48 to -6.32) | -8.90 (-11.10 to -6.70) | -8.90 (-11.47 to -6.33) | -8.90 (-11.10 to -6.70) |  | -8.90 (-11.10 to -6.70) | -8.90 (-11.46 to -6.34) | -8.90 (-11.46 to -6.34) |  | -8.90 (-11.45 to -6.35) | -8.90 (-11.52 to -6.28) | -8.90 (-11.45 to -6.35) |
| Pemvi SC 2.4mg QW | -10.23 (-12.54 to -7.84) | -10.69 (-13.19 to -8.10) | -10.30 (-12.78 to -7.82) | -10.30 (-12.78 to -7.82) | -10.30 (-12.80 to -7.80) | -10.30 (-12.40 to -8.20) | -10.30 (-12.78 to -7.82) | -10.30 (-12.40 to -8.20) |  | -10.30 (-12.40 to -8.20) | -10.30 (-12.78 to -7.82) | -10.30 (-12.78 to -7.82) |  | -10.30 (-12.77 to -7.83) | -10.30 (-12.83 to -7.77) | -10.30 (-12.76 to -7.84) |
| Reta SC 12mg QW | -19.80 (-21.81 to -17.78) | -0.71 (-27.47 to 25.07) | -20.00 (-21.93 to -18.07) | -20.00 (-21.93 to -18.07) | -20.00 (-21.94 to -18.06) |  | -20.00 (-21.93 to -18.07) |  |  | -20.00 (-21.40 to -18.60) | -20.00 (-21.92 to -18.08) | -20.00 (-21.92 to -18.08) | -22.10 (-24.90 to -19.30) | -20.00 (-21.91 to -18.09) | -20.00 (-21.99 to -18.01) | -20.00 (-21.90 to -18.10) |
| Reta SC 4mg QW | -13.21 (-15.16 to -11.24) | -0.16 (-27.16 to 25.56) | -13.20 (-15.06 to -11.34) | -13.20 (-15.06 to -11.34) | -13.20 (-15.07 to -11.33) |  | -13.20 (-15.06 to -11.34) |  |  | -13.20 (-14.50 to -11.90) | -13.20 (-15.05 to -11.35) | -13.20 (-15.05 to -11.35) |  | -13.20 (-15.04 to -11.36) | -13.20 (-15.12 to -11.28) | -13.20 (-15.03 to -11.37) |
| Reta SC 9mg QW | -18.64 (-20.62 to -16.67) | -0.77 (-27.38 to 25.25) | -18.80 (-20.73 to -16.87) | -18.80 (-20.73 to -16.87) | -18.80 (-20.74 to -16.86) |  | -18.80 (-20.73 to -16.87) |  |  | -18.80 (-20.20 to -17.40) | -18.80 (-20.72 to -16.88) | -18.80 (-20.72 to -16.88) |  | -18.80 (-20.71 to -16.89) | -18.80 (-20.79 to -16.81) | -18.80 (-20.70 to -16.90) |
| Sema PO 25mg QD | -11.51 (-14.15 to -8.72) | -11.32 (-15.68 to -6.85) | -11.90 (-14.73 to -9.07) | -11.90 (-14.73 to -9.07) | -11.90 (-14.74 to -9.06) |  | -11.90 (-14.73 to -9.07) |  |  | -11.90 (-14.40 to -9.40) | -11.90 (-14.72 to -9.08) | -11.90 (-14.72 to -9.08) |  | -11.90 (-14.72 to -9.08) | -11.90 (-14.77 to -9.03) | -11.90 (-14.71 to -9.09) |
| Sema PO 50mg QD | -12.69 (-14.44 to -10.75) | -12.67 (-18.01 to -7.18) | -12.90 (-14.83 to -10.97) | -12.90 (-14.83 to -10.97) | -12.90 (-14.84 to -10.96) |  | -12.90 (-14.83 to -10.97) |  |  | -12.90 (-14.30 to -11.50) | -12.90 (-14.82 to -10.98) | -12.90 (-14.82 to -10.98) |  | -12.90 (-14.81 to -10.99) | -12.90 (-14.89 to -10.91) | -12.90 (-14.80 to -11.00) |
| Sema SC 0.05mg QD | -3.50 (-5.42 to -1.48) | -3.53 (-5.83 to -1.36) | -2.98 (-5.24 to -0.72) | -2.98 (-5.24 to -0.72) | -3.01 (-5.28 to -0.74) | -3.11 (-5.06 to -1.16) | -2.98 (-5.24 to -0.72) |  |  | -3.70 (-5.80 to -1.60) | -3.32 (-5.52 to -1.12) | -3.32 (-5.52 to -1.12) |  | -3.24 (-5.61 to -0.88) | -2.68 (-4.84 to -0.53) | -2.99 (-5.23 to -0.75) |
| Sema SC 0.1mg QD | -5.97 (-7.88 to -3.97) | -6.02 (-8.14 to -3.90) | -5.58 (-7.84 to -3.32) | -5.58 (-7.84 to -3.32) | -5.61 (-7.88 to -3.34) | -5.71 (-7.66 to -3.76) | -5.58 (-7.84 to -3.32) |  |  | -6.30 (-8.40 to -4.20) | -5.92 (-8.12 to -3.72) | -5.92 (-8.12 to -3.72) |  | -5.84 (-8.21 to -3.48) | -5.28 (-7.44 to -3.13) | -5.59 (-7.83 to -3.35) |
| Sema SC 0.2mg QD | -8.79 (-10.76 to -6.86) | -8.89 (-11.09 to -6.67) | -8.58 (-10.84 to -6.32) | -8.58 (-10.84 to -6.32) | -8.61 (-10.88 to -6.34) | -8.71 (-10.66 to -6.76) | -8.58 (-10.84 to -6.32) |  |  | -9.30 (-11.40 to -7.20) | -8.92 (-11.12 to -6.72) | -8.92 (-11.12 to -6.72) |  | -8.84 (-11.21 to -6.48) | -8.28 (-10.44 to -6.13) | -8.59 (-10.83 to -6.35) |
| Sema SC 0.3mg QD fast | -8.68 (-10.59 to -6.64) | -8.68 (-10.85 to -6.48) | -8.39 (-10.65 to -6.13) | -8.39 (-10.65 to -6.13) | -8.42 (-10.69 to -6.15) | -8.52 (-10.47 to -6.57) | -8.39 (-10.65 to -6.13) |  |  | -9.11 (-11.21 to -7.01) | -8.73 (-10.93 to -6.53) | -8.73 (-10.93 to -6.53) |  | -8.65 (-11.02 to -6.29) | -8.07 (-10.20 to -5.93) | -8.40 (-10.64 to -6.16) |
| Sema SC 0.3mg QD standard | -8.41 (-10.36 to -6.47) | -8.47 (-10.68 to -6.35) | -8.18 (-10.44 to -5.92) | -8.18 (-10.44 to -5.92) | -8.21 (-10.48 to -5.94) | -8.31 (-10.26 to -6.36) | -8.18 (-10.44 to -5.92) |  |  | -8.90 (-11.00 to -6.80) | -8.52 (-10.72 to -6.32) | -8.52 (-10.72 to -6.32) |  | -8.44 (-10.81 to -6.08) | -7.86 (-9.99 to -5.72) | -8.19 (-10.43 to -5.95) |
| Sema SC 0.4mg QD fast | -13.26 (-15.22 to -11.29) | -13.33 (-15.54 to -11.23) | -13.26 (-15.53 to -10.98) | -13.26 (-15.53 to -10.98) | -13.30 (-15.57 to -11.00) | -13.39 (-15.35 to -11.43) | -13.26 (-15.53 to -10.98) |  |  | -13.95 (-16.05 to -11.85) | -13.58 (-15.80 to -11.36) | -13.58 (-15.80 to -11.36) |  | -13.33 (-15.60 to -11.06) | -13.16 (-15.47 to -10.84) | -13.27 (-15.52 to -11.01) |
| Sema SC 0.4mg QD standard | -10.93 (-12.86 to -8.90) | -10.99 (-13.17 to -8.87) | -10.78 (-13.04 to -8.52) | -10.78 (-13.04 to -8.52) | -10.81 (-13.08 to -8.54) | -10.91 (-12.86 to -8.96) | -10.78 (-13.04 to -8.52) |  |  | -11.50 (-13.60 to -9.40) | -11.12 (-13.32 to -8.92) | -11.12 (-13.32 to -8.92) |  | -11.04 (-13.41 to -8.68) | -10.46 (-12.59 to -8.32) | -10.79 (-13.03 to -8.55) |
| Sema SC 2.4mg QW | -11.60 (-12.32 to -10.89) | -11.55 (-12.87 to -10.36) | -11.49 (-12.23 to -10.75) | -11.49 (-12.23 to -10.75) | -11.81 (-12.68 to -10.94) | -10.86 (-11.89 to -9.84) | -11.49 (-12.23 to -10.75) |  | -11.49 (-12.23 to -10.75) |  | -11.52 (-12.25 to -10.79) | -11.52 (-12.25 to -10.79) |  | -11.48 (-12.21 to -10.75) | -11.52 (-12.27 to -10.76) | -11.60 (-12.29 to -10.91) |
| Sema SC 7.2mg QW | -14.26 (-15.76 to -12.75) | -13.61 (-21.49 to -6.52) | -14.34 (-16.03 to -12.65) | -14.34 (-16.03 to -12.65) | -14.48 (-16.20 to -12.77) |  | -14.34 (-16.03 to -12.65) |  |  | -14.30 (-15.70 to -12.90) | -14.35 (-16.04 to -12.67) | -14.35 (-16.04 to -12.67) |  | -14.33 (-16.01 to -12.66) | -14.35 (-16.10 to -12.61) | -14.39 (-16.05 to -12.73) |
| Survo SC 3.6mg QW | -7.32 (-9.61 to -4.68) | -10.25 (-15.63 to -4.62) | -6.50 (-8.90 to -4.10) | -6.50 (-8.90 to -4.10) | -6.50 (-8.91 to -4.09) |  | -6.50 (-8.90 to -4.10) |  |  | -6.50 (-8.50 to -4.50) | -6.50 (-8.89 to -4.11) | -6.50 (-8.89 to -4.11) |  | -6.50 (-8.88 to -4.12) | -6.50 (-8.95 to -4.05) | -6.50 (-8.88 to -4.12) |
| Survo SC 6mg QW | -8.10 (-10.42 to -5.56) | -10.44 (-15.94 to -4.53) | -7.40 (-9.88 to -4.92) | -7.40 (-9.88 to -4.92) | -7.40 (-9.90 to -4.90) |  | -7.40 (-9.88 to -4.92) |  |  | -7.40 (-9.50 to -5.30) | -7.40 (-9.88 to -4.92) | -7.40 (-9.88 to -4.92) |  | -7.40 (-9.87 to -4.93) | -7.40 (-9.93 to -4.87) | -7.40 (-9.86 to -4.94) |
| TZP SC 10mg QW | -15.19 (-16.52 to -13.79) | -12.38 (-15.53 to -8.96) | -14.89 (-16.36 to -13.43) | -14.89 (-16.36 to -13.43) | -14.94 (-16.42 to -13.46) | -11.30 (-14.30 to -8.30) | -14.89 (-16.36 to -13.43) |  | -14.89 (-16.36 to -13.43) |  | -14.90 (-16.36 to -13.44) | -14.90 (-16.36 to -13.44) |  | -14.88 (-16.35 to -13.41) | -14.89 (-16.39 to -13.40) | -14.93 (-16.37 to -13.49) |
| TZP SC 15mg QW | -16.87 (-18.07 to -15.51) | -15.31 (-18.40 to -12.30) | -16.90 (-18.15 to -15.64) | -16.90 (-18.15 to -15.64) | -17.01 (-18.28 to -15.73) | -15.10 (-18.15 to -12.05) | -16.90 (-18.15 to -15.64) |  | -16.90 (-18.15 to -15.64) |  | -16.91 (-18.16 to -15.66) | -16.91 (-18.16 to -15.66) |  | -16.94 (-18.18 to -15.69) | -16.86 (-18.15 to -15.56) | -16.94 (-18.18 to -15.70) |
| TZP SC 5mg QW | -11.26 (-12.66 to -9.80) | -11.43 (-18.86 to -1.10) | -11.16 (-12.76 to -9.56) | -11.16 (-12.76 to -9.56) | -11.22 (-12.83 to -9.60) |  | -11.16 (-12.76 to -9.56) |  | -11.16 (-12.76 to -9.56) |  | -11.17 (-12.77 to -9.58) | -11.17 (-12.77 to -9.58) |  | -11.21 (-12.83 to -9.59) | -11.12 (-12.74 to -9.50) | -11.19 (-12.76 to -9.62) |
| TZP SC MTD 10-15mg QW | -16.47 (-18.04 to -14.85) | -16.38 (-18.04 to -14.60) | -16.66 (-18.29 to -15.02) | -16.66 (-18.29 to -15.02) |  | -16.64 (-17.97 to -15.30) | -16.66 (-18.29 to -15.02) |  | -16.66 (-18.29 to -15.02) |  | -16.66 (-18.29 to -15.03) | -16.66 (-18.29 to -15.03) |  | -16.66 (-18.28 to -15.03) | -16.66 (-18.33 to -14.99) | -16.66 (-18.28 to -15.04) |
| NB PO 32/360mg QD |  |  |  |  |  |  |  |  |  |  | -4.80 (-6.34 to -3.26) | -4.80 (-6.34 to -3.26) |  |  |  |  |
| PHEN/TPM PO 15/92mg QD |  |  |  |  |  |  |  |  |  |  | -9.37 (-11.06 to -7.68) | -9.37 (-11.06 to -7.68) |  |  |  |  |
| PHEN/TPM PO 3.75/23mg QD |  |  |  |  |  |  |  |  |  |  | -3.55 (-5.42 to -1.68) | -3.55 (-5.42 to -1.68) |  |  |  |  |
| Reta SC 1mg QW |  |  |  |  |  |  |  |  |  |  |  |  | -6.60 (-8.95 to -4.25) |  |  |  |
| Reta SC 4mg QW fast |  |  |  |  |  |  |  |  |  |  |  |  | -15.70 (-19.05 to -12.35) |  |  |  |
| Reta SC 4mg QW slow |  |  |  |  |  |  |  |  |  |  |  |  | -14.20 (-17.60 to -10.80) |  |  |  |
| Reta SC 8mg QW fast |  |  |  |  |  |  |  |  |  |  |  |  | -21.80 (-25.10 to -18.50) |  |  |  |
| Reta SC 8mg QW slow |  |  |  |  |  |  |  |  |  |  |  |  | -19.60 (-22.70 to -16.50) |  |  |  |

**Table S13.** Leave-one-parent-trial-out analysis (maximum influence for interventions supported by >=2 trials) Single-trial nodes (39 node[s]) are omitted because leaving out their sole trial disconnects the node and renders its effect non-estimable.

| Intervention | Trials in network | Main MD | MD without trial | Left-out parent trial | Shift |
|---|---|---|---|---|---|
| TZP SC 10mg QW | 2 | -14.894 | -11.922 | p027 | 2.9718 |
| Lira SC 3mg QD | 3 | -4.067 | -3.415 | p017 | 0.65203 |
| TZP SC 15mg QW | 3 | -16.896 | -17.272 | p030 | -0.37536 |
| Sema SC 2.4mg QW | 8 | -11.49 | -11.117 | p001 | 0.37347 |
| Orli PO 120mg TID | 3 | -3.1296 | -2.9032 | p164 | 0.22641 |
| TZP SC MTD 10-15mg QW | 2 | -16.658 | -16.637 | p001 | 0.020586 |

**Table S14.**
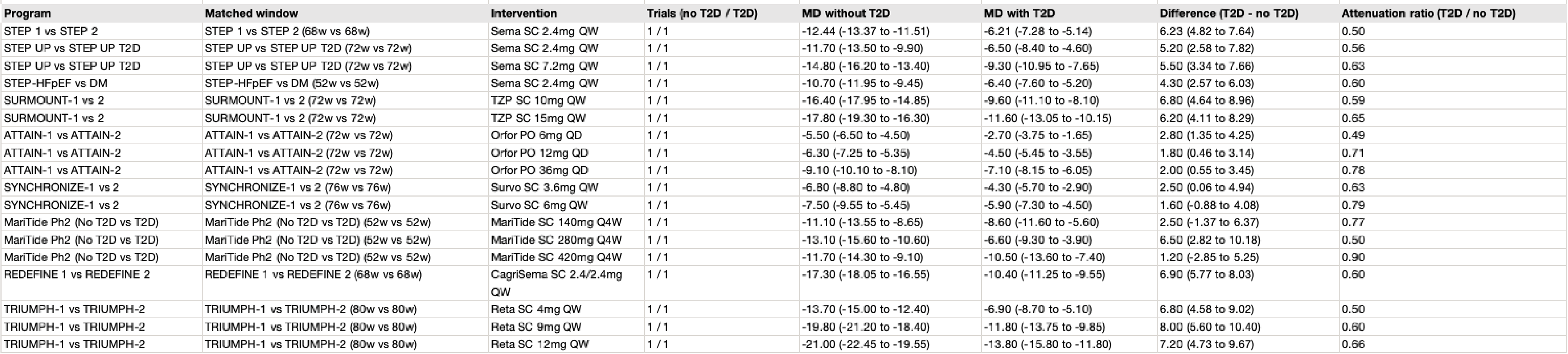
Timepoint-matched comparison of weight reduction by type 2 diabetes status (multilevel ratio-of-means model) Multilevel random-effects ratio-of-means model (clustering doses within 9 timepoint-matched trial programs, 18 dose pairs): placebo-adjusted percentage weight reduction in adults with T20 was 62.3% of that in adults without T20 (ratio 0.62, 95% Cl 0.58 to 0.66; relative attenuation 37.7%, 95% Cl 33.69).

| Program | Matched window | Intervention | Trials (no T2D / T2D) | MD without T2D | MD with T2D | Difference (T2D - no T2D) | Attenuation ratio (T2D / no T2D) |
|---|---|---|---|---|---|---|---|
| STEP 1 vs STEP 2 | STEP 1 vs STEP 2 (68w vs 68w) | Sema SC 2.4mg QW | 1 / 1 | -12.44 (-13.37 to -11.51) | -6.21 (-7.28 to -5.14) | 6.23 (4.82 to 7.64) | 0.50 |
| STEP UP vs STEP UP T2D | STEP UP vs STEP UP T2D (72w vs 72w) | Sema SC 2.4mg QW | 1 / 1 | -11.70 (-13.50 to -9.90) | -6.50 (-8.40 to -4.60) | 5.20 (2.58 to 7.82) | 0.56 |
| STEP UP vs STEP UP T2D | STEP UP vs STEP UP T2D (72w vs 72w) | Sema SC 7.2mg QW | 1 / 1 | -14.80 (-16.20 to -13.40) | -9.30 (-10.95 to -7.65) | 5.50 (3.34 to 7.66) | 0.63 |
| STEP-HFpEF vs DM | STEP-HFpEF vs DM (52w vs 52w) | Sema SC 2.4mg QW | 1 / 1 | -10.70 (-11.95 to -9.45) | -6.40 (-7.60 to -5.20) | 4.30 (2.57 to 6.03) | 0.60 |
| SURMOUNT-1 vs 2 | SURMOUNT-1 vs 2 (72w vs 72w) | TZP SC 10mg QW | 1 / 1 | -16.40 (-17.95 to -14.85) | -9.60 (-11.10 to -8.10) | 6.80 (4.64 to 8.96) | 0.59 |
| SURMOUNT-1 vs 2 | SURMOUNT-1 vs 2 (72w vs 72w) | TZP SC 15mg QW | 1 / 1 | -17.80 (-19.30 to -16.30) | -11.60 (-13.05 to -10.15) | 6.20 (4.11 to 8.29) | 0.65 |
| ATTAIN-1 vs ATTAIN-2 | ATTAIN-1 vs ATTAIN-2 (72w vs 72w) | Orfor PO 6mg QD | 1 / 1 | -5.50 (-6.50 to -4.50) | -2.70 (-3.75 to -1.65) | 2.80 (1.35 to 4.25) | 0.49 |
| ATTAIN-1 vs ATTAIN-2 | ATTAIN-1 vs ATTAIN-2 (72w vs 72w) | Orfor PO 12mg QD | 1 / 1 | -6.30 (-7.25 to -5.35) | -4.50 (-5.45 to -3.55) | 1.80 (0.46 to 3.14) | 0.71 |
| ATTAIN-1 vs ATTAIN-2 | ATTAIN-1 vs ATTAIN-2 (72w vs 72w) | Orfor PO 36mg QD | 1 / 1 | -9.10 (-10.10 to -8.10) | -7.10 (-8.15 to -6.05) | 2.00 (0.55 to 3.45) | 0.78 |
| SYNCHRONIZE-1 vs 2 | SYNCHRONIZE-1 vs 2 (76w vs 76w) | Survo SC 3.6mg QW | 1 / 1 | -6.80 (-8.80 to -4.80) | -4.30 (-5.70 to -2.90) | 2.50 (0.06 to 4.94) | 0.63 |
| SYNCHRONIZE-1 vs 2 | SYNCHRONIZE-1 vs 2 (76w vs 76w) | Survo SC 6mg QW | 1 / 1 | -7.50 (-9.55 to -5.45) | -5.90 (-7.30 to -4.50) | 1.60 (-0.88 to 4.08) | 0.79 |
| MariTide Ph2 (No T2D vs T2D) | MariTide Ph2 (No T2D vs T2D) (52w vs 52w) | MariTide SC 140mg Q4W | 1 / 1 | -11.10 (-13.55 to -8.65) | -8.60 (-11.60 to -5.60) | 2.50 (-1.37 to 6.37) | 0.77 |
| MariTide Ph2 (No T2D vs T2D) | MariTide Ph2 (No T2D vs T2D) (52w vs 52w) | MariTide SC 280mg Q4W | 1 / 1 | -13.10 (-15.60 to -10.60) | -6.60 (-9.30 to -3.90) | 6.50 (2.82 to 10.18) | 0.50 |
| MariTide Ph2 (No T2D vs T2D) | MariTide Ph2 (No T2D vs T2D) (52w vs 52w) | MariTide SC 420mg Q4W | 1 / 1 | -11.70 (-14.30 to -9.10) | -10.50 (-13.60 to -7.40) | 1.20 (-2.85 to 5.25) | 0.90 |
| REDEFINE 1 vs REDEFINE 2 | REDEFINE 1 vs REDEFINE 2 (68w vs 68w) | CagriSema SC 2.4/2.4mg QW | 1 / 1 | -17.30 (-18.05 to -16.55) | -10.40 (-11.25 to -9.55) | 6.90 (5.77 to 8.03) | 0.60 |
| TRIUMPH-1 vs TRIUMPH-2 | TRIUMPH-1 vs TRIUMPH-2 (80w vs 80w) | Reta SC 4mg QW | 1 / 1 | -13.70 (-15.00 to -12.40) | -6.90 (-8.70 to -5.10) | 6.80 (4.58 to 9.02) | 0.50 |
| TRIUMPH-1 vs TRIUMPH-2 | TRIUMPH-1 vs TRIUMPH-2 (80w vs 80w) | Reta SC 9mg QW | 1 / 1 | -19.80 (-21.20 to -18.40) | -11.80 (-13.75 to -9.85) | 8.00 (5.60 to 10.40) | 0.60 |
| TRIUMPH-1 vs TRIUMPH-2 | TRIUMPH-1 vs TRIUMPH-2 (80w vs 80w) | Reta SC 12mg QW | 1 / 1 | -21.00 (-22.45 to -19.55) | -13.80 (-15.80 to -11.80) | 7.20 (4.73 to 9.67) | 0.66 |

**Table S15.** Missing results and reporting status at 48-56 weeks and 60·80 weeks (with ROB-MEN reporting-bias summary) Distinguishes outcomes directly available at 48-56 weeks, outcomes reported at 60-80 weeks (incorporated into the primary longitudinal model), and outcomes not reported in the primary source. ROB-MEN evaluation across 71 vs-placebo network comparisons identified 27 comparison(s) at high risk and 5 at some concerns.

| population | outcome | not_reported_or_not_extracted_at_48_56w | reported_at_60_80w_primary_endpoint_in_longitudinal | reported_usable_at_48_56w | reported_at_48_56w_other_estimand_or_role |
|---|---|---|---|---|---|
| mixed population | AE-related permanent discontinuation | 6 | 4 | 3 | 0 |
| mixed population | All-cause permanent discontinuation | 12 | 1 | 0 | 0 |
| mixed population | Constipation | 10 | 1 | 2 | 0 |
| mixed population | Diarrhoea | 10 | 2 | 1 | 0 |
| mixed population | Nausea | 9 | 2 | 2 | 0 |
| mixed population | Serious adverse events | 6 | 4 | 3 | 0 |
| mixed population | Vomiting | 10 | 2 | 1 | 0 |
| mixed population | % body-weight change | 5 | 4 | 4 | 0 |
| mixed population | >=10% weight reduction | 7 | 3 | 2 | 1 |
| mixed population | >=15% weight reduction | 9 | 3 | 0 | 1 |
| mixed population | >=20% weight reduction | 9 | 3 | 0 | 1 |
| mixed population | >=5% weight reduction | 6 | 3 | 3 | 1 |
| without T2D | AE-related permanent discontinuation | 15 | 15 | 17 | 0 |
| without T2D | All-cause permanent discontinuation | 21 | 12 | 14 | 0 |
| without T2D | Constipation | 36 | 7 | 4 | 0 |
| without T2D | Diarrhoea | 34 | 7 | 6 | 0 |
| without T2D | Nausea | 30 | 7 | 10 | 0 |
| without T2D | Serious adverse events | 18 | 14 | 15 | 0 |
| without T2D | Vomiting | 35 | 7 | 5 | 0 |
| without T2D | % body-weight change | 10 | 2 | 35 | 0 |
| without T2D | >=10% weight reduction | 25 | 13 | 7 | 2 |
| without T2D | >=15% weight reduction | 28 | 13 | 5 | 1 |
| without T2D | >=20% weight reduction | 31 | 13 | 3 | 0 |
| without T2D | >=5% weight reduction | 25 | 13 | 6 | 3 |
| with T2D | AE-related permanent discontinuation | 5 | 7 | 6 | 0 |
| with T2D | All-cause permanent discontinuation | 16 | 2 | 0 | 0 |
| with T2D | Constipation | 14 | 2 | 2 | 0 |
| with T2D | Diarrhoea | 11 | 5 | 2 | 0 |
| with T2D | Nausea | 11 | 5 | 2 | 0 |
| with T2D | Serious adverse events | 6 | 7 | 5 | 0 |
| with T2D | Vomiting | 13 | 3 | 2 | 0 |
| with T2D | % body-weight change | 3 | 2 | 13 | 0 |
| with T2D | >=10% weight reduction | 10 | 6 | 2 | 0 |
| with T2D | >=15% weight reduction | 12 | 5 | 1 | 0 |
| with T2D | >=20% weight reduction | 12 | 5 | 1 | 0 |
| with T2D | >=5% weight reduction | 10 | 6 | 2 | 0 |

**Table S16.** Dose-escalation benefit-discontinuation trade-off across adjacent maintenance doses (non-extrapolated 52-week nodes) Restricted to target maintenance doses with non-extrapolated 52-week support (protocol sections 20, 41 and 56). Fixed-dose combinations are compared only across identical companion-component doses. Prespecified probability thresholds: high (P >= 0.80), moderate/uncertain (0.50 <= P < 0.80), low (P < 0.50).

| Comparison | Efficacy source | Safety source | Tolerance (pp) | Incremental weight MD (95% CI) | P(incremental benefit) | All-cause disc OR (95% CI) | All-cause disc RD /1000 (95% CI) | AE discontinuation OR (95% CI) | AE disc RD /1000 (95% CI) | Interpretation |
|---|---|---|---|---|---|---|---|---|---|---|
| Ecnoglutide (SUBCUTANEOUS QW): 1.8mg vs 1.2mg | longitudinal_52w_primary | w52_strict_48_56w | 0.5 | -3.00 (-5.62 to -0.38) | 0.96915 |  |  | 1.41 (0.27 to 7.24) | +5 (-9 to +69) | Favours higher dose for weight loss (P >= 0.80), with imprecisely estimated AE discontinuation difference (95% CI crosses 1.0). |
| Ecnoglutide (SUBCUTANEOUS QW): 1.8mg vs 1.2mg | longitudinal_52w_primary | w52_strict_48_56w | 1 | -3.00 (-5.62 to -0.38) | 0.93253 |  |  | 1.41 (0.27 to 7.24) | +5 (-9 to +69) | Favours higher dose for weight loss (P >= 0.80), with imprecisely estimated AE discontinuation difference (95% CI crosses 1.0). |
| Ecnoglutide (SUBCUTANEOUS QW): 1.8mg vs 1.2mg | longitudinal_52w_primary | w52_strict_48_56w | 2 | -3.00 (-5.62 to -0.38) | 0.77269 |  |  | 1.41 (0.27 to 7.24) | +5 (-9 to +69) | Moderate/uncertain probability (0.50 <= P < 0.80) that incremental weight loss exceeds threshold; individualize based on tolerability. |
| Ecnoglutide (SUBCUTANEOUS QW): 2.4mg vs 1.8mg | longitudinal_52w_primary | w52_strict_48_56w | 0.5 | -1.98 (-4.61 to 0.66) | 0.8636 |  |  | 1.58 (0.41 to 6.15) | +10 (-11 to +84) | Favours higher dose for weight loss (P >= 0.80), with imprecisely estimated AE discontinuation difference (95% CI crosses 1.0). |
| Ecnoglutide (SUBCUTANEOUS QW): 2.4mg vs 1.8mg | longitudinal_52w_primary | w52_strict_48_56w | 1 | -1.98 (-4.61 to 0.66) | 0.76579 |  |  | 1.58 (0.41 to 6.15) | +10 (-11 to +84) | Moderate/uncertain probability (0.50 <= P < 0.80) that incremental weight loss exceeds threshold; individualize based on tolerability. |
| Ecnoglutide (SUBCUTANEOUS QW): 2.4mg vs 1.8mg | longitudinal_52w_primary | w52_strict_48_56w | 2 | -1.98 (-4.61 to 0.66) | 0.49278 |  |  | 1.58 (0.41 to 6.15) | +10 (-11 to +84) | Favours maintaining lower dose: low probability (P < 0.50) that incremental weight loss exceeds the prespecified threshold. |
| Eloralintide (SUBCUTANEOUS QW): 3mg vs 1mg | longitudinal_52w_primary | not_reported_at_48_80w | 0.5 | -1.62 (-6.21 to 2.97) | 0.68365 | 0.23 (0.06 to 0.85) | -298 (-413 to -40) |  |  | Moderate/uncertain probability (0.50 <= P < 0.80) that incremental weight loss exceeds threshold; individualize based on tolerability. |
| Eloralintide (SUBCUTANEOUS QW): 3mg vs 1mg | longitudinal_52w_primary | not_reported_at_48_80w | 1 | -1.62 (-6.21 to 2.97) | 0.60429 | 0.23 (0.06 to 0.85) | -298 (-413 to -40) |  |  | Moderate/uncertain probability (0.50 <= P < 0.80) that incremental weight loss exceeds threshold; individualize based on tolerability. |
| Eloralintide (SUBCUTANEOUS QW): 3mg vs 1mg | longitudinal_52w_primary | not_reported_at_48_80w | 2 | -1.62 (-6.21 to 2.97) | 0.43547 | 0.23 (0.06 to 0.85) | -298 (-413 to -40) |  |  | Favours maintaining lower dose: low probability (P < 0.50) that incremental weight loss exceeds the prespecified threshold. |
| Eloralintide (SUBCUTANEOUS QW): 6mg vs 3mg | longitudinal_52w_primary | not_reported_at_48_80w | 0.5 | -2.27 (-6.88 to 2.34) | 0.77376 | 1.67 (0.42 to 6.58) | +83 (-89 to +401) |  |  | Moderate/uncertain probability (0.50 <= P < 0.80) that incremental weight loss exceeds threshold; individualize based on tolerability. |
| Eloralintide (SUBCUTANEOUS QW): 6mg vs 3mg | longitudinal_52w_primary | not_reported_at_48_80w | 1 | -2.27 (-6.88 to 2.34) | 0.70499 | 1.67 (0.42 to 6.58) | +83 (-89 to +401) |  |  | Moderate/uncertain probability (0.50 <= P < 0.80) that incremental weight loss exceeds threshold; individualize based on tolerability. |
| Eloralintide (SUBCUTANEOUS QW): 6mg vs 3mg | longitudinal_52w_primary | not_reported_at_48_80w | 2 | -2.27 (-6.88 to 2.34) | 0.54533 | 1.67 (0.42 to 6.58) | +83 (-89 to +401) |  |  | Moderate/uncertain probability (0.50 <= P < 0.80) that incremental weight loss exceeds threshold; individualize based on tolerability. |
| Eloralintide (SUBCUTANEOUS QW): 9mg vs 6mg | longitudinal_52w_primary | not_reported_at_48_80w | 0.5 | -2.75 (-7.23 to 1.73) | 0.83704 | 0.52 (0.17 to 1.63) | -102 (-197 to +102) |  |  | High probability (P >= 0.80) of exceeding weight-loss threshold; AE discontinuation not reported at 48-80 weeks. |
| Eloralintide (SUBCUTANEOUS QW): 9mg vs 6mg | longitudinal_52w_primary | not_reported_at_48_80w | 1 | -2.75 (-7.23 to 1.73) | 0.77746 | 0.52 (0.17 to 1.63) | -102 (-197 to +102) |  |  | Moderate/uncertain probability (0.50 <= P < 0.80) that incremental weight loss exceeds threshold; individualize based on tolerability. |
| Eloralintide (SUBCUTANEOUS QW): 9mg vs 6mg | longitudinal_52w_primary | not_reported_at_48_80w | 2 | -2.75 (-7.23 to 1.73) | 0.62786 | 0.52 (0.17 to 1.63) | -102 (-197 to +102) |  |  | Moderate/uncertain probability (0.50 <= P < 0.80) that incremental weight loss exceeds threshold; individualize based on tolerability. |
| Maridebart_cafraglutide (SUBCUTANEOUS Q4W): 280mg vs 140mg | longitudinal_52w_primary | w52_strict_48_56w | 0.5 | -1.46 (-3.68 to 0.77) | 0.80002 | 0.78 (0.39 to 1.56) | -52 (-167 to +104) | 1.00 (0.41 to 2.47) | -0 (-80 to +148) | Favours higher dose for weight loss (P >= 0.80), with imprecisely estimated AE discontinuation difference (95% CI crosses 1.0). |
| Maridebart_cafraglutide (SUBCUTANEOUS Q4W): 280mg vs 140mg | longitudinal_52w_primary | w52_strict_48_56w | 1 | -1.46 (-3.68 to 0.77) | 0.65602 | 0.78 (0.39 to 1.56) | -52 (-167 to +104) | 1.00 (0.41 to 2.47) | -0 (-80 to +148) | Moderate/uncertain probability (0.50 <= P < 0.80) that incremental weight loss exceeds threshold; individualize based on tolerability. |
| Maridebart_cafraglutide (SUBCUTANEOUS Q4W): 280mg vs 140mg | longitudinal_52w_primary | w52_strict_48_56w | 2 | -1.46 (-3.68 to 0.77) | 0.31614 | 0.78 (0.39 to 1.56) | -52 (-167 to +104) | 1.00 (0.41 to 2.47) | -0 (-80 to +148) | Favours maintaining lower dose: low probability (P < 0.50) that incremental weight loss exceeds the prespecified threshold. |
| Maridebart_cafraglutide (SUBCUTANEOUS Q4W): 420mg vs 280mg | longitudinal_52w_primary | w52_strict_48_56w | 0.5 | 1.03 (-1.18 to 3.23) | 0.087095 | 0.90 (0.49 to 1.65) | -20 (-117 to +109) | 1.65 (0.71 to 3.79) | +72 (-36 to +244) | Favours maintaining lower dose: low probability (P < 0.50) that incremental weight loss exceeds the prespecified threshold. |
| Maridebart_cafraglutide (SUBCUTANEOUS Q4W): 420mg vs 280mg | longitudinal_52w_primary | w52_strict_48_56w | 1 | 1.03 (-1.18 to 3.23) | 0.035636 | 0.90 (0.49 to 1.65) | -20 (-117 to +109) | 1.65 (0.71 to 3.79) | +72 (-36 to +244) | Favours maintaining lower dose: low probability (P < 0.50) that incremental weight loss exceeds the prespecified threshold. |
| Maridebart_cafraglutide (SUBCUTANEOUS Q4W): 420mg vs 280mg | longitudinal_52w_primary | w52_strict_48_56w | 2 | 1.03 (-1.18 to 3.23) | 0.0035352 | 0.90 (0.49 to 1.65) | -20 (-117 to +109) | 1.65 (0.71 to 3.79) | +72 (-36 to +244) | Favours maintaining lower dose: low probability (P < 0.50) that incremental weight loss exceeds the prespecified threshold. |
| Mazdutide (SUBCUTANEOUS QW): 6mg vs 4mg | longitudinal_52w_primary | not_reported_at_48_80w | 0.5 | -2.67 (-4.07 to -1.27) | 0.99883 | 1.01 (0.51 to 1.99) | +0 (-42 to +74) |  |  | High probability (P >= 0.80) of exceeding weight-loss threshold; AE discontinuation not reported at 48-80 weeks. |
| Mazdutide (SUBCUTANEOUS QW): 6mg vs 4mg | longitudinal_52w_primary | not_reported_at_48_80w | 1 | -2.67 (-4.07 to -1.27) | 0.99042 | 1.01 (0.51 to 1.99) | +0 (-42 to +74) |  |  | High probability (P >= 0.80) of exceeding weight-loss threshold; AE discontinuation not reported at 48-80 weeks. |
| Mazdutide (SUBCUTANEOUS QW): 6mg vs 4mg | longitudinal_52w_primary | not_reported_at_48_80w | 2 | -2.67 (-4.07 to -1.27) | 0.82703 | 1.01 (0.51 to 1.99) | +0 (-42 to +74) |  |  | High probability (P >= 0.80) of exceeding weight-loss threshold; AE discontinuation not reported at 48-80 weeks. |
| Orforglipron (ORAL QD): 12mg vs 6mg | longitudinal_52w_primary | long_term_42_80w | 0.5 | -0.79 (-1.78 to 0.20) | 0.71877 | 1.04 (0.44 to 2.43) | +6 (-108 to +186) | 1.54 (1.01 to 2.35) | +26 (+0 to +63) | Uncertain incremental weight-loss benefit (0.50 <= P < 0.80) accompanied by significantly higher AE-related discontinuation; maintain lower dose unless greater weight loss is prioritized. |
| Orforglipron (ORAL QD): 12mg vs 6mg | longitudinal_52w_primary | long_term_42_80w | 1 | -0.79 (-1.78 to 0.20) | 0.33937 | 1.04 (0.44 to 2.43) | +6 (-108 to +186) | 1.54 (1.01 to 2.35) | +26 (+0 to +63) | Does not favour dose escalation: low probability (P < 0.50) of exceeding weight-loss threshold while AE-related discontinuation significantly increases. |
| Orforglipron (ORAL QD): 12mg vs 6mg | longitudinal_52w_primary | long_term_42_80w | 2 | -0.79 (-1.78 to 0.20) | 0.0081764 | 1.04 (0.44 to 2.43) | +6 (-108 to +186) | 1.54 (1.01 to 2.35) | +26 (+0 to +63) | Does not favour dose escalation: low probability (P < 0.50) of exceeding weight-loss threshold while AE-related discontinuation significantly increases. |
| Orforglipron (ORAL QD): 36mg vs 12mg | longitudinal_52w_primary | long_term_42_80w | 0.5 | -2.59 (-3.57 to -1.60) | 0.99998 | 1.11 (0.48 to 2.60) | +19 (-104 to +205) | 1.34 (0.94 to 1.93) | +24 (-5 to +63) | Favours higher dose for weight loss (P >= 0.80), with imprecisely estimated AE discontinuation difference (95% CI crosses 1.0). |
| Orforglipron (ORAL QD): 36mg vs 12mg | longitudinal_52w_primary | long_term_42_80w | 1 | -2.59 (-3.57 to -1.60) | 0.99921 | 1.11 (0.48 to 2.60) | +19 (-104 to +205) | 1.34 (0.94 to 1.93) | +24 (-5 to +63) | Favours higher dose for weight loss (P >= 0.80), with imprecisely estimated AE discontinuation difference (95% CI crosses 1.0). |
| Orforglipron (ORAL QD): 36mg vs 12mg | longitudinal_52w_primary | long_term_42_80w | 2 | -2.59 (-3.57 to -1.60) | 0.8786 | 1.11 (0.48 to 2.60) | +19 (-104 to +205) | 1.34 (0.94 to 1.93) | +24 (-5 to +63) | Favours higher dose for weight loss (P >= 0.80), with imprecisely estimated AE discontinuation difference (95% CI crosses 1.0). |
| Orlistat (ORAL TID): 120mg vs 60mg | longitudinal_52w_primary | not_reported_at_48_80w | 0.5 | -0.97 (-2.07 to 0.13) | 0.7987 | 1.06 (0.72 to 1.54) | +11 (-55 to +89) |  |  | Moderate/uncertain probability (0.50 <= P < 0.80) that incremental weight loss exceeds threshold; individualize based on tolerability. |
| Orlistat (ORAL TID): 120mg vs 60mg | longitudinal_52w_primary | not_reported_at_48_80w | 1 | -0.97 (-2.07 to 0.13) | 0.47958 | 1.06 (0.72 to 1.54) | +11 (-55 to +89) |  |  | Favours maintaining lower dose: low probability (P < 0.50) that incremental weight loss exceeds the prespecified threshold. |
| Orlistat (ORAL TID): 120mg vs 60mg | longitudinal_52w_primary | not_reported_at_48_80w | 2 | -0.97 (-2.07 to 0.13) | 0.033807 | 1.06 (0.72 to 1.54) | +11 (-55 to +89) |  |  | Favours maintaining lower dose: low probability (P < 0.50) that incremental weight loss exceeds the prespecified threshold. |
| Pemvidutide (SUBCUTANEOUS QW): 1.8mg vs 1.2mg | longitudinal_52w_primary | not_reported_at_48_80w | 0.5 | -1.46 (-3.66 to 0.75) | 0.80297 | 1.14 (0.59 to 2.20) | +23 (-79 to +165) |  |  | High probability (P >= 0.80) of exceeding weight-loss threshold; AE discontinuation not reported at 48-80 weeks. |
| Pemvidutide (SUBCUTANEOUS QW): 1.8mg vs 1.2mg | longitudinal_52w_primary | not_reported_at_48_80w | 1 | -1.46 (-3.66 to 0.75) | 0.65822 | 1.14 (0.59 to 2.20) | +23 (-79 to +165) |  |  | Moderate/uncertain probability (0.50 <= P < 0.80) that incremental weight loss exceeds threshold; individualize based on tolerability. |
| Pemvidutide (SUBCUTANEOUS QW): 1.8mg vs 1.2mg | longitudinal_52w_primary | not_reported_at_48_80w | 2 | -1.46 (-3.66 to 0.75) | 0.31499 | 1.14 (0.59 to 2.20) | +23 (-79 to +165) |  |  | Favours maintaining lower dose: low probability (P < 0.50) that incremental weight loss exceeds the prespecified threshold. |
| Pemvidutide (SUBCUTANEOUS QW): 2.4mg vs 1.8mg | longitudinal_52w_primary | not_reported_at_48_80w | 0.5 | -1.16 (-3.34 to 1.01) | 0.72424 | 1.30 (0.69 to 2.44) | +52 (-63 to +198) |  |  | Moderate/uncertain probability (0.50 <= P < 0.80) that incremental weight loss exceeds threshold; individualize based on tolerability. |
| Pemvidutide (SUBCUTANEOUS QW): 2.4mg vs 1.8mg | longitudinal_52w_primary | not_reported_at_48_80w | 1 | -1.16 (-3.34 to 1.01) | 0.55761 | 1.30 (0.69 to 2.44) | +52 (-63 to +198) |  |  | Moderate/uncertain probability (0.50 <= P < 0.80) that incremental weight loss exceeds threshold; individualize based on tolerability. |
| Pemvidutide (SUBCUTANEOUS QW): 2.4mg vs 1.8mg | longitudinal_52w_primary | not_reported_at_48_80w | 2 | -1.16 (-3.34 to 1.01) | 0.22475 | 1.30 (0.69 to 2.44) | +52 (-63 to +198) |  |  | Favours maintaining lower dose: low probability (P < 0.50) that incremental weight loss exceeds the prespecified threshold. |
| Retatrutide (SUBCUTANEOUS QW): 9mg vs 4mg | longitudinal_52w_primary | long_term_42_80w | 0.5 | -5.42 (-6.70 to -4.14) | 1 |  |  | 1.63 (0.96 to 2.75) | +24 (-1 to +64) | Favours higher dose for weight loss (P >= 0.80), with imprecisely estimated AE discontinuation difference (95% CI crosses 1.0). |
| Retatrutide (SUBCUTANEOUS QW): 9mg vs 4mg | longitudinal_52w_primary | long_term_42_80w | 1 | -5.42 (-6.70 to -4.14) | 1 |  |  | 1.63 (0.96 to 2.75) | +24 (-1 to +64) | Favours higher dose for weight loss (P >= 0.80), with imprecisely estimated AE discontinuation difference (95% CI crosses 1.0). |
| Retatrutide (SUBCUTANEOUS QW): 9mg vs 4mg | longitudinal_52w_primary | long_term_42_80w | 2 | -5.42 (-6.70 to -4.14) | 1 |  |  | 1.63 (0.96 to 2.75) | +24 (-1 to +64) | Favours higher dose for weight loss (P >= 0.80), with imprecisely estimated AE discontinuation difference (95% CI crosses 1.0). |
| Retatrutide (SUBCUTANEOUS QW): 12mg vs 9mg | longitudinal_52w_primary | long_term_42_80w | 0.5 | -1.16 (-2.47 to 0.15) | 0.83927 |  |  | 1.84 (1.21 to 2.79) | +48 (+13 to +98) | Trade-off required: high probability (P >= 0.80) of incremental weight loss, but statistically significant increase in AE-related discontinuation. |
| Retatrutide (SUBCUTANEOUS QW): 12mg vs 9mg | longitudinal_52w_primary | long_term_42_80w | 1 | -1.16 (-2.47 to 0.15) | 0.59632 |  |  | 1.84 (1.21 to 2.79) | +48 (+13 to +98) | Uncertain incremental weight-loss benefit (0.50 <= P < 0.80) accompanied by significantly higher AE-related discontinuation; maintain lower dose unless greater weight loss is prioritized. |
| Retatrutide (SUBCUTANEOUS QW): 12mg vs 9mg | longitudinal_52w_primary | long_term_42_80w | 2 | -1.16 (-2.47 to 0.15) | 0.1054 |  |  | 1.84 (1.21 to 2.79) | +48 (+13 to +98) | Does not favour dose escalation: low probability (P < 0.50) of exceeding weight-loss threshold while AE-related discontinuation significantly increases. |
| Semaglutide (ORAL QD): 50mg vs 25mg | longitudinal_52w_primary | long_term_42_80w | 0.5 | -1.17 (-4.44 to 2.10) | 0.65663 | 0.86 (0.22 to 3.31) | -21 (-137 to +244) | 1.37 (0.40 to 4.70) | +23 (-40 to +189) | Moderate/uncertain probability (0.50 <= P < 0.80) that incremental weight loss exceeds threshold; individualize based on tolerability. |
| Semaglutide (ORAL QD): 50mg vs 25mg | longitudinal_52w_primary | long_term_42_80w | 1 | -1.17 (-4.44 to 2.10) | 0.54122 | 0.86 (0.22 to 3.31) | -21 (-137 to +244) | 1.37 (0.40 to 4.70) | +23 (-40 to +189) | Moderate/uncertain probability (0.50 <= P < 0.80) that incremental weight loss exceeds threshold; individualize based on tolerability. |
| Semaglutide (ORAL QD): 50mg vs 25mg | longitudinal_52w_primary | long_term_42_80w | 2 | -1.17 (-4.44 to 2.10) | 0.30995 | 0.86 (0.22 to 3.31) | -21 (-137 to +244) | 1.37 (0.40 to 4.70) | +23 (-40 to +189) | Favours maintaining lower dose: low probability (P < 0.50) that incremental weight loss exceeds the prespecified threshold. |
| Semaglutide (SUBCUTANEOUS QD): 0.1mg vs 0.05mg | longitudinal_52w_primary | w52_strict_48_56w | 0.5 | -2.46 (-4.46 to -0.46) | 0.97288 | 0.47 (0.23 to 0.97) | -115 (-180 to -6) | 1.17 (0.41 to 3.35) | +10 (-39 to +128) | Favours higher dose for weight loss (P >= 0.80), with imprecisely estimated AE discontinuation difference (95% CI crosses 1.0). |
| Semaglutide (SUBCUTANEOUS QD): 0.1mg vs 0.05mg | longitudinal_52w_primary | w52_strict_48_56w | 1 | -2.46 (-4.46 to -0.46) | 0.92427 | 0.47 (0.23 to 0.97) | -115 (-180 to -6) | 1.17 (0.41 to 3.35) | +10 (-39 to +128) | Favours higher dose for weight loss (P >= 0.80), with imprecisely estimated AE discontinuation difference (95% CI crosses 1.0). |
| Semaglutide (SUBCUTANEOUS QD): 0.1mg vs 0.05mg | longitudinal_52w_primary | w52_strict_48_56w | 2 | -2.46 (-4.46 to -0.46) | 0.67477 | 0.47 (0.23 to 0.97) | -115 (-180 to -6) | 1.17 (0.41 to 3.35) | +10 (-39 to +128) | Moderate/uncertain probability (0.50 <= P < 0.80) that incremental weight loss exceeds threshold; individualize based on tolerability. |
| Semaglutide (SUBCUTANEOUS QD): 0.2mg vs 0.1mg | longitudinal_52w_primary | w52_strict_48_56w | 0.5 | -2.83 (-4.85 to -0.81) | 0.98817 | 1.16 (0.53 to 2.51) | +18 (-59 to +148) | 0.60 (0.19 to 1.90) | -30 (-63 to +61) | Favours dose escalation: high probability (P >= 0.80) of exceeding threshold without excess discontinuation signal. |
| Semaglutide (SUBCUTANEOUS QD): 0.2mg vs 0.1mg | longitudinal_52w_primary | w52_strict_48_56w | 1 | -2.83 (-4.85 to -0.81) | 0.96226 | 1.16 (0.53 to 2.51) | +18 (-59 to +148) | 0.60 (0.19 to 1.90) | -30 (-63 to +61) | Favours dose escalation: high probability (P >= 0.80) of exceeding threshold without excess discontinuation signal. |
| Semaglutide (SUBCUTANEOUS QD): 0.2mg vs 0.1mg | longitudinal_52w_primary | w52_strict_48_56w | 2 | -2.83 (-4.85 to -0.81) | 0.79035 | 1.16 (0.53 to 2.51) | +18 (-59 to +148) | 0.60 (0.19 to 1.90) | -30 (-63 to +61) | Moderate/uncertain probability (0.50 <= P < 0.80) that incremental weight loss exceeds threshold; individualize based on tolerability. |
| Semaglutide (SUBCUTANEOUS QW): 7.2mg vs 2.4mg | longitudinal_52w_primary | long_term_42_80w | 0.5 | -2.67 (-4.14 to -1.21) | 0.99821 | 0.72 (0.33 to 1.61) | -34 (-87 to +66) | 1.57 (0.79 to 3.13) | +34 (-13 to +116) | Favours higher dose for weight loss (P >= 0.80), with imprecisely estimated AE discontinuation difference (95% CI crosses 1.0). |
| Semaglutide (SUBCUTANEOUS QW): 7.2mg vs 2.4mg | longitudinal_52w_primary | long_term_42_80w | 1 | -2.67 (-4.14 to -1.21) | 0.98756 | 0.72 (0.33 to 1.61) | -34 (-87 to +66) | 1.57 (0.79 to 3.13) | +34 (-13 to +116) | Favours higher dose for weight loss (P >= 0.80), with imprecisely estimated AE discontinuation difference (95% CI crosses 1.0). |
| Semaglutide (SUBCUTANEOUS QW): 7.2mg vs 2.4mg | longitudinal_52w_primary | long_term_42_80w | 2 | -2.67 (-4.14 to -1.21) | 0.8167 | 0.72 (0.33 to 1.61) | -34 (-87 to +66) | 1.57 (0.79 to 3.13) | +34 (-13 to +116) | Favours higher dose for weight loss (P >= 0.80), with imprecisely estimated AE discontinuation difference (95% CI crosses 1.0). |
| Survodutide (SUBCUTANEOUS QW): 6mg vs 3.6mg | longitudinal_52w_primary | long_term_42_80w | 0.5 | -0.83 (-2.81 to 1.14) | 0.62945 | 0.96 (0.39 to 2.35) | -10 (-177 to +209) | 1.05 (0.69 to 1.58) | +8 (-60 to +92) | Moderate/uncertain probability (0.50 <= P < 0.80) that incremental weight loss exceeds threshold; individualize based on tolerability. |
| Survodutide (SUBCUTANEOUS QW): 6mg vs 3.6mg | longitudinal_52w_primary | long_term_42_80w | 1 | -0.83 (-2.81 to 1.14) | 0.43416 | 0.96 (0.39 to 2.35) | -10 (-177 to +209) | 1.05 (0.69 to 1.58) | +8 (-60 to +92) | Favours maintaining lower dose: low probability (P < 0.50) that incremental weight loss exceeds the prespecified threshold. |
| Survodutide (SUBCUTANEOUS QW): 6mg vs 3.6mg | longitudinal_52w_primary | long_term_42_80w | 2 | -0.83 (-2.81 to 1.14) | 0.1234 | 0.96 (0.39 to 2.35) | -10 (-177 to +209) | 1.05 (0.69 to 1.58) | +8 (-60 to +92) | Favours maintaining lower dose: low probability (P < 0.50) that incremental weight loss exceeds the prespecified threshold. |
| Tirzepatide (SUBCUTANEOUS QW): 10mg vs 5mg | longitudinal_52w_primary | long_term_42_80w | 0.5 | -3.92 (-4.97 to -2.87) | 1 |  |  | 1.60 (0.98 to 2.60) | +24 (-1 to +61) | Favours higher dose for weight loss (P >= 0.80), with imprecisely estimated AE discontinuation difference (95% CI crosses 1.0). |
| Tirzepatide (SUBCUTANEOUS QW): 10mg vs 5mg | longitudinal_52w_primary | long_term_42_80w | 1 | -3.92 (-4.97 to -2.87) | 1 |  |  | 1.60 (0.98 to 2.60) | +24 (-1 to +61) | Favours higher dose for weight loss (P >= 0.80), with imprecisely estimated AE discontinuation difference (95% CI crosses 1.0). |
| Tirzepatide (SUBCUTANEOUS QW): 10mg vs 5mg | longitudinal_52w_primary | long_term_42_80w | 2 | -3.92 (-4.97 to -2.87) | 0.99984 |  |  | 1.60 (0.98 to 2.60) | +24 (-1 to +61) | Favours higher dose for weight loss (P >= 0.80), with imprecisely estimated AE discontinuation difference (95% CI crosses 1.0). |
| Tirzepatide (SUBCUTANEOUS QW): 15mg vs 10mg | longitudinal_52w_primary | w52_strict_48_56w | 0.5 | -1.67 (-2.66 to -0.69) | 0.99015 | 1.83 (0.68 to 4.96) | +69 (-30 to +255) | 2.58 (0.48 to 13.74) | +83 (-31 to +413) | Favours higher dose for weight loss (P >= 0.80), with imprecisely estimated AE discontinuation difference (95% CI crosses 1.0). |
| Tirzepatide (SUBCUTANEOUS QW): 15mg vs 10mg | longitudinal_52w_primary | w52_strict_48_56w | 1 | -1.67 (-2.66 to -0.69) | 0.90956 | 1.83 (0.68 to 4.96) | +69 (-30 to +255) | 2.58 (0.48 to 13.74) | +83 (-31 to +413) | Favours higher dose for weight loss (P >= 0.80), with imprecisely estimated AE discontinuation difference (95% CI crosses 1.0). |
| Tirzepatide (SUBCUTANEOUS QW): 15mg vs 10mg | longitudinal_52w_primary | w52_strict_48_56w | 2 | -1.67 (-2.66 to -0.69) | 0.25785 | 1.83 (0.68 to 4.96) | +69 (-30 to +255) | 2.58 (0.48 to 13.74) | +83 (-31 to +413) | Favours maintaining lower dose: low probability (P < 0.50) that incremental weight loss exceeds the prespecified threshold. |
| Pemvidutide (SUBCUTANEOUS QW): 2.4mg vs 1.2mg | longitudinal_52w_primary | not_reported_at_48_80w | 0.5 | -2.62 (-4.83 to -0.40) | 0.96959 | 1.47 (0.77 to 2.80) | +74 (-41 to +224) |  |  | High probability (P >= 0.80) of exceeding weight-loss threshold; AE discontinuation not reported at 48-80 weeks. |
| Pemvidutide (SUBCUTANEOUS QW): 2.4mg vs 1.2mg | longitudinal_52w_primary | not_reported_at_48_80w | 1 | -2.62 (-4.83 to -0.40) | 0.924 | 1.47 (0.77 to 2.80) | +74 (-41 to +224) |  |  | High probability (P >= 0.80) of exceeding weight-loss threshold; AE discontinuation not reported at 48-80 weeks. |
| Pemvidutide (SUBCUTANEOUS QW): 2.4mg vs 1.2mg | longitudinal_52w_primary | not_reported_at_48_80w | 2 | -2.62 (-4.83 to -0.40) | 0.70807 | 1.47 (0.77 to 2.80) | +74 (-41 to +224) |  |  | Moderate/uncertain probability (0.50 <= P < 0.80) that incremental weight loss exceeds threshold; individualize based on tolerability. |

**Table S17.** Frechet-Hoeffding bounds for responding without all-cause permanent discontinuation (arm-level matched) Computed strictly within individual randomized arms at 48-56 weeks using treatment-policy response and all-cause permanent treatment discontinuation with compatible denominators.

| Intervention | Response | Discontinuation | Matched arms | Trials | P(response) | P(discontinuation) | Lower bound | Upper bound | Bound width |
|---|---|---|---|---|---|---|---|---|---|
| Lira SC 3mg QD | >=10% weight reduction | All-cause permanent discontinuation | 1 | z017 | 0.33981 | 0.16505 | 0.17476 | 0.33981 | 0.16505 |
| Placebo | >=10% weight reduction | All-cause permanent discontinuation | 4 | z009;z017;z024;z030 | 0.10056 | 0.17037 | 0.0018622 | 0.10056 | 0.098696 |
| Placebo | >=15% weight reduction | All-cause permanent discontinuation | 3 | z009;z024;z030 | 0.022444 | 0.14604 | 0 | 0.022444 | 0.022444 |
| Sema SC 0.05mg QD | >=10% weight reduction | All-cause permanent discontinuation | 1 | z017 | 0.19417 | 0.25243 | 0 | 0.19417 | 0.19417 |
| Sema SC 0.1mg QD | >=10% weight reduction | All-cause permanent discontinuation | 1 | z017 | 0.37255 | 0.13725 | 0.23529 | 0.37255 | 0.13725 |
| Sema SC 0.2mg QD | >=10% weight reduction | All-cause permanent discontinuation | 1 | z017 | 0.56311 | 0.15534 | 0.40777 | 0.56311 | 0.15534 |
| Sema SC 0.3mg QD fast | >=10% weight reduction | All-cause permanent discontinuation | 1 | z017 | 0.57843 | 0.26471 | 0.31373 | 0.57843 | 0.26471 |
| Sema SC 0.3mg QD standard | >=10% weight reduction | All-cause permanent discontinuation | 1 | z017 | 0.58252 | 0.14563 | 0.43689 | 0.58252 | 0.14563 |
| Sema SC 0.4mg QD fast | >=10% weight reduction | All-cause permanent discontinuation | 1 | z017 | 0.71845 | 0.1165 | 0.60194 | 0.71845 | 0.1165 |
| Sema SC 0.4mg QD standard | >=10% weight reduction | All-cause permanent discontinuation | 1 | z017 | 0.64706 | 0.19608 | 0.45098 | 0.64706 | 0.19608 |
| Sema SC 2.4mg QW | >=10% weight reduction | All-cause permanent discontinuation | 2 | z009;z024 | 0.68367 | 0.14963 | 0.53361 | 0.68367 | 0.15007 |
| Sema SC 2.4mg QW | >=15% weight reduction | All-cause permanent discontinuation | 2 | z009;z024 | 0.45153 | 0.14963 | 0.30146 | 0.45153 | 0.15007 |
| TZP SC 10mg QW | >=10% weight reduction | All-cause permanent discontinuation | 1 | z030 | 0.64286 | 0.1 | 0.54286 | 0.64286 | 0.1 |
| TZP SC 10mg QW | >=15% weight reduction | All-cause permanent discontinuation | 1 | z030 | 0.45714 | 0.1 | 0.35714 | 0.45714 | 0.1 |
| TZP SC 15mg QW | >=10% weight reduction | All-cause permanent discontinuation | 1 | z030 | 0.71831 | 0.16901 | 0.5493 | 0.71831 | 0.16901 |
| TZP SC 15mg QW | >=15% weight reduction | All-cause permanent discontinuation | 1 | z030 | 0.61972 | 0.16901 | 0.4507 | 0.61972 | 0.16901 |

**Table S18.** Layer B: randomized support-increment comparisons (identical medication core)

| Outcome | Window | Population | Estimand | Comparison | k | MD (95% CI) | Conclusion |
|---|---|---|---|---|---|---|---|
| Body-weight change (kg) | 10-14 wk | without T2D | Treatment-policy | Orli 60mg TID + gsh group vs Orli PO 60mg TID | 1 | -1.15 (-6.26 to 3.96) | Sparse direct randomized evidence (single-trial contrasts); insufficient to establish a generalizable incremental support effect across medication classes. |
| Body-weight change (kg) | 10-14 wk | without T2D | Historical LOCF | shared medication + structured ww support vs shared medication standard advice | 1 | -1.78 (-3.42 to -0.14) | Sparse direct randomized evidence (single-trial contrasts); insufficient to establish a generalizable incremental support effect across medication classes. |
| Body-weight change (kg) | 20-28 wk | without T2D | Historical LOCF | Orli 120mg TID + lifestyle counselling vs Orli PO 120mg TID | 1 | -0.60 (-2.18 to 0.98) | Sparse direct randomized evidence (single-trial contrasts); insufficient to establish a generalizable incremental support effect across medication classes. |
| Body-weight change (kg) | 20-28 wk | without T2D | Completer / per-protocol | Orli 120mg TID + lifestyle counselling vs Orli PO 120mg TID | 1 | -1.00 (-2.94 to 0.94) | Sparse direct randomized evidence (single-trial contrasts); insufficient to establish a generalizable incremental support effect across medication classes. |
| Body-weight change (kg) | 48-56 wk | without T2D | Historical LOCF | Orli 120mg TID + lifestyle counselling vs Orli PO 120mg TID | 1 | 0.00 (-2.02 to 2.02) | Sparse direct randomized evidence (single-trial contrasts); insufficient to establish a generalizable incremental support effect across medication classes. |
| Body-weight change (kg) | 48-56 wk | without T2D | Completer / per-protocol | Orli 120mg TID + lifestyle counselling vs Orli PO 120mg TID | 1 | -0.60 (-3.19 to 1.99) | Sparse direct randomized evidence (single-trial contrasts); insufficient to establish a generalizable incremental support effect across medication classes. |
| % body-weight change | 10-14 wk | without T2D | Historical LOCF | shared medication + structured ww support vs shared medication standard advice | 1 | -1.64 (-3.24 to -0.04) | Sparse direct randomized evidence (single-trial contrasts); insufficient to establish a generalizable incremental support effect across medication classes. |

**Table S19.**
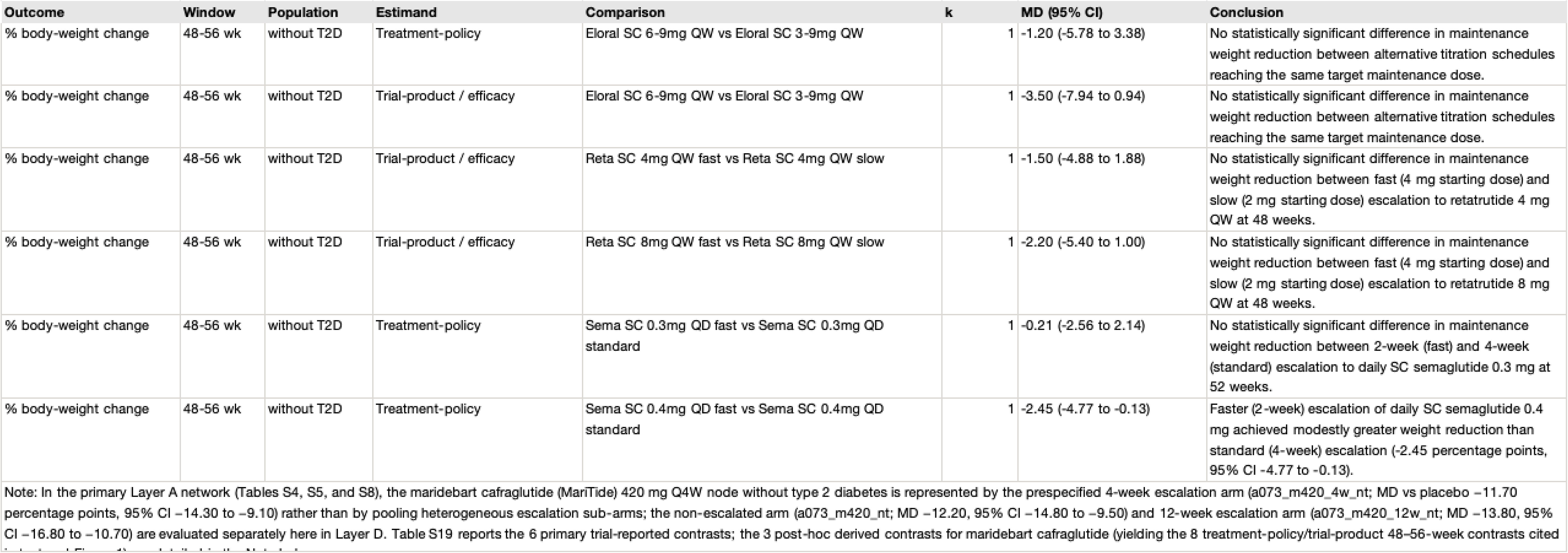
Layer D: randomized titration comparisons at an identical target maintenance dose.

| Outcome | Window | Population | Estimand | Comparison | k | MD (95% CI) | Conclusion |
|---|---|---|---|---|---|---|---|
| % body-weight change | 48-56 wk | without T2D | Treatment-policy | Eloral SC 6-9mg QW vs Eloral SC 3-9mg QW | 1 | -1.20 (-5.78 to 3.38) | No statistically significant difference in maintenance weight reduction between alternative titration schedules reaching the same target maintenance dose. |
| % body-weight change | 48-56 wk | without T2D | Trial-product / efficacy | Eloral SC 6-9mg QW vs Eloral SC 3-9mg QW | 1 | -3.50 (-7.94 to 0.94) | No statistically significant difference in maintenance weight reduction between alternative titration schedules reaching the same target maintenance dose. |
| % body-weight change | 48-56 wk | without T2D | Trial-product / efficacy | Reta SC 4mg QW fast vs Reta SC 4mg QW slow | 1 | -1.50 (-4.88 to 1.88) | No statistically significant difference in maintenance weight reduction between fast (4 mg starting dose) and slow (2 mg starting dose) escalation to retatrutide 4 mg QW at 48 weeks. |
| % body-weight change | 48-56 wk | without T2D | Trial-product / efficacy | Reta SC 8mg QW fast vs Reta SC 8mg QW slow | 1 | -2.20 (-5.40 to 1.00) | No statistically significant difference in maintenance weight reduction between fast (4 mg starting dose) and slow (2 mg starting dose) escalation to retatrutide 8 mg QW at 48 weeks. |
| % body-weight change | 48-56 wk | without T2D | Treatment-policy | Sema SC 0.3mg QD fast vs Sema SC 0.3mg QD standard | 1 | -0.21 (-2.56 to 2.14) | No statistically significant difference in maintenance weight reduction between 2-week (fast) and 4-week (standard) escalation to daily SC semaglutide 0.3 mg at 52 weeks. |
| % body-weight change | 48-56 wk | without T2D | Treatment-policy | Sema SC 0.4mg QD fast vs Sema SC 0.4mg QD standard | 1 | -2.45 (-4.77 to -0.13) | Faster (2-week) escalation of daily SC semaglutide 0.4 mg achieved modestly greater weight reduction than standard (4-week) escalation (-2.45 percentage points, 95% CI -4.77 to -0.13). |

Note: In the primary Layer A network (Tables S4, S5, and S8), the maridebart cafraglutide (MariTide) 420 mg Q4W node without type 2 diabetes is represented by the prespecified 4-week escalation arm (a073_m420_4w_nt; MD vs placebo −11.70 percentage points, 95% CI −14.30 to −9.10) rather than by pooling heterogeneous escalation sub-arms; the non-escalated arm (a073_m420_nt; MD −12.20, 95% CI −14.80 to −9.50) and 12-week escalation arm (a073_m420_12w_nt; MD −13.80, 95% CI −16.80 to −10.70) are evaluated separately here in Layer D. Table S19 reports the 6 primary trial-reported contrasts; the 3 post-hoc derived contrasts for maridebart cafraglutide (yielding the 8 treatment-policy/trial-product 48–56-week contrasts cited [illegible]

**Table S20.**
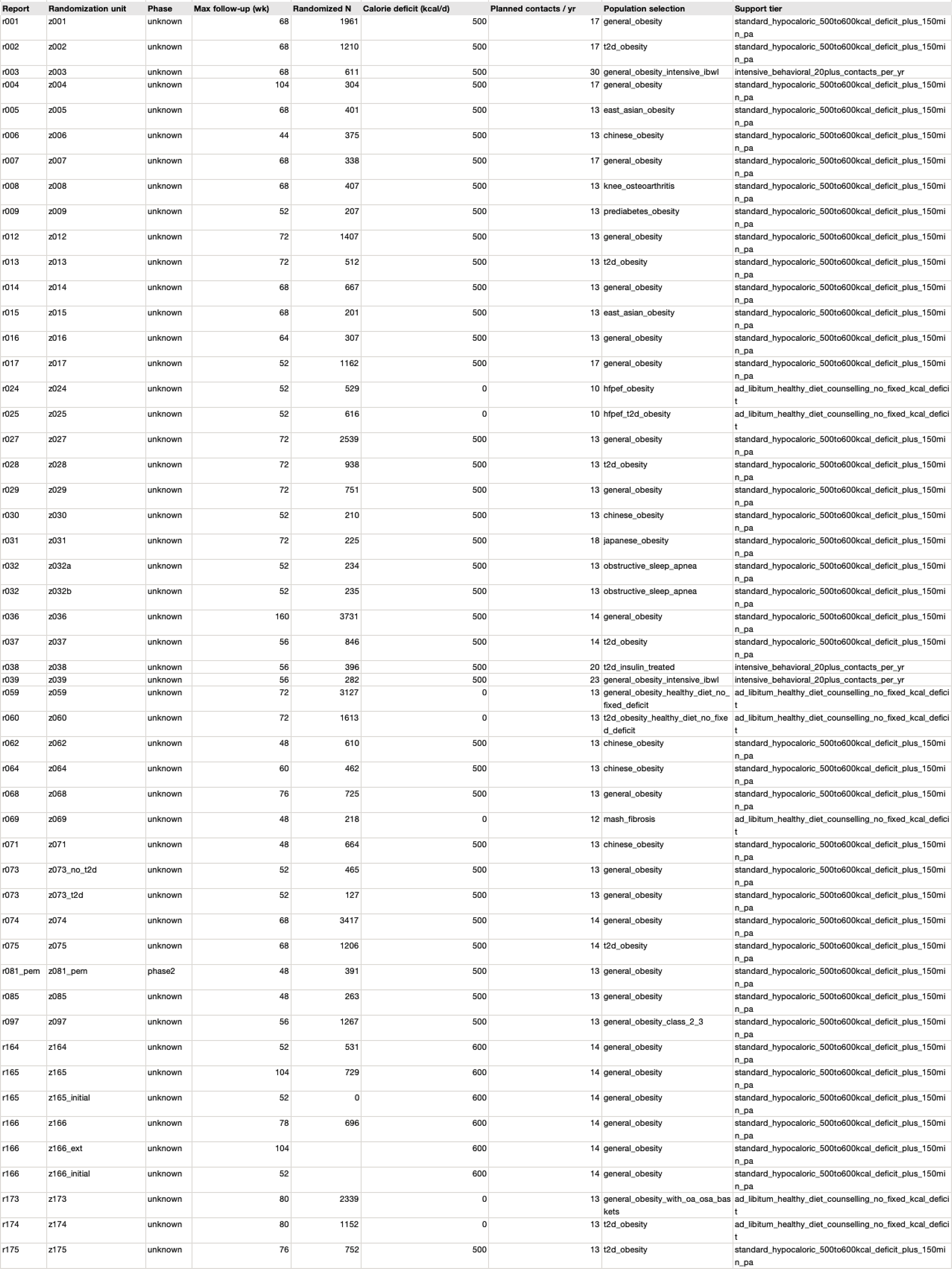
Layer C: cross-trial background lifestyle support characteristics. Cross-trial background lifestyle intensity descriptor (layer C). Because background support is colinear with trial family and era, Layer C meta-regression is underpowered and reported descriptivety.

| Report | Randomization unit | Phase | Max follow-up (wk) | Randomized N | Calorie deficit (kcal/d) | Planned contacts / yr | Population selection | Support tier |
|---|---|---|---|---|---|---|---|---|
| r001 | z001 | unknown | 68 | 1961 | 500 | 17 | general_obesity | standard_hypocaloric_500to600kcal_deficit_plus_150min_pa |
| r002 | z002 | unknown | 68 | 1210 | 500 | 17 | t2d_obesity | standard_hypocaloric_500to600kcal_deficit_plus_150min_pa |
| r003 | z003 | unknown | 68 | 611 | 500 | 30 | general_obesity_intensive_ibwl | intensive_behavioral_20plus_contacts_per_yr |
| r004 | z004 | unknown | 104 | 304 | 500 | 17 | general_obesity | standard_hypocaloric_500to600kcal_deficit_plus_150min_pa |
| r005 | z005 | unknown | 68 | 401 | 500 | 13 | east_asian_obesity | standard_hypocaloric_500to600kcal_deficit_plus_150min_pa |
| r006 | z006 | unknown | 44 | 375 | 500 | 13 | chinese_obesity | standard_hypocaloric_500to600kcal_deficit_plus_150min_pa |
| r007 | z007 | unknown | 68 | 338 | 500 | 17 | general_obesity | standard_hypocaloric_500to600kcal_deficit_plus_150min_pa |
| r008 | z008 | unknown | 68 | 407 | 500 | 13 | knee_osteoarthritis | standard_hypocaloric_500to600kcal_deficit_plus_150min_pa |
| r009 | z009 | unknown | 52 | 207 | 500 | 13 | prediabetes_obesity | standard_hypocaloric_500to600kcal_deficit_plus_150min_pa |
| r012 | z012 | unknown | 72 | 1407 | 500 | 13 | general_obesity | standard_hypocaloric_500to600kcal_deficit_plus_150min_pa |
| r013 | z013 | unknown | 72 | 512 | 500 | 13 | t2d_obesity | standard_hypocaloric_500to600kcal_deficit_plus_150min_pa |
| r014 | z014 | unknown | 68 | 667 | 500 | 13 | general_obesity | standard_hypocaloric_500to600kcal_deficit_plus_150min_pa |
| r015 | z015 | unknown | 68 | 201 | 500 | 13 | east_asian_obesity | standard_hypocaloric_500to600kcal_deficit_plus_150min_pa |
| r016 | z016 | unknown | 64 | 307 | 500 | 13 | general_obesity | standard_hypocaloric_500to600kcal_deficit_plus_150min_pa |
| r017 | z017 | unknown | 52 | 1162 | 500 | 17 | general_obesity | standard_hypocaloric_500to600kcal_deficit_plus_150min_pa |
| r024 | z024 | unknown | 52 | 529 | 0 | 10 | hfpef_obesity | ad_libitum_healthy_diet_counselling_no_fixed_kcal_deficit |
| r025 | z025 | unknown | 52 | 616 | 0 | 10 | hfpef_t2d_obesity | ad_libitum_healthy_diet_counselling_no_fixed_kcal_deficit |
| r027 | z027 | unknown | 72 | 2539 | 500 | 13 | general_obesity | standard_hypocaloric_500to600kcal_deficit_plus_150min_pa |
| r028 | z028 | unknown | 72 | 938 | 500 | 13 | t2d_obesity | standard_hypocaloric_500to600kcal_deficit_plus_150min_pa |
| r029 | z029 | unknown | 72 | 751 | 500 | 13 | general_obesity | standard_hypocaloric_500to600kcal_deficit_plus_150min_pa |
| r030 | z030 | unknown | 52 | 210 | 500 | 13 | chinese_obesity | standard_hypocaloric_500to600kcal_deficit_plus_150min_pa |
| r031 | z031 | unknown | 72 | 225 | 500 | 18 | japanese_obesity | standard_hypocaloric_500to600kcal_deficit_plus_150min_pa |
| r032 | z032a | unknown | 52 | 234 | 500 | 13 | obstructive_sleep_apnea | standard_hypocaloric_500to600kcal_deficit_plus_150min_pa |
| r032 | z032b | unknown | 52 | 235 | 500 | 13 | obstructive_sleep_apnea | standard_hypocaloric_500to600kcal_deficit_plus_150min_pa |
| r036 | z036 | unknown | 160 | 3731 | 500 | 14 | general_obesity | standard_hypocaloric_500to600kcal_deficit_plus_150min_pa |
| r037 | z037 | unknown | 56 | 846 | 500 | 14 | t2d_obesity | standard_hypocaloric_500to600kcal_deficit_plus_150min_pa |
| r038 | z038 | unknown | 56 | 396 | 500 | 20 | t2d_insulin_treated | intensive_behavioral_20plus_contacts_per_yr |
| r039 | z039 | unknown | 56 | 282 | 500 | 23 | general_obesity_intensive_ibwl | intensive_behavioral_20plus_contacts_per_yr |
| r059 | z059 | unknown | 72 | 3127 | 0 | 13 | general_obesity_healthy_diet_no_fixed_deficit | ad_libitum_healthy_diet_counselling_no_fixed_kcal_deficit |
| r060 | z060 | unknown | 72 | 1613 | 0 | 13 | t2d_obesity_healthy_diet_no_fixed_deficit | ad_libitum_healthy_diet_counselling_no_fixed_kcal_deficit |
| r062 | z062 | unknown | 48 | 610 | 500 | 13 | chinese_obesity | standard_hypocaloric_500to600kcal_deficit_plus_150min_pa |
| r064 | z064 | unknown | 60 | 462 | 500 | 13 | chinese_obesity | standard_hypocaloric_500to600kcal_deficit_plus_150min_pa |
| r068 | z068 | unknown | 76 | 725 | 500 | 13 | general_obesity | standard_hypocaloric_500to600kcal_deficit_plus_150min_pa |
| r069 | z069 | unknown | 48 | 218 | 0 | 12 | mash_fibrosis | ad_libitum_healthy_diet_counselling_no_fixed_kcal_deficit |
| r071 | z071 | unknown | 48 | 664 | 500 | 13 | chinese_obesity | standard_hypocaloric_500to600kcal_deficit_plus_150min_pa |
| r073 | z073_no_t2d | unknown | 52 | 465 | 500 | 13 | general_obesity | standard_hypocaloric_500to600kcal_deficit_plus_150min_pa |
| r073 | z073_t2d | unknown | 52 | 127 | 500 | 13 | general_obesity | standard_hypocaloric_500to600kcal_deficit_plus_150min_pa |
| r074 | z074 | unknown | 68 | 3417 | 500 | 14 | general_obesity | standard_hypocaloric_500to600kcal_deficit_plus_150min_pa |
| r075 | z075 | unknown | 68 | 1206 | 500 | 14 | t2d_obesity | standard_hypocaloric_500to600kcal_deficit_plus_150min_pa |
| r081_pem | z081_pem | phase2 | 48 | 391 | 500 | 13 | general_obesity | standard_hypocaloric_500to600kcal_deficit_plus_150min_pa |
| r085 | z085 | unknown | 48 | 263 | 500 | 13 | general_obesity | standard_hypocaloric_500to600kcal_deficit_plus_150min_pa |
| r097 | z097 | unknown | 56 | 1267 | 500 | 13 | general_obesity_class_2_3 | standard_hypocaloric_500to600kcal_deficit_plus_150min_pa |
| r164 | z164 | unknown | 52 | 531 | 600 | 14 | general_obesity | standard_hypocaloric_500to600kcal_deficit_plus_150min_pa |
| r165 | z165 | unknown | 104 | 729 | 600 | 14 | general_obesity | standard_hypocaloric_500to600kcal_deficit_plus_150min_pa |
| r165 | z165_initial | unknown | 52 | 0 | 600 | 14 | general_obesity | standard_hypocaloric_500to600kcal_deficit_plus_150min_pa |
| r166 | z166 | unknown | 78 | 696 | 600 | 14 | general_obesity | standard_hypocaloric_500to600kcal_deficit_plus_150min_pa |
| r166 | z166_ext | unknown | 104 |  | 600 | 14 | general_obesity | standard_hypocaloric_500to600kcal_deficit_plus_150min_pa |
| r166 | z166_initial | unknown | 52 |  | 600 | 14 | general_obesity | standard_hypocaloric_500to600kcal_deficit_plus_150min_pa |
| r173 | z173 | unknown | 80 | 2339 | 0 | 13 | general_obesity_with_oa_osa_baskets | ad_libitum_healthy_diet_counselling_no_fixed_kcal_deficit |
| r174 | z174 | unknown | 80 | 1152 | 0 | 13 | t2d_obesity | ad_libitum_healthy_diet_counselling_no_fixed_kcal_deficit |
| r175 | z175 | unknown | 76 | 752 | 500 | 13 | t2d_obesity | standard_hypocaloric_500to600kcal_deficit_plus_150min_pa |

**Table S21.**
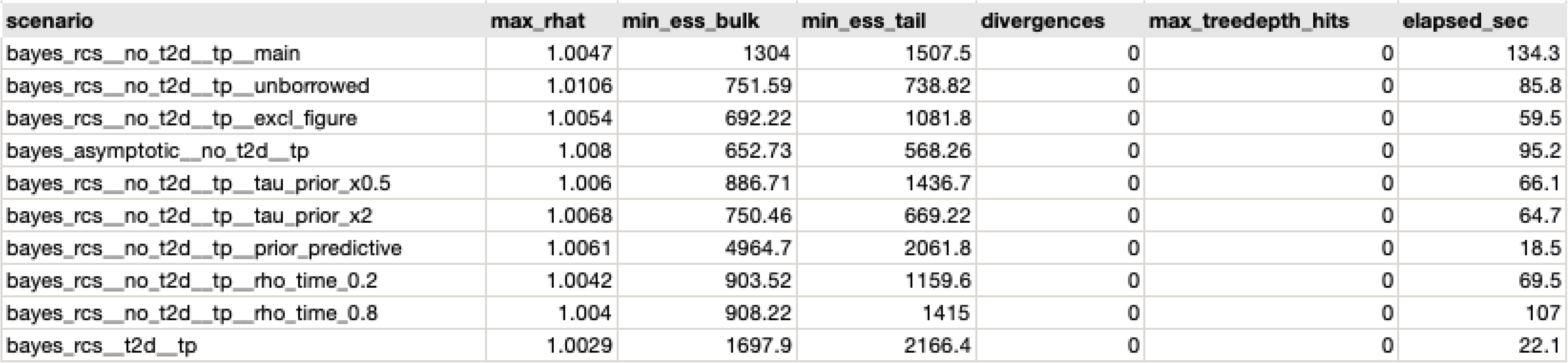
Longitudinal model convergence diagnostics.

**Table S22.** Sources of within-trial covariances. Within-trial covariances between active contrasts sharing a common comparator arm were obtained from: (1) exact shared control-arm variance when arm-level SEs were reported (shared_arm_variance_exact); (2) exact algebraic recovery from reported active-versus-active and active-versus-placebo contrast variances Var(A-C)+Var(B-C)-Var(A-B) (recovered_from_three_contrast_variances): (3) study-reported covariance (reported); or (4) prespecified default correlation rho = 0.50 (sensitivity range 0.30 to 0.70 in Table S12) when neither arm-level SEs nor head-to-head active contrasts were reported.

| Source | Pairs |
|---|---|
| assumed_rho_0.5 | 134 |
| recovered_from_three_contrast_variances | 8 |
| reported | 15 |
| shared_arm_variance_exact | 65 |

**Table S23.**
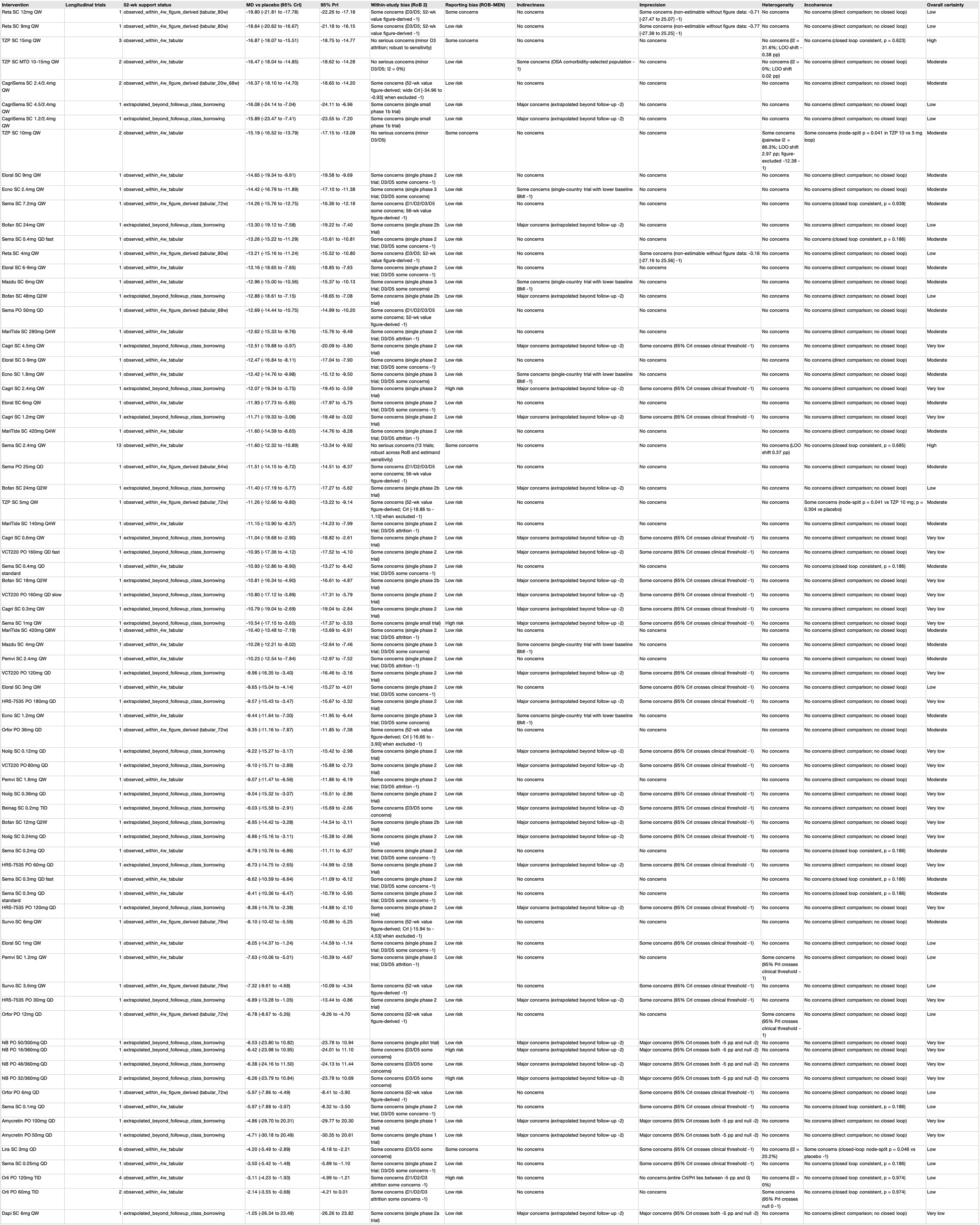
CINeMA and GRADE certainty-of-evidence assessment across six domains for primary 52-week weight reduction. Assessed per protocol section 37 across six CINeMA/GRADE domains using a clinically important threshold of −5 percentage points versus placebo.

| Intervention | Longitudinal trials | 52-wk support status | MD vs placebo (95% CrI) | 95% PrI | Within-study bias (RoB 2) | Reporting bias (ROB-MEN) | Indirectness | Imprecision | Heterogeneity | Incoherence | Overall certainty |
|---|---|---|---|---|---|---|---|---|---|---|---|
| Reta SC 12mg QW | 1 | observed_within_4w_figure_derived (tabular_80w) | -19.80 (-21.81 to -17.78) | -22.26 to -17.18 | Some concerns (D3/D5; 52-wk value figure-derived -1) | Some concerns | No concerns | Some concerns (non-estimable without figure data: -0.71 [-27.47 to 25.07] -1) | No concerns | No concerns (direct comparison; no closed loop) | Low |
| Reta SC 9mg QW | 1 | observed_within_4w_figure_derived (tabular_80w) | -18.64 (-20.62 to -16.67) | -21.18 to -16.15 | Some concerns (D3/D5; 52-wk value figure-derived -1) | Low risk | No concerns | Some concerns (non-estimable without figure data: -0.77 [-27.38 to 25.25] -1) | No concerns | No concerns (direct comparison; no closed loop) | Low |
| TZP SC 15mg QW | 3 | observed_within_4w_tabular | -16.87 (-18.07 to -15.51) | -18.75 to -14.77 | No serious concerns (minor D3 attrition; robust to sensitivity) | Some concerns | No concerns | No concerns | No concerns (I2 = 31.6%; LOO shift 0.38 pp) | No concerns (closed loop consistent, p = 0.623) | High |
| TZP SC MTD 10-15mg QW | 2 | observed_within_4w_tabular | -16.47 (-18.04 to -14.85) | -18.62 to -14.28 | No serious concerns (minor D3/D5; I2 = 0%) | Low risk | Some concerns (OSA comorbidity-selected population -1) | No concerns | No concerns (I2 = 0%; LOO shift 0.02 pp) | No concerns (direct comparison; no closed loop) | Moderate |
| CagriSema SC 2.4/2.4mg QW | 2 | observed_within_4w_figure_derived (tabular_20w_68w) | -16.37 (-18.10 to -14.70) | -18.65 to -14.20 | Some concerns (52-wk value figure-derived; wide CrI [-34.96 to -0.93] when excluded -1) | Low risk | No concerns | No concerns | No concerns | No concerns (direct comparison; no closed loop) | Moderate |
| CagriSema SC 4.5/2.4mg QW | 1 | extrapolated_beyond_followup_class_borrowing | -16.08 (-24.14 to -7.04) | -24.11 to -6.96 | Some concerns (single small phase 1b trial) | Low risk | Major concerns (extrapolated beyond follow-up -2) | No concerns | No concerns | No concerns (direct comparison; no closed loop) | Low |
| CagriSema SC 1.2/2.4mg QW | 1 | extrapolated_beyond_followup_class_borrowing | -15.89 (-23.47 to -7.41) | -23.55 to -7.20 | Some concerns (single small phase 1b trial) | Low risk | Major concerns (extrapolated beyond follow-up -2) | No concerns | No concerns | No concerns (direct comparison; no closed loop) | Low |
| TZP SC 10mg QW | 2 | observed_within_4w_tabular | -15.19 (-16.52 to -13.79) | -17.15 to -13.09 | No serious concerns (minor D3/D5) | Some concerns | No concerns | No concerns | Some concerns (pairwise I2 = 86.3%; LOO shift 2.97 pp; figure-excluded -12.38 -1) | Some concerns (node-split p = 0.041 in TZP 10 vs 5 mg loop) | Moderate |
| Eloral SC 9mg QW | 1 | observed_within_4w_tabular | -14.65 (-19.34 to -9.91) | -19.58 to -9.69 | Some concerns (single phase 2 trial; D3/D5 some concerns -1) | Low risk | No concerns | No concerns | No concerns | No concerns (direct comparison; no closed loop) | Moderate |
| Ecno SC 2.4mg QW | 1 | observed_within_4w_tabular | -14.42 (-16.79 to -11.89) | -17.10 to -11.38 | Some concerns (single phase 3 trial; D3/D5 some concerns) | Low risk | Some concerns (single-country trial with lower baseline BMI -1) | No concerns | No concerns | No concerns (direct comparison; no closed loop) | Moderate |
| Sema SC 7.2mg QW | 1 | observed_within_4w_figure_derived (tabular_72w) | -14.26 (-15.76 to -12.75) | -16.36 to -12.18 | Some concerns (D1/D2/D3/D5 some concerns; 56-wk value figure-derived -1) | Low risk | No concerns | No concerns | No concerns | No concerns (closed loop consistent, p = 0.939) | Moderate |
| Bofan SC 24mg QW | 1 | extrapolated_beyond_followup_class_borrowing | -13.30 (-19.12 to -7.58) | -19.22 to -7.40 | Some concerns (single phase 2b trial) | Low risk | Major concerns (extrapolated beyond follow-up -2) | No concerns | No concerns | No concerns (direct comparison; no closed loop) | Low |
| Sema SC 0.4mg QD fast | 1 | observed_within_4w_tabular | -13.26 (-15.22 to -11.29) | -15.61 to -10.81 | Some concerns (single phase 2 trial; D3/D5 some concerns -1) | Low risk | No concerns | No concerns | No concerns | No concerns (closed loop consistent, p = 0.186) | Moderate |
| Reta SC 4mg QW | 1 | observed_within_4w_figure_derived (tabular_80w) | -13.21 (-15.16 to -11.24) | -15.52 to -10.80 | Some concerns (D3/D5; 52-wk value figure-derived -1) | Low risk | No concerns | Some concerns (non-estimable without figure data: -0.16 [-27.16 to 25.56] -1) | No concerns | No concerns (direct comparison; no closed loop) | Low |
| Eloral SC 6-9mg QW | 1 | observed_within_4w_tabular | -13.16 (-18.65 to -7.65) | -18.85 to -7.63 | Some concerns (single phase 2 trial; D3/D5 some concerns -1) | Low risk | No concerns | No concerns | No concerns | No concerns (direct comparison; no closed loop) | Moderate |
| Mazdu SC 6mg QW | 1 | observed_within_4w_tabular | -12.96 (-15.00 to -10.56) | -15.37 to -10.13 | Some concerns (single phase 3 trial; D3/D5 some concerns) | Low risk | Some concerns (single-country trial with lower baseline BMI -1) | No concerns | No concerns | No concerns (direct comparison; no closed loop) | Moderate |
| Bofan SC 48mg Q2W | 1 | extrapolated_beyond_followup_class_borrowing | -12.88 (-18.61 to -7.15) | -18.65 to -7.08 | Some concerns (single phase 2b trial) | Low risk | Major concerns (extrapolated beyond follow-up -2) | No concerns | No concerns | No concerns (direct comparison; no closed loop) | Low |
| Sema PO 50mg QD | 1 | observed_within_4w_figure_derived (tabular_68w) | -12.69 (-14.44 to -10.75) | -14.99 to -10.20 | Some concerns (D1/D2/D3/D5 some concerns; 52-wk value figure-derived -1) | Low risk | No concerns | No concerns | No concerns | No concerns (direct comparison; no closed loop) | Moderate |
| MariTide SC 280mg Q4W | 1 | observed_within_4w_tabular | -12.62 (-15.33 to -9.76) | -15.76 to -9.49 | Some concerns (single phase 2 trial; D3/D5 attrition -1) | Low risk | No concerns | No concerns | No concerns | No concerns (direct comparison; no closed loop) | Moderate |
| Cagri SC 4.5mg QW | 1 | extrapolated_beyond_followup_class_borrowing | -12.51 (-19.88 to -3.97) | -20.09 to -3.80 | Some concerns (single phase 2 trial) | Low risk | Major concerns (extrapolated beyond follow-up -2) | Some concerns (95% CrI crosses clinical threshold -1) | No concerns | No concerns (direct comparison; no closed loop) | Very low |
| Eloral SC 3-9mg QW | 1 | observed_within_4w_tabular | -12.47 (-16.84 to -8.11) | -17.04 to -7.90 | Some concerns (single phase 2 trial; D3/D5 some concerns -1) | Low risk | No concerns | No concerns | No concerns | No concerns (direct comparison; no closed loop) | Moderate |
| Ecno SC 1.8mg QW | 1 | observed_within_4w_tabular | -12.42 (-14.76 to -9.98) | -15.12 to -9.50 | Some concerns (single phase 3 trial; D3/D5 some concerns) | Low risk | Some concerns (single-country trial with lower baseline BMI -1) | No concerns | No concerns | No concerns (direct comparison; no closed loop) | Moderate |
| Cagri SC 2.4mg QW | 1 | extrapolated_beyond_followup_class_borrowing | -12.07 (-19.34 to -3.75) | -19.45 to -3.59 | Some concerns (single phase 2 trial) | High risk | Major concerns (extrapolated beyond follow-up -2) | Some concerns (95% CrI crosses clinical threshold -1) | No concerns | No concerns (direct comparison; no closed loop) | Very low |
| Eloral SC 6mg QW | 1 | observed_within_4w_tabular | -11.93 (-17.73 to -5.85) | -17.97 to -5.75 | Some concerns (single phase 2 trial; D3/D5 some concerns -1) | Low risk | No concerns | No concerns | No concerns | No concerns (direct comparison; no closed loop) | Moderate |
| Cagri SC 1.2mg QW | 1 | extrapolated_beyond_followup_class_borrowing | -11.71 (-19.33 to -3.06) | -19.48 to -3.02 | Some concerns (single phase 2 trial) | Low risk | Major concerns (extrapolated beyond follow-up -2) | Some concerns (95% CrI crosses clinical threshold -1) | No concerns | No concerns (direct comparison; no closed loop) | Very low |
| MariTide SC 420mg Q4W | 1 | observed_within_4w_tabular | -11.60 (-14.39 to -8.65) | -14.76 to -8.28 | Some concerns (single phase 2 trial; D3/D5 attrition -1) | Low risk | No concerns | No concerns | No concerns | No concerns (direct comparison; no closed loop) | Moderate |
| Sema SC 2.4mg QW | 13 | observed_within_4w_tabular | -11.60 (-12.32 to -10.89) | -13.34 to -9.92 | No serious concerns (13 trials; robust across RoB and estimand sensitivity) | Some concerns | No concerns | No concerns | No concerns (LOO shift 0.37 pp) | No concerns (closed loop consistent, p = 0.685) | High |
| Sema PO 25mg QD | 1 | observed_within_4w_figure_derived (tabular_64w) | -11.51 (-14.15 to -8.72) | -14.51 to -8.37 | Some concerns (D1/D2/D3/D5 some concerns; 56-wk value figure-derived -1) | Low risk | No concerns | No concerns | No concerns | No concerns (direct comparison; no closed loop) | Moderate |
| Bofan SC 24mg Q2W | 1 | extrapolated_beyond_followup_class_borrowing | -11.40 (-17.19 to -5.77) | -17.27 to -5.62 | Some concerns (single phase 2b trial) | Low risk | Major concerns (extrapolated beyond follow-up -2) | No concerns | No concerns | No concerns (direct comparison; no closed loop) | Low |
| TZP SC 5mg QW | 1 | observed_within_4w_figure_derived (tabular_72w) | -11.26 (-12.66 to -9.80) | -13.22 to -9.14 | Some concerns (52-wk value figure-derived; CrI [-18.86 to -1.10] when excluded -1) | Low risk | No concerns | No concerns | No concerns | Some concerns (node-split p = 0.041 vs TZP 10 mg; p = 0.304 vs placebo) | Moderate |
| MariTide SC 140mg Q4W | 1 | observed_within_4w_tabular | -11.15 (-13.90 to -8.37) | -14.23 to -7.99 | Some concerns (single phase 2 trial; D3/D5 attrition -1) | Low risk | No concerns | No concerns | No concerns | No concerns (direct comparison; no closed loop) | Moderate |
| Cagri SC 0.6mg QW | 1 | extrapolated_beyond_followup_class_borrowing | -11.04 (-18.68 to -2.90) | -18.82 to -2.61 | Some concerns (single phase 2 trial) | Low risk | Major concerns (extrapolated beyond follow-up -2) | Some concerns (95% CrI crosses clinical threshold -1) | No concerns | No concerns (direct comparison; no closed loop) | Very low |
| VCT220 PO 160mg QD fast | 1 | extrapolated_beyond_followup_class_borrowing | -10.95 (-17.36 to -4.12) | -17.52 to -4.10 | Some concerns (single phase 2 trial) | Low risk | Major concerns (extrapolated beyond follow-up -2) | Some concerns (95% CrI crosses clinical threshold -1) | No concerns | No concerns (direct comparison; no closed loop) | Very low |
| Sema SC 0.4mg QD standard | 1 | observed_within_4w_tabular | -10.93 (-12.86 to -8.90) | -13.27 to -8.42 | Some concerns (single phase 2 trial; D3/D5 some concerns -1) | Low risk | No concerns | No concerns | No concerns | No concerns (closed loop consistent, p = 0.186) | Moderate |
| Bofan SC 18mg Q2W | 1 | extrapolated_beyond_followup_class_borrowing | -10.81 (-16.34 to -4.90) | -16.61 to -4.87 | Some concerns (single phase 2b trial) | Low risk | Major concerns (extrapolated beyond follow-up -2) | Some concerns (95% CrI crosses clinical threshold -1) | No concerns | No concerns (direct comparison; no closed loop) | Very low |
| VCT220 PO 160mg QD slow | 1 | extrapolated_beyond_followup_class_borrowing | -10.80 (-17.12 to -3.89) | -17.31 to -3.79 | Some concerns (single phase 2 trial) | Low risk | Major concerns (extrapolated beyond follow-up -2) | Some concerns (95% CrI crosses clinical threshold -1) | No concerns | No concerns (direct comparison; no closed loop) | Very low |
| Cagri SC 0.3mg QW | 1 | extrapolated_beyond_followup_class_borrowing | -10.79 (-19.04 to -2.69) | -19.04 to -2.84 | Some concerns (single phase 2 trial) | Low risk | Major concerns (extrapolated beyond follow-up -2) | Some concerns (95% CrI crosses clinical threshold -1) | No concerns | No concerns (direct comparison; no closed loop) | Very low |
| Sema SC 1mg QW | 1 | extrapolated_beyond_followup_class_borrowing | -10.54 (-17.15 to -3.65) | -17.37 to -3.53 | Some concerns (single small trial) | High risk | Major concerns (extrapolated beyond follow-up -2) | Some concerns (95% CrI crosses clinical threshold -1) | No concerns | No concerns (direct comparison; no closed loop) | Very low |
| MariTide SC 420mg Q8W | 1 | observed_within_4w_tabular | -10.40 (-13.48 to -7.19) | -13.69 to -6.91 | Some concerns (single phase 2 trial; D3/D5 attrition -1) | Low risk | No concerns | No concerns | No concerns | No concerns (direct comparison; no closed loop) | Moderate |
| Mazdu SC 4mg QW | 1 | observed_within_4w_tabular | -10.28 (-12.21 to -8.02) | -12.64 to -7.46 | Some concerns (single phase 3 trial; D3/D5 some concerns) | Low risk | Some concerns (single-country trial with lower baseline BMI -1) | No concerns | No concerns | No concerns (direct comparison; no closed loop) | Moderate |
| Pemvi SC 2.4mg QW | 1 | observed_within_4w_tabular | -10.23 (-12.54 to -7.84) | -12.97 to -7.52 | Some concerns (single phase 2 trial; D3/D5 attrition -1) | Low risk | No concerns | No concerns | No concerns | No concerns (direct comparison; no closed loop) | Moderate |
| VCT220 PO 120mg QD | 1 | extrapolated_beyond_followup_class_borrowing | -9.96 (-16.35 to -3.40) | -16.46 to -3.16 | Some concerns (single phase 2 trial) | Low risk | Major concerns (extrapolated beyond follow-up -2) | Some concerns (95% CrI crosses clinical threshold -1) | No concerns | No concerns (direct comparison; no closed loop) | Very low |
| Eloral SC 3mg QW | 1 | observed_within_4w_tabular | -9.65 (-15.04 to -4.14) | -15.27 to -4.01 | Some concerns (single phase 2 trial; D3/D5 some concerns -1) | Low risk | No concerns | Some concerns (95% CrI crosses clinical threshold -1) | No concerns | No concerns (direct comparison; no closed loop) | Low |
| HRS-7535 PO 180mg QD | 1 | extrapolated_beyond_followup_class_borrowing | -9.57 (-15.43 to -3.47) | -15.67 to -3.32 | Some concerns (single phase 2 trial) | Low risk | Major concerns (extrapolated beyond follow-up -2) | Some concerns (95% CrI crosses clinical threshold -1) | No concerns | No concerns (direct comparison; no closed loop) | Very low |
| Ecno SC 1.2mg QW | 1 | observed_within_4w_tabular | -9.44 (-11.64 to -7.00) | -11.95 to -6.44 | Some concerns (single phase 3 trial; D3/D5 some concerns) | Low risk | Some concerns (single-country trial with lower baseline BMI -1) | No concerns | No concerns | No concerns (direct comparison; no closed loop) | Moderate |
| Orfor PO 36mg QD | 1 | observed_within_4w_figure_derived (tabular_72w) | -9.35 (-11.16 to -7.87) | -11.85 to -7.38 | Some concerns (52-wk value figure-derived; CrI [-16.66 to -3.90] when excluded -1) | Low risk | No concerns | No concerns | No concerns | No concerns (direct comparison; no closed loop) | Moderate |
| Noiig SC 0.12mg QD | 1 | extrapolated_beyond_followup_class_borrowing | -9.22 (-15.27 to -3.17) | -15.42 to -2.98 | Some concerns (single phase 2 trial) | Low risk | Major concerns (extrapolated beyond follow-up -2) | Some concerns (95% CrI crosses clinical threshold -1) | No concerns | No concerns (direct comparison; no closed loop) | Very low |
| VCT220 PO 80mg QD | 1 | extrapolated_beyond_followup_class_borrowing | -9.10 (-15.71 to -2.89) | -15.88 to -2.73 | Some concerns (single phase 2 trial) | Low risk | Major concerns (extrapolated beyond follow-up -2) | Some concerns (95% CrI crosses clinical threshold -1) | No concerns | No concerns (direct comparison; no closed loop) | Very low |
| Pemvi SC 1.8mg QW | 1 | observed_within_4w_tabular | -9.07 (-11.47 to -6.56) | -11.86 to -6.19 | Some concerns (single phase 2 trial; D3/D5 attrition -1) | Low risk | No concerns | No concerns | No concerns | No concerns (direct comparison; no closed loop) | Moderate |
| Noiig SC 0.36mg QD | 1 | extrapolated_beyond_followup_class_borrowing | -9.04 (-15.32 to -3.07) | -15.51 to -2.86 | Some concerns (single phase 2 trial) | Low risk | Major concerns (extrapolated beyond follow-up -2) | Some concerns (95% CrI crosses clinical threshold -1) | No concerns | No concerns (direct comparison; no closed loop) | Very low |
| Beinag SC 0.2mg TID | 1 | extrapolated_beyond_followup_class_borrowing | -9.03 (-15.58 to -2.91) | -15.69 to -2.66 | Some concerns (D3/D5 some concerns) | Low risk | Major concerns (extrapolated beyond follow-up -2) | Some concerns (95% CrI crosses clinical threshold -1) | No concerns | No concerns (direct comparison; no closed loop) | Very low |
| Bofan SC 12mg Q2W | 1 | extrapolated_beyond_followup_class_borrowing | -8.95 (-14.42 to -3.28) | -14.54 to -3.11 | Some concerns (single phase 2b trial) | Low risk | Major concerns (extrapolated beyond follow-up -2) | Some concerns (95% CrI crosses clinical threshold -1) | No concerns | No concerns (direct comparison; no closed loop) | Very low |
| Noiig SC 0.24mg QD | 1 | extrapolated_beyond_followup_class_borrowing | -8.86 (-15.16 to -3.11) | -15.38 to -2.86 | Some concerns (single phase 2 trial) | Low risk | Major concerns (extrapolated beyond follow-up -2) | Some concerns (95% CrI crosses clinical threshold -1) | No concerns | No concerns (direct comparison; no closed loop) | Very low |
| Sema SC 0.2mg QD | 1 | observed_within_4w_tabular | -8.79 (-10.76 to -6.86) | -11.11 to -6.37 | Some concerns (single phase 2 trial; D3/D5 some concerns -1) | Low risk | No concerns | No concerns | No concerns | No concerns (closed loop consistent, p = 0.186) | Moderate |
| HRS-7535 PO 60mg QD | 1 | extrapolated_beyond_followup_class_borrowing | -8.73 (-14.75 to -2.65) | -14.99 to -2.58 | Some concerns (single phase 2 trial) | Low risk | Major concerns (extrapolated beyond follow-up -2) | Some concerns (95% CrI crosses clinical threshold -1) | No concerns | No concerns (direct comparison; no closed loop) | Very low |
| Sema SC 0.3mg QD fast | 1 | observed_within_4w_tabular | -8.62 (-10.59 to -6.64) | -11.09 to -6.12 | Some concerns (single phase 2 trial; D3/D5 some concerns -1) | Low risk | No concerns | No concerns | No concerns | No concerns (closed loop consistent, p = 0.186) | Moderate |
| Sema SC 0.3mg QD standard | 1 | observed_within_4w_tabular | -8.41 (-10.36 to -6.47) | -10.78 to -5.95 | Some concerns (single phase 2 trial; D3/D5 some concerns -1) | Low risk | No concerns | No concerns | No concerns | No concerns (closed loop consistent, p = 0.186) | Moderate |
| HRS-7535 PO 120mg QD | 1 | extrapolated_beyond_followup_class_borrowing | -8.36 (-14.76 to -2.38) | -14.88 to -2.10 | Some concerns (single phase 2 trial) | Low risk | Major concerns (extrapolated beyond follow-up -2) | Some concerns (95% CrI crosses clinical threshold -1) | No concerns | No concerns (direct comparison; no closed loop) | Very low |
| Survo SC 6mg QW | 1 | observed_within_4w_figure_derived (tabular_76w) | -8.10 (-10.42 to -5.56) | -10.86 to -5.25 | Some concerns (52-wk value figure-derived; CrI [-15.94 to -4.53] when excluded -1) | Low risk | No concerns | No concerns | No concerns | No concerns (direct comparison; no closed loop) | Moderate |
| Eloral SC 1mg QW | 1 | observed_within_4w_tabular | -8.05 (-14.37 to -1.24) | -14.59 to -1.14 | Some concerns (single phase 2 trial; D3/D5 some concerns -1) | Low risk | No concerns | Some concerns (95% CrI crosses clinical threshold -1) | No concerns | No concerns (direct comparison; no closed loop) | Low |
| Pemvi SC 1.2mg QW | 1 | observed_within_4w_tabular | -7.63 (-10.06 to -5.01) | -10.39 to -4.67 | Some concerns (single phase 2 trial; D3/D5 attrition -1) | Low risk | No concerns | No concerns | Some concerns (95% PrI crosses clinical threshold -1) | No concerns (direct comparison; no closed loop) | Low |
| Survo SC 3.6mg QW | 1 | observed_within_4w_figure_derived (tabular_76w) | -7.32 (-9.61 to -4.68) | -10.09 to -4.34 | Some concerns (52-wk value figure-derived -1) | Low risk | No concerns | Some concerns (95% CrI crosses clinical threshold -1) | No concerns | No concerns (direct comparison; no closed loop) | Low |
| HRS-7535 PO 30mg QD | 1 | extrapolated_beyond_followup_class_borrowing | -6.89 (-13.28 to -1.05) | -13.44 to -0.86 | Some concerns (single phase 2 trial) | Low risk | Major concerns (extrapolated beyond follow-up -2) | Some concerns (95% CrI crosses clinical threshold -1) | No concerns | No concerns (direct comparison; no closed loop) | Very low |
| Orfor PO 12mg QD | 1 | observed_within_4w_figure_derived (tabular_72w) | -6.78 (-8.67 to -5.26) | -9.26 to -4.70 | Some concerns (52-wk value figure-derived -1) | Low risk | No concerns | No concerns | Some concerns (95% PrI crosses clinical threshold -1) | No concerns (direct comparison; no closed loop) | Low |
| NB PO 50/300mg QD | 1 | extrapolated_beyond_followup_class_borrowing | -6.53 (-23.80 to 10.82) | -23.78 to 10.94 | Some concerns (single pilot trial) | Low risk | Major concerns (extrapolated beyond follow-up -2) | Major concerns (95% CrI crosses both -5 pp and null -2) | No concerns | No concerns (direct comparison; no closed loop) | Very low |
| NB PO 16/360mg QD | 1 | extrapolated_beyond_followup_class_borrowing | -6.42 (-23.98 to 10.95) | -24.01 to 11.10 | Some concerns (D3/D5 some concerns) | High risk | Major concerns (extrapolated beyond follow-up -2) | Major concerns (95% CrI crosses both -5 pp and null -2) | No concerns | No concerns (direct comparison; no closed loop) | Very low |
| NB PO 48/360mg QD | 1 | extrapolated_beyond_followup_class_borrowing | -6.38 (-24.16 to 11.50) | -24.13 to 11.44 | Some concerns (D3/D5 some concerns) | Low risk | Major concerns (extrapolated beyond follow-up -2) | Major concerns (95% CrI crosses both -5 pp and null -2) | No concerns | No concerns (direct comparison; no closed loop) | Very low |
| NB PO 32/360mg QD | 2 | extrapolated_beyond_followup_class_borrowing | -6.26 (-23.79 to 10.84) | -23.78 to 10.69 | Some concerns (D3/D5 some concerns) | High risk | Major concerns (extrapolated beyond follow-up -2) | Major concerns (95% CrI crosses both -5 pp and null -2) | No concerns | No concerns (direct comparison; no closed loop) | Very low |
| Orfor PO 6mg QD | 1 | observed_within_4w_figure_derived (tabular_72w) | -5.97 (-7.86 to -4.49) | -8.41 to -3.90 | Some concerns (52-wk value figure-derived -1) | Low risk | No concerns | Some concerns (95% CrI crosses clinical threshold -1) | No concerns | No concerns (direct comparison; no closed loop) | Low |
| Sema SC 0.1mg QD | 1 | observed_within_4w_tabular | -5.97 (-7.88 to -3.97) | -8.32 to -3.50 | Some concerns (single phase 2 trial; D3/D5 some concerns -1) | Low risk | No concerns | Some concerns (95% CrI crosses clinical threshold -1) | No concerns | No concerns (closed loop consistent, p = 0.186) | Low |
| Amycretin PO 100mg QD | 1 | extrapolated_beyond_followup_class_borrowing | -4.86 (-29.70 to 20.31) | -29.77 to 20.30 | Some concerns (single phase 1 trial) | Low risk | Major concerns (extrapolated beyond follow-up -2) | Major concerns (95% CrI crosses both -5 pp and null -2) | No concerns | No concerns (direct comparison; no closed loop) | Very low |
| Amycretin PO 50mg QD | 1 | extrapolated_beyond_followup_class_borrowing | -4.71 (-30.18 to 20.49) | -30.35 to 20.61 | Some concerns (single phase 1 trial) | Low risk | Major concerns (extrapolated beyond follow-up -2) | Major concerns (95% CrI crosses both -5 pp and null -2) | No concerns | No concerns (direct comparison; no closed loop) | Very low |
| Lira SC 3mg QD | 6 | observed_within_4w_tabular | -4.20 (-5.49 to -2.89) | -6.18 to -2.21 | Some concerns (D3/D5 some concerns) | Some concerns | No concerns | Some concerns (95% CrI crosses clinical threshold -1) | No concerns (I2 = 20.2%) | Some concerns (closed-loop node-split p = 0.046 vs placebo -1) | Low |
| Sema SC 0.05mg QD | 1 | observed_within_4w_tabular | -3.50 (-5.42 to -1.48) | -5.89 to -1.10 | Some concerns (single phase 2 trial; D3/D5 some concerns -1) | Low risk | No concerns | Some concerns (95% CrI crosses clinical threshold -1) | No concerns | No concerns (closed loop consistent, p = 0.186) | Low |
| Orli PO 120mg TID | 4 | observed_within_4w_tabular | -3.11 (-4.23 to -1.93) | -4.99 to -1.21 | Some concerns (D1/D2/D3 attrition some concerns -1) | High risk | No concerns | No concerns (entire CrI/PrI lies between -5 pp and 0) | No concerns (I2 = 0%) | No concerns (closed loop consistent, p = 0.974) | Low |
| Orli PO 60mg TID | 2 | observed_within_4w_tabular | -2.14 (-3.55 to -0.68) | -4.21 to 0.01 | Some concerns (D1/D2/D3 attrition some concerns -1) | High risk | No concerns | No concerns | Some concerns (95% PrI crosses null 0 -1) | No concerns (closed loop consistent, p = 0.974) | Low |
| Dapi SC 6mg QW | 1 | extrapolated_beyond_followup_class_borrowing | -1.05 (-26.34 to 23.49) | -26.26 to 23.82 | Some concerns (single phase 2a trial) | Low risk | Major concerns (extrapolated beyond follow-up -2) | Major concerns (95% CrI crosses both -5 pp and null -2) | No concerns | No concerns (direct comparison; no closed loop) | Very low |

**Table S24.** Evidence funnel and analytical attrition cascade from eligible mapped trials to specific synthesis layers (denominators in independent randomisation units)

| Funnel level | Analytical stage and prespecified eligibility gate | Adults without T2D | Adults with T2D | Mixed or unstratified populations | Disposition of non-advancing units and exact reconciliation | Source tables |
|---|---|---|---|---|---|---|
| L0 Screening | Records screened after deduplication across 10 databases and registries | See PRISMA flow diagram | See PRISMA flow diagram | See PRISMA flow diagram | Excluded records failing PICO criteria, non-randomised studies, and trials with planned treatment shorter than 12 weeks | PRISMA flow diagram |
| L1 Eligible and mapped evidence base | All randomised trials meeting review inclusion criteria and retained in the master linked database | 138 randomisation units (across 178 total parent trials, 180 reports, 196 total randomisation units, 81,014 non-overlapping participants, and 160 target drug-strategy nodes) | 29 randomisation units | 29 randomisation units | All 196 independent randomisation units are retained in Clean NMA Database v1.2 and routed to specific analytical windows and sensitivity layers according to prespecified protocol gates (§45 and §55) | Table S1, Table S2 |
| L2 One-year reporting assessment set | Initiation-stage randomisation units with planned follow-up of at least 42 weeks evaluated for 48–80-week reporting status and ROB-MEN bias | 47 randomisation units | 18 randomisation units | 13 randomisation units | Total of 78 medium-to-long-term units evaluated in Table S15. The remaining 118 units (of 196) comprise 116 short-duration Phase 1/2 or mechanistic units (<42 weeks; routed to 10–14-week and 20–28-week cross-sectional networks and longitudinal extrapolation) and 2 isolated long-term extension cohorts (z027_longterm at 176 weeks and z206 at 160 weeks; isolated in sensitivity analyses to prevent double-counting) | Table S15, Table S20 |
| L3 Usable percentage weight-change data at 48–56 weeks | Randomisation units reporting usable percentage body-weight change within the 48–56-week window (plus units reporting only at 60–104 weeks for longitudinal anchoring) | 35 randomisation units at 48–56 weeks (plus 2 units reporting percentage weight change only at 60–104 weeks: STEP 5 z004 at 104 weeks and Tronieri z246_study1 at 60 weeks) | 13 randomisation units at 48–56 weeks (plus 2 units reporting percentage weight change only at 76–80 weeks: TRIUMPH-2 z174 and SYNCHRONIZE-2 z175) | 4 randomisation units at 48–56 weeks (plus 4 units reporting only at 60–72 weeks) | In adults without T2D, 10 of 47 units did not report compatible 48–56-week percentage weight change (reported kg change only, binary/safety outcomes only, or trial-product/efficacy estimands only, such as Retatrutide Phase 2 z057). In adults with T2D, 3 of 18 units reported kg weight change or safety outcomes only | Table S15 |
| L4 Primary 48–56-week reference network | Randomisation units meeting strict treatment-policy-compatible estimand criteria and connected to the primary 48–56-week reference network | 28 randomisation units / trials connecting 46 nodes (45 active strategies and placebo; includes 27 placebo-controlled units and 1 active head-to-head unit, SURMOUNT-5 z029) | 9 randomisation units / trials connecting 15 nodes (14 active strategies and placebo) | Not pooled into a primary ranking network (routed to binary responder networks and comorbidity sensitivity analyses) | Exact reconciliation for adults without T2D (35 minus 28 equals 7 units): 5 units used historical LOCF imputation (SCALE Obesity z036, EQUIP z097, COR-I z102, COR-II z103_initial, COR-BMOD z104) and were isolated to Table S12 sensitivity analyses; 1 unit (Wadden 2019 z040) used intensive behavioural therapy as comparator without a drug-placebo arm; 1 unit (BELIEVE z077, bimagrumab plus semaglutide Phase 2) evaluated non-target connector arms under a trial-product primary estimand and contributed to binary safety networks only. Exact reconciliation for adults with T2D (13 minus 9 equals 4 units): all 4 units (OB-202/DM-230 z100, COR-Diabetes z105, Hollander 1998 z118, Miles 2002 z123) used historical LOCF imputation and were isolated to sensitivity analyses | Table S3, Table S4, Table S12 |
| L4' Primary Bayesian longitudinal model | Randomisation units with treatment-policy-compatible percentage weight-change contrasts across any follow-up timepoint (12–104 weeks) informing the restricted cubic spline longitudinal NMA at 52 weeks | 50 randomisation units (from 48 parent trials; informing 77 target strategy nodes) | 11 randomisation units (from 11 parent trials; informing 19 target strategy nodes) | Isolated in mixed-population sensitivity analyses | Incorporates the 28 (without T2D) and 9 (with T2D) reference-network units plus units reporting at 60–104 weeks and short-duration (12–40-week) treatment-policy units | Table S5, Table S21 |
| L5 One-year ranking and clinical decision set | Target strategies with non-extrapolated 52-week support eligible for SUCRA ranking and adjacent dose-escalation trade-off evaluation | 45 ranked strategies (32 supported by 48–56-week tabular endpoints; 13 supported by 48–56-week figure-derived trajectories paired with 64–80-week tabular endpoints); 32 short-duration strategies (<48 weeks) labelled extrapolated and excluded from ranking | 16 non-extrapolated strategies (14 with direct 48–56-week support; 2 anchored by 76–80-week endpoints) | Excluded from primary ranking | Prevents short-term (12–36-week) Phase 1/2 strategies (such as cagrilintide monotherapy, bofanglutide, VCT220, HRS-7535, and oral amycretin) from distorting 1-year comparative rankings | Table S5, Table S23 |
| L6 Implementation and decision contrasts | Specific initiation-stage clinical decision analyses (dose escalation, titration speed, lifestyle-support increment, joint response-persistence bounds, and T2D attenuation) | 22 adjacent maintenance-dose steps (69 tolerance rows across 0.5, 1.0, and 2.0 percentage points); 8 randomised titration contrasts at 48–56 weeks across identical maintenance doses (Layer D: 6 treatment-policy, 2 trial-product; 4 trials; Table S19); 7 randomised lifestyle-support increment contrasts (Layer B, all single-trial); 4 units (16 arm-level rows) for Fréchet-Hoeffding joint bounds; 9 matched programmes (18 dose pairs) for T2D attenuation | 9 timepoint-matched trial programmes (18 dose pairs) informing the multilevel T2D ratio-of-means model | Evaluated in comorbidity-exclusion sensitivity analyses | Identifies a major evidence gap: zero randomised trials test structured lifestyle-support increments under a shared modern incretin medication at 52 weeks using a treatment-policy estimand | Table S14, Table S16, Table S17, Table S18, Table S19 |

**Table S25.**
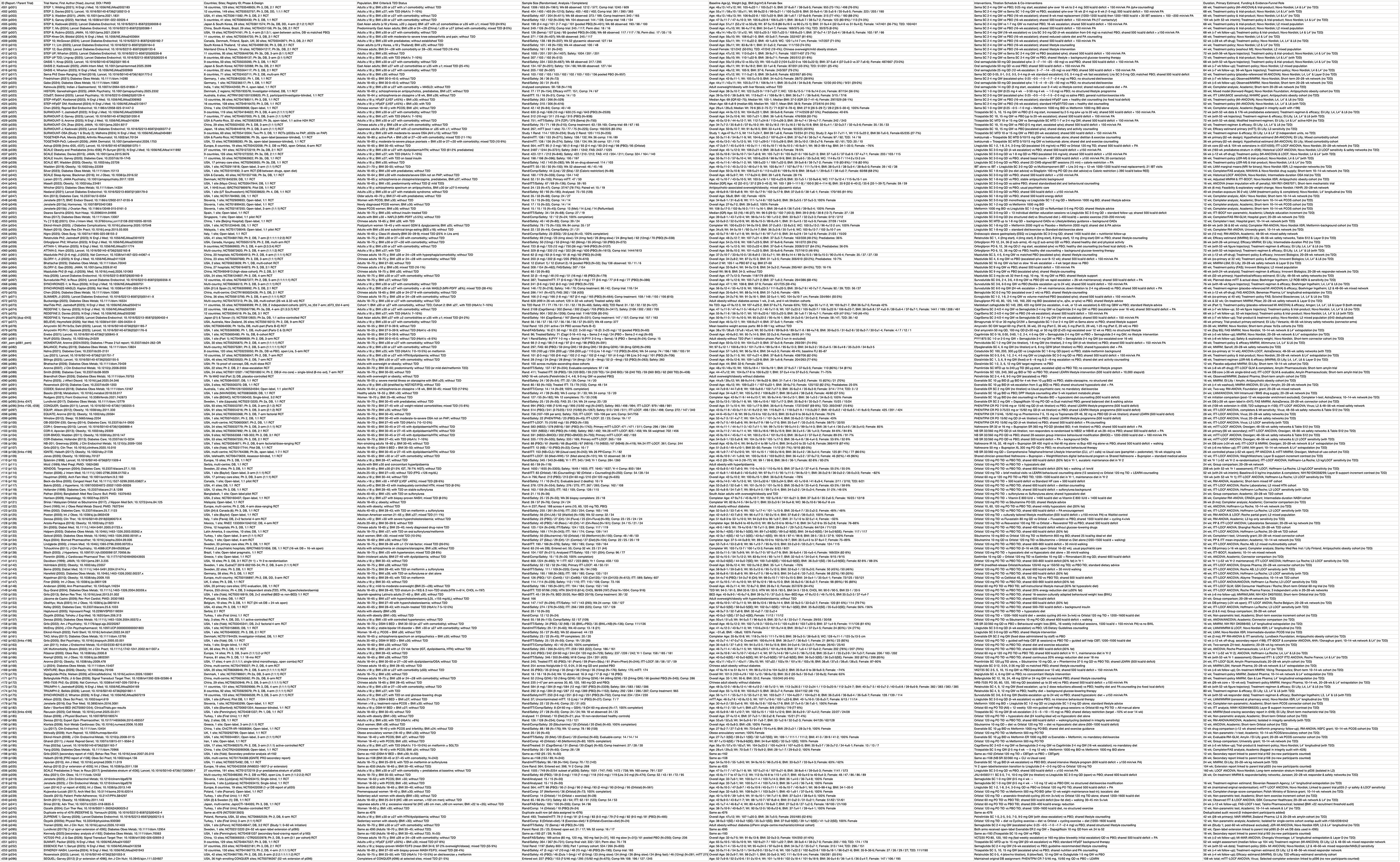
Full Baseline Characteristics Table.

